# Social Distancing with Movement Restrictions and the Effective Replication Number of COVID-19: Multi-Country Analysis Based on Phone Mobility Data

**DOI:** 10.1101/2020.10.08.20209064

**Authors:** Mounir Ould Setti, Ari Voutilainen

## Abstract

Linking phone mobility data to the effective replication number (Rt) could help evaluation of the impact of social distancing on the coronavirus disease 2019 (COVID-19) spread and estimate the time lag (TL) needed for the effect of movement restrictions to appear. We used a time-series analysis to discover how patterns of five indicators of mobility data relate to changes in Rt of 125 countries distributed over three groups based on Rt-mobility correlation. Group 1 included 71 countries in which Rt correlates negatively with residential and positively with other mobility indicators. Group 2 included 25 countries showing an opposite correlation pattern to Group 1. Group 3 included the 29 remaining countries. We chose the best-fit TL based on forecast and linear regression models. We used linear mixed models to evaluate how mobility indicators and the stringency index (SI) relate with Rt. SI reflects the strictness of governmental responses to COVID-19. With a median of 14 days, TLs varied across countries as well as across groups of countries. There was a strong negative correlation between SI and Rt in most countries belonging to Group 1 as opposed to Group 2. SI (units of 10%) associated with decreasing Rt in Group 1 [β −0.15, 95% CI −0.15 – (−0.14)] and Group 3 [-0.05, −0.07 – (−0.03)], whereas, in Group 2, SI associated with increasing Rt (0.13, 0.11 – 0.16). Mobile phone mobility data could contribute evaluations of the impact of social distancing with movement restrictions on the spread of the COVID-19.

## Introduction

By October 6, 2020, nearly 36 million humans have come down with the coronavirus disease 2019 (COVID-19) with the death toll exceeding one million.[1] While the virus continues to spread exponentially in some countries and others brace for a second wave of the disease, governments are tracking the effective reproduction number (Rt) to evaluate disease spread and the effectiveness of social distancing and movement restrictions.[2] Whereas the more commonly known basic reproduction number (R0) reflects the innate capacity of an infectious agent to spread,[3] Rt captures the dynamic changes of its spread that vary over time as a result of societies changing their behavior and gaining immunity. To reduce an individual’s capacity to transmit COVID-19, governments have been implementing a variety of movement restrictions. To evaluate the effectiveness of these measures, researchers have attempted to systematically associate changes in COVID-19 incidence with changes in Rt to schedule restriction actions. For example, Alfano and Ercolano[4] used the ACAPS #COVID19 Government Measures Dataset, which gathers implementation dates of restriction interventions in several countries and regions.[5] Despite including some regional data, the dataset does not allow proper aggregation of restriction interventions at the country level. Moreover, the dataset lacks information regarding the strength of restriction actions.

To account for the strength of restriction actions, researchers from Blavatnik School of Government developed the stringency index (SI), an indicator of the strictness of governmental responses to COVID-19.[6] Based on qualitative measures of restriction policies, SI, however, does not reflect the extent to which the legislated measures are enforced in practice or the social compliance and response, which could vary across cultures, regions, and times independently of the restriction policies.

Aggregated mobile community mobility data track the population mobility behavior and, during a pandemic, the data could provide good indicators of the regional extent of social distancing. Multiple studies have used mobility data to evaluate the social adherence to movement restriction policies against COVID-19[7] and to investigate how changes in mobility relate to the COVID-19 incidence and Rt.[8]

As a function of the performance of health systems in tracking, testing, and reporting cases, a time lag (TL) is expected between the implementation of a restriction action and the possible change in disease incidence. Linka et al. have identified great variability in TLs across countries regarding COVID-19 incidence.[8] TLs can i) provide an idea of countries’ early detection capabilities of COVID-19 cases, ii) improve planning of lockdown exit strategies, iii) guide the length of recommended quarantines, and iv) permit a timeframe to prepare for predicted disease peaks.

The object of this study is to analyze how patterns of mobility data can be linked to COVID-19 Rt changes in 125 countries with and without consideration of SI.

## Methods

### Data Sources and Variables of Interest

COVID-19 Community Mobility Reports[9] provided country-specific mobility data. Google aggregates anonymized position data from users who have the Location History activated on their mobile phones to reflect regional trends of community mobility in different regions and varied places. The dataset consists of daily regional percentages of change from the median value of the corresponding weekday during the 5-week baseline period (Jan 3 to Feb 6, 2020.) By September 18, 2020, the dataset was publicly available. In addition to the mobility data, we obtained data on COVID-19 daily case incidence from the GitHub COVID-19 repository of Johns Hopkins University[1] and data on policy responses and stringency of government measures for 180 countries from the Oxford COVID-19 Government Response Tracker (OxCGRT).[6]

In this study, we focused on the period from 15 February to 11 September 2020 with days as a time unit. For each country, we considered days starting from the 2nd confirmed COVID-19 case. After excluding countries with less than 60 days reporting new cases, mobility, or SI, we retained 125 countries for the analysis. The analyzed countries had a median observation time of 188 days. COVID-19 Open-Data GitHub repository eased access to data sources.[10] This study needed no ethical review since the observations concerned countries, not identified persons.[11]

With the help of the R-package Epi-Estim,[12] we calculated daily changes in Rt for each country based on the COVID-19 incidence. As the virus is suggested to spread faster than SARS but slower than H1N1,[13] we used, as a parameter in our computations of Rt, the serial interval of COVID-19 proposed by Nishiura et al.[14] [mean 4.7, standard deviation (SD) 2.9]. From Google’s COVID-19 Community Mobility Reports we obtained the indicators of daily changes in mobility in five categories: retails and recreation places (named in analyses as ‘retail’), grocery stores and pharmacies (grocery), transit stations (transit), residential places (residential), and workplaces (work). Google Community Mobility Reports also comprises information on mobility in parks, but like Vokó and Pitter,[15] we omitted this variable from our analyses as the effect of the time spent in parks on disease spread is unclear.

We used the daily SI computed by the OxCGRT[6] as a single numerical measure to aggregate different types of governmental responses and levels of restriction policies in each country. From Wikidata we obtained demographics, the number of internet users, and the Human Development index (HDI) for each country.

### Data Analysis

We performed all analyses by means of the R version 4.0.2.[16]

#### Preliminary Analysis

First, we organized the data as timeseries of daily mobility, SI, and Rt. Second, we derived 30 datasets from the timeseries for each country by lagging the mobility observations by 1 to 30 days forward (corresponding to possible TL values).[17] As a preliminary analysis, we evaluated trends in mobility indicators and SI and estimated change-points in country-specific trends for each mobility indicator and SI using the Buishand range test.[18] Moreover, based on the directions of Pearson’s correlation coefficients (r) between mobility indicators and Rt changes, we identified three groups of countries. Group 1 included 71 countries with positive correlations of Rt with retail, grocery, transit, and work mobility but a negative correlation of Rt with residential mobility. The reason of that choice was that increased residential mobility is expected to associate with a decrease in disease spread as opposed to the other mobility indicators. Group 2 included 25 countries showing an opposite correlation pattern to Group 1. Group 3 included 29 remaining countries. We compared the groups’ sociodemographic characteristics using Kruskal-Wallis Ranks Sum test.

#### Predictive Modeling

We applied Facebook Prophet, an open-source forecasting package for timeseries,[19] to fit additive non-linear models regarding changes of Rt in each country over its 30 datasets (the possible TL values) and used the five mobility variables as regressors. We did not adjust for seasonality or trend. We evaluated the accuracy of each Prophet forecast using the mean absolute percentage error (MAPE), estimated by means of the Prophet’s cross validation function, and the adjusted explanatory power, R^2^.

Similarly, for each country’s 30 datasets, we fitted a linear regression model with Rt as the dependent variable and the five lagged mobility indicators as independent variables.

#### Selection of the Best Fitting Time Lag

We assumed that the TL remains constant over time and does not vary across mobility indicators. To select the bestfit TL value for each country, we evaluated the following parameters for the Prophet’s forecast models: MAPE and the adjusted R^2^, and the following parameters for the linear regression models: p-value, the adjusted R^2^, the F-test value, and the likelihood-ratio. We also examined cross-correlation plots of Rt with all mobility indicators.[20]

We based the final choice regarding the best-fit TL on manual inspections of country-anonymized plots of rescaled performance indicators (Supplementary material). We prioritized early change points over the late ones, criteria concerning the regression models over Prophet-forecasts’ performances, MAPE over Prophet-forecasts’ adjusted R^2^, and rapid changes in curves over extreme values.

#### The Stringency Index

After lagging each country’s mobility data using its best-fit TL, we fitted a linear mixed model for each Group to evaluate the role of mobility indicators and stringency as fixed effects factors and countries as random factors in predicting Rt. We also fitted linear mixed models to predict Rt with SI alone. We applied the maximum likelihood method and we scaled the fixed factors to their respective datasets prior to modeling. We used the R packages ‘lme4’[21], ‘afex’[22], and ‘insight’[23] to fit and assess the mixed effects models.

## Results

The inspection of mobility trends suggested the existence of two major changing points in most countries. The first changing point in mobility trends corresponded to the first change from normal to social distancing with an increase of residential mobility and a decrease of grocery, transit, retail, and workplace mobility. It occurred mostly (in 40 – 60 countries) during the second and third weeks of March coinciding with the beginning of rigorous lockdown policies, as the US and European countries started closing borders. The second changing point of mobility trends was more widely scattered and occurred around mid-June, corresponding to the slow return to baseline patterns of mobility.

The mean best-fit TL was 14.6 days [median 14, interquartile range (IQR) 10 – 19] (Figs. 1 and 2). Continent-wise, Europe had the lowest median TL (12.5 days, IQR 10 – 14.75) followed by Asia & Oceania (14, 10 – 20), the Americas (16, 10 – 19), and Africa (16, 9.5 – 22).

**Fig. 1.**
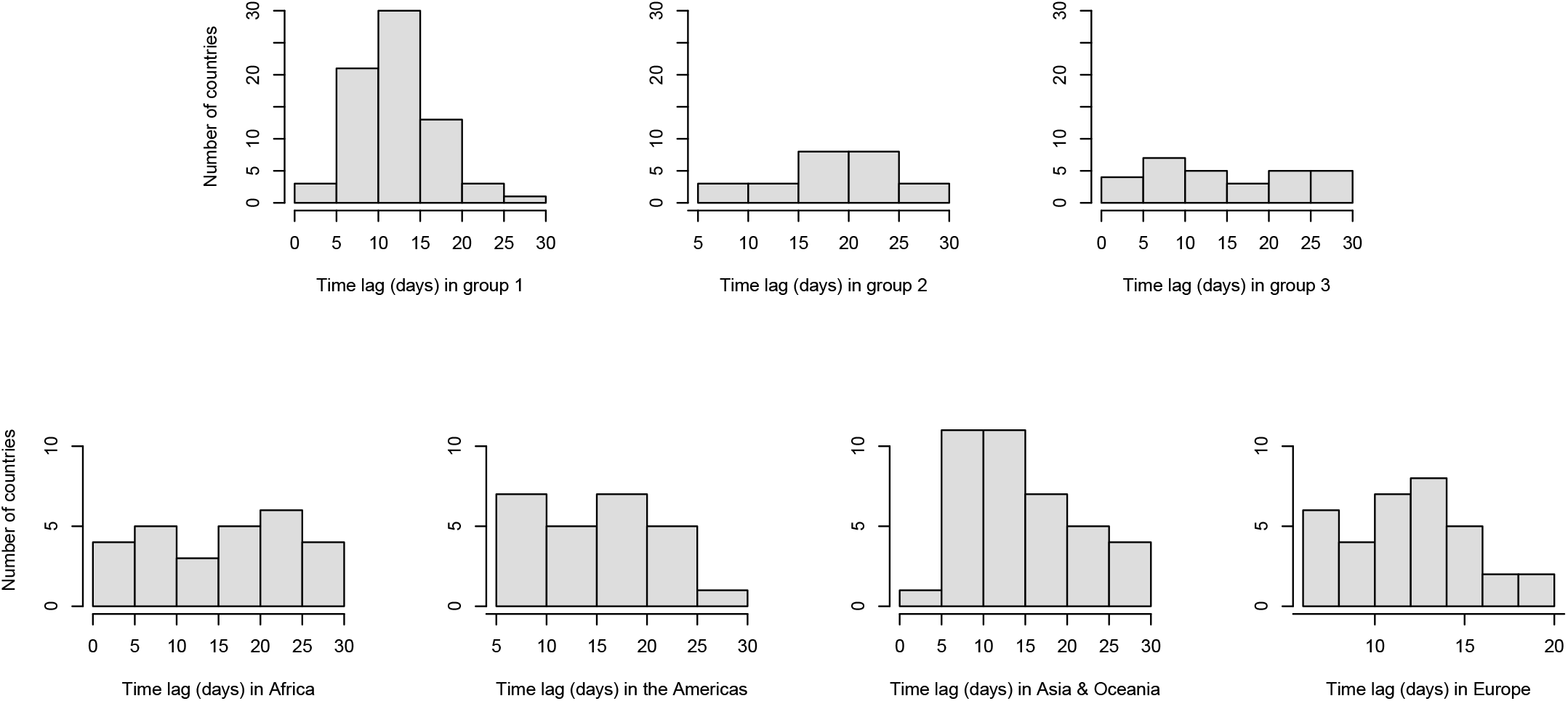
Distribution of the estimated time lag over groups and continents. Group 1 (n = 71) includes countries with a positive correlation of the effective replication number with mobility in retail, grocery, transit, and workplace, and a negative correlation of the effective replication number with residential mobility; group 2 (n = 25) includes countries with a negative correlation of the effective replication number with mobility in retail, grocery, transit, and workplace, and a positive correlation of the effective replication number with residential mobility; group 3 (n = 29) includes the remaining countries.

**Fig. 2.**
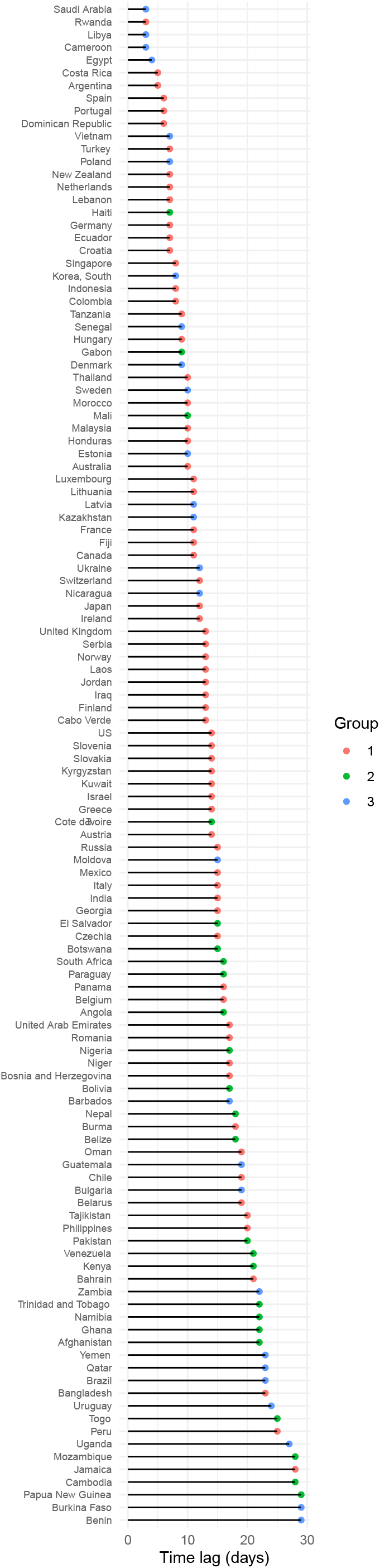
Estimated time lag by country. Note. The following countries had a p-value > 0.05 of the regression model of their effective reproduction number on lagged mobility, and were so excluded: Sri Lanka, Mongolia, Mauritius, Taiwan, and Zimbabwe.

In comparison to Group 2, Group 1 had significantly shorter TLs, lower percentages of forecasting error (MAPE), an older population, and a higher HDI (Table 1). The forecasting models performed well, especially in Group 1, with MAPE as low as 6% in the US for example (Table 2).

**Table 1.**
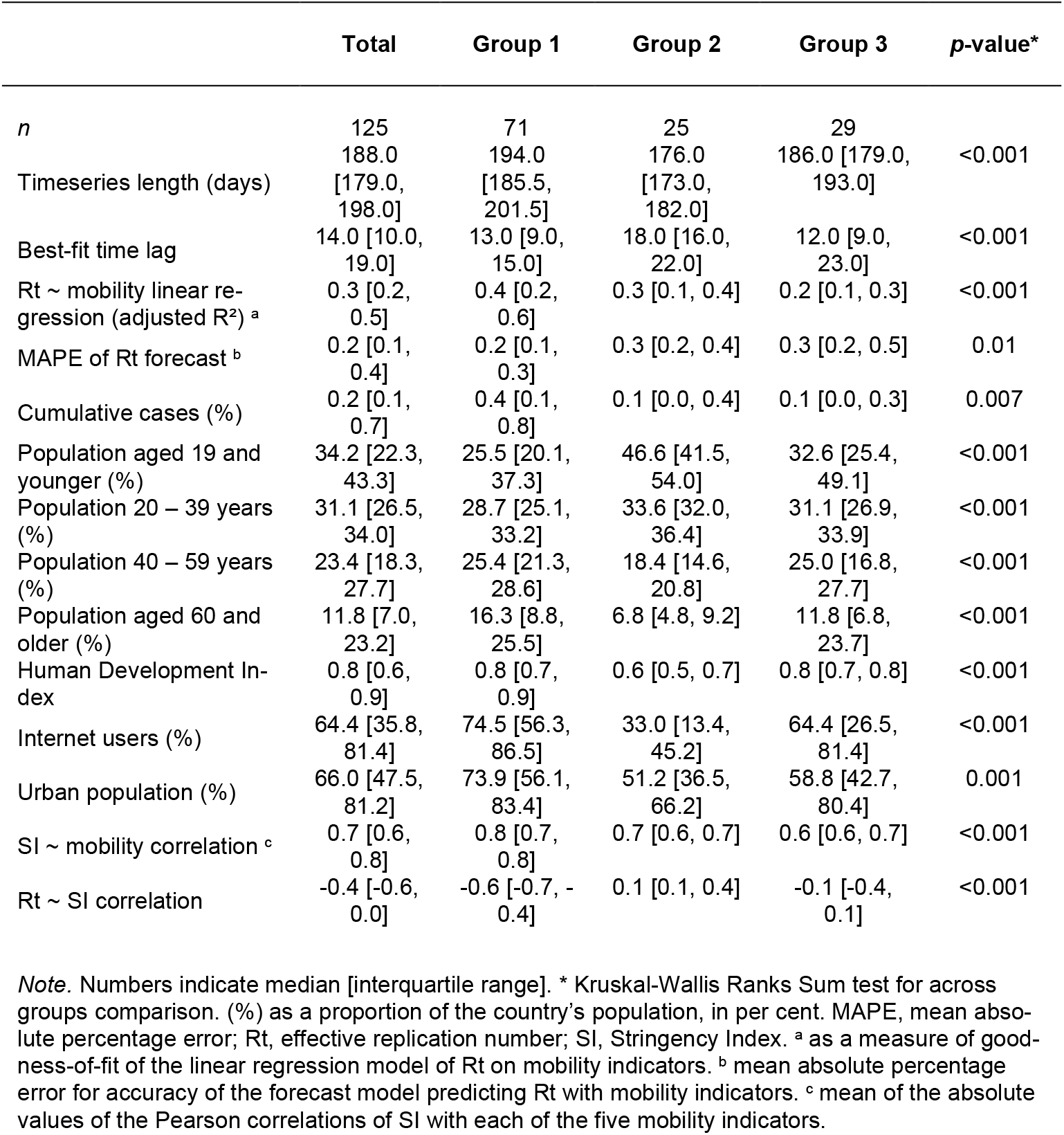
Characteristics of the studied countries by Group.

|  | Total | Group 1 | Group 2 | Group 3 | p-value* |
| --- | --- | --- | --- | --- | --- |
| *n* | 125 | 71 | 25 | 29 |  |
| Timeseries length (days) | 188.0<br>[179.0, 198.0] | 194.0<br>[185.5, 201.5] | 176.0<br>[173.0, 182.0] | 186.0 [179.0, 193.0] | <0.001 |
| Best-fit time lag | 14.0 [10.0, 19.0] | 13.0 [9.0, 15.0] | 18.0 [16.0, 22.0] | 12.0 [9.0, 23.0] | <0.001 |
| Rt ~ mobility linear regression (adjusted R<sup>2</sup>)<sup>a</sup> | 0.3 [0.2, 0.5] | 0.4 [0.2, 0.6] | 0.3 [0.1, 0.4] | 0.2 [0.1, 0.3] | <0.001 |
| MAPE of Rt forecast<sup>b</sup> | 0.2 [0.1, 0.4] | 0.2 [0.1, 0.3] | 0.3 [0.2, 0.4] | 0.3 [0.2, 0.5] | 0.01 |
| Cumulative cases (%) | 0.2 [0.1, 0.7] | 0.4 [0.1, 0.8] | 0.1 [0.0, 0.4] | 0.1 [0.0, 0.3] | 0.007 |
| Population aged 19 and younger (%) | 34.2 [22.3, 43.3] | 25.5 [20.1, 37.3] | 46.6 [41.5, 54.0] | 32.6 [25.4, 49.1] | <0.001 |
| Population 20 – 39 years (%) | 31.1 [26.5, 34.0] | 28.7 [25.1, 33.2] | 33.6 [32.0, 36.4] | 31.1 [26.9, 33.9] | <0.001 |
| Population 40 – 59 years (%) | 23.4 [18.3, 27.7] | 25.4 [21.3, 28.6] | 18.4 [14.6, 20.8] | 25.0 [16.8, 27.7] | <0.001 |
| Population aged 60 and older (%) | 11.8 [7.0, 23.2] | 16.3 [8.8, 25.5] | 6.8 [4.8, 9.2] | 11.8 [6.8, 23.7] | <0.001 |
| Human Development Index | 0.8 [0.6, 0.9] | 0.8 [0.7, 0.9] | 0.6 [0.5, 0.7] | 0.8 [0.7, 0.8] | <0.001 |
| Internet users (%) | 64.4 [35.8, 81.4] | 74.5 [56.3, 86.5] | 33.0 [13.4, 45.2] | 64.4 [26.5, 81.4] | <0.001 |
| Urban population (%) | 66.0 [47.5, 81.2] | 73.9 [56.1, 83.4] | 51.2 [36.5, 66.2] | 58.8 [42.7, 80.4] | 0.001 |
| SI ~ mobility correlation<sup>c</sup> | 0.7 [0.6, 0.8] | 0.8 [0.7, 0.8] | 0.7 [0.6, 0.7] | 0.6 [0.6, 0.7] | <0.001 |
| Rt ~ SI correlation | -0.4 [-0.6, 0.0] | -0.6 [-0.7, -0.4] | 0.1 [0.1, 0.4] | -0.1 [-0.4, 0.1] | <0.001 |

*Note.* Numbers indicate median [interquartile range]. \* Kruskal-Wallis Ranks Sum test for across groups comparison. (%) as a proportion of the country's population, in per cent. MAPE, mean absolute percentage error; Rt, effective replication number; SI, Stringency Index. <sup>a</sup> as a measure of goodness-of-fit of the linear regression model of Rt on mobility indicators. <sup>b</sup> mean absolute percentage error for accuracy of the forecast model predicting Rt with mobility indicators. <sup>c</sup> mean of the absolute values of the Pearson correlations of SI with each of the five mobility indicators.

**Table 2.** Time lag and prediction performances by country.

| Country | TL | MAPE | Adj. R<sup>2</sup> | Country | TL | MAPE | Adj. R<sup>2</sup> | Country | TL | MAPE | Adj. R<sup>2</sup> | Country | TL | MAPE | Adj. R<sup>2</sup> |
| --- | --- | --- | --- | --- | --- | --- | --- | --- | --- | --- | --- | --- | --- | --- | --- |
| Afghanistan | 22 | 0.17 | 0.62 | Egypt | 4 | 0.37 | 0.51 | Libya | 3 | 0.3 | 0.22 | Saudi Arabia | 3 | 0.2 | 0.65 |
| Angola | 16 | 0.41 | 0.12 | El Salvador | 15 | 0.3 | 0.38 | Lithuania | 11 | 0.12 | 0.2 | Senegal | 9 | 0.15 | 0.33 |
| Argentina | 5 | 0.1 | 0.72 | Estonia | 10 | 0.43 | 0.3 | Luxembourg | 11 | 0.29 | 0.45 | Serbia | 13 | 0.28 | 0.47 |
| Australia | 10 | 0.17 | 0.56 | Fiji | 11 | 0.88 | 0.09 | Malaysia | 10 | 0.2 | 0.33 | Singapore | 8 | 0.49 | 0.25 |
| Austria | 14 | 0.19 | 0.55 | Finland | 13 | 0.07 | 0.42 | Mali | 10 | 0.33 | 0.25 | Slovakia | 14 | 0.19 | 0.32 |
| Bahrain | 21 | 0.23 | 0.13 | France | 11 | 0.25 | 0.41 | Mauritius | 16 | 1.01 | 0.01 | Slovenia | 14 | 0.41 | 0.45 |
| Bangladesh | 23 | 0.08 | 0.58 | Gabon | 9 | 0.18 | 0.42 | Mexico | 15 | 0.04 | 0.68 | South Africa | 16 | 0.21 | 0.57 |
| Barbados | 17 | 0.91 | 0.09 | Georgia | 15 | 0.54 | 0.1 | Moldova | 15 | 0.08 | 0.43 | Spain | 6 | 0.09 | 0.55 |
| Belarus | 19 | 0.16 | 0.39 | Germany | 7 | 0.1 | 0.49 | Mongolia | 27 | 0.82 | 0.03 | Sri Lanka | 28 | 0.36 | 0.01 |
| Belgium | 16 | 0.28 | 0.52 | Ghana | 22 | 0.55 | 0.26 | Morocco | 10 | 0.08 | 0.43 | Sweden | 10 | 0.21 | 0.28 |
| Belize | 18 | 0.33 | 0.32 | Greece | 14 | 0.2 | 0.25 | Mozambique | 28 | 0.15 | 0.19 | Switzerland | 12 | 0.08 | 0.57 |
| Benin | 29 | 0.24 | 0.09 | Guatemala | 19 | 0.08 | 0.28 | Namibia | 22 | 0.47 | 0.47 | Taiwan | 8 | 1.64 | 0.02 |
| Bolivia | 17 | 0.05 | 0.36 | Haiti | 7 | 0.26 | 0.26 | Nepal | 18 | 0.12 | 0.25 | Tajikistan | 20 | 0.07 | 0.52 |
| Bosnia and Herzegovina | 17 | 0.18 | 0.4 | Honduras | 10 | 0.27 | 0.38 | Netherlands | 7 | 0.52 | 0.62 | Tanzania | 9 | 0.29 | 0.66 |
| Botswana | 15 | 1.45 | 0.12 | Hungary | 9 | 0.17 | 0.41 | New Zealand | 7 | 0.49 | 0.16 | Thailand | 10 | 0.23 | 0.41 |
| Brazil | 23 | 0.04 | 0.58 | India | 15 | 0.05 | 0.7 | Nicaragua | 12 | 0.61 | 0.11 | Togo | 25 | 0.16 | 0.1 |
| Bulgaria | 19 | 0.19 | 0.11 | Indonesia | 8 | 0.06 | 0.61 | Niger | 17 | 0.56 | 0.09 | Trinidad and Tobago | 22 | 0.32 | 0.13 |
| Burkina Faso | 29 | 0.41 | 0.1 | Iraq | 13 | 0.12 | 0.39 | Nigeria | 17 | 0.17 | 0.45 | Turkey | 7 | 0.09 | 0.29 |
| Burma | 18 | 0.43 | 0.24 | Ireland | 12 | 0.21 | 0.48 | Norway | 13 | 0.37 | 0.34 | Uganda | 27 | 0.34 | 0.2 |
| Cabo Verde | 13 | 0.16 | 0.12 | Israel | 14 | 0.14 | 0.43 | Oman | 19 | 0.25 | 0.41 | Ukraine | 12 | 0.05 | 0.4 |
| Cambodia | 28 | 4.97 | 0.32 | Italy | 15 | 0.1 | 0.7 | Pakistan | 20 | 0.25 | 0.22 | United Arab Emirates | 17 | 0.26 | 0.46 |
| Cameroon | 3 | 0.49 | 0.12 | Jamaica | 28 | 0.35 | 0.16 | Panama | 16 | 0.15 | 0.23 | United Kingdom | 13 | 0.11 | 0.72 |
| Canada | 11 | 0.09 | 0.83 | Japan | 12 | 0.31 | 0.48 | Papua New Guinea | 29 | 1.17 | 0.33 | Uruguay | 24 | 0.46 | 0.08 |
| Chile | 19 | 0.05 | 0.65 | Jordan | 13 | 0.35 | 0.12 | Paraguay | 16 | 0.18 | 0.04 | US | 14 | 0.06 | 0.76 |
| Colombia | 8 | 0.08 | 0.58 | Kazakhstan | 11 | 0.25 | 0.19 | Peru | 25 | 0.29 | 0.37 | Venezuela | 21 | 0.26 | 0.06 |
| Costa Rica | 5 | 0.11 | 0.66 | Kenya | 21 | 0.21 | 0.49 | Philippines | 20 | 0.2 | 0.58 | Vietnam | 7 | 0.46 | 0.1 |
| Ivory Coast | 14 | 0.25 | 0.13 | South Korea | 8 | 0.38 | 0.24 | Poland | 7 | 0.13 | 0.4 | Yemen | 23 | 0.35 | 0.18 |
| Croatia | 7 | 0.25 | 0.15 | Kuwait | 14 | 0.11 | 0.35 | Portugal | 6 | 0.16 | 0.8 | Zambia | 22 | 0.56 | 0.17 |
| Czechia | 15 | 0.06 | 0.33 | Kyrgyzstan | 14 | 0.22 | 0.19 | Qatar | 23 | 0.12 | 0.7 | Zimbabwe | 24 | 1.15 | 0.04 |
| Denmark | 9 | 0.36 | 0.19 | Laos | 13 | 1.5 | 0.21 | Romania | 17 | 0.08 | 0.73 |  |  |  |  |
| Dominican Republic | 6 | 0.08 | 0.59 | Latvia | 11 | 0.55 | 0.22 | Russia | 15 | 0.08 | 0.58 |  |  |  |  |
| Ecuador | 7 | 0.37 | 0.53 | Lebanon | 7 | 0.09 | 0.28 | Rwanda | 3 | 0.42 | 0.18 |  |  |  |  |

Note. TL, time lag; MAPE, mean absolute percentage error of the forecast model (the lower, the better is the forecast); Adj. R<sup>2</sup>, adjusted r-squared of the regression model.

In all countries, SI associated with mobility indicators through correlation and linear regressions (p-values < 0.001 in the models of 124 countries). There was a strong negative correlation between SI and Rt in most countries belonging to Group 1. In most countries belonging to Group 2, the correlation was positive (Fig. 3).

**Fig. 3.**
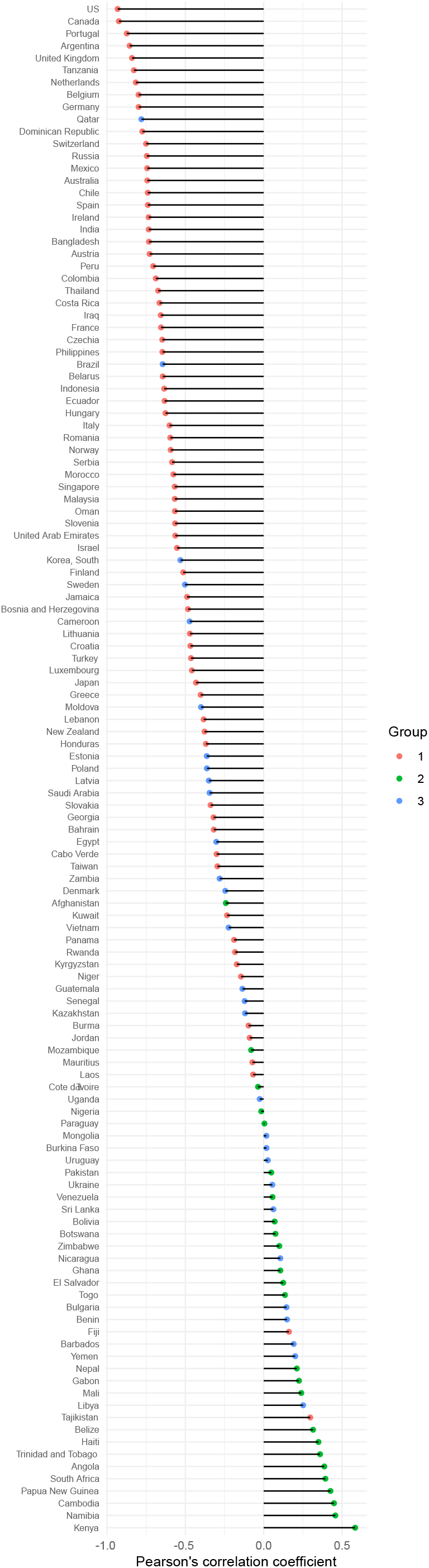
Pearson’s correlation coefficient of the association between the effective reproduction number and the stringency index by country. Note. Positive correlation values suggest that a high stringency was favoring disease spread.

Table 3 summarizes results of the analysis of the full linear mixed models with effect sizes labelled according to Funder & Ozer.[24] Models fitted to Groups 1 and 2 showed substantial explanatory power (conditional R^2^ = 0.44 and 0.42, respectively), unlike the model fitted to Group 3 (conditional R^2^ = 0.05). Removing SI from the mixed models brought significance to retail mobility in Group 1 [β 0.09, 95% confidence interval (CI) 0.05 – 0.14], and to grocery (− 0.14, −0.22 – −0.06) and transit (0.12, 0.01 – 0.23) mobility in Group 3; the other factors retained the same pattern. The analysis of the mixed models of Rt on SI without mobility variables (rescaling not needed) suggested an association of high stringency with disease spread in Group 2, with an increase of 0.13 in Rt for every 10% of increase in SI (95% CI, 0.11 – 0.16). SI without mobility variables associated with a decrease in Rt in Group 1 (β −0.15 for units of 10% of SI, 95% CI −0.15 – −0.14) and Group 3 (−0.05, −0.07 – −0.03).

**Table 3.** Results of linear mixed models predicting COVID-19 effective replication number (Rt) based on mobility and stringency.

| Predictors <sup>a</sup> | Rt - Group 1 |  |  | Rt - Group 2 |  |  | Rt - Group 3 |  |  |
| --- | --- | --- | --- | --- | --- | --- | --- | --- | --- |
|  | Estimates | CI | p | Estimates | CI | p | Estimates | CI | p |
| (Intercept) | 1.3 | 1.18 – 1.42 | **<0.001** | 1.41 | 1.14 – 1.68 | **<0.001** | 1.19 | 1.11 – 1.27 | **<0.001** |
| retail mobility | 0 | -0.05 – 0.04 | 0.937 | -0.68 | -0.85 – -0.50 | **<0.001** | -0.02 | -0.13 – 0.09 | 0.756 |
| grocery mobility | -0.11 | -0.14 – -0.08 | **<0.001** | 0.16 | 0.02 – 0.30 | **0.024** | -0.07 | -0.16 – 0.01 | 0.096 |
| transit mobility | 0.28 | 0.23 – 0.33 | **<0.001** | 0.02 | -0.12 – 0.16 | 0.78 | 0.07 | -0.05 – 0.18 | 0.256 |
| residential mobility | 0.26 | 0.22 – 0.31 | **<0.001** | -0.54 | -0.68 – -0.40 | **<0.001** | 0.06 | -0.03 – 0.16 | 0.188 |
| workplace mobility | 0.14 | 0.11 – 0.17 | **<0.001** | -0.39 | -0.48 – -0.30 | **<0.001** | 0.03 | -0.03 – 0.09 | 0.372 |
| Stringency Index | -0.28 | -0.31 – -0.25 | **<0.001** | 0.06 | 0.00 – 0.12 | **0.047** | -0.15 | -0.21 – -0.10 | **<0.001** |
| **Random Effects** |  |  |  |  |  |  |  |  |  |
| $\sigma^2$ | 0.52 | | | 1.01 | | | 1.29 | | |
| $\tau_{00}$ | 0.28 | | | 0.40 | | | 0.04 | | |
| ICC | 0.35 |  |  | 0.32 |  |  | 0.03 |  |  |
| n (countries) | 71 |  |  | 25 |  |  | 29 |  |  |
| Observations (days) | 12736 |  |  | 3928 |  |  | 4930 |  |  |
| Marginal R<sup>2</sup> / Conditional R<sup>2</sup> | 0.143 / 0.444 |  |  | 0.154 / 0.423 |  |  | 0.017 / 0.047 |  |  |

Note. CI, 95% confidence interval; ICC, Intraclass Correlation Coefficient; Rt, effective replication number; $\sigma^2$ , total variance; $\tau_{00}$ , random-intercept-variance. <sup>a</sup> all predictors' scales have been standardized.

## Discussion

Through a timeseries regression study, we examined how mobility patterns from aggregated mobile phones data relate to the variation of COVID-19 Rt in 125 countries during the period from February 15 to September 11, 2020. Our analyses showed that changes in community mobility for their part explain the variation in Rt and that community mobility forecasts Rt even with an accuracy of 90%. Based on our findings, specifically the directions of correlations between community mobility indicators and Rt, we suggest that it is possible to distribute countries into distinct groups that differ from each other regarding the effectiveness of social distancing and movement restrictions in reducing Rt. Most European countries belonged to Group 1 and none belonged to Group 2.

Countries differ from each other regarding how promptly they report diseased persons as confirmed COVID-19 cases. This TL is termed as the time delay,[8] or the positivity detection time,[25] and it could correspond to the necessary time before the effect of restriction actions is seen on COVID-19 disease spread. As an estimator of this TL, we propose a country-specific value relying on cross-correlation and performance criteria of regression models. Based on our results, the effects of restriction actions appear after a median of 14 days. This estimation is in accordance with TLs proposed by Alfano and Ercolano[4] (10 days), and Linka et al.[8] (17 days). Furthermore, our results indicate that the median TL in Europe (12.5 days) is shorter than in the rest of the world (15 days).

If we assume that social distancing works at reducing Rt, the association between mobility data and Rt could function as an indicator of the accuracy of daily case incidence estimates. However, our findings propose that social distancing could have the opposite effect on COVID-19 spread in some countries. Whether or not this paradoxical effect is due to biased mobility data or to residential community spread, we confirmed the observation with the results of the mixed models regarding SI and Rt. Undoubtedly, also factors other than social distancing and restriction measures, such as the non-compliance with hygiene measures[26] or the lack of public trust in their government[27], can affect Rt.

In the absence of a single criterium not contradicting with other performance criteria, we had to select the best model based on plots inspections. This approximate inspection is often used in sensitivity analyses for variable selection, but although we tried to reduce the bias by anonymizing the plots, we recognized the reproducibility limit and the human error susceptibility of this method. Another limitation in our TL estimation is the lack of confidence intervals.

## Conclusion

Mobile phone data could contribute to predictions of the Rt of COVID-19 and to estimations of the TL between social distancing with movement restrictions and Rt, which, according to our results, greatly vary across countries. In general, social distancing and high stringency indices are associated with a reduction in the spread of COVID-19. We recommend authorities to estimate country-specific TLs through a more accurate method, such as using individualized tracking data. The country-specific TL could be particularly practical in revising the commonly enforced 14-day quarantine period for susceptible individuals and travelers returning to their home country.

## Data Availability

Data is publicly available from the COVID-19 Open-Data GitHub Repository (Wahltinez et al. 2020)

https://github.com/GoogleCloudPlatform/covid-19-open-data

## Acknowledgments

Many thanks to Redouane Benazzouz for his comments and support, and to Kerstin Gackle and Caroline Allen for their advice on academic writing.

## References

1. Dong E, Du H, Gardner L. An interactive web-based dashboard to track COVID-19 in real time. Lancet Infect Dis. 2020 May;20(5):533–4.

2. Kuniya T. Evaluation of the effect of the state of emergency for the first wave of COVID-19 in Japan. Infect Dis Model. 2020;5:580–7.

3. Diekmann O, Heesterbeek JA, Metz JA. On the definition and the computation of the basic reproduction ratio R0 in models for infectious diseases in heterogeneous populations. J Math Biol. 1990;28(4):365–82.

4. Alfano V, Ercolano S. The Efficacy of Lockdown Against COVID-19: A Cross-Country Panel Analysis. Appl Health Econ Health Policy. 2020 Aug 1;18(4):509–17.

5. ACAPS. #COVID19 Government Measures Dataset [Internet]. ACAPS. 2020 [cited 2020 Aug 29]. Available from: https://www.acaps.org/covid19-government-measures-dataset.

6. Hale T, Sam Webster, Anna Petherick, Toby Phillips, Beatriz Kira. Oxford COVID-19 Government Response Tracker [Internet]. Blavatnik School of Government; 2020. Available from: http://www.bsg.ox.ac.uk/covidtracker

7. Saha J, Barman B, Chouhan P. Lockdown for COVID-19 and its impact on community mobility in India: An analysis of the COVID-19 Community Mobility Reports, 2020. Child Youth Serv Rev. 2020 Sep;116:105160.

8. Linka K, Peirlinck M, Kuhl E. The reproduction number of COVID-19 and its correlation with public health interventions. Comput Mech. 2020 Jul 28;1–16.

9. Google. COVID-19 Community Mobility Report [Internet]. COVID-19 Community Mobility Report. 2020 [cited 2020 Aug 31]. Available from https://www.google.com/covid19/mobility?hl=en

10. Wahltinez O, Lee M, Erlinger A, Daswani M, Yawalkar P, Murphy K, et al. COVID-19 Open-Data: curating a fine-grained, global-scale data repository for SARS-CoV-2. 2020; Available from: https://github.com/GoogleCloudPlatform/covid-19-open-data

11. Ienca M, Vayena E. On the responsible use of digital data to tackle the COVID-19 pandemic. Nat Med. 2020 Mar 27;1–2.

12. Cori A, Ferguson NM, Fraser C, Cauchemez S. A New Framework and Software to Estimate Time-Varying Reproduction Numbers During Epidemics. Am J Epidemiol. 2013 Nov 1;178(9):1505–12.

13. Swerdlow DL, Finelli L. Preparation for Possible Sustained Transmission of 2019 Novel Coronavirus: Lessons From Previous Epidemics. JAMA. 2020 Mar 24;323(12):1129–30.

14. Nishiura H, Linton NM, Akhmetzhanov AR. Serial interval of novel coronavirus (COVID-19) infections. Int J Infect Dis. 2020 Apr;93:284–6.

15. Vokó Z, Pitter JG. The effect of social distance measures on COVID-19 epidemics in Europe: an interrupted time series analysis. GeroScience. 2020 Jun 11;1–8.

16. R Core Team. R: A Language and Environment for Statistical Computing [Internet]. Vienna, Austria: R Foundation for Statistical Computing; 2020. Available from:https://www.R-project.org

17. Guan W, Ni Z, Hu Y, Liang W, Ou C, He J, et al. Clinical Characteristics of Coronavirus Disease 2019 in China. N Engl J Med. 2020 Apr 30;382(18):1708–20.

18. Buishand TA. Some methods for testing the homogeneity of rainfall records. J Hydrol. 1982 Aug 1;58(1):11–27.

19. Taylor SJ, Letham B. Forecasting at scale [Internet]. PeerJ Inc.; 2017 Sep [cited 2020 Sep 6]. Report No.: e3190v2. Available from: https://peerj.com/preprints/3190

20. Venables WN, Ripley BD. Modern Applied Statistics with S [Internet]. 4th ed. New York: Springer-Verlag; 2002 [cited 2020 Sep 20]. (Statistics and Computing, Statistics, Computing Venables, W.N.:Statistics w.S- PLUS). Available from: https://www.springer.com/gp/book/9780387954578.

21. Bates D, Maechler M, Bolker[aut B cre, Walker S, Christensen RHB, et al. lme4: Linear Mixed-Effects Models using “Eigen” and S4 [Internet]. 2020 [cited 2020 Sep 7]. Available from: https://CRAN.R-project.org/package=lme4

22. Singmann H, Bolker B, Westfall J, Aust F, Ben-Shachar MS, Højsgaard S, et al. afex: Analysis of Factorial Experiments [Internet]. 2020 [cited 2020 Sep 7]. Available from: https://CRAN.R-project.org/package=afex

23. Lüdecke D, Makowski D, Patil I, Waggoner P. insight: Easy Access to Model Information for Various Model Objects [Internet]. 2020 [cited 2020 Sep 7]. Available from: https://CRAN.R-project.org/package=insight

24. Funder DC, Ozer DJ. Evaluating Effect Size in Psychological Research: Sense and Nonsense. Adv Methods Pract Psychol Sci. 2019 Jun 1;2(2):156–68.

25. Cartenì A, Francesco LD, Martino M. How mobility habits influenced the spread of the COVID-19 pandemic: Results from the Italian case study. Sci Total Environ. 2020 Nov 1;741:140489.

26. Nivette A, Ribeaud D, Murray A, Steinhoff A, Bechtiger L, Hepp U, et al. Non-compliance with COVID-19-related public health measures among young adults in Switzerland: Insights from a longitudinal cohort study. Soc Sci Med 1982. 2020 Sep 16;268:113370.

27. Elgar FJ, Stefaniak A, Wohl MJA. The trouble with trust: Time-series analysis of social capital, income inequality, and COVID-19 deaths in 84 countries. Soc Sci Med 1982. 2020 Sep 16;113365.

