## Supplementary Material for "Social Distancing with Movement Restrictions and the Effective Replication Number of COVID-19: Multi-Country Analysis Based on Phone Mobility Data"

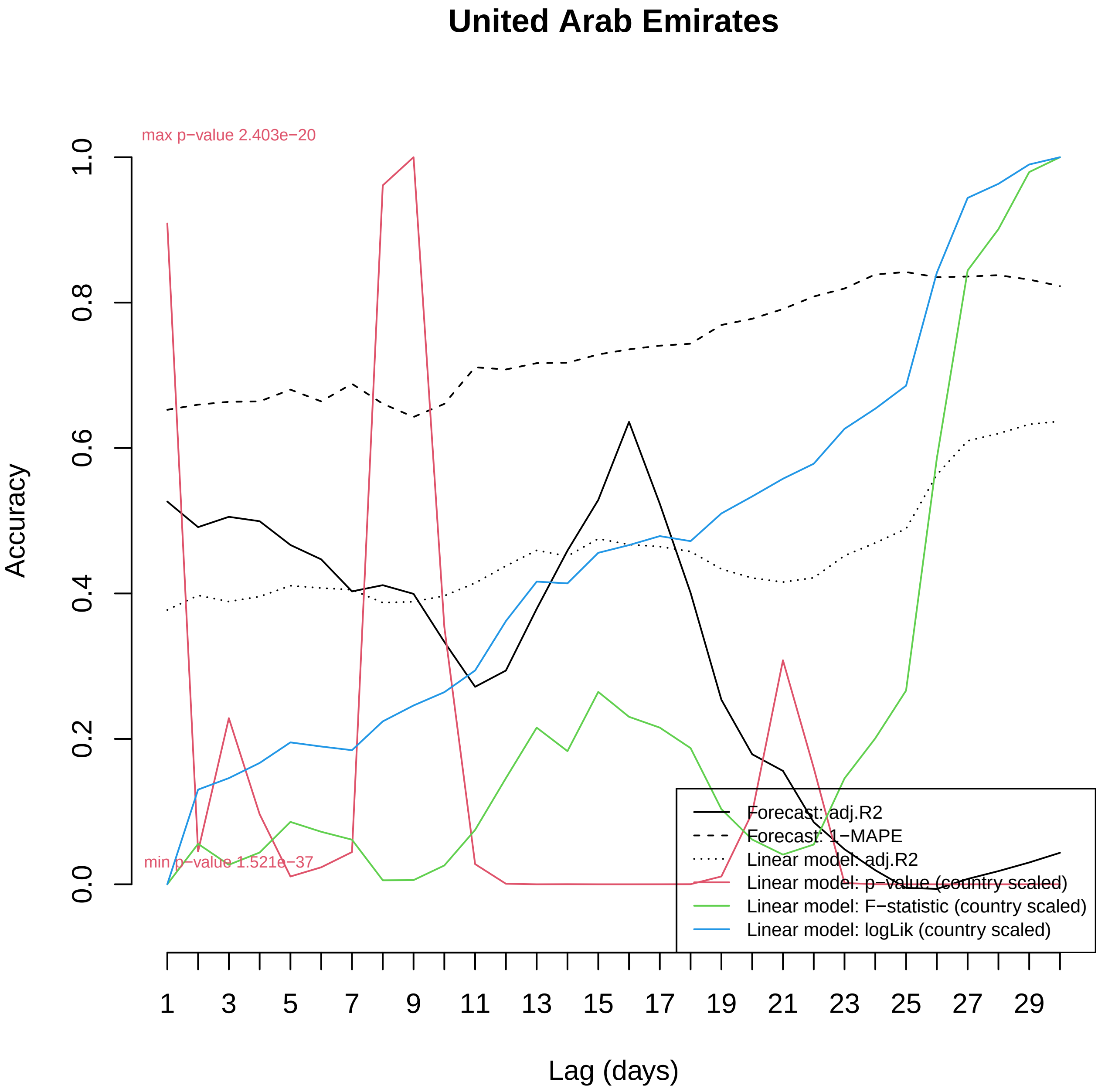

United Arab Emirates
max p−value 2.403e−20
min p−value 1.521e−37
Accuracy
0.0
0.2
0.4
0.6
0.8
1.0
Lag (days)
1
3
5
7
9
11
13
15
17
19
21
23
25
27
29
Forecast: adj.R2
Forecast: 1−MAPE
Linear model: adj.R2
Linear model: p−value (country scaled)
Linear model: F−statistic (country scaled)
Linear model: logLik (country scaled)

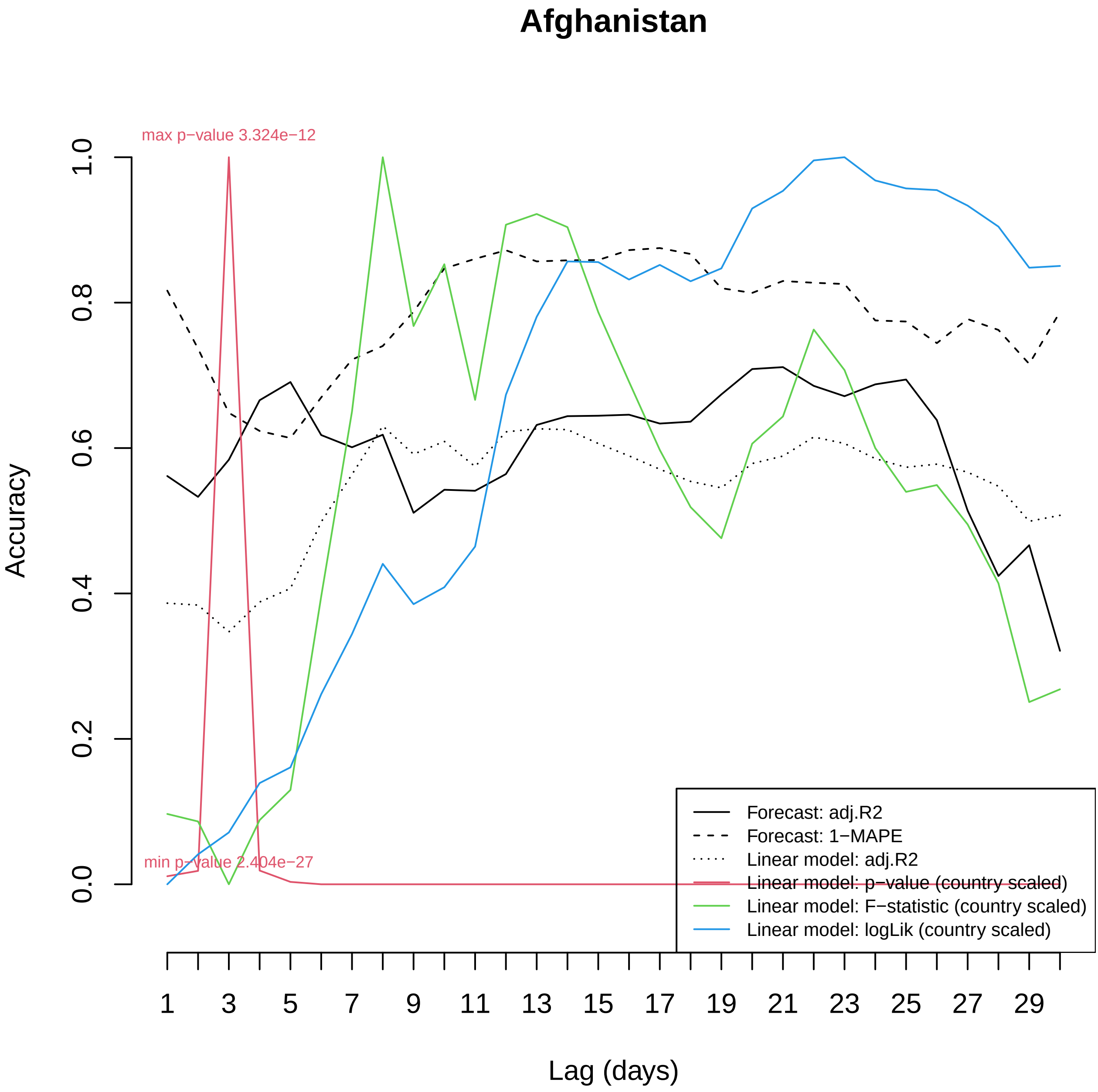
Afghanistan
max p−value 3.324e−12
min p−value 2.404e−27
Accuracy
Lag (days)
Forecast: adj.R2
Forecast: 1−MAPE
Linear model: adj.R2
Linear model: p−value (country scaled)
Linear model: F−statistic (country scaled)
Linear model: logLik (country scaled)

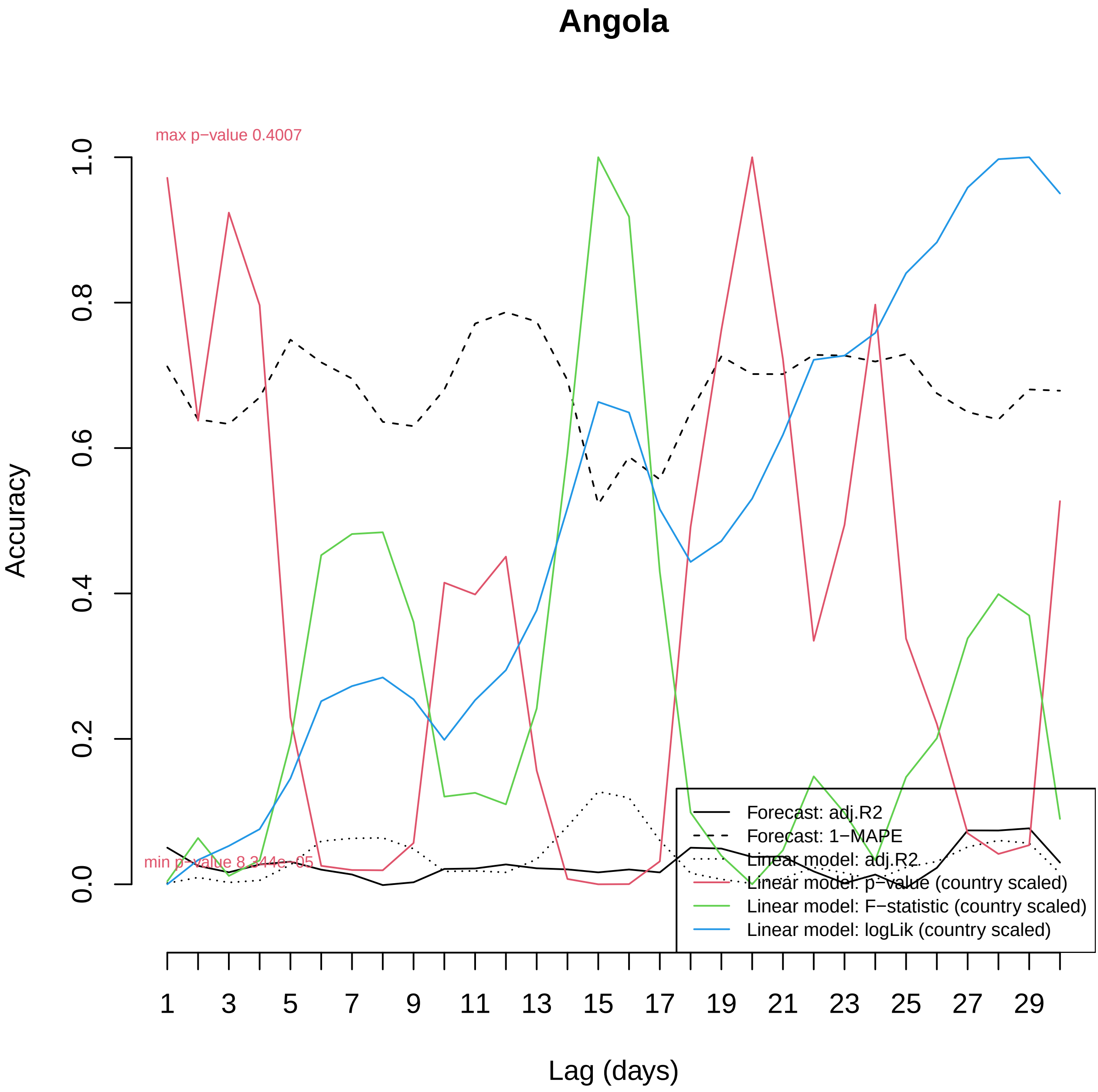

Angola
max p−value 0.4007
min p−value 8.344e−05
Accuracy
0.0
0.2
0.4
0.6
0.8
1.0
1
3
5
7
9
11
13
15
17
19
21
23
25
27
29
Lag (days)
Forecast: adj.R2
Forecast: 1−MAPE
Linear model: adj.R2
Linear model: p−value (country scaled)
Linear model: F−statistic (country scaled)
Linear model: logLik (country scaled)

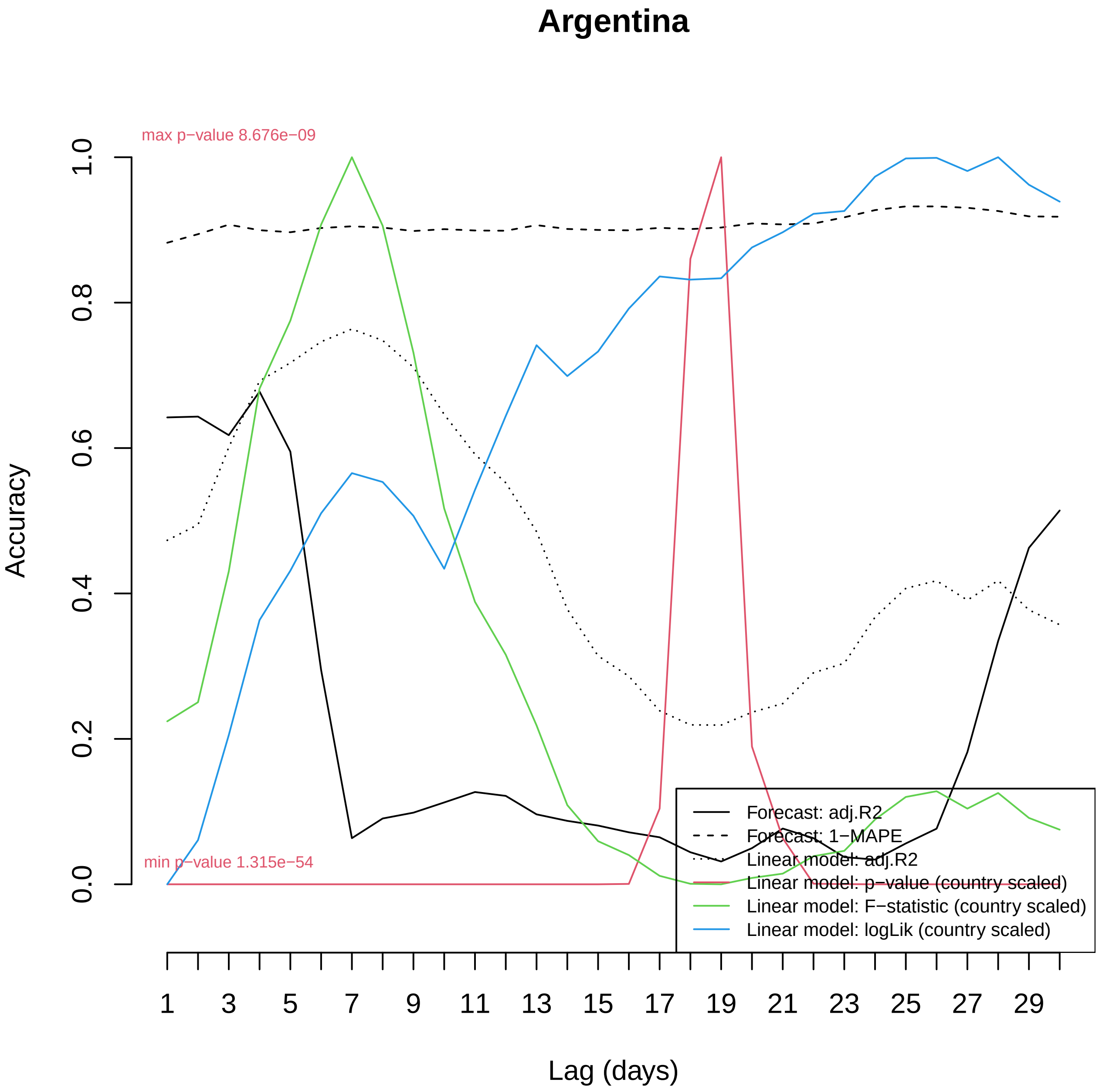
Argentina
max p−value 8.676e−09
min p−value 1.315e−54
Accuracy
0.0
0.2
0.4
0.6
0.8
1.0
Lag (days)
1
3
5
7
9
11
13
15
17
19
21
23
25
27
29
Forecast: adj.R2
Forecast: 1−MAPE
Linear model: adj.R2
Linear model: p−value (country scaled)
Linear model: F−statistic (country scaled)
Linear model: logLik (country scaled)

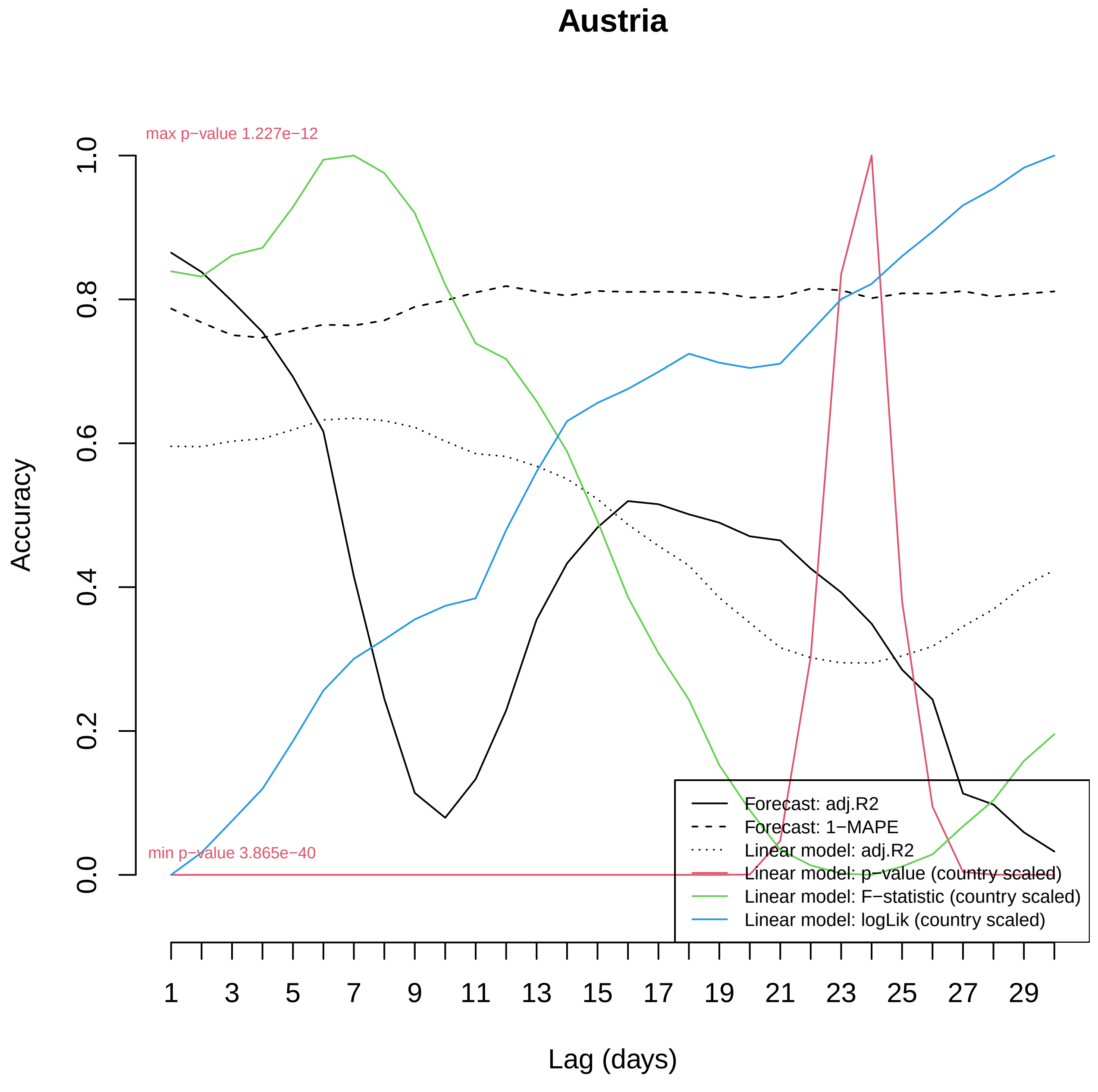

Austria
max p−value 1.227e−12
min p−value 3.865e−40
Accuracy
0.0
0.2
0.4
0.6
0.8
1.0
1
3
5
7
9
11
13
15
17
19
21
23
25
27
29
Lag (days)
Forecast: adj.R2
Forecast: 1−MAPE
Linear model: adj.R2
Linear model: p−value (country scaled)
Linear model: F−statistic (country scaled)
Linear model: logLik (country scaled)

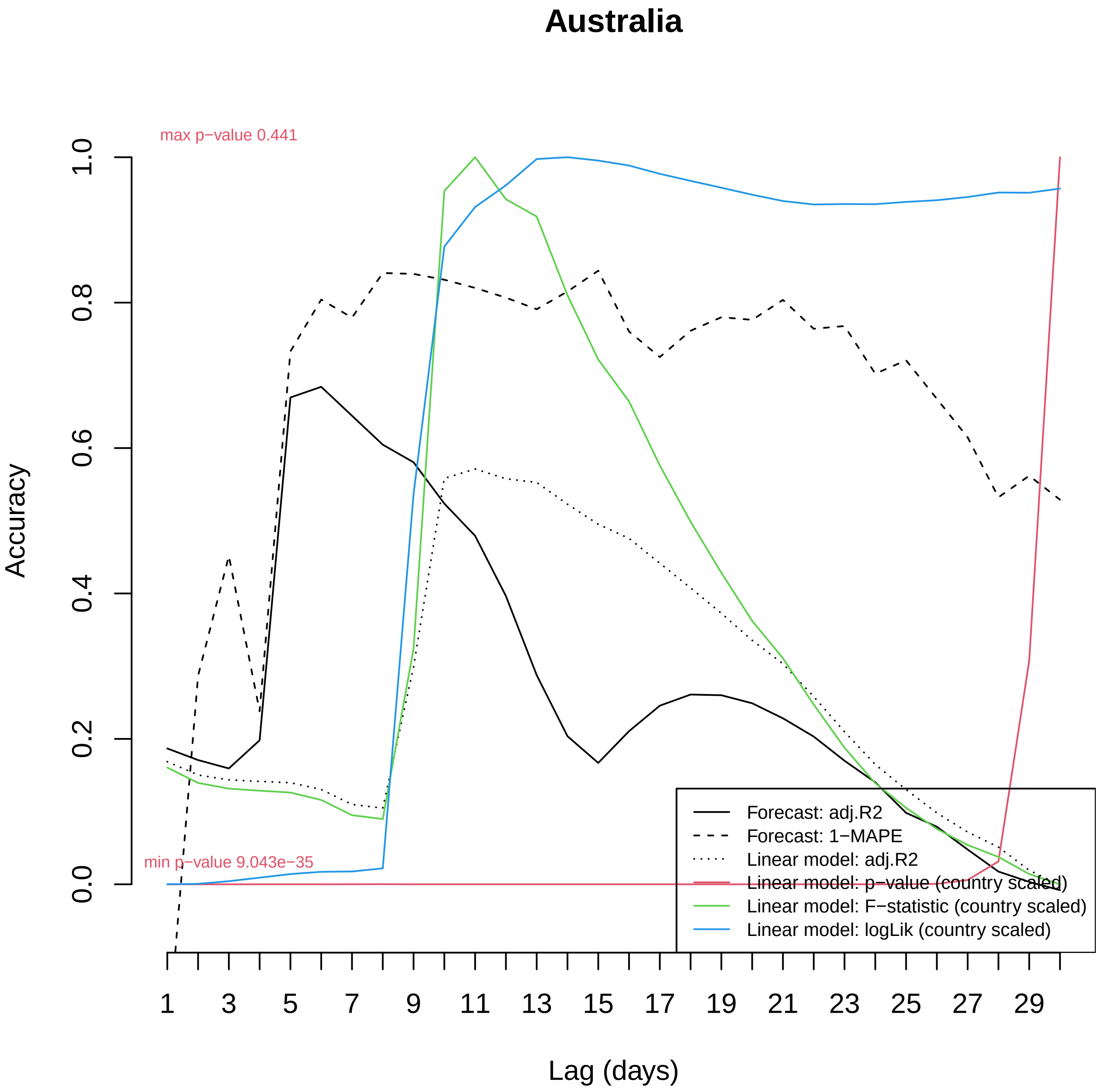

Australia
max p−value 0.441
min p−value 9.043e−35
Accuracy
0.0
0.2
0.4
0.6
0.8
1.0
Lag (days)
1
3
5
7
9
11
13
15
17
19
21
23
25
27
29
Forecast: adj.R2
Forecast: 1−MAPE
Linear model: adj.R2
Linear model: p−value (country scaled)
Linear model: F−statistic (country scaled)
Linear model: logLik (country scaled)

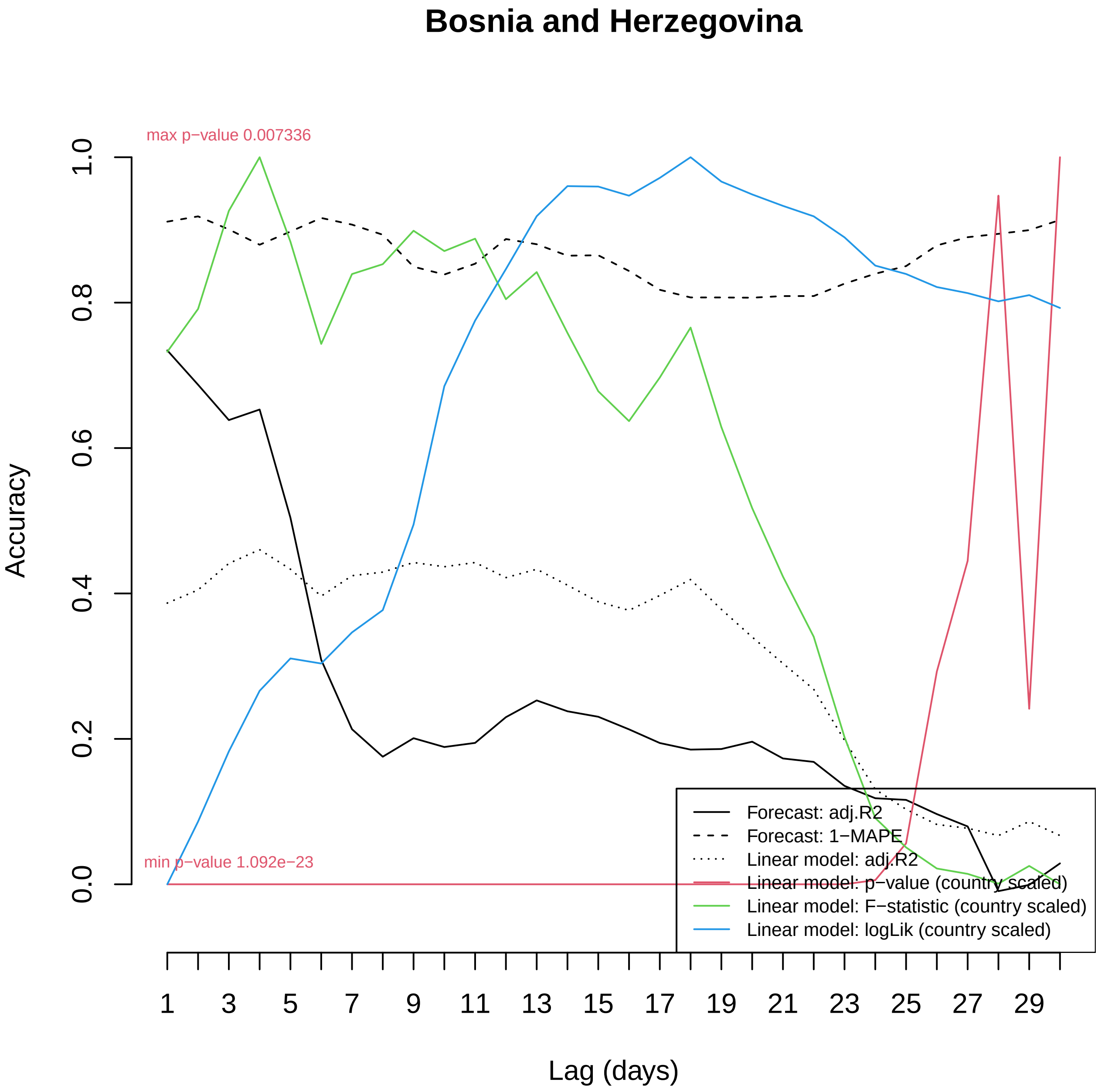
Bosnia and Herzegovina
max p−value 0.007336
min p−value 1.092e−23
Accuracy
0.0
0.2
0.4
0.6
0.8
1.0
1
3
5
7
9
11
13
15
17
19
21
23
25
27
29
Lag (days)
Forecast: adj.R2
Forecast: 1−MAPE
Linear model: adj.R2
Linear model: p−value (country scaled)
Linear model: F−statistic (country scaled)
Linear model: logLik (country scaled)

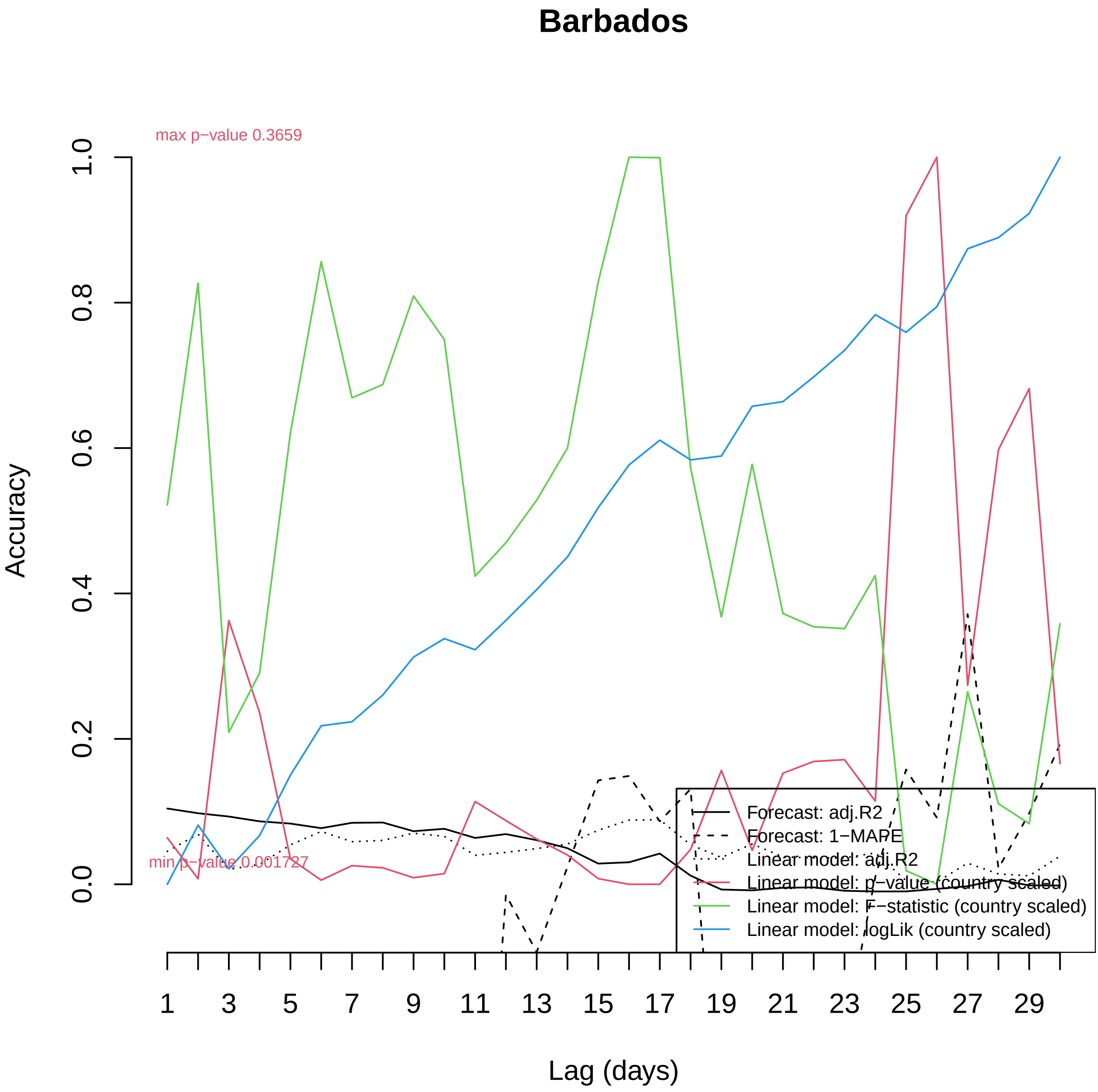
Barbados
max p−value 0.3659
min p−value 0.001727
Accuracy
0.0
0.2
0.4
0.6
0.8
1.0
1
3
5
7
9
11
13
15
17
19
21
23
25
27
29
Lag (days)
Forecast: adj.R2
Forecast: 1−MAPE
Linear model: adj.R2
Linear model: p−value (country scaled)
Linear model: F−statistic (country scaled)
Linear model: logLik (country scaled)

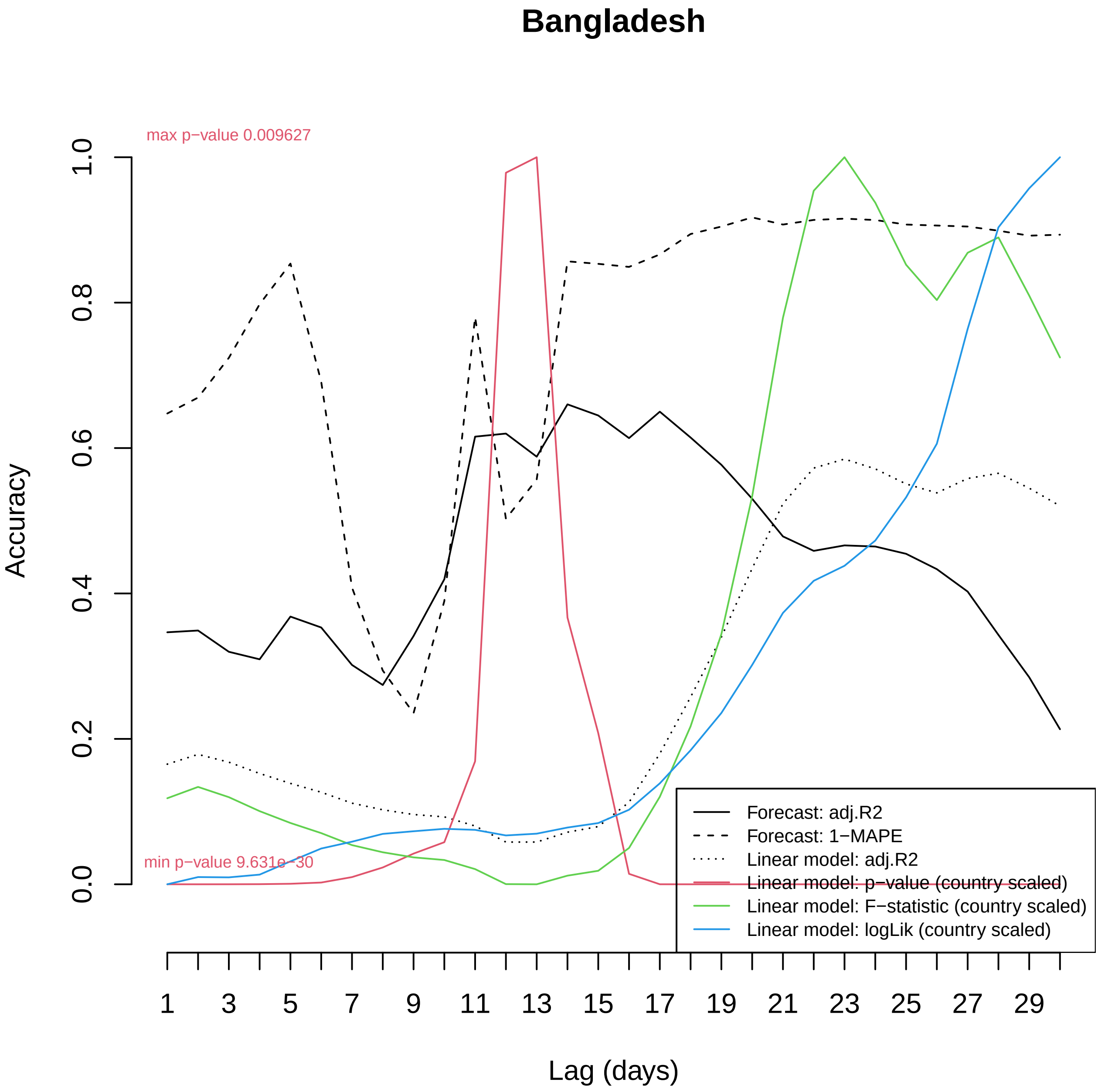

Bangladesh
max p−value 0.009627
min p−value 9.631e−30
Accuracy
0.0
0.2
0.4
0.6
0.8
1.0
Lag (days)
1
3
5
7
9
11
13
15
17
19
21
23
25
27
29
Forecast: adj.R2
Forecast: 1−MAPE
Linear model: adj.R2
Linear model: p−value (country scaled)
Linear model: F−statistic (country scaled)
Linear model: logLik (country scaled)

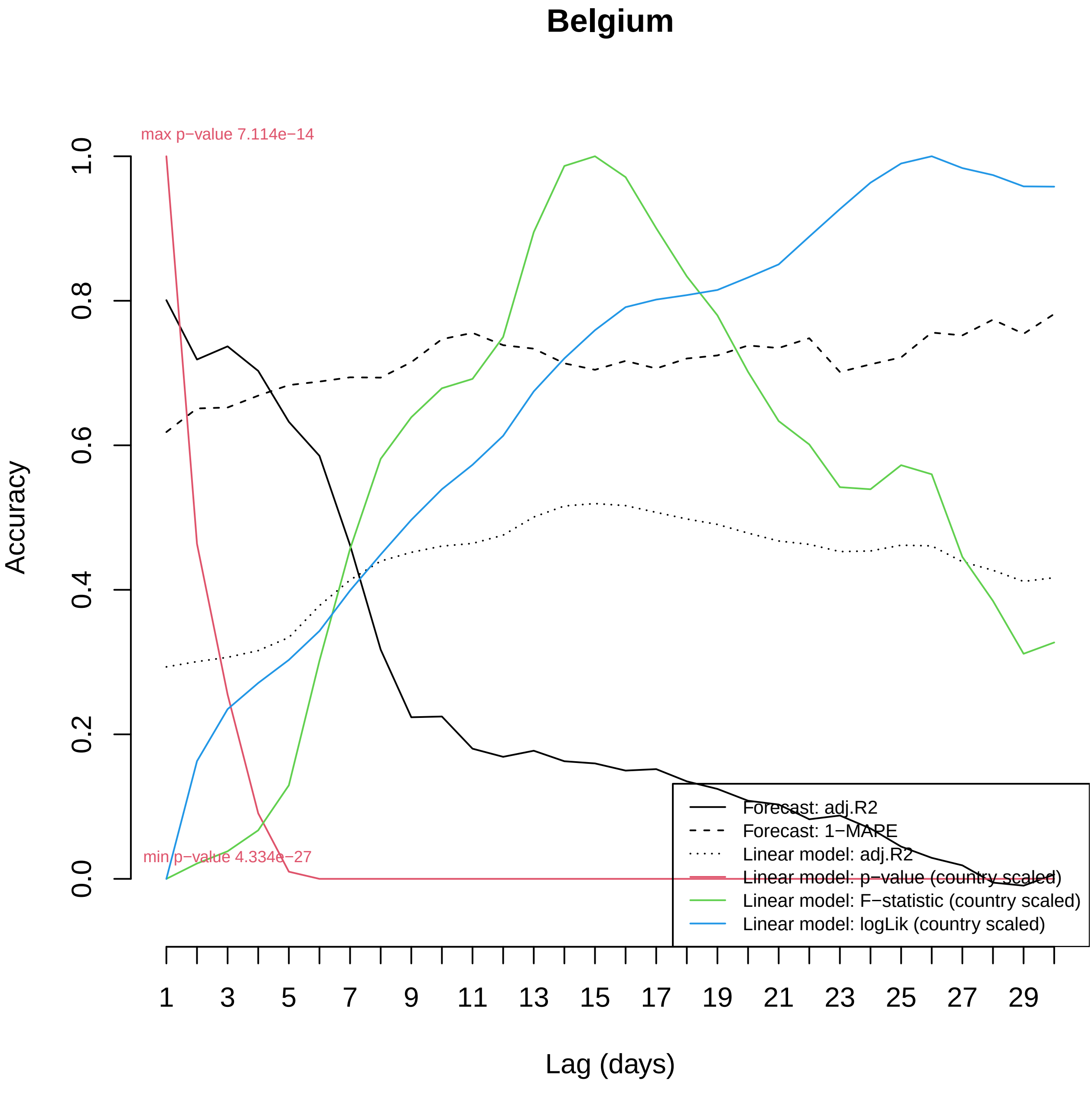
Belgium
max p−value 7.114e−14
min p−value 4.334e−27
Accuracy
0.0
0.2
0.4
0.6
0.8
1.0
Lag (days)
1
3
5
7
9
11
13
15
17
19
21
23
25
27
29
Forecast: adj.R2
Forecast: 1−MAPE
Linear model: adj.R2
Linear model: p−value (country scaled)
Linear model: F−statistic (country scaled)
Linear model: logLik (country scaled)

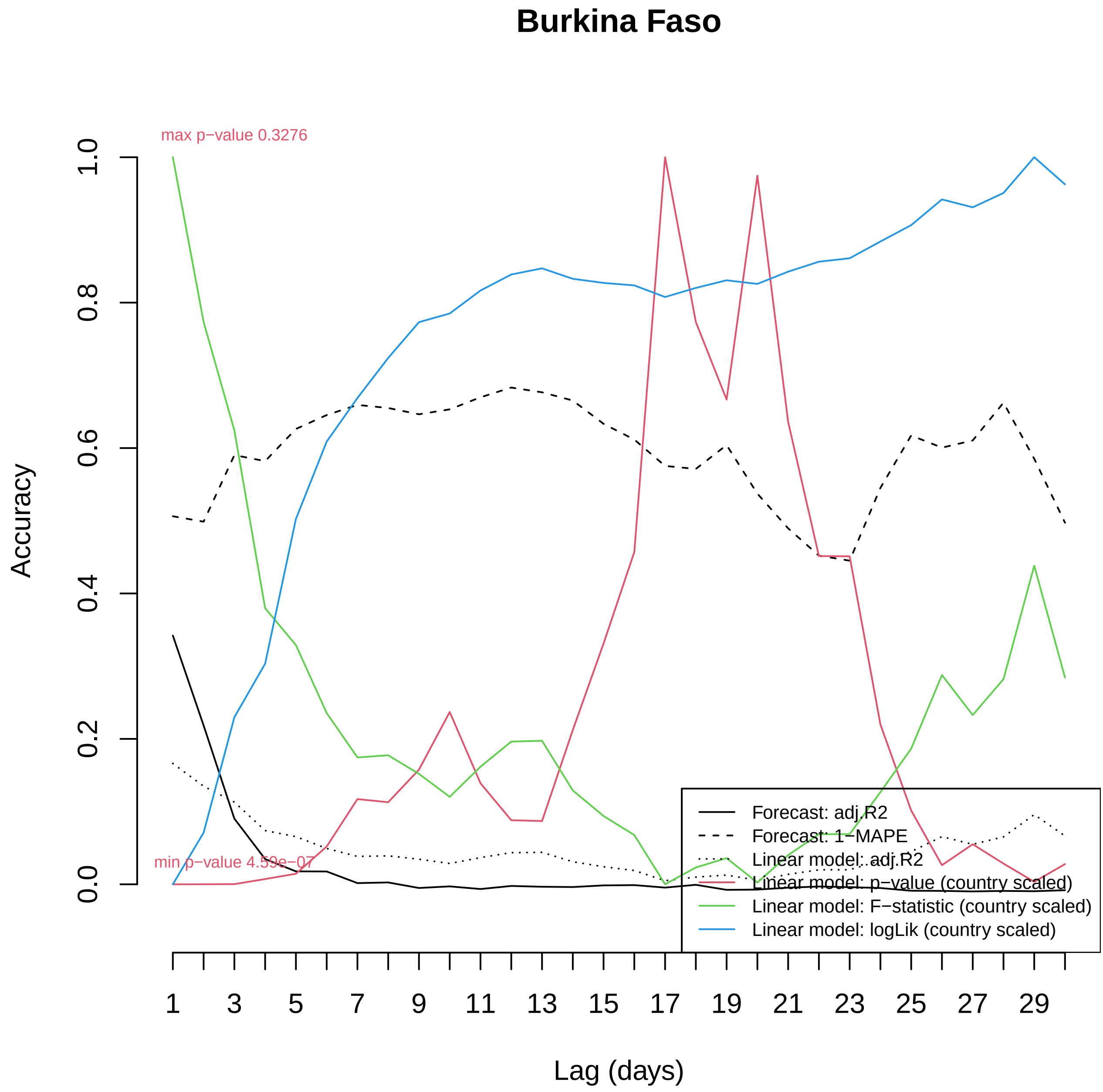

Burkina Faso
max p−value 0.3276
min p−value 4.59e−07
Accuracy
1.0
0.8
0.6
0.4
0.2
0.0
Lag (days)
1
3
5
7
9
11
13
15
17
19
21
23
25
27
29
Forecast: adj.R2
Forecast: 1−MAPE
Linear model: adj.R2
Linear model: p−value (country scaled)
Linear model: F−statistic (country scaled)
Linear model: logLik (country scaled)

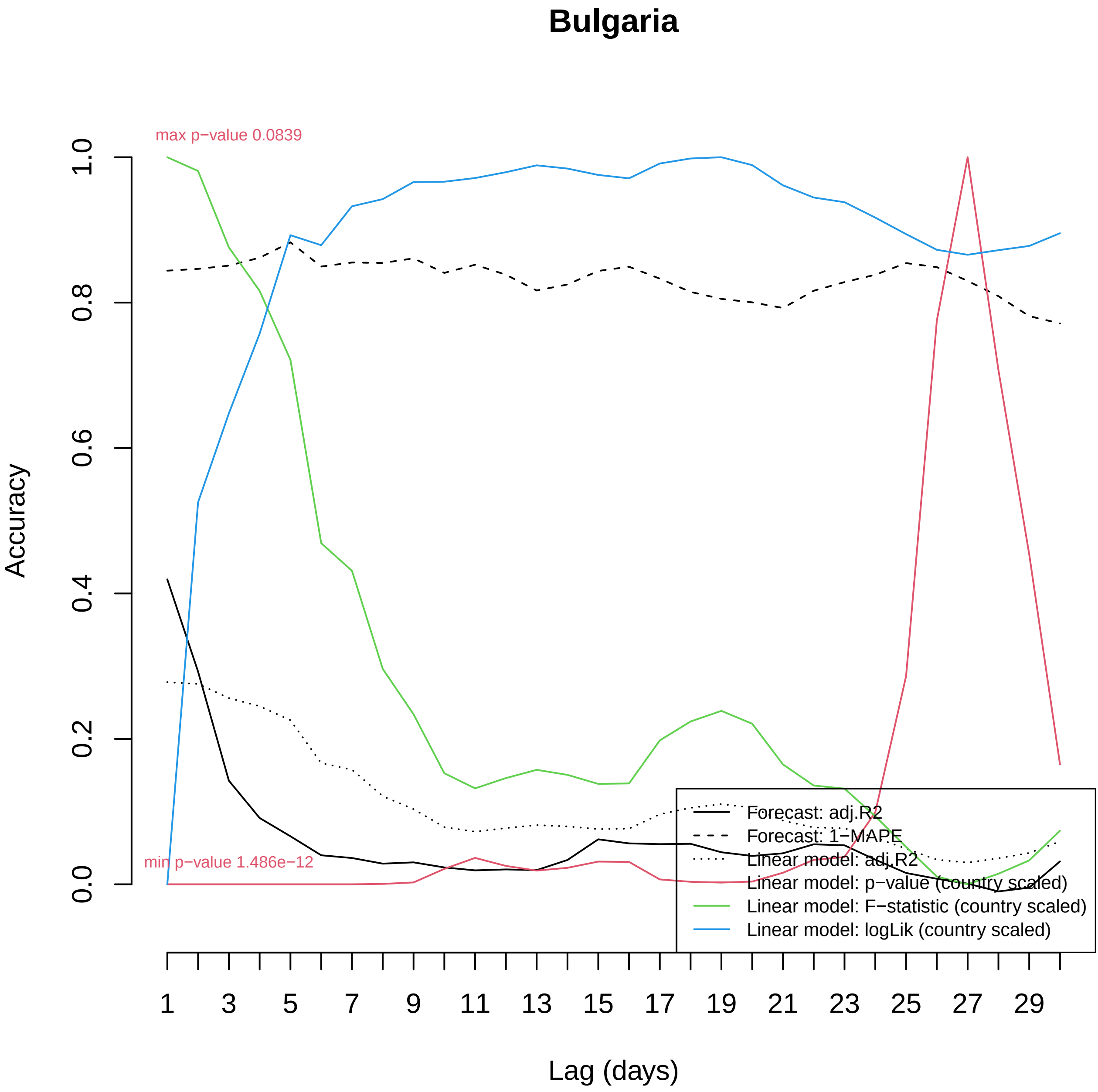

Bulgaria
max p−value 0.0839
min p−value 1.486e−12
Accuracy
0.0
0.2
0.4
0.6
0.8
1.0
Lag (days)
1
3
5
7
9
11
13
15
17
19
21
23
25
27
29
Forecast: adj.R2
Forecast: 1−MAPE
Linear model: adj.R2
Linear model: p−value (country scaled)
Linear model: F−statistic (country scaled)
Linear model: logLik (country scaled)

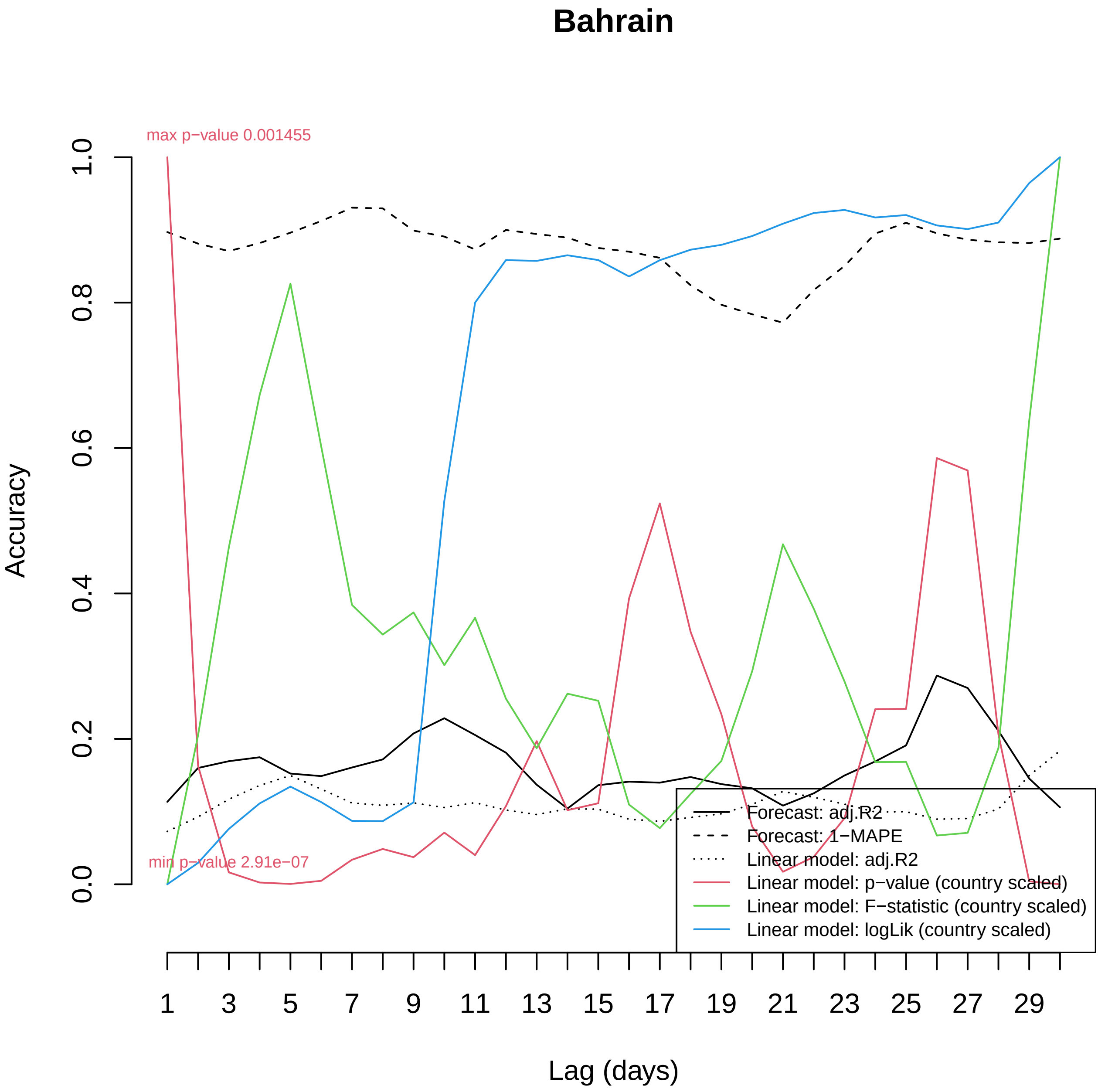
Bahrain
max p−value 0.001455
min p−value 2.91e−07
Accuracy
0.0
0.2
0.4
0.6
0.8
1.0
Lag (days)
1
3
5
7
9
11
13
15
17
19
21
23
25
27
29
Forecast: adj.R2
Forecast: 1−MAPE
Linear model: adj.R2
Linear model: p−value (country scaled)
Linear model: F−statistic (country scaled)
Linear model: logLik (country scaled)

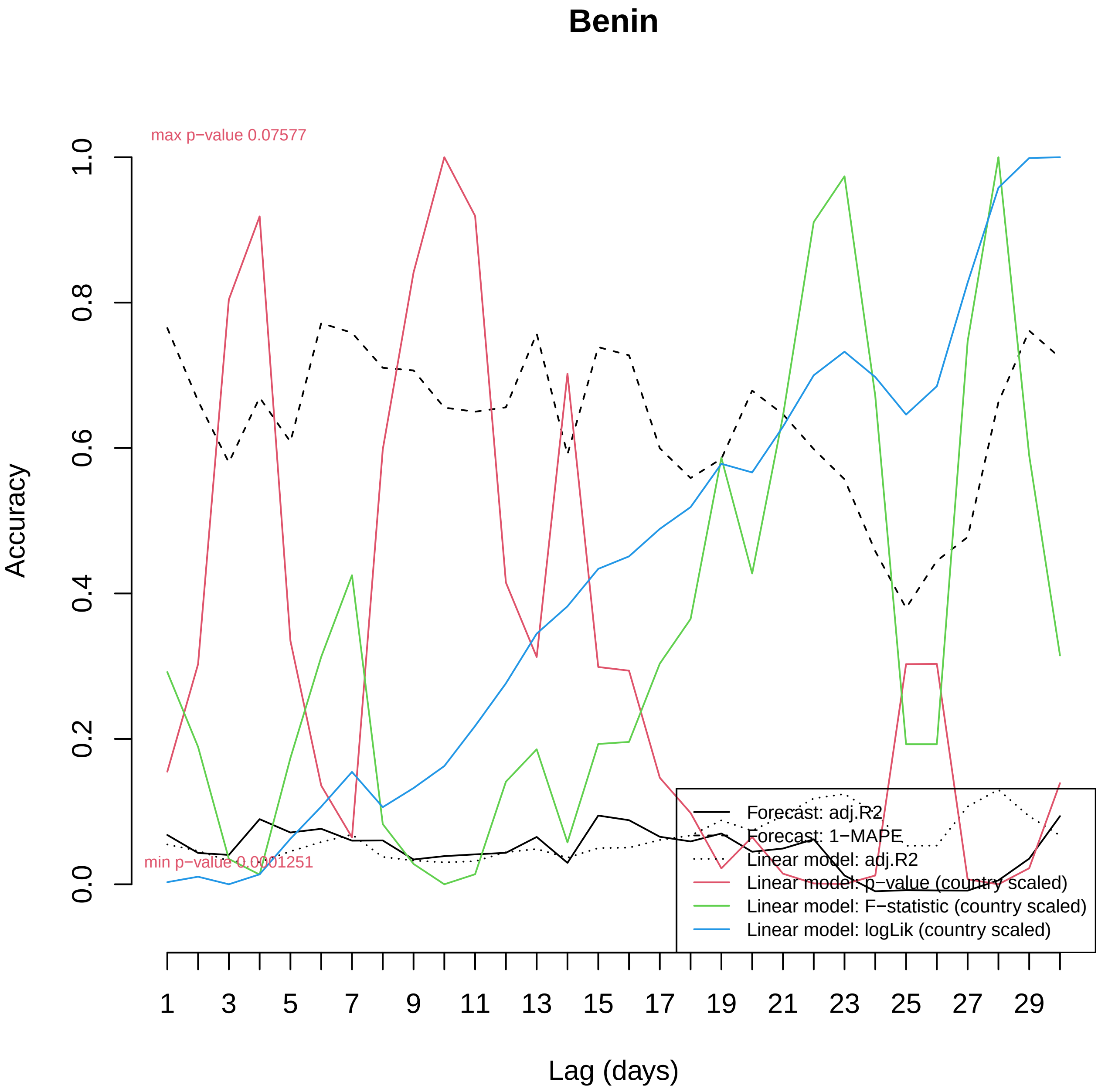

Benin
max p−value 0.07577
min p−value 0.0001251
Accuracy
0.0
0.2
0.4
0.6
0.8
1.0
Lag (days)
1
3
5
7
9
11
13
15
17
19
21
23
25
27
29
Forecast: adj.R2
Forecast: 1−MAPE
Linear model: adj.R2
Linear model: p−value (country scaled)
Linear model: F−statistic (country scaled)
Linear model: logLik (country scaled)

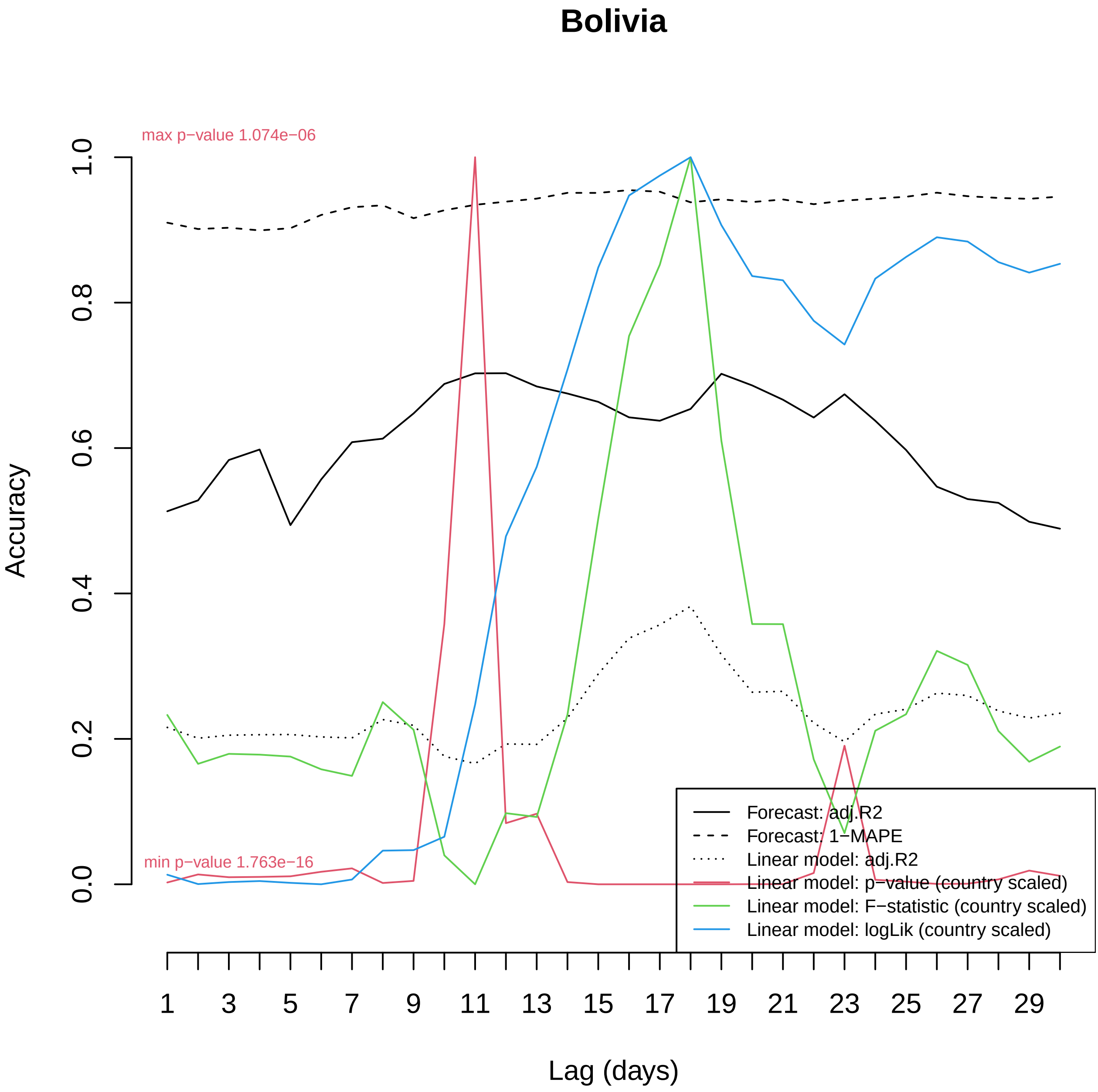
Bolivia
max p−value 1.074e−06
min p−value 1.763e−16
Accuracy
0.0
0.2
0.4
0.6
0.8
1.0
Lag (days)
1
3
5
7
9
11
13
15
17
19
21
23
25
27
29
Forecast: adj.R2
Forecast: 1−MAPE
Linear model: adj.R2
Linear model: p−value (country scaled)
Linear model: F−statistic (country scaled)
Linear model: logLik (country scaled)

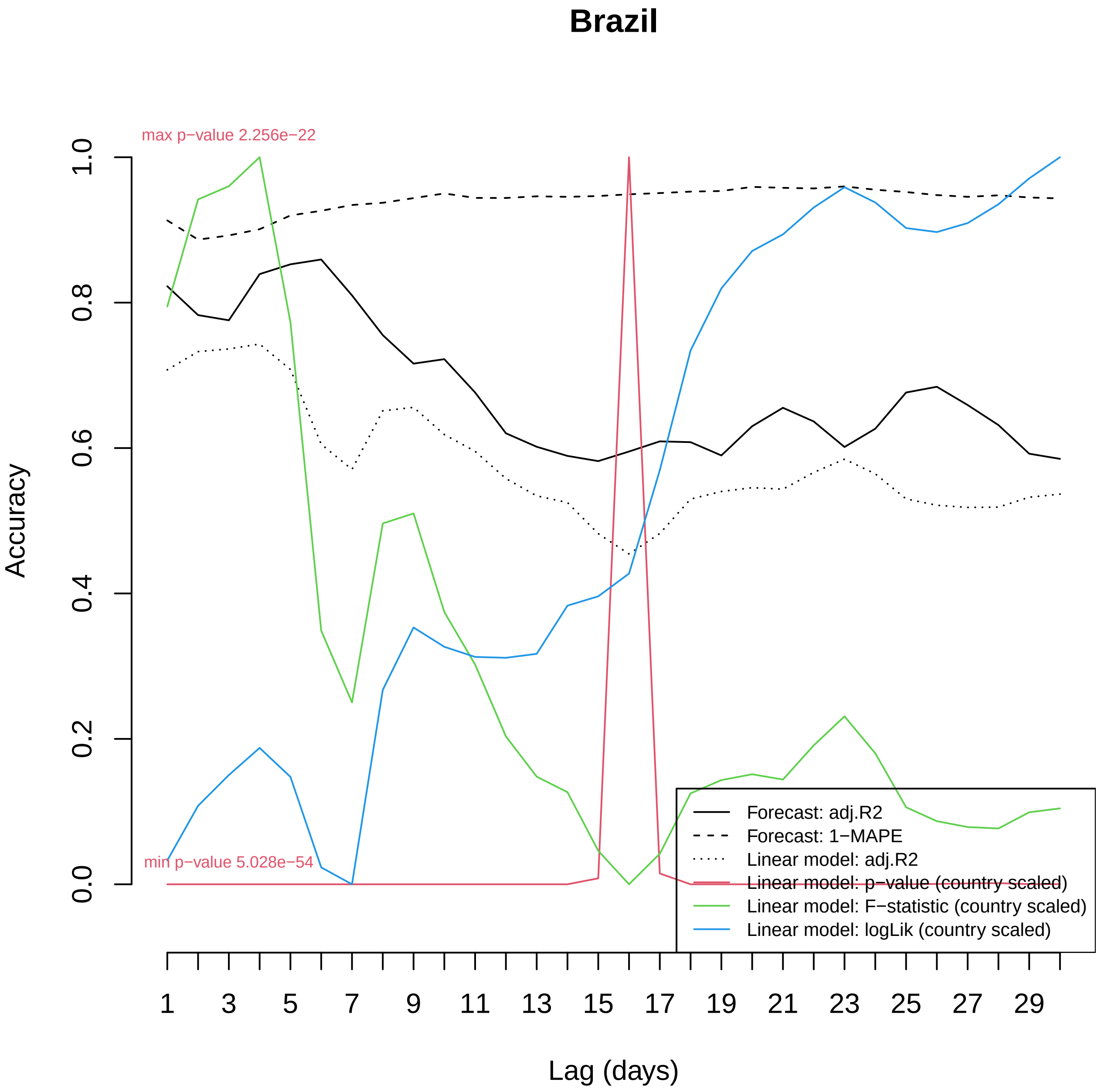

Brazil
max p−value 2.256e−22
min p−value 5.028e−54
Accuracy
0.0
0.2
0.4
0.6
0.8
1.0
Lag (days)
1
3
5
7
9
11
13
15
17
19
21
23
25
27
29
Forecast: adj.R2
Forecast: 1−MAPE
Linear model: adj.R2
Linear model: p−value (country scaled)
Linear model: F−statistic (country scaled)
Linear model: logLik (country scaled)

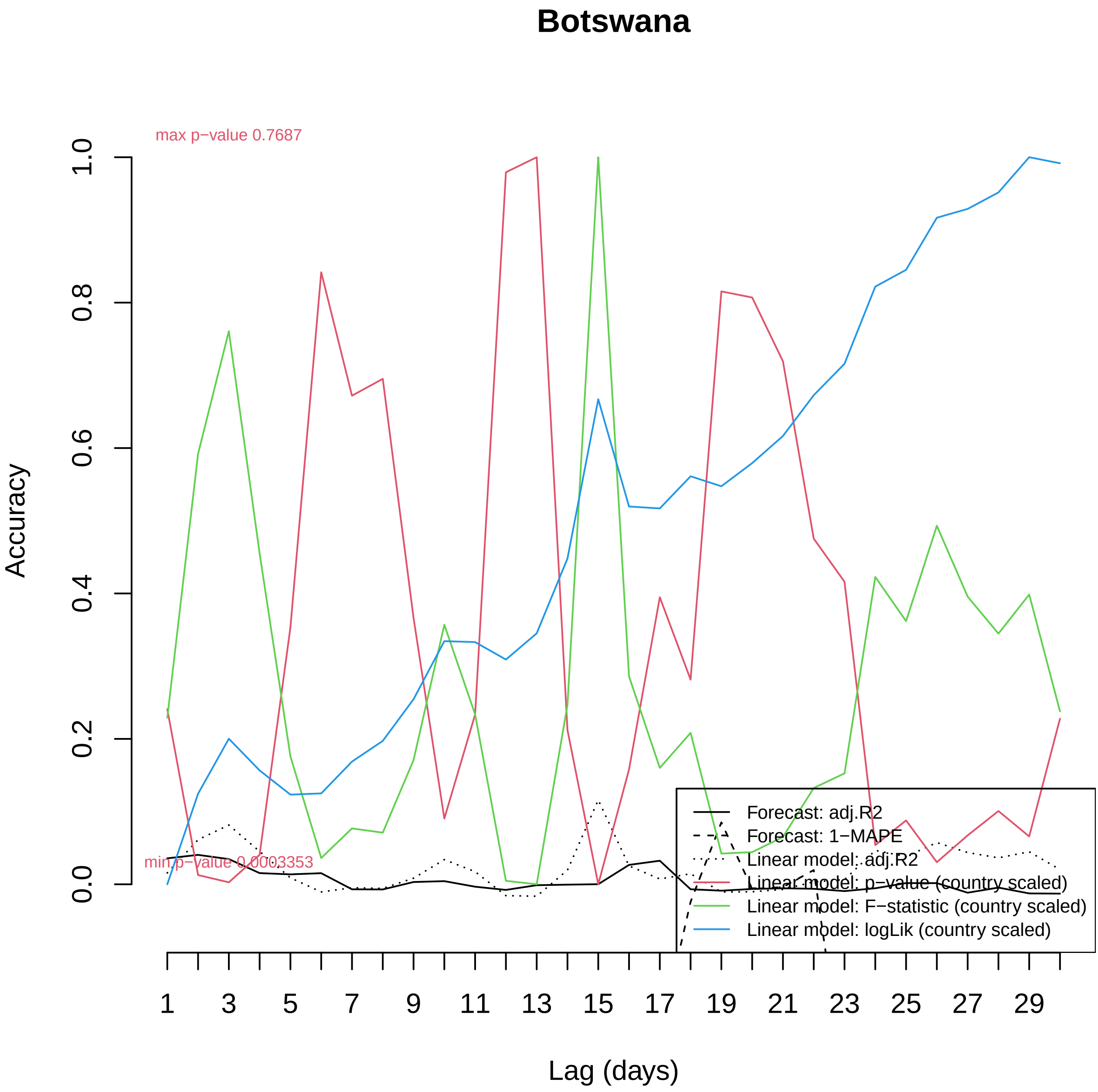

Botswana
max p−value 0.7687
min p−value 0.0003353
Accuracy
0.0
0.2
0.4
0.6
0.8
1.0
Lag (days)
1
3
5
7
9
11
13
15
17
19
21
23
25
27
29
Forecast: adj.R2
Forecast: 1−MAPE
Linear model: adj.R2
Linear model: p−value (country scaled)
Linear model: F−statistic (country scaled)
Linear model: logLik (country scaled)

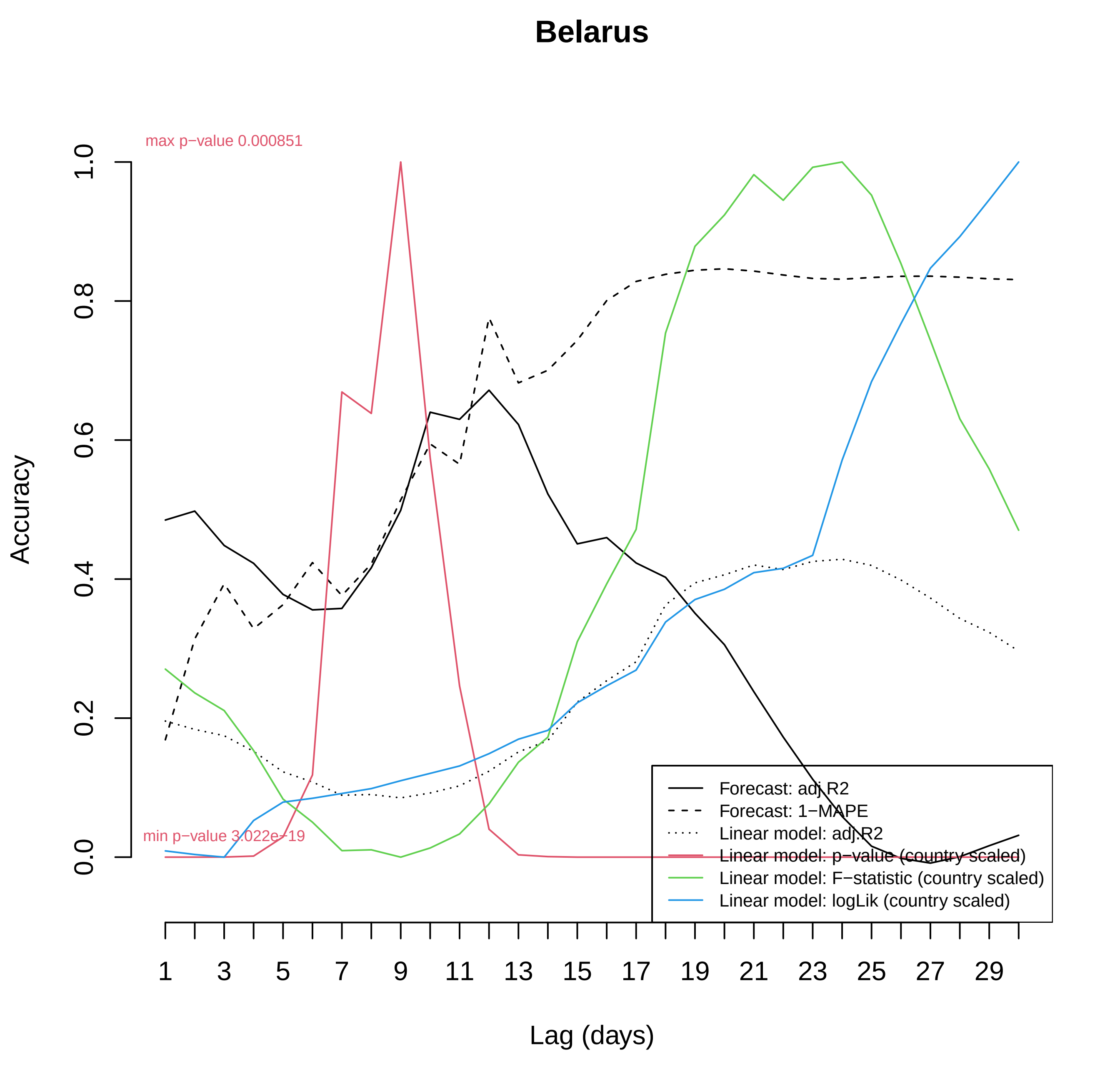

Belarus
max p−value 0.000851
min p−value 3.022e−19
Accuracy
0.0
0.2
0.4
0.6
0.8
1.0
1
3
5
7
9
11
13
15
17
19
21
23
25
27
29
Lag (days)
Forecast: adj.R2
Forecast: 1−MAPE
Linear model: adj.R2
Linear model: p−value (country scaled)
Linear model: F−statistic (country scaled)
Linear model: logLik (country scaled)

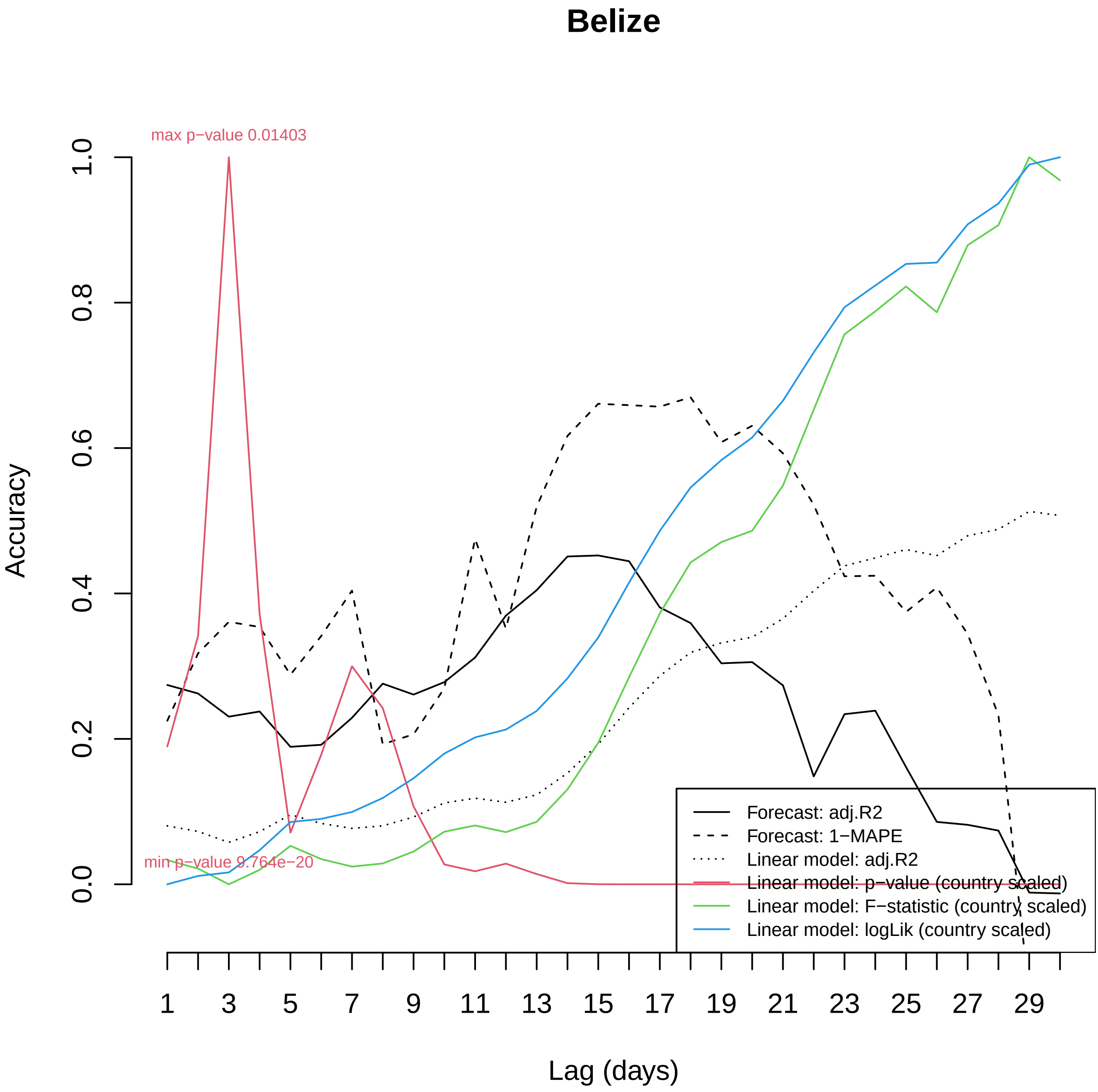

Belize
max p−value 0.01403
min p−value 9.764e−20
Accuracy
0.0
0.2
0.4
0.6
0.8
1.0
Lag (days)
1
3
5
7
9
11
13
15
17
19
21
23
25
27
29
Forecast: adj.R2
Forecast: 1−MAPE
Linear model: adj.R2
Linear model: p−value (country scaled)
Linear model: F−statistic (country scaled)
Linear model: logLik (country scaled)

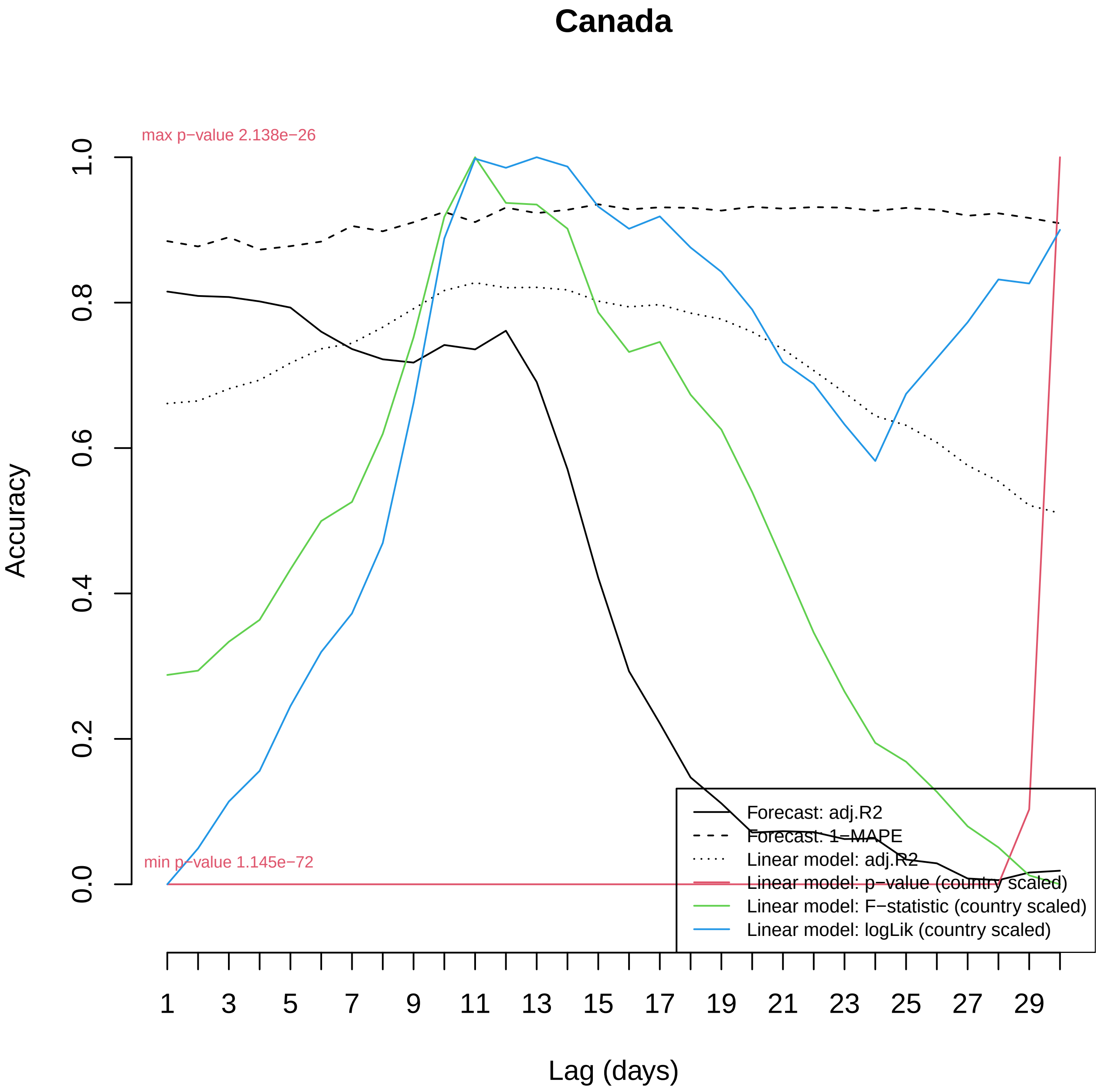
Canada
max p−value 2.138e−26
min p−value 1.145e−72
Accuracy
0.0
0.2
0.4
0.6
0.8
1.0
1
3
5
7
9
11
13
15
17
19
21
23
25
27
29
Lag (days)
Forecast: adj.R2
Forecast: 1−MAPE
Linear model: adj.R2
Linear model: p−value (country scaled)
Linear model: F−statistic (country scaled)
Linear model: logLik (country scaled)

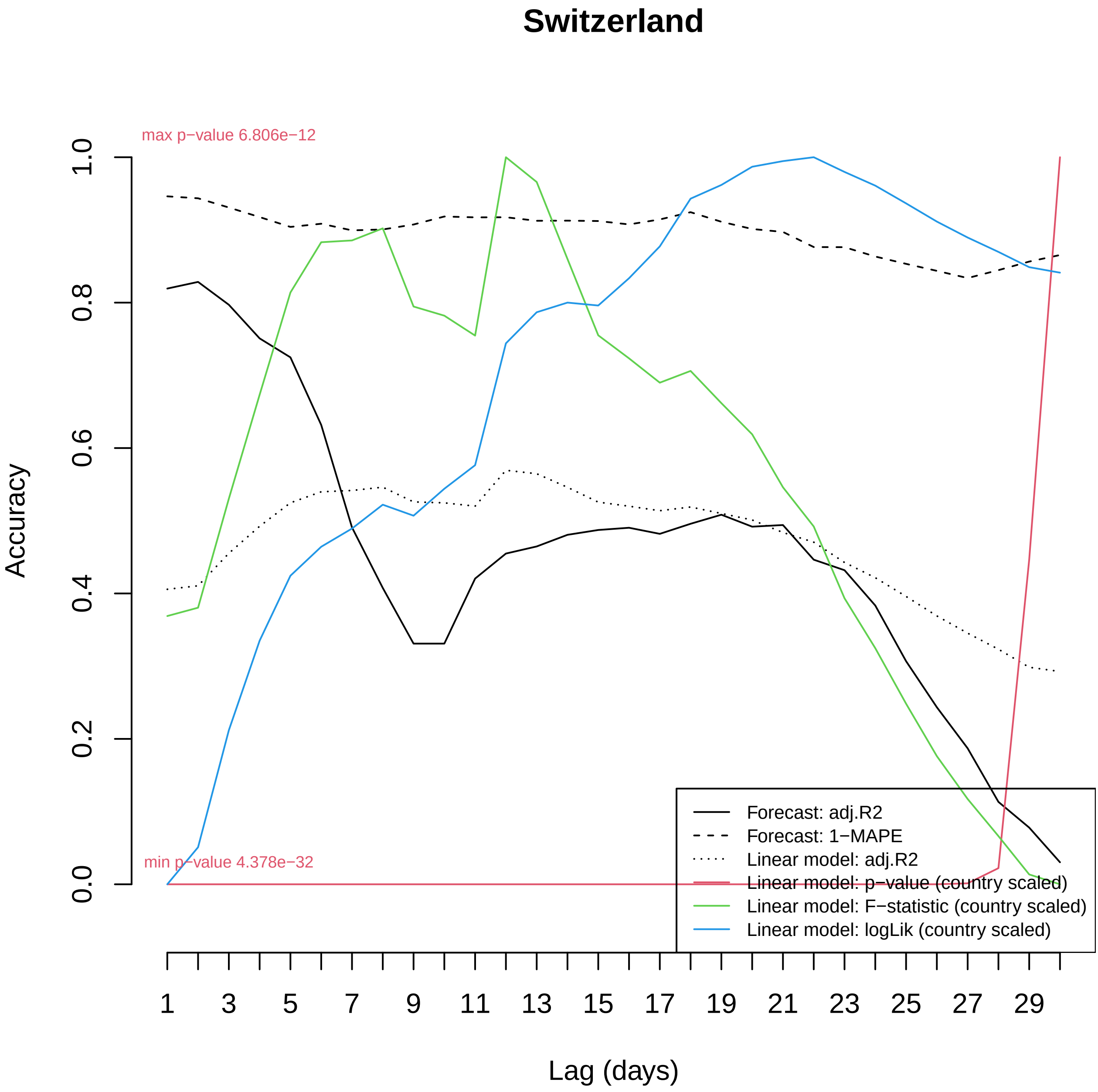
Switzerland
max p−value 6.806e−12
min p−value 4.378e−32
Accuracy
1.0
0.8
0.6
0.4
0.2
0.0
1
3
5
7
9
11
13
15
17
19
21
23
25
27
29
Lag (days)
Forecast: adj.R2
Forecast: 1−MAPE
Linear model: adj.R2
Linear model: p−value (country scaled)
Linear model: F−statistic (country scaled)
Linear model: logLik (country scaled)

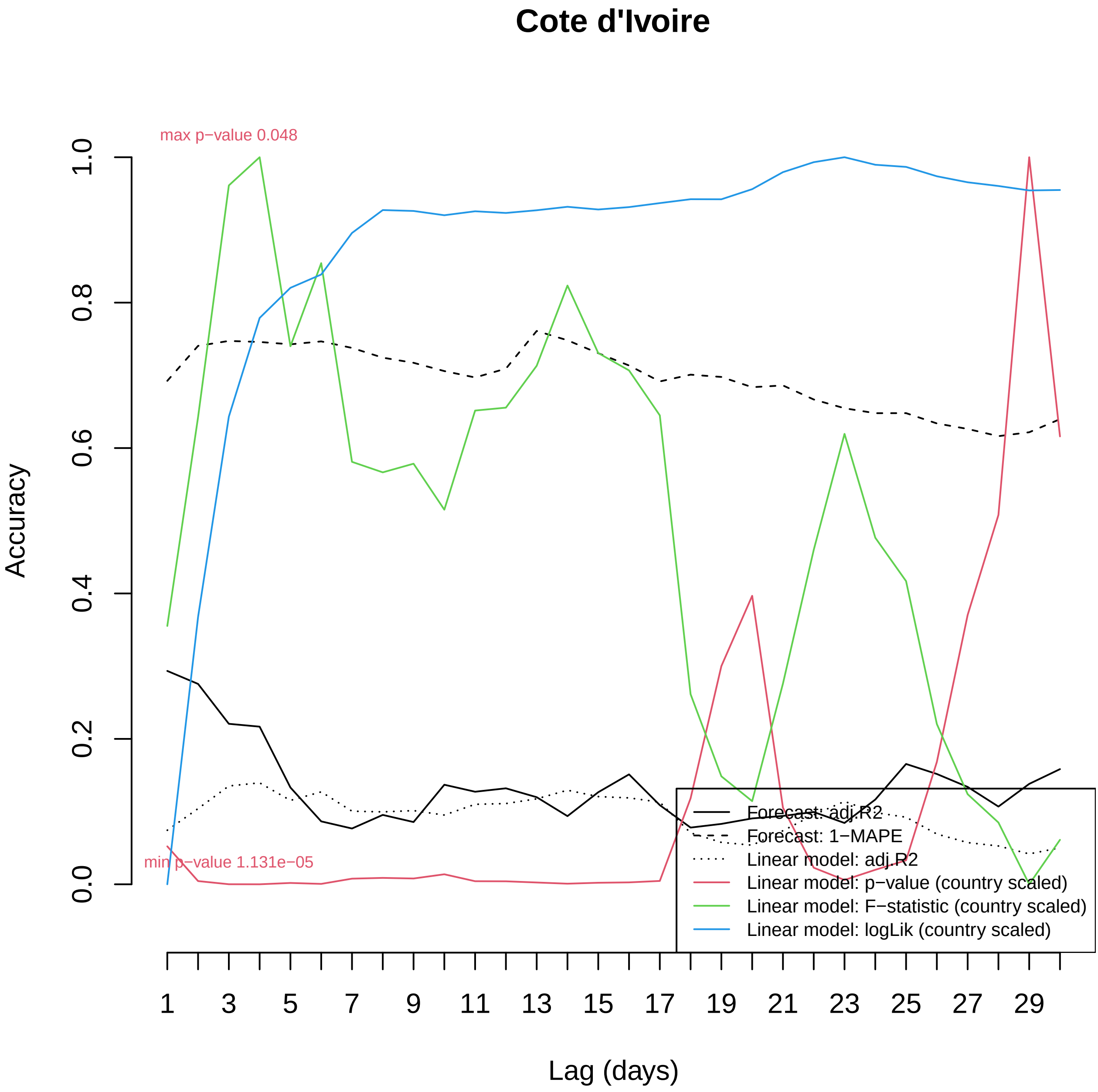

Cote d'Ivoire
max p−value 0.048
min p−value 1.131e−05
Accuracy
0.0
0.2
0.4
0.6
0.8
1.0
1
3
5
7
9
11
13
15
17
19
21
23
25
27
29
Lag (days)
Forecast: adj.R2
Forecast: 1−MAPE
Linear model: adj.R2
Linear model: p−value (country scaled)
Linear model: F−statistic (country scaled)
Linear model: logLik (country scaled)

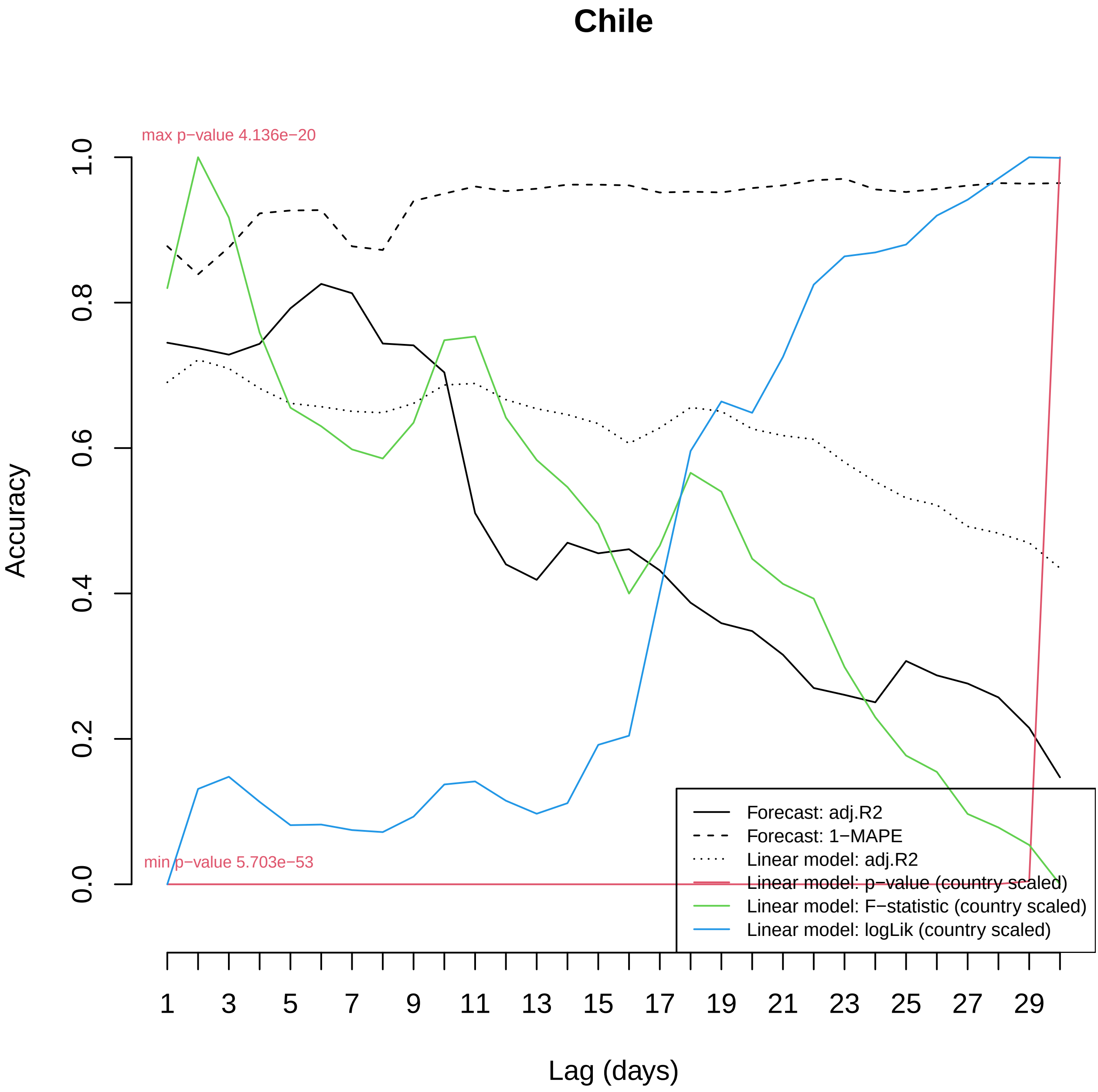

Chile
max p−value 4.136e−20
min p−value 5.703e−53
Accuracy
0.0
0.2
0.4
0.6
0.8
1.0
Lag (days)
1
3
5
7
9
11
13
15
17
19
21
23
25
27
29
Forecast: adj.R2
Forecast: 1−MAPE
Linear model: adj.R2
Linear model: p−value (country scaled)
Linear model: F−statistic (country scaled)
Linear model: logLik (country scaled)

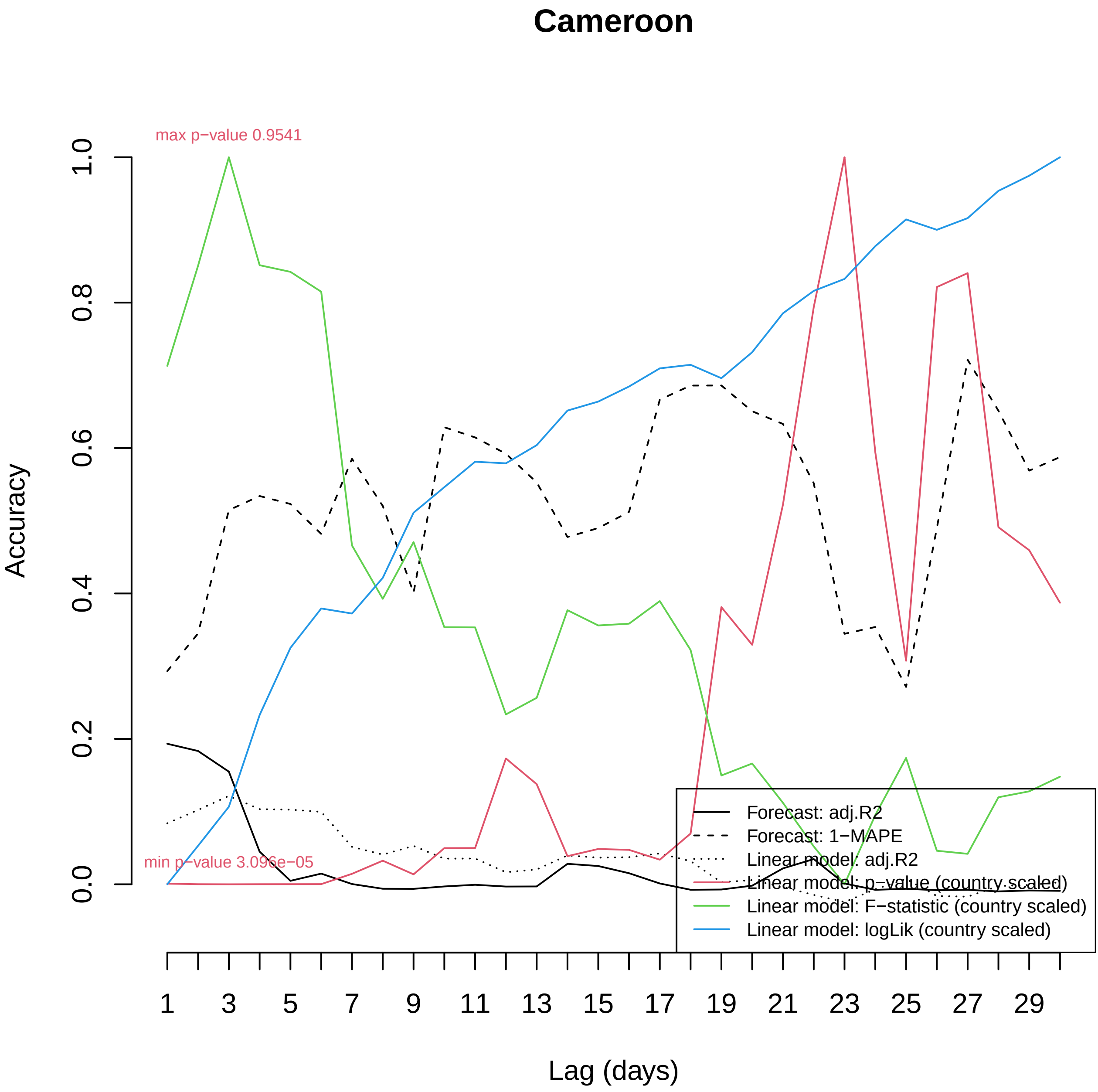
Cameroon
max p−value 0.9541
min p−value 3.096e−05
Accuracy
0.0
0.2
0.4
0.6
0.8
1.0
Lag (days)
1
3
5
7
9
11
13
15
17
19
21
23
25
27
29
Forecast: adj.R2
Forecast: 1−MAPE
Linear model: adj.R2
Linear model: p−value (country scaled)
Linear model: F−statistic (country scaled)
Linear model: logLik (country scaled)

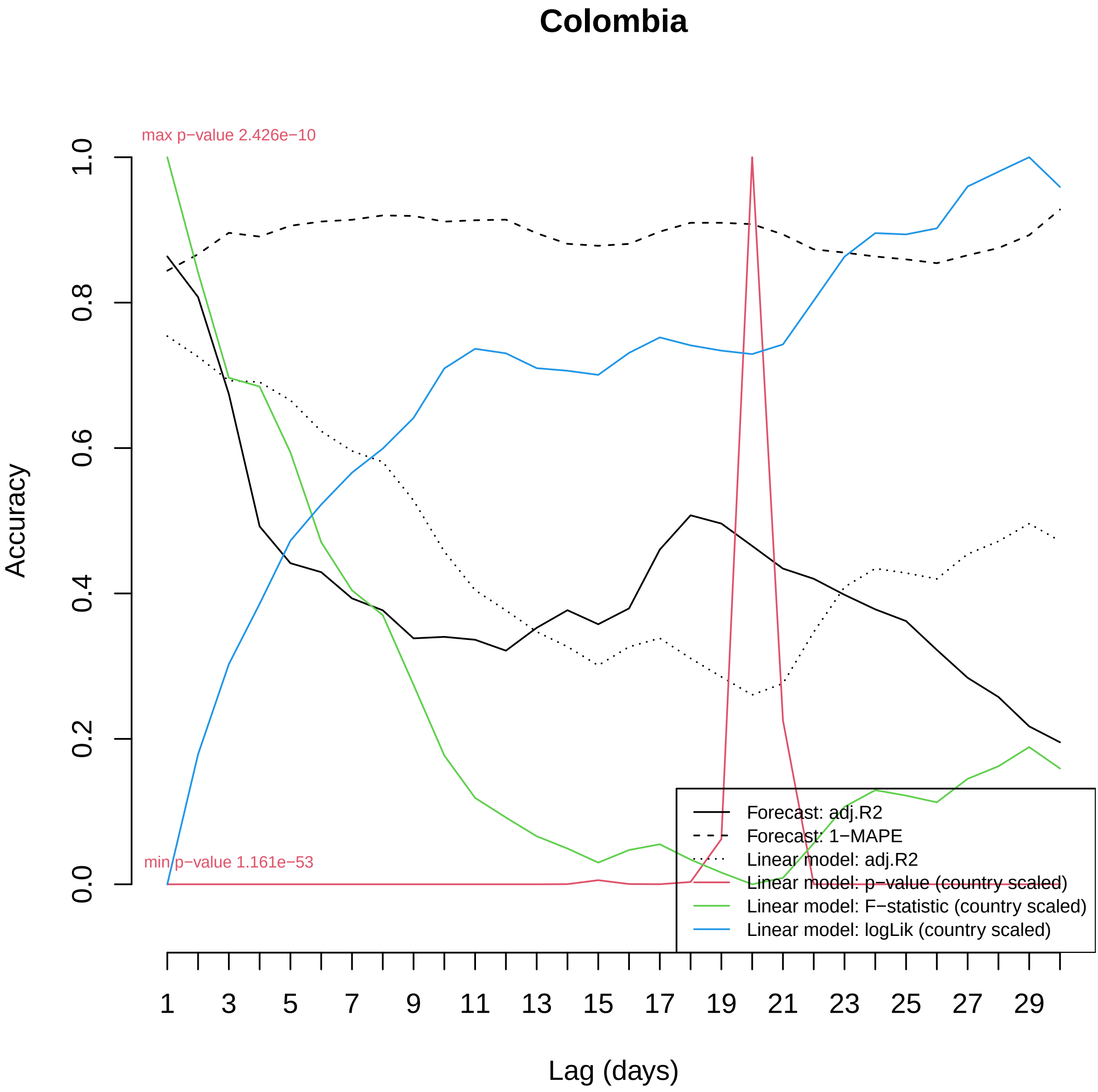

Colombia
max p−value 2.426e−10
min p−value 1.161e−53
Accuracy
0.0
0.2
0.4
0.6
0.8
1.0
Lag (days)
1
3
5
7
9
11
13
15
17
19
21
23
25
27
29
Forecast: adj.R2
Forecast: 1−MAPE
Linear model: adj.R2
Linear model: p−value (country scaled)
Linear model: F−statistic (country scaled)
Linear model: logLik (country scaled)

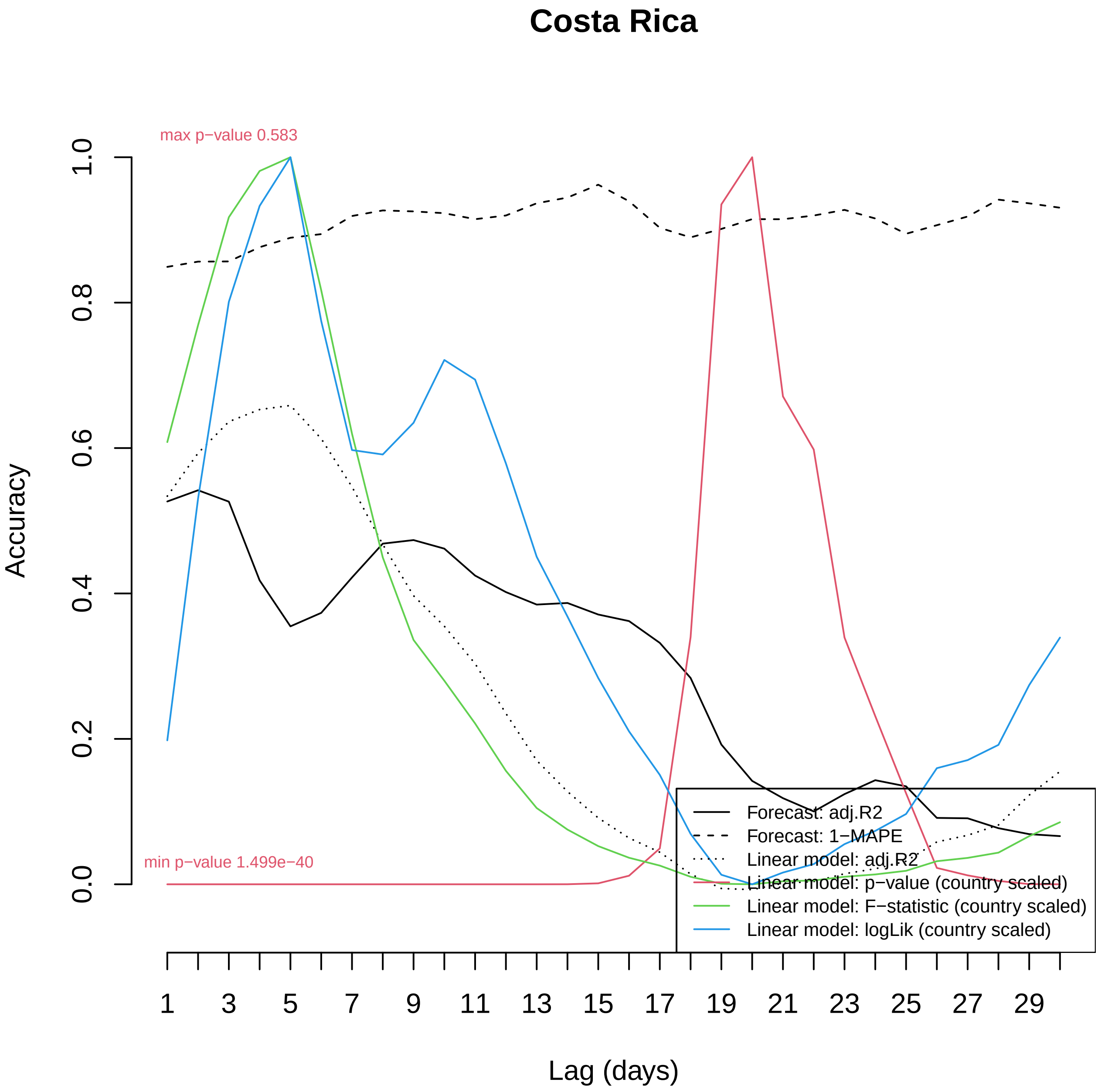

Costa Rica
max p−value 0.583
min p−value 1.499e−40
Accuracy
1.0
0.8
0.6
0.4
0.2
0.0
1
3
5
7
9
11
13
15
17
19
21
23
25
27
29
Lag (days)
Forecast: adj.R2
Forecast: 1−MAPE
Linear model: adj.R2
Linear model: p−value (country scaled)
Linear model: F−statistic (country scaled)
Linear model: logLik (country scaled)

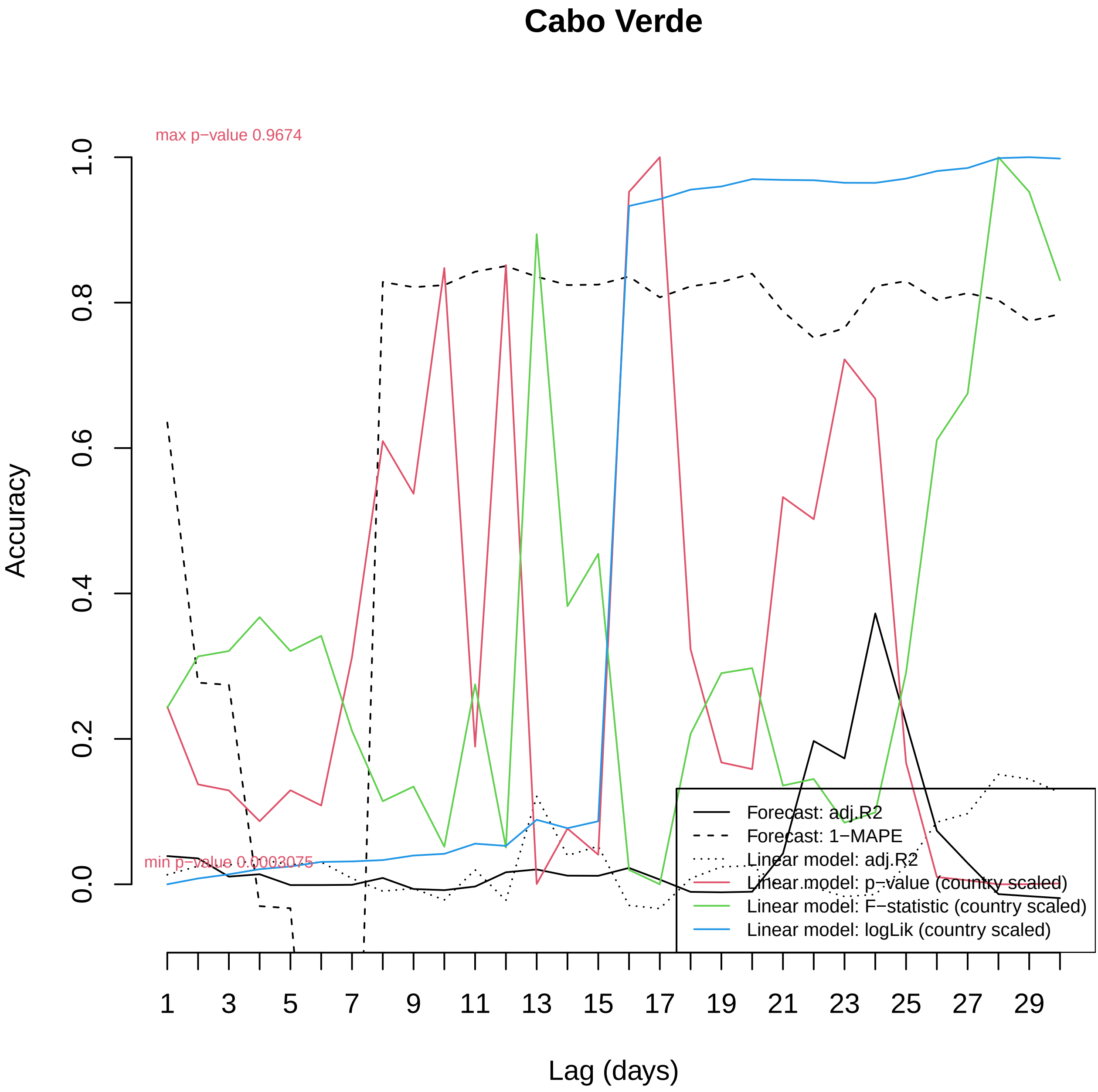

Cabo Verde
max p−value 0.9674
min p−value 0.0003075
Accuracy
1.0
0.8
0.6
0.4
0.2
0.0
1
3
5
7
9
11
13
15
17
19
21
23
25
27
29
Lag (days)
Forecast: adj.R2
Forecast: 1−MAPE
Linear model: adj.R2
Linear model: p−value (country scaled)
Linear model: F−statistic (country scaled)
Linear model: logLik (country scaled)

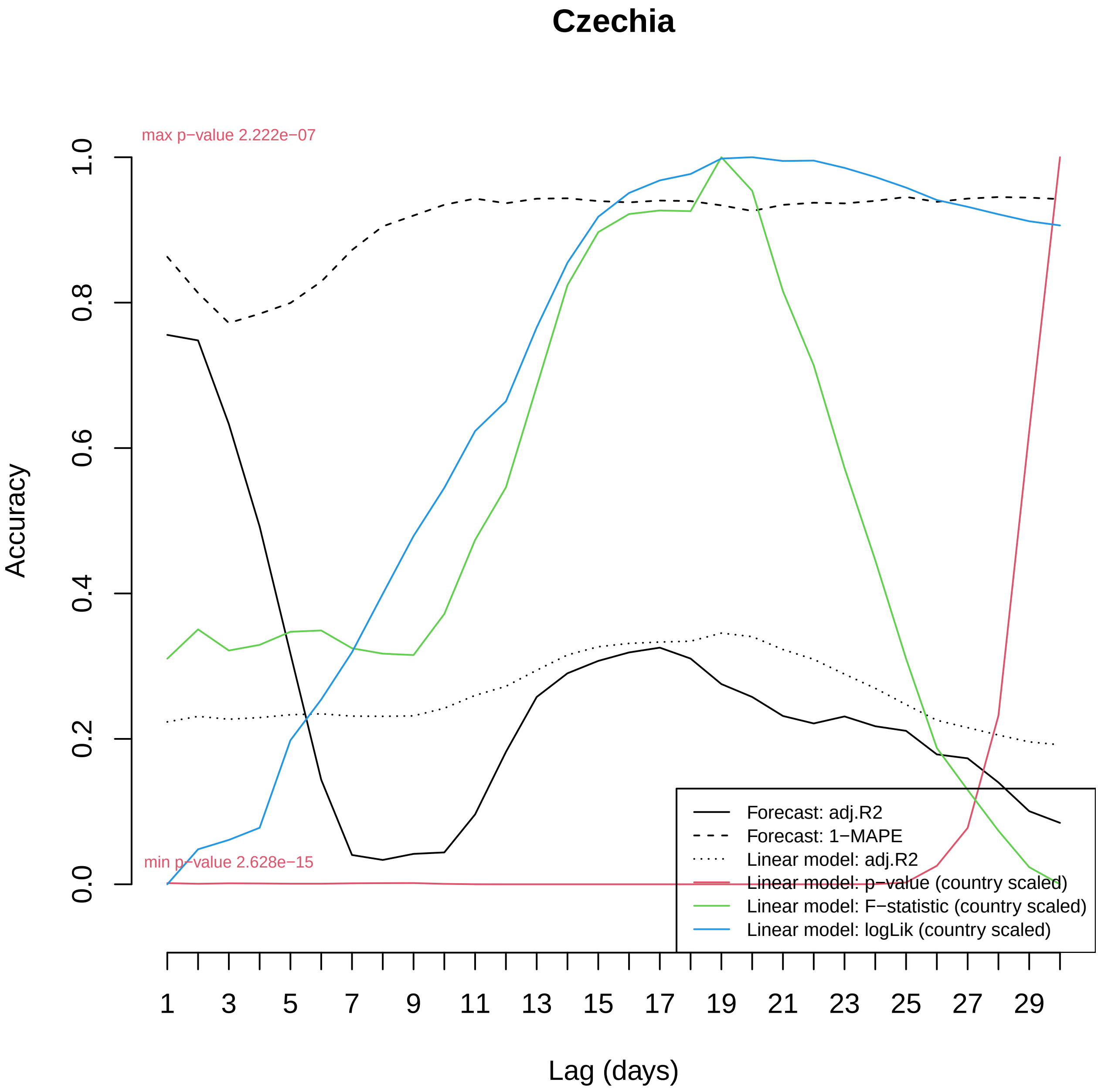

Czechia
max p−value 2.222e−07
min p−value 2.628e−15
Accuracy
0.0
0.2
0.4
0.6
0.8
1.0
1
3
5
7
9
11
13
15
17
19
21
23
25
27
29
Lag (days)
Forecast: adj.R2
Forecast: 1−MAPE
Linear model: adj.R2
Linear model: p−value (country scaled)
Linear model: F−statistic (country scaled)
Linear model: logLik (country scaled)

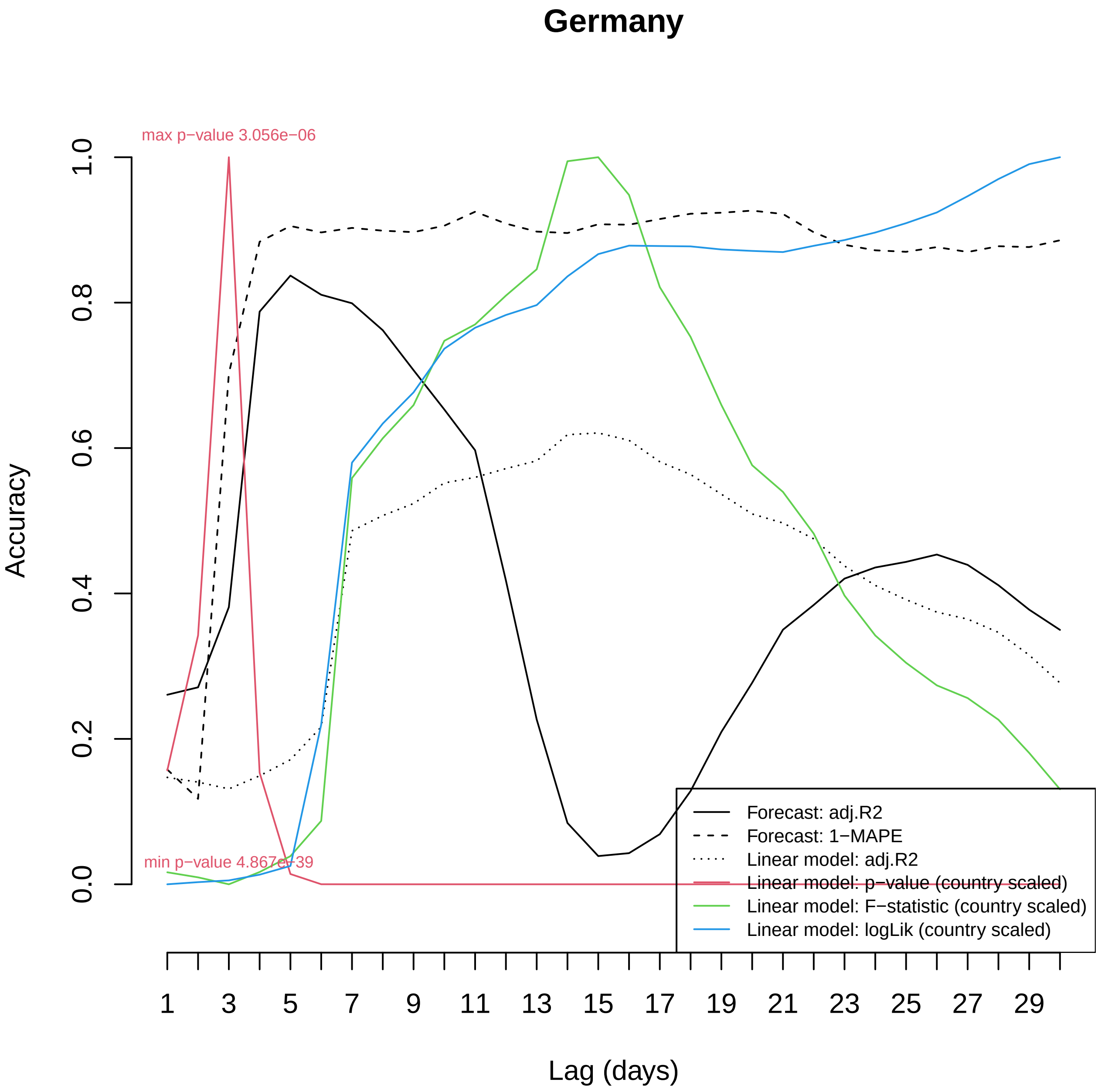

Germany
max p−value 3.056e−06
min p−value 4.867e−39
Accuracy
0.0
0.2
0.4
0.6
0.8
1.0
Lag (days)
1
3
5
7
9
11
13
15
17
19
21
23
25
27
29
Forecast: adj.R2
Forecast: 1−MAPE
Linear model: adj.R2
Linear model: p−value (country scaled)
Linear model: F−statistic (country scaled)
Linear model: logLik (country scaled)

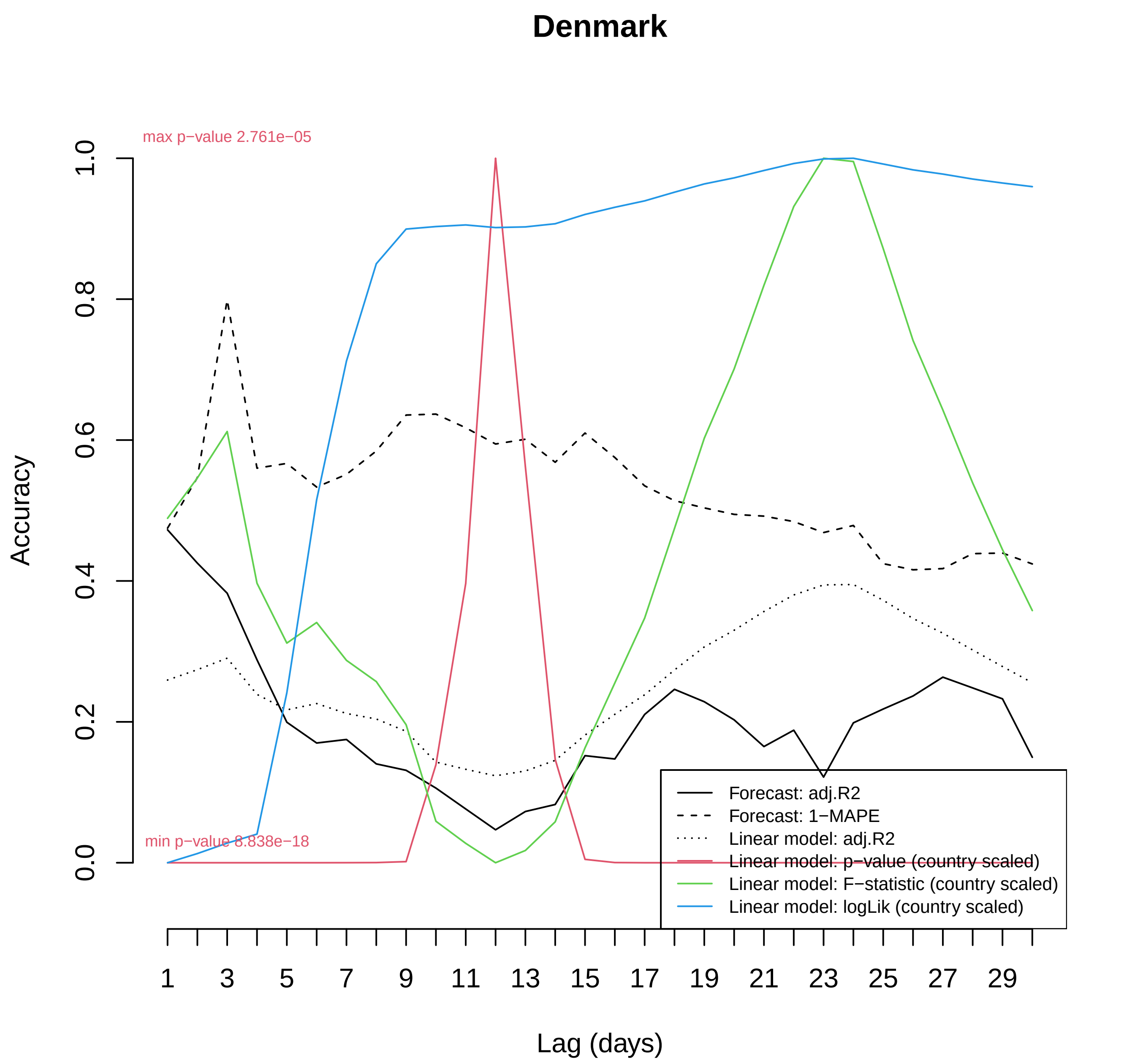

Denmark
max p−value 2.761e−05
min p−value 8.838e−18
Accuracy
0.0
0.2
0.4
0.6
0.8
1.0
1
3
5
7
9
11
13
15
17
19
21
23
25
27
29
Lag (days)
Forecast: adj.R2
Forecast: 1−MAPE
Linear model: adj.R2
Linear model: p−value (country scaled)
Linear model: F−statistic (country scaled)
Linear model: logLik (country scaled)

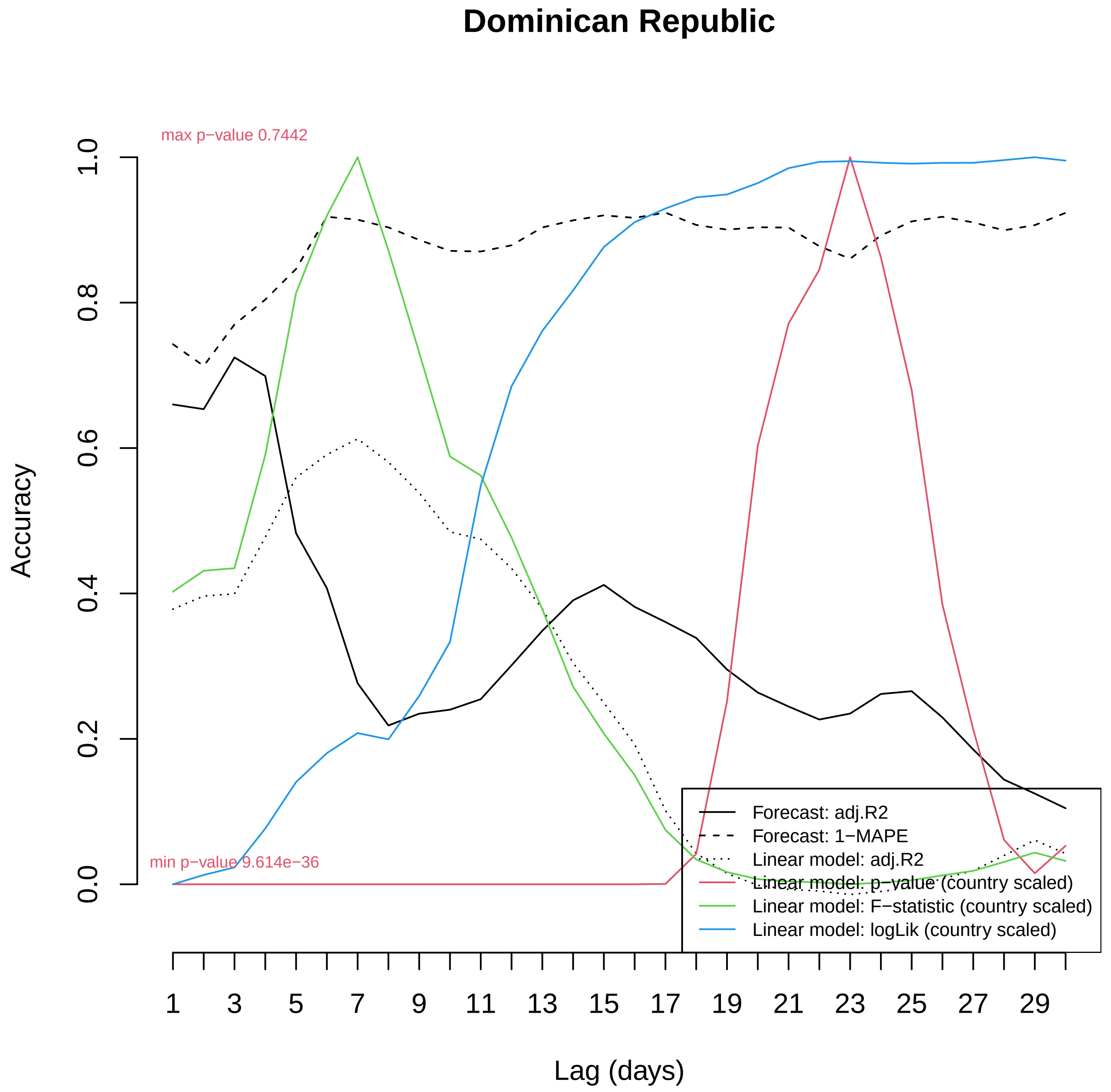
Dominican Republic
max p−value 0.7442
min p−value 9.614e−36
Accuracy
0.0
0.2
0.4
0.6
0.8
1.0
Lag (days)
1
3
5
7
9
11
13
15
17
19
21
23
25
27
29
Forecast: adj.R2
Forecast: 1−MAPE
Linear model: adj.R2
Linear model: p−value (country scaled)
Linear model: F−statistic (country scaled)
Linear model: logLik (country scaled)

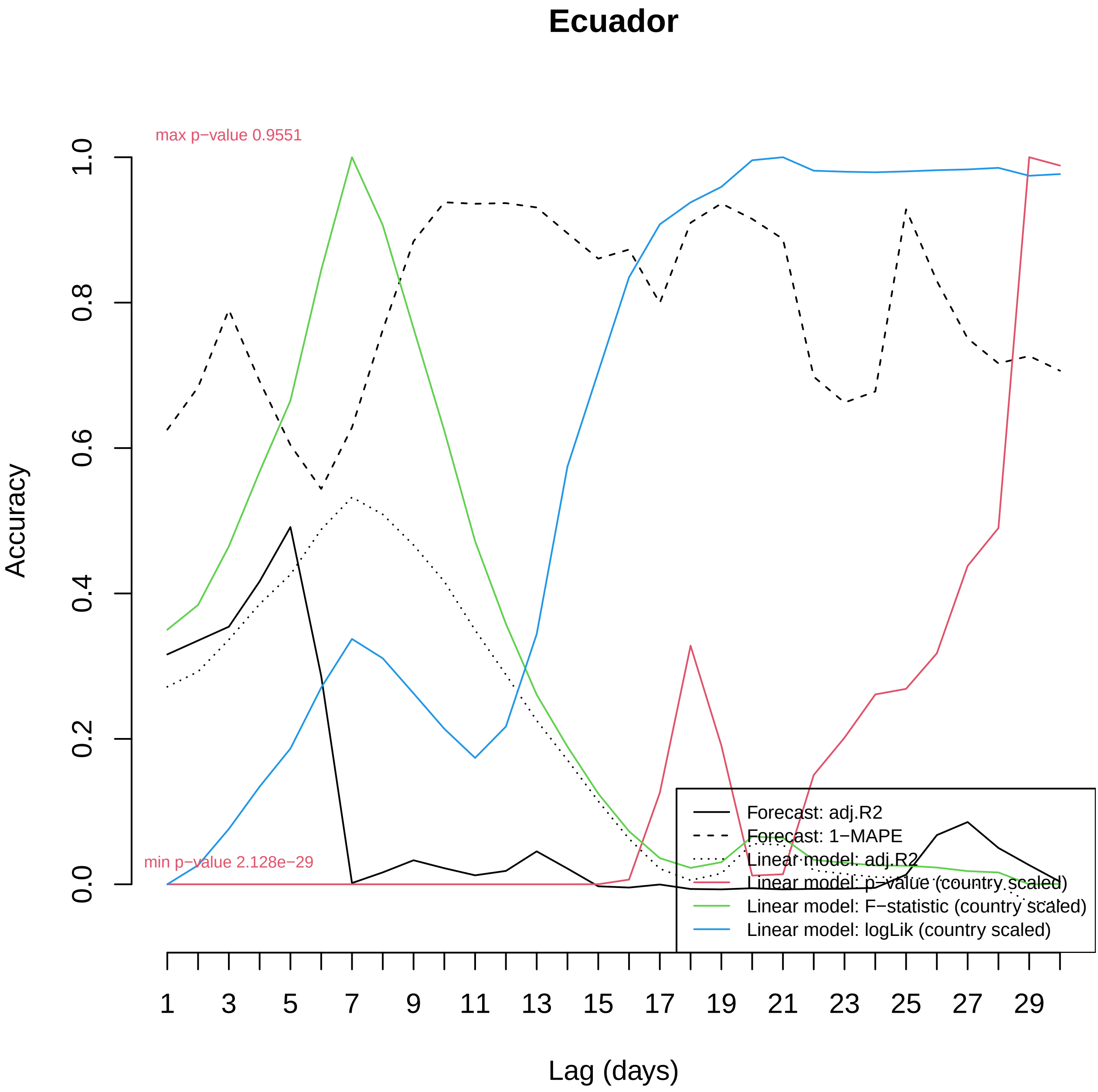

Ecuador
max p−value 0.9551
min p−value 2.128e−29
Accuracy
0.0
0.2
0.4
0.6
0.8
1.0
1
3
5
7
9
11
13
15
17
19
21
23
25
27
29
Lag (days)
Forecast: adj.R2
Forecast: 1−MAPE
Linear model: adj.R2
Linear model: p−value (country scaled)
Linear model: F−statistic (country scaled)
Linear model: logLik (country scaled)

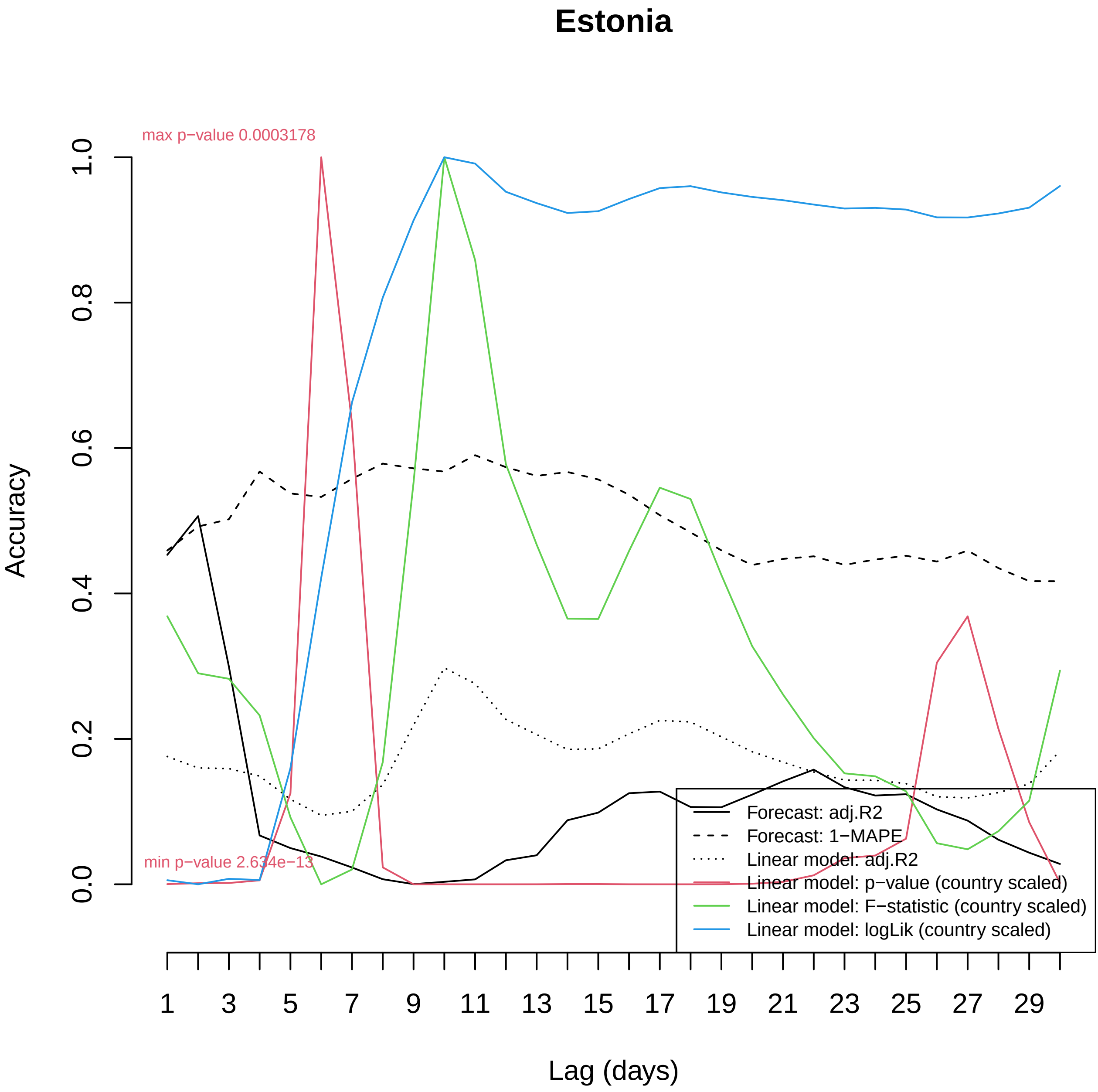
Estonia
max p−value 0.0003178
min p−value 2.634e−13
Accuracy
0.0
0.2
0.4
0.6
0.8
1.0
Lag (days)
1
3
5
7
9
11
13
15
17
19
21
23
25
27
29
Forecast: adj.R2
Forecast: 1−MAPE
Linear model: adj.R2
Linear model: p−value (country scaled)
Linear model: F−statistic (country scaled)
Linear model: logLik (country scaled)

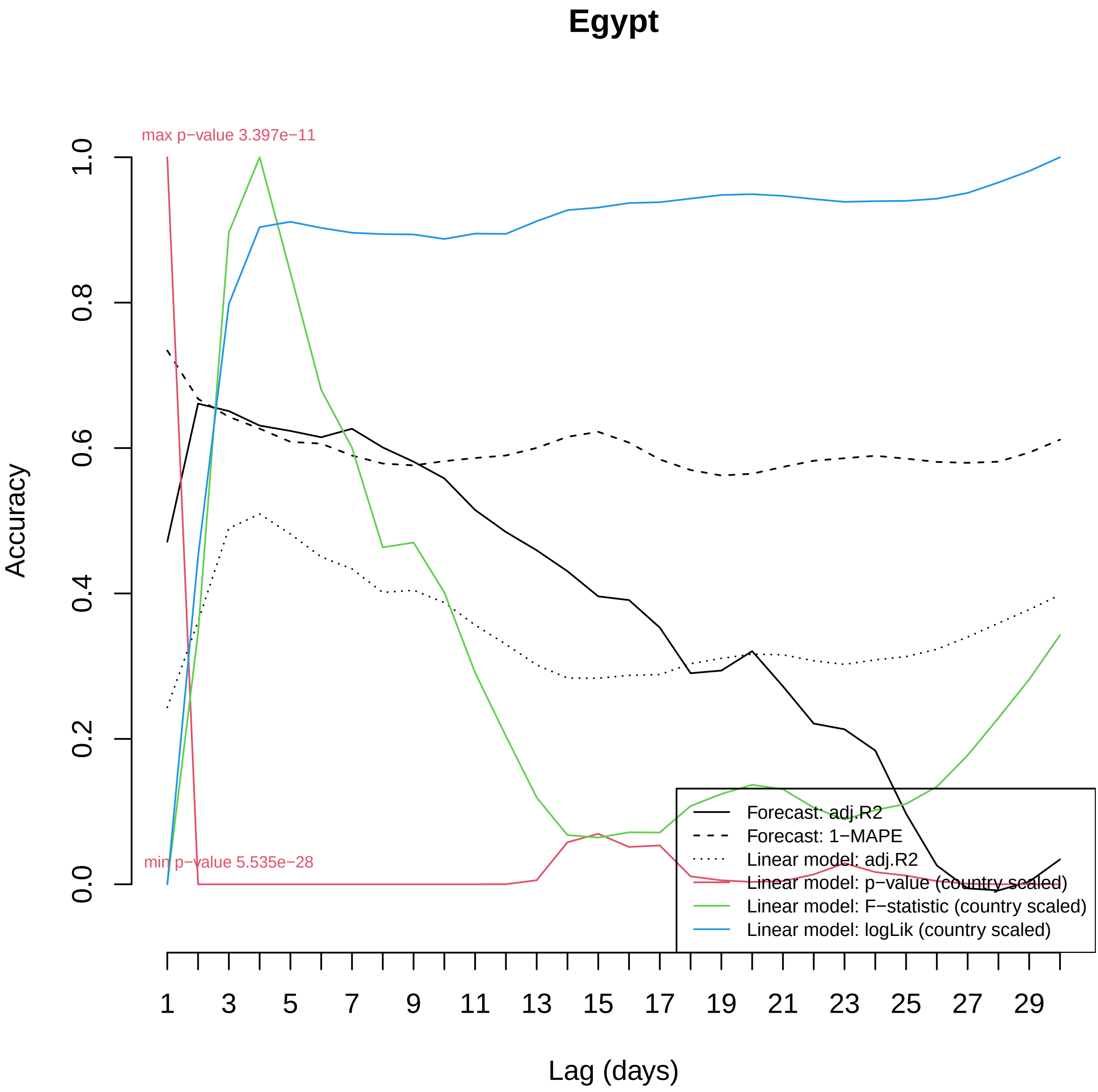
Egypt
max p−value 3.397e−11
min p−value 5.535e−28
Accuracy
0.0
0.2
0.4
0.6
0.8
1.0
Lag (days)
1
3
5
7
9
11
13
15
17
19
21
23
25
27
29
Forecast: adj.R2
Forecast: 1−MAPE
Linear model: adj.R2
Linear model: p−value (country scaled)
Linear model: F−statistic (country scaled)
Linear model: logLik (country scaled)

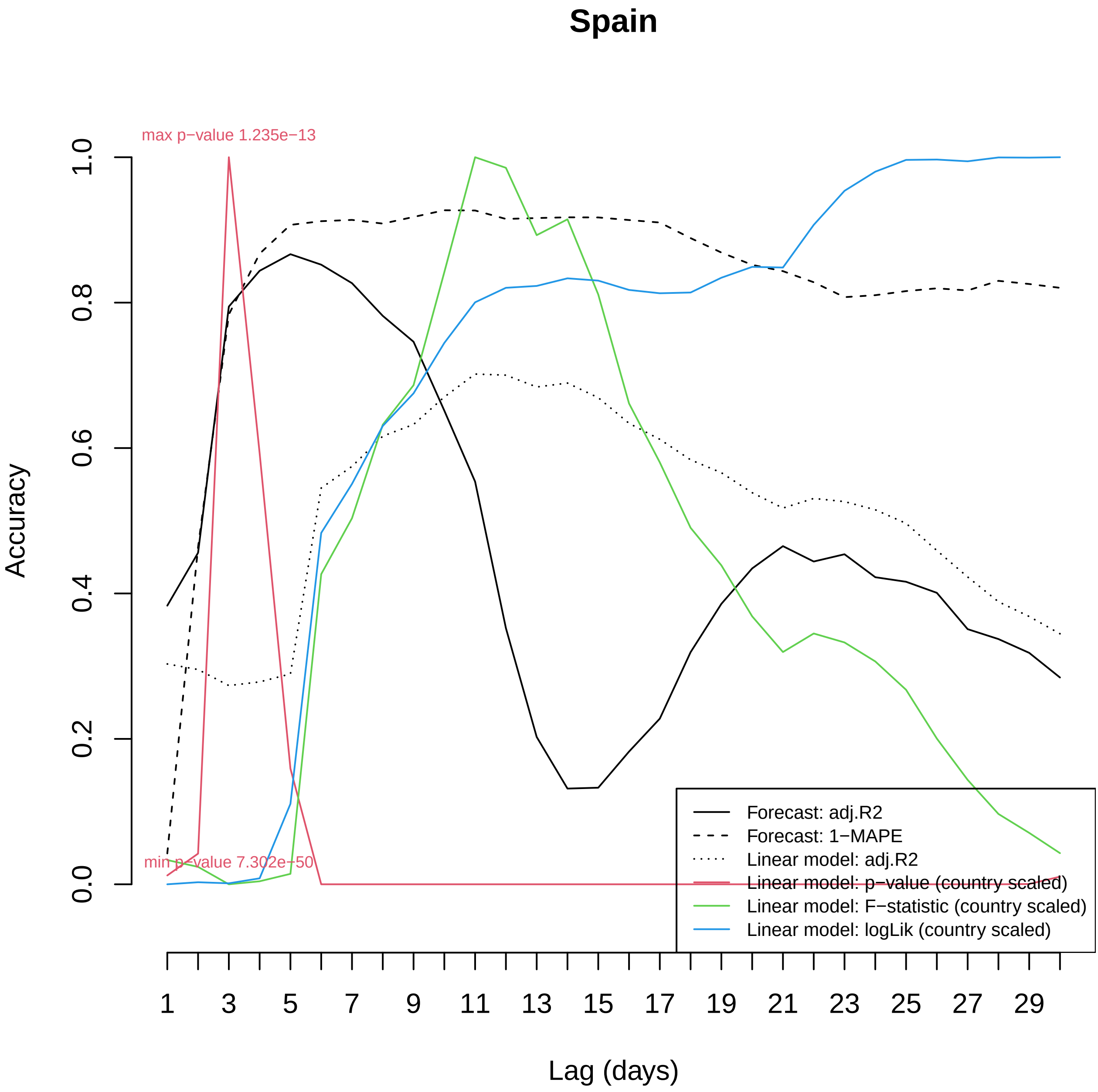
Spain
max p−value 1.235e−13
min p−value 7.302e−50
Accuracy
1.0
0.8
0.6
0.4
0.2
0.0
1
3
5
7
9
11
13
15
17
19
21
23
25
27
29
Lag (days)
Forecast: adj.R2
Forecast: 1−MAPE
Linear model: adj.R2
Linear model: p−value (country scaled)
Linear model: F−statistic (country scaled)
Linear model: logLik (country scaled)

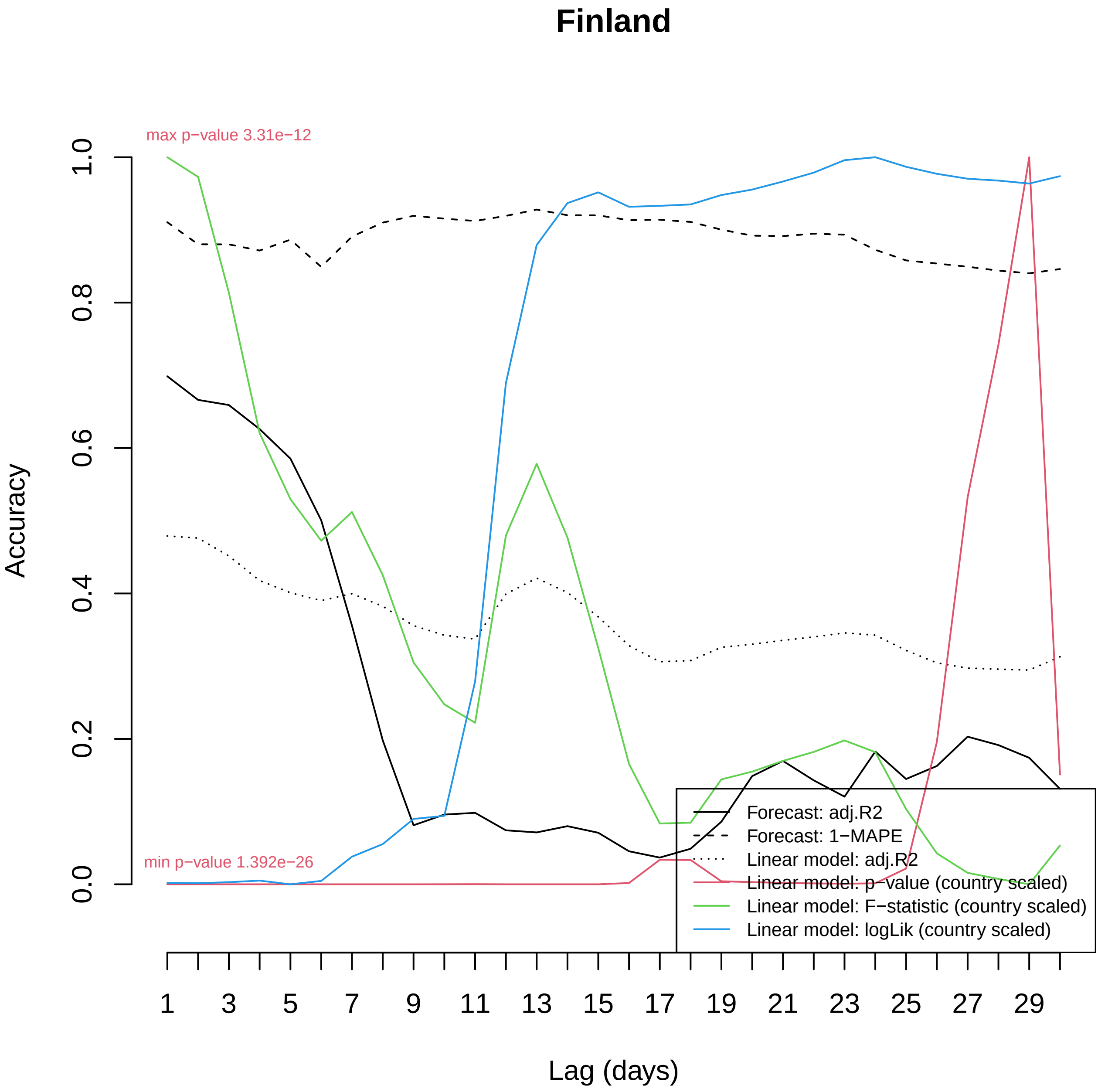
Finland
max p−value 3.31e−12
min p−value 1.392e−26
Accuracy
0.0
0.2
0.4
0.6
0.8
1.0
1
3
5
7
9
11
13
15
17
19
21
23
25
27
29
Lag (days)
Forecast: adj.R2
Forecast: 1−MAPE
Linear model: adj.R2
Linear model: p−value (country scaled)
Linear model: F−statistic (country scaled)
Linear model: logLik (country scaled)

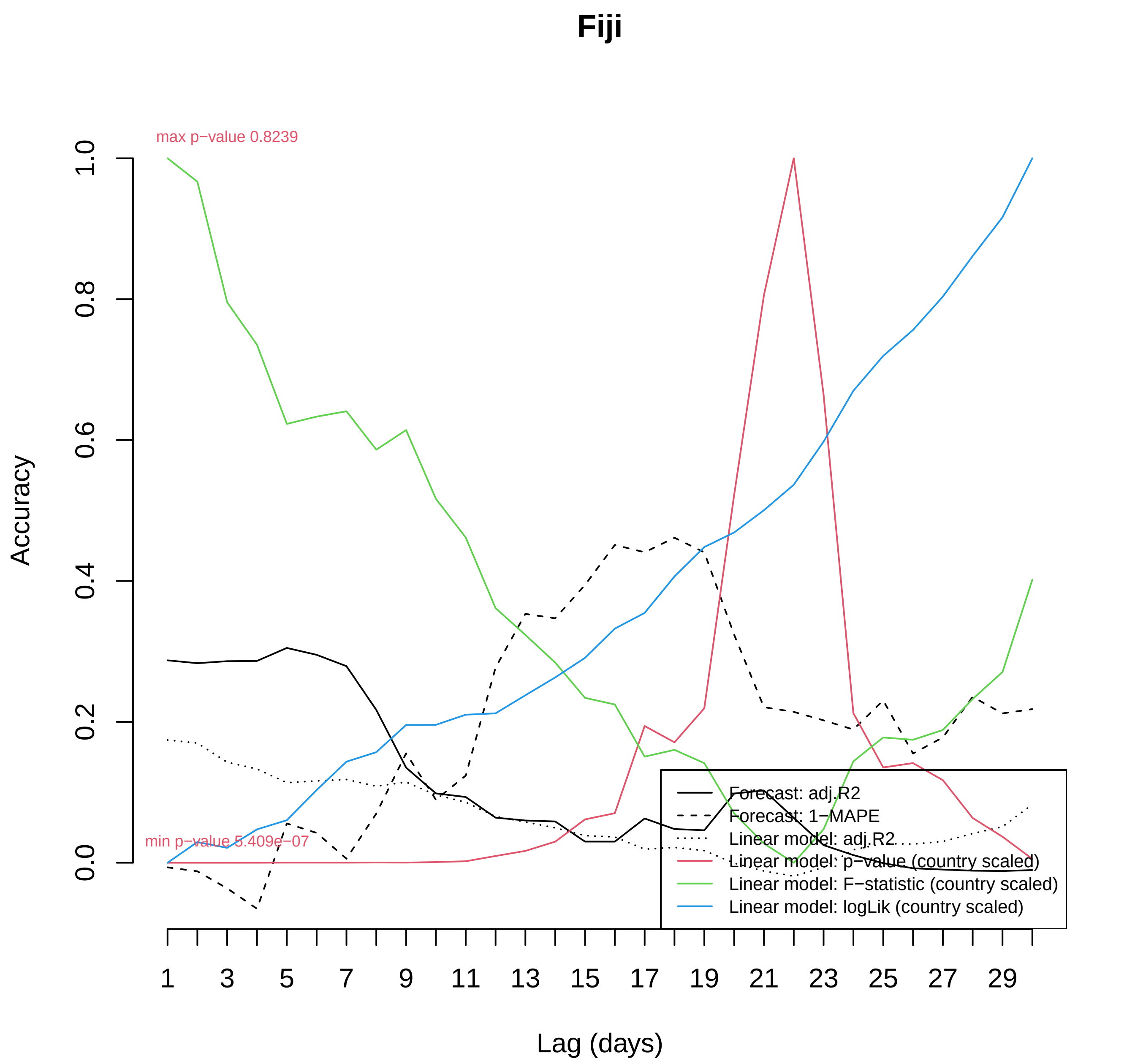

Fiji
max p−value 0.8239
min p−value 5.409e−07
Accuracy
0.0
0.2
0.4
0.6
0.8
1.0
Lag (days)
1
3
5
7
9
11
13
15
17
19
21
23
25
27
29
Forecast: adj.R2
Forecast: 1−MAPE
Linear model: adj.R2
Linear model: p−value (country scaled)
Linear model: F−statistic (country scaled)
Linear model: logLik (country scaled)

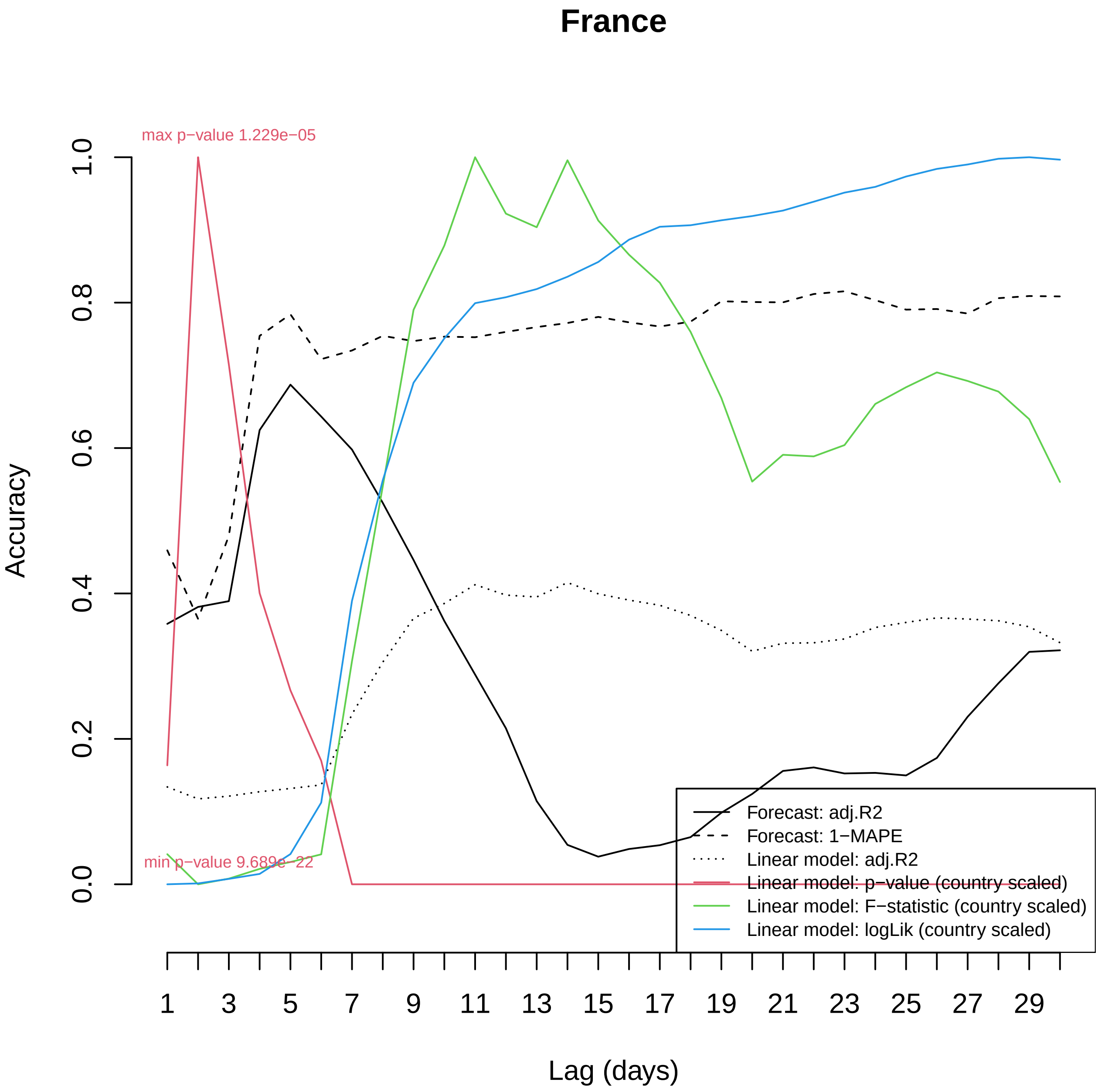
France
max p−value 1.229e−05
min p−value 9.689e−22
Accuracy
0.0
0.2
0.4
0.6
0.8
1.0
Lag (days)
1
3
5
7
9
11
13
15
17
19
21
23
25
27
29
Forecast: adj.R2
Forecast: 1−MAPE
Linear model: adj.R2
Linear model: p−value (country scaled)
Linear model: F−statistic (country scaled)
Linear model: logLik (country scaled)

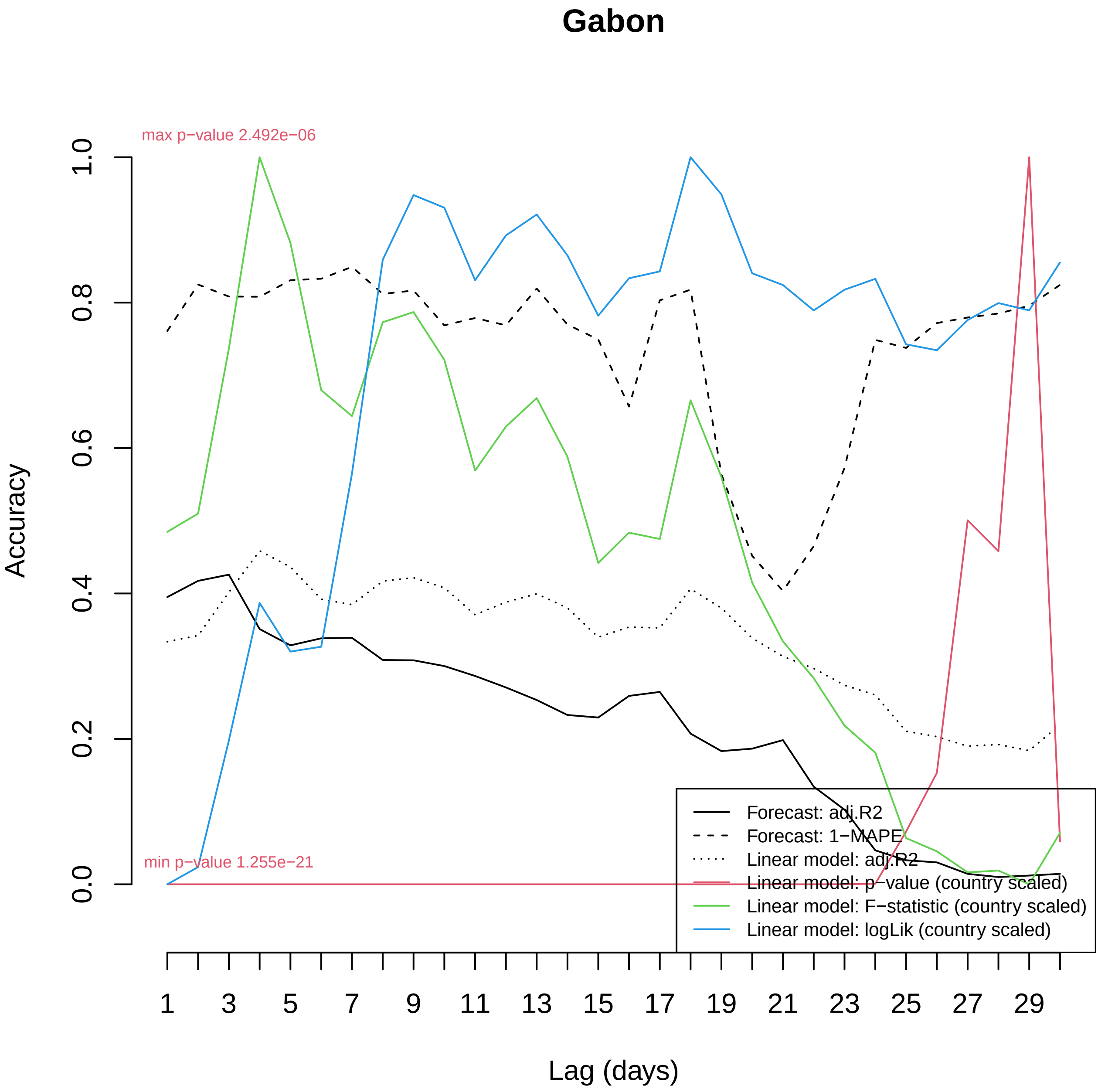

Gabon
max p−value 2.492e−06
min p−value 1.255e−21
Accuracy
0.0
0.2
0.4
0.6
0.8
1.0
Lag (days)
1
3
5
7
9
11
13
15
17
19
21
23
25
27
29
Forecast: adj.R2
Forecast: 1−MAPE
Linear model: adj.R2
Linear model: p−value (country scaled)
Linear model: F−statistic (country scaled)
Linear model: logLik (country scaled)

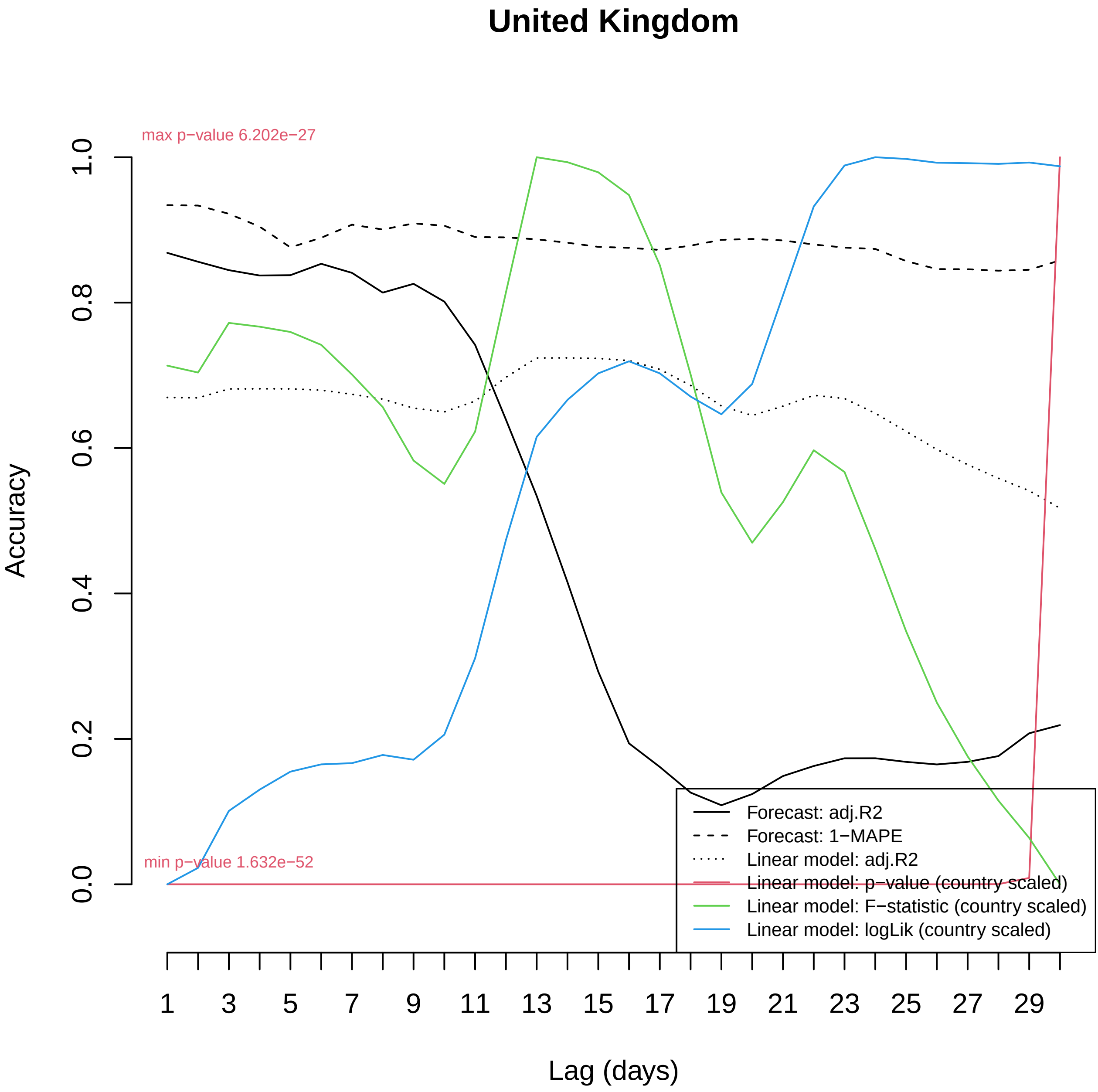

United Kingdom
max p−value 6.202e−27
min p−value 1.632e−52
Accuracy
0.0
0.2
0.4
0.6
0.8
1.0
1
3
5
7
9
11
13
15
17
19
21
23
25
27
29
Lag (days)
Forecast: adj.R2
Forecast: 1−MAPE
Linear model: adj.R2
Linear model: p−value (country scaled)
Linear model: F−statistic (country scaled)
Linear model: logLik (country scaled)

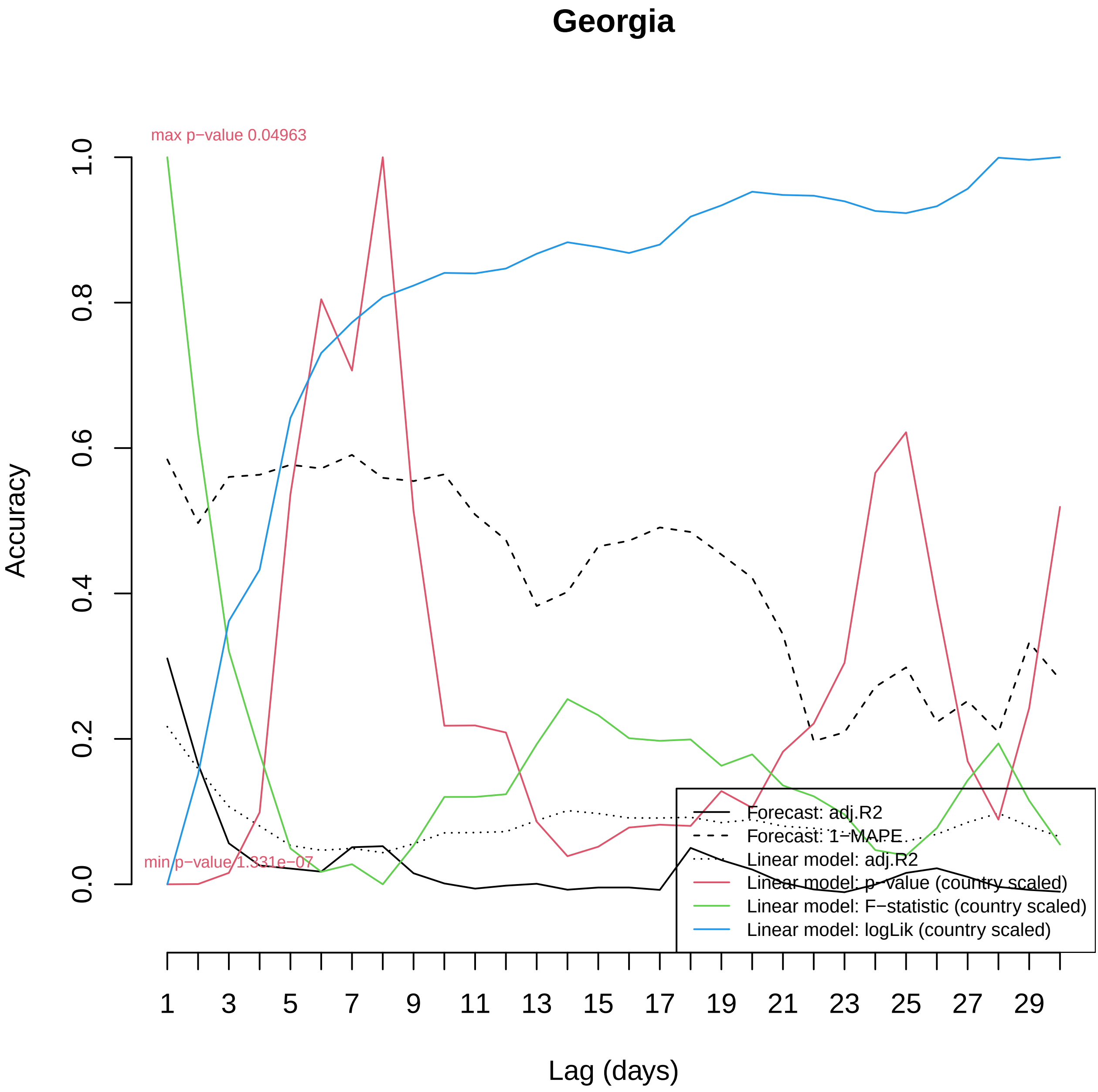

Georgia
max p−value 0.04963
min p−value 1.331e−07
Accuracy
0.0
0.2
0.4
0.6
0.8
1.0
Lag (days)
1
3
5
7
9
11
13
15
17
19
21
23
25
27
29
Forecast: adj.R2
Forecast: 1−MAPE
Linear model: adj.R2
Linear model: p−value (country scaled)
Linear model: F−statistic (country scaled)
Linear model: logLik (country scaled)

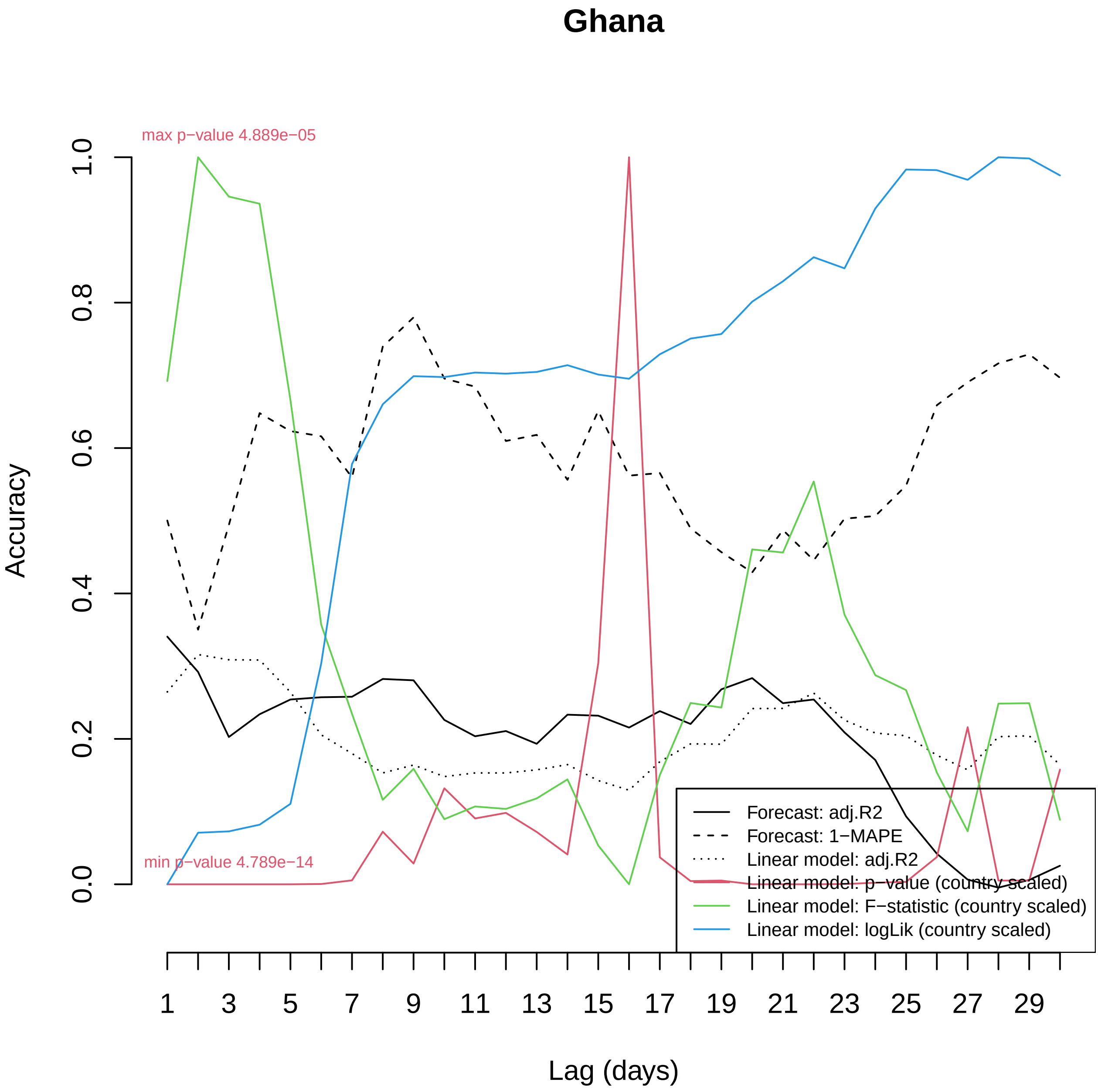

Ghana
max p−value 4.889e−05
min p−value 4.789e−14
Accuracy
0.0
0.2
0.4
0.6
0.8
1.0
Lag (days)
1
3
5
7
9
11
13
15
17
19
21
23
25
27
29
Forecast: adj.R2
Forecast: 1−MAPE
Linear model: adj.R2
Linear model: p−value (country scaled)
Linear model: F−statistic (country scaled)
Linear model: logLik (country scaled)

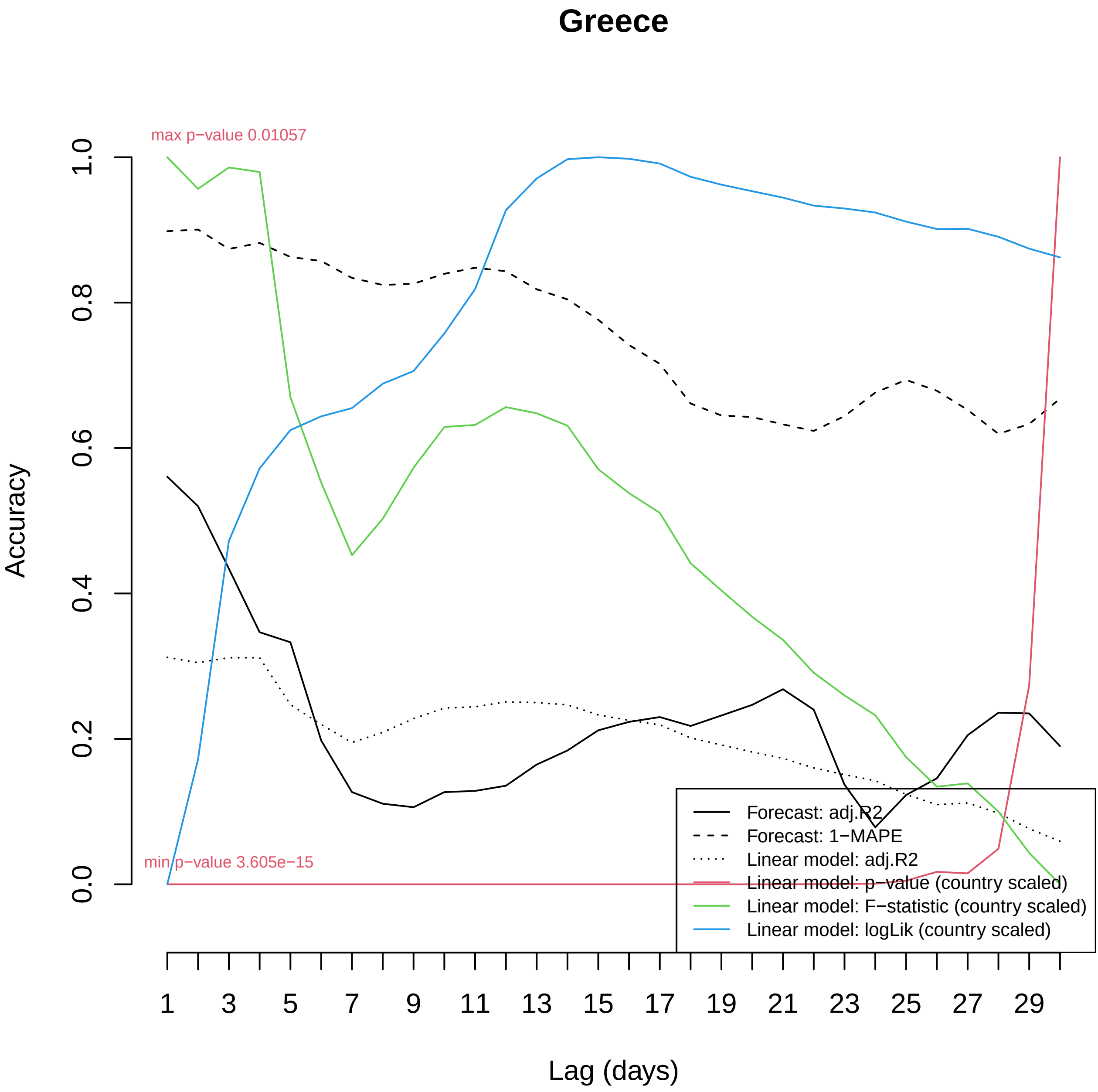

Greece
max p−value 0.01057
min p−value 3.605e−15
Accuracy
0.0
0.2
0.4
0.6
0.8
1.0
Lag (days)
1
3
5
7
9
11
13
15
17
19
21
23
25
27
29
Forecast: adj.R2
Forecast: 1−MAPE
Linear model: adj.R2
Linear model: p−value (country scaled)
Linear model: F−statistic (country scaled)
Linear model: logLik (country scaled)

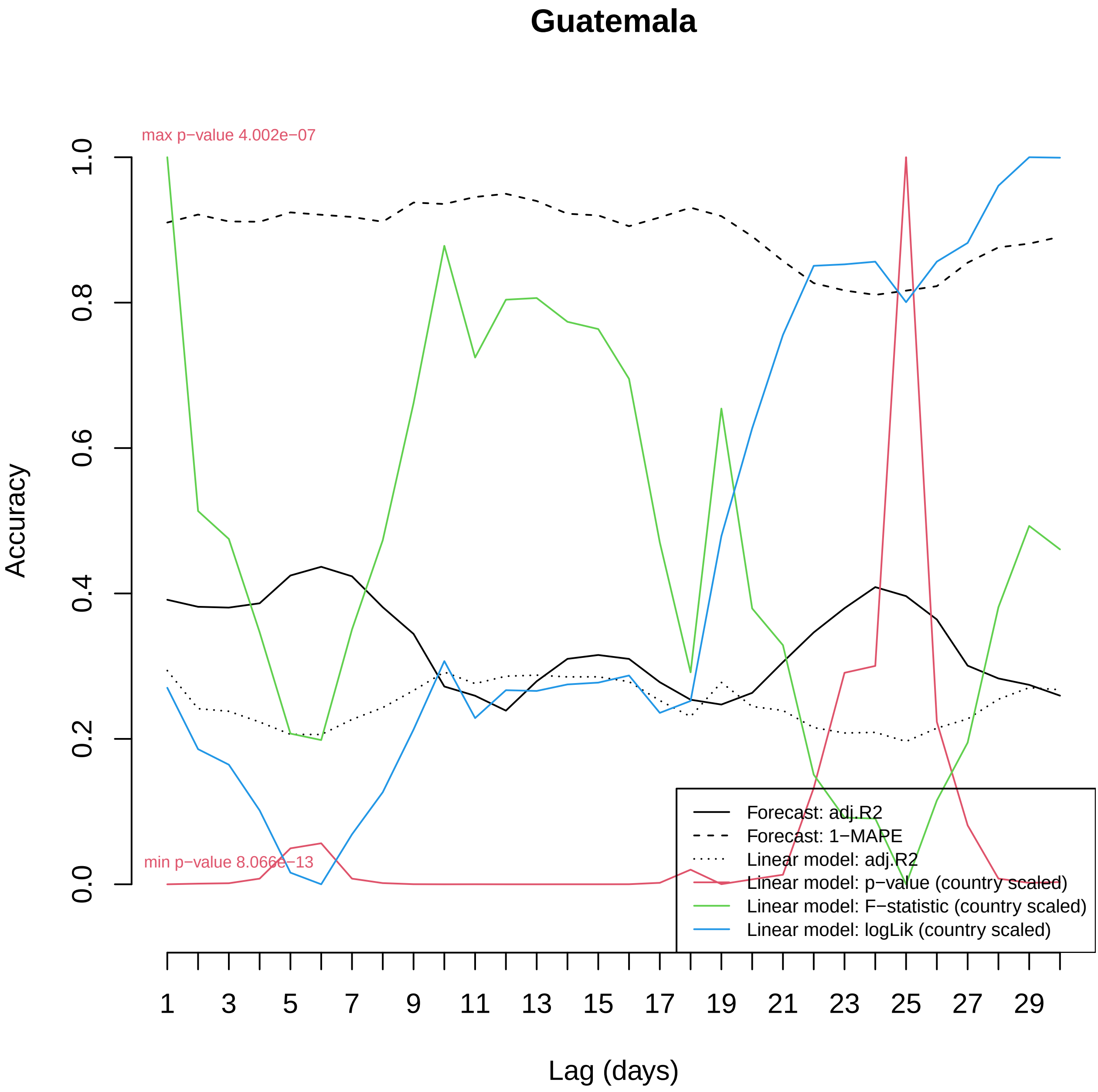
Guatemala
max p−value 4.002e−07
min p−value 8.066e−13
Accuracy
0.0
0.2
0.4
0.6
0.8
1.0
Lag (days)
1
3
5
7
9
11
13
15
17
19
21
23
25
27
29
Forecast: adj.R2
Forecast: 1−MAPE
Linear model: adj.R2
Linear model: p−value (country scaled)
Linear model: F−statistic (country scaled)
Linear model: logLik (country scaled)

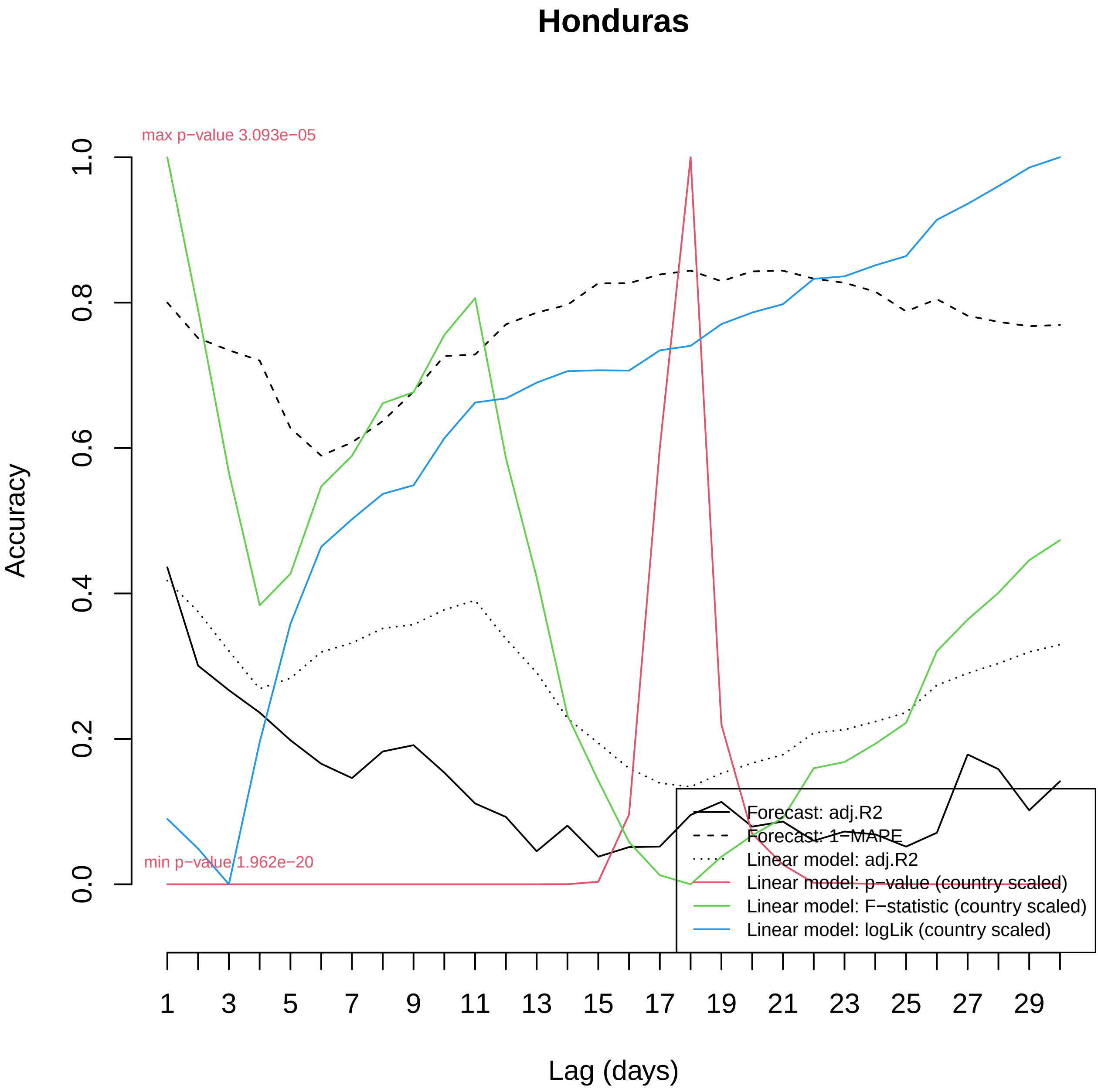

Honduras
max p−value 3.093e−05
min p−value 1.962e−20
Accuracy
0.0
0.2
0.4
0.6
0.8
1.0
1
3
5
7
9
11
13
15
17
19
21
23
25
27
29
Lag (days)
Forecast: adj.R2
Forecast: 1−MAPE
Linear model: adj.R2
Linear model: p−value (country scaled)
Linear model: F−statistic (country scaled)
Linear model: logLik (country scaled)

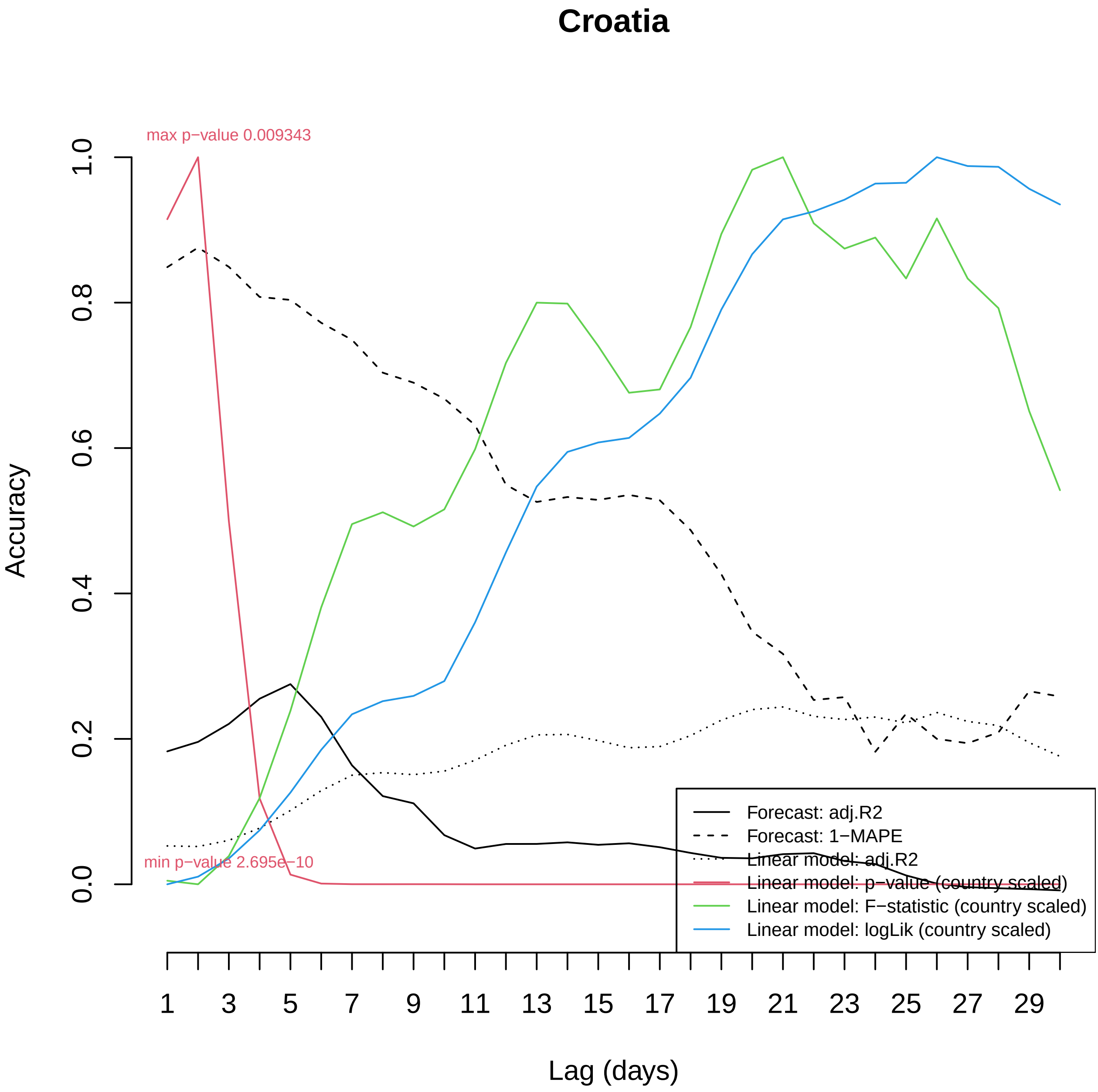
Croatia
max p−value 0.009343
min p−value 2.695e−10
Accuracy
0.0
0.2
0.4
0.6
0.8
1.0
Lag (days)
1
3
5
7
9
11
13
15
17
19
21
23
25
27
29
Forecast: adj.R2
Forecast: 1−MAPE
Linear model: adj.R2
Linear model: p−value (country scaled)
Linear model: F−statistic (country scaled)
Linear model: logLik (country scaled)

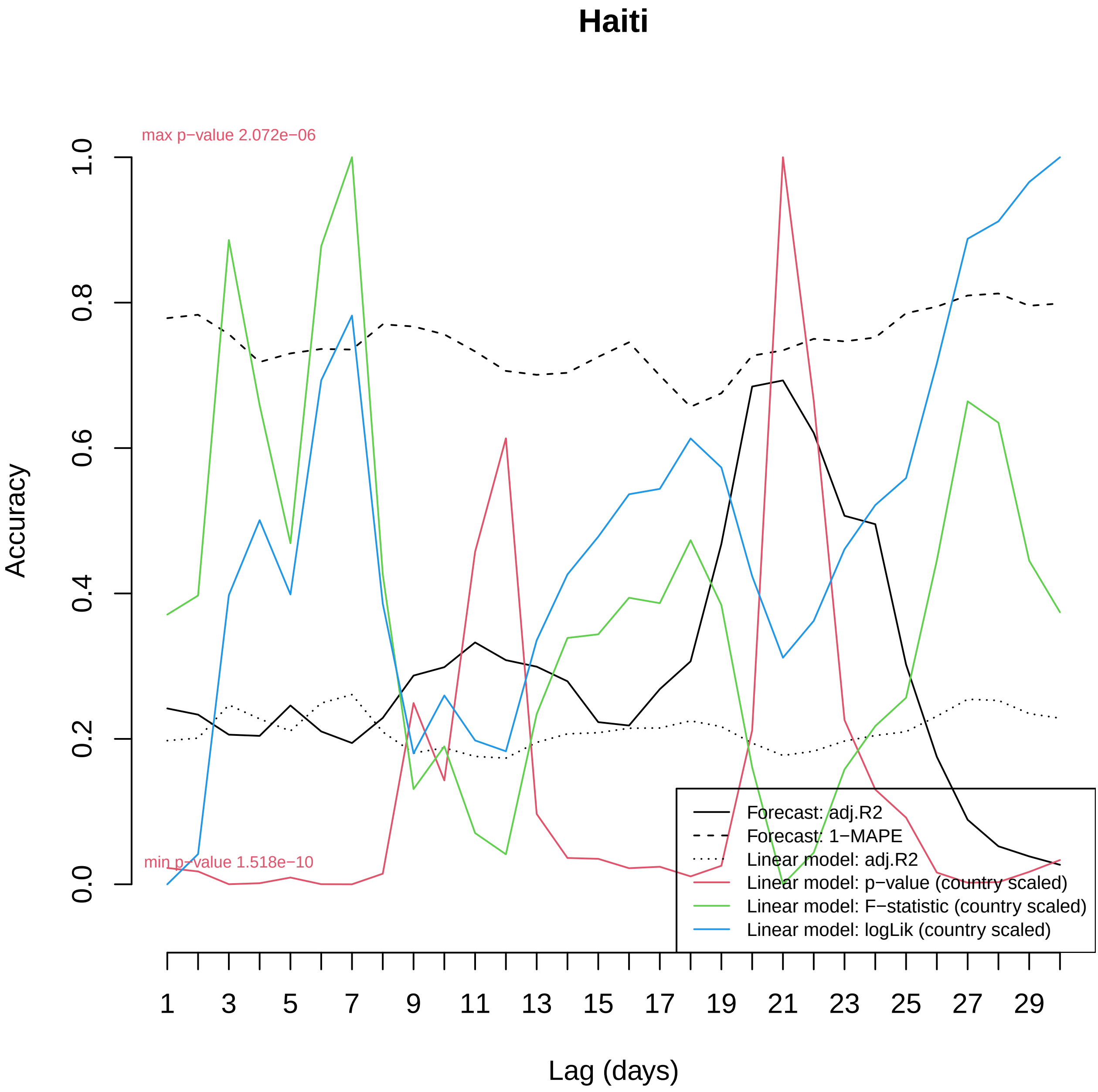

Haiti
max p−value 2.072e−06
min p−value 1.518e−10
Accuracy
0.0
0.2
0.4
0.6
0.8
1.0
Lag (days)
1
3
5
7
9
11
13
15
17
19
21
23
25
27
29
Forecast: adj.R2
Forecast: 1−MAPE
Linear model: adj.R2
Linear model: p−value (country scaled)
Linear model: F−statistic (country scaled)
Linear model: logLik (country scaled)

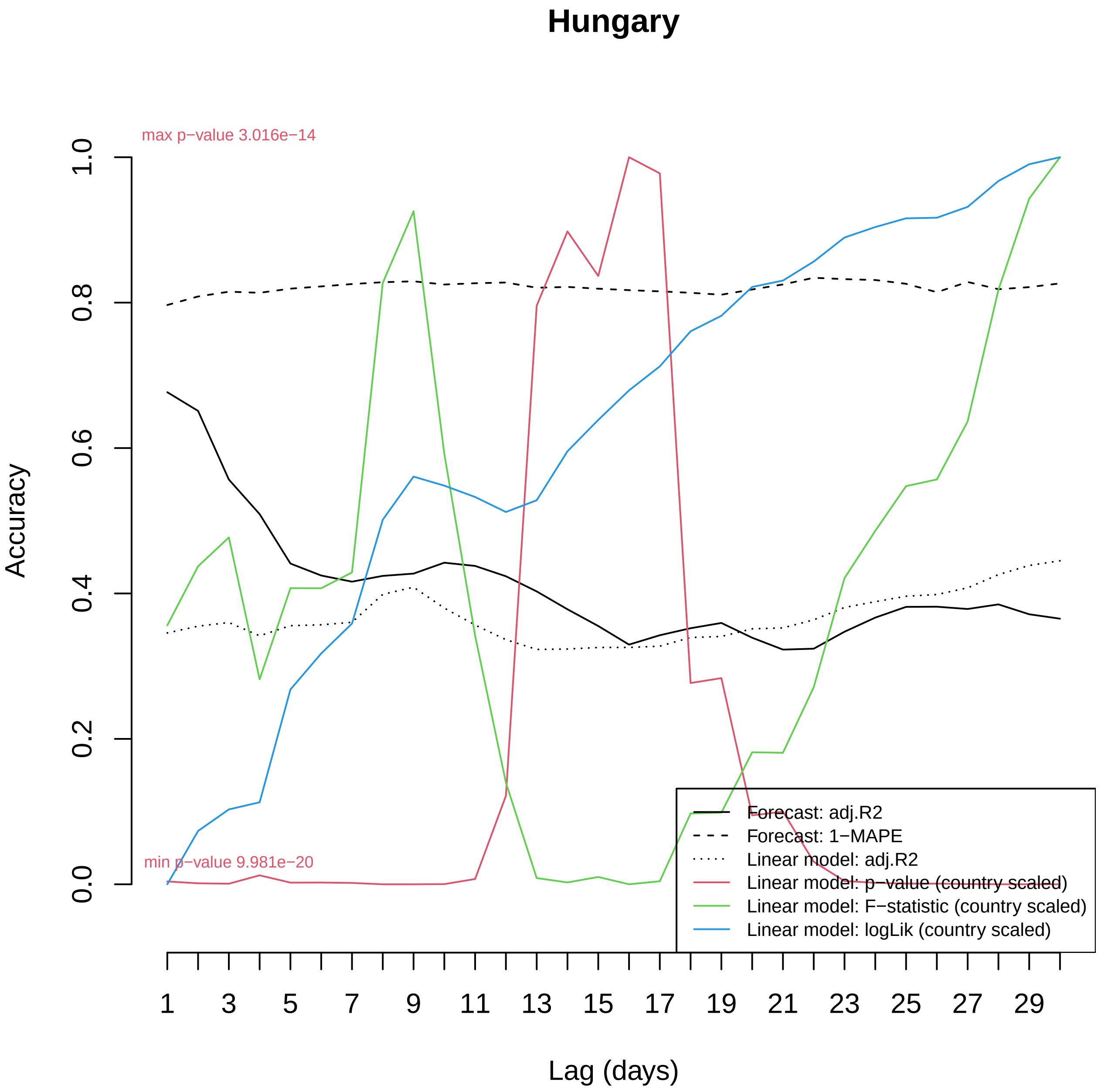
Hungary
max p−value 3.016e−14
min p−value 9.981e−20
Accuracy
0.0
0.2
0.4
0.6
0.8
1.0
1
3
5
7
9
11
13
15
17
19
21
23
25
27
29
Lag (days)
Forecast: adj.R2
Forecast: 1−MAPE
Linear model: adj.R2
Linear model: p−value (country scaled)
Linear model: F−statistic (country scaled)
Linear model: logLik (country scaled)

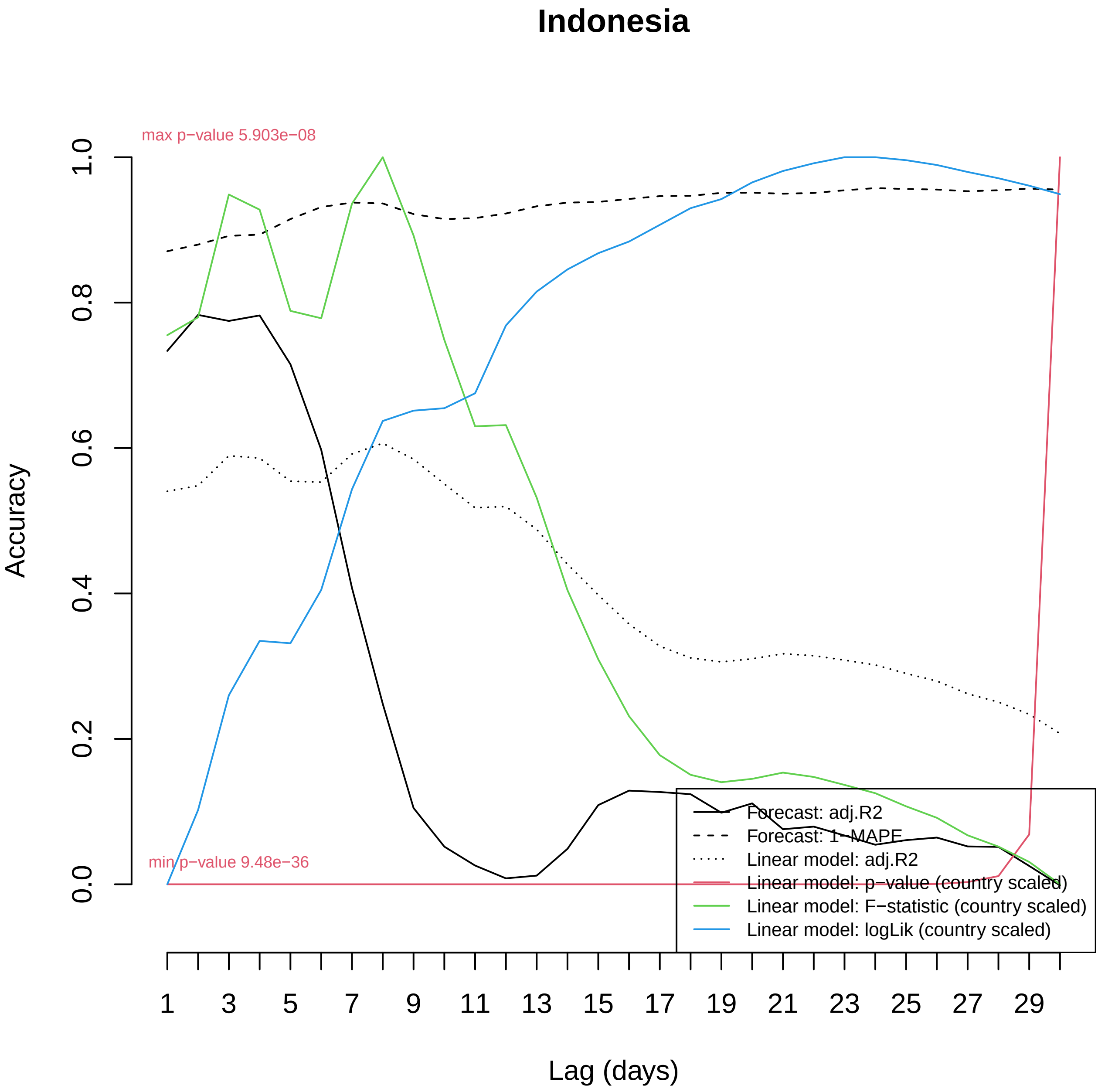
Indonesia
max p−value 5.903e−08
min p−value 9.48e−36
Accuracy
1.0
0.8
0.6
0.4
0.2
0.0
1
3
5
7
9
11
13
15
17
19
21
23
25
27
29
Lag (days)
Forecast: adj.R2
Forecast: 1−MAPE
Linear model: adj.R2
Linear model: p−value (country scaled)
Linear model: F−statistic (country scaled)
Linear model: logLik (country scaled)

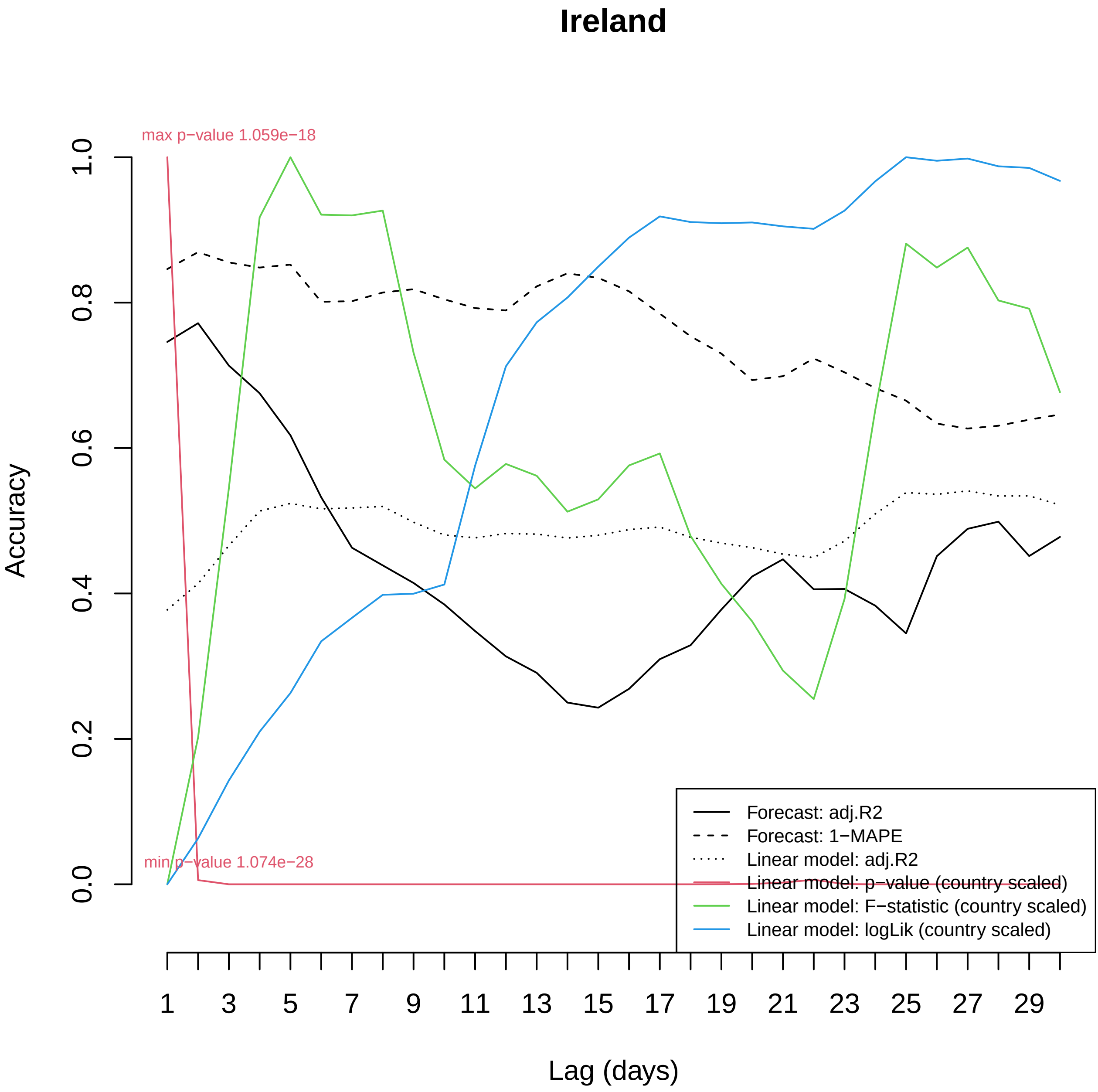

Ireland
max p−value 1.059e−18
min p−value 1.074e−28
Accuracy
0.0
0.2
0.4
0.6
0.8
1.0
Lag (days)
1
3
5
7
9
11
13
15
17
19
21
23
25
27
29
Forecast: adj.R2
Forecast: 1−MAPE
Linear model: adj.R2
Linear model: p−value (country scaled)
Linear model: F−statistic (country scaled)
Linear model: logLik (country scaled)

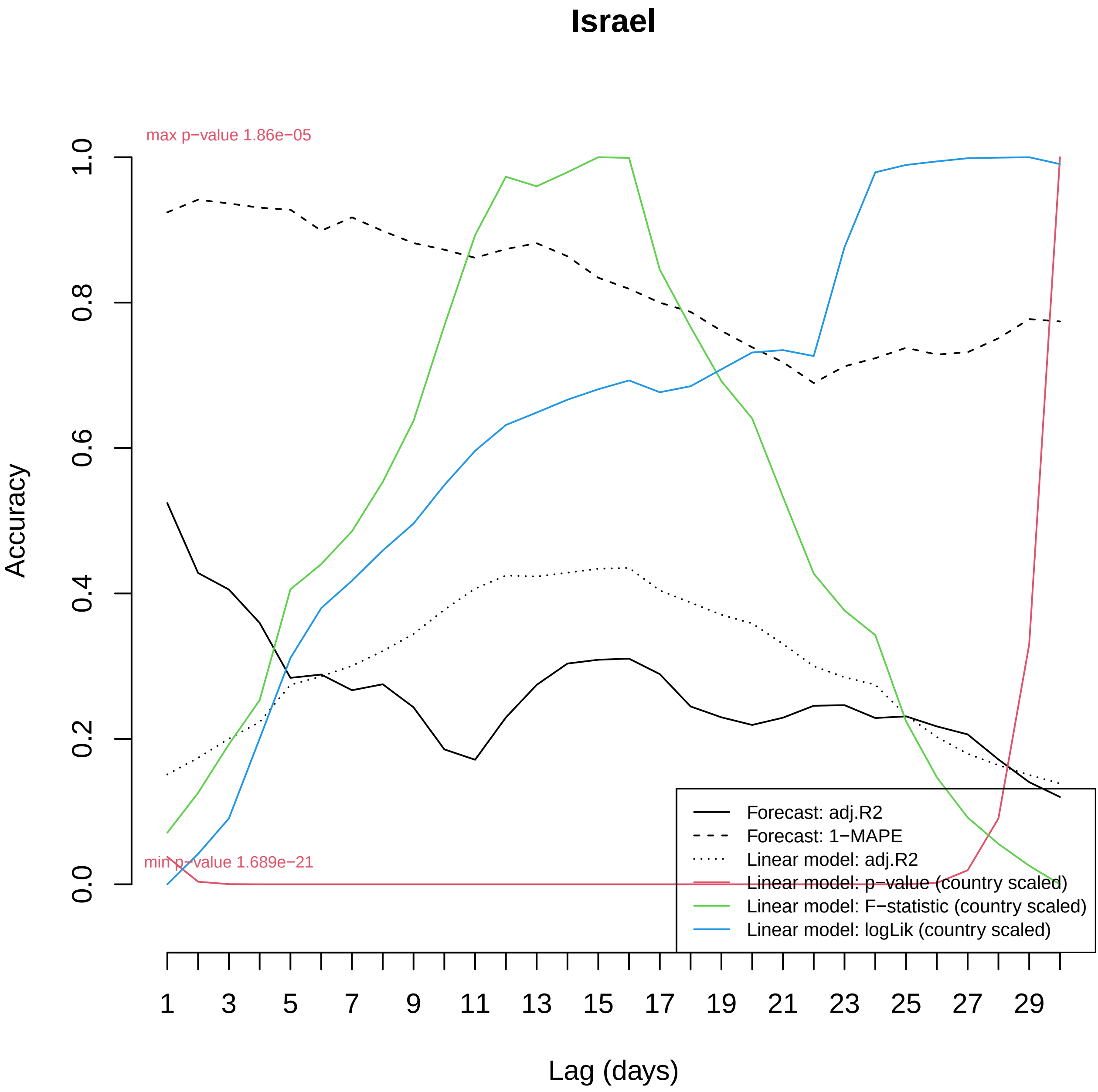

Israel
max p−value 1.86e−05
min p−value 1.689e−21
Accuracy
0.0
0.2
0.4
0.6
0.8
1.0
1
3
5
7
9
11
13
15
17
19
21
23
25
27
29
Lag (days)
Forecast: adj.R2
Forecast: 1−MAPE
Linear model: adj.R2
Linear model: p−value (country scaled)
Linear model: F−statistic (country scaled)
Linear model: logLik (country scaled)

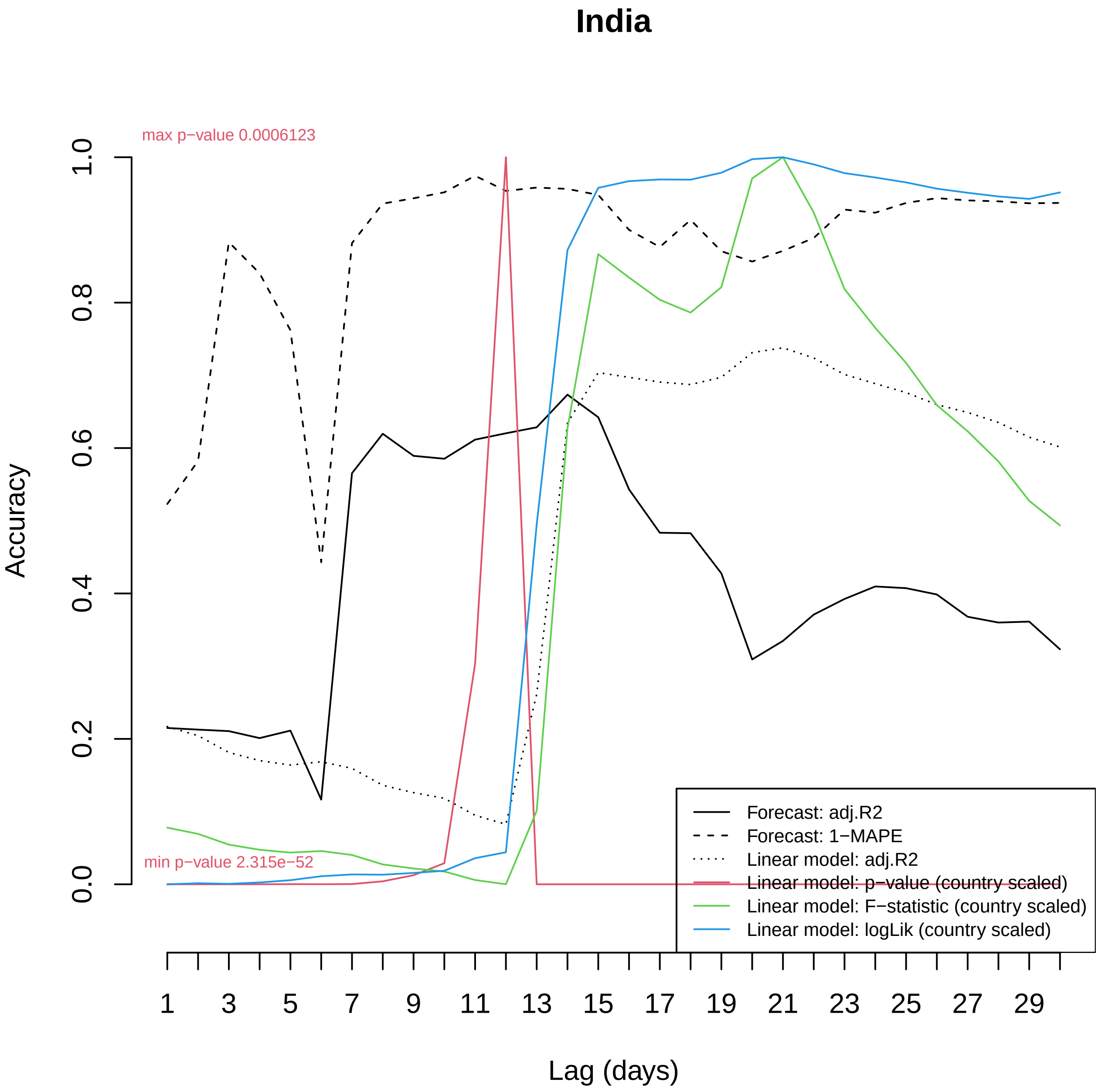

India
max p−value 0.0006123
min p−value 2.315e−52
Accuracy
0.0
0.2
0.4
0.6
0.8
1.0
1
3
5
7
9
11
13
15
17
19
21
23
25
27
29
Lag (days)
Forecast: adj.R2
Forecast: 1−MAPE
Linear model: adj.R2
Linear model: p−value (country scaled)
Linear model: F−statistic (country scaled)
Linear model: logLik (country scaled)

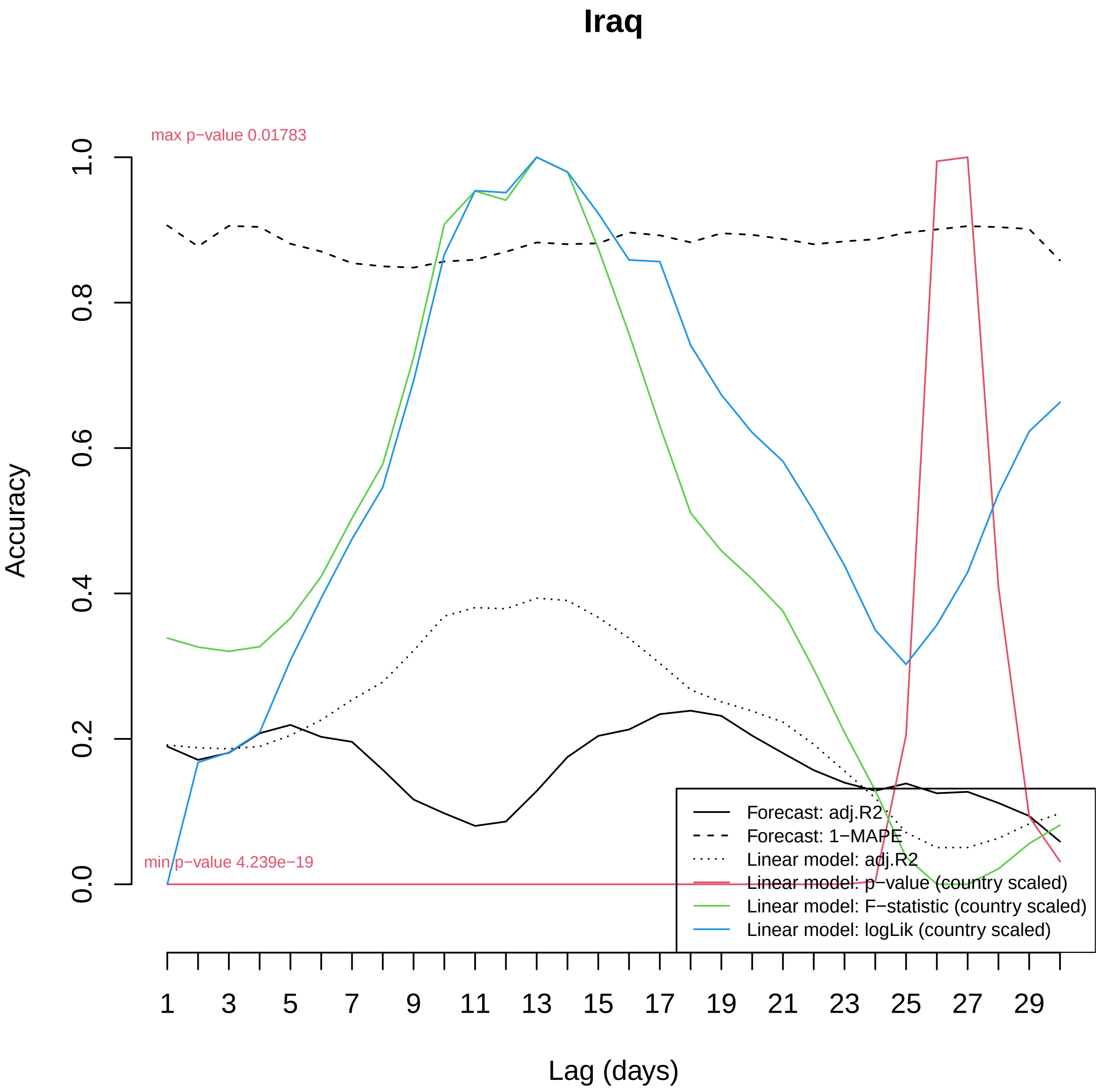

Iraq
max p−value 0.01783
min p−value 4.239e−19
Accuracy
1.0
0.8
0.6
0.4
0.2
0.0
1
3
5
7
9
11
13
15
17
19
21
23
25
27
29
Lag (days)
Forecast: adj.R2
Forecast: 1−MAPE
Linear model: adj.R2
Linear model: p−value (country scaled)
Linear model: F−statistic (country scaled)
Linear model: logLik (country scaled)

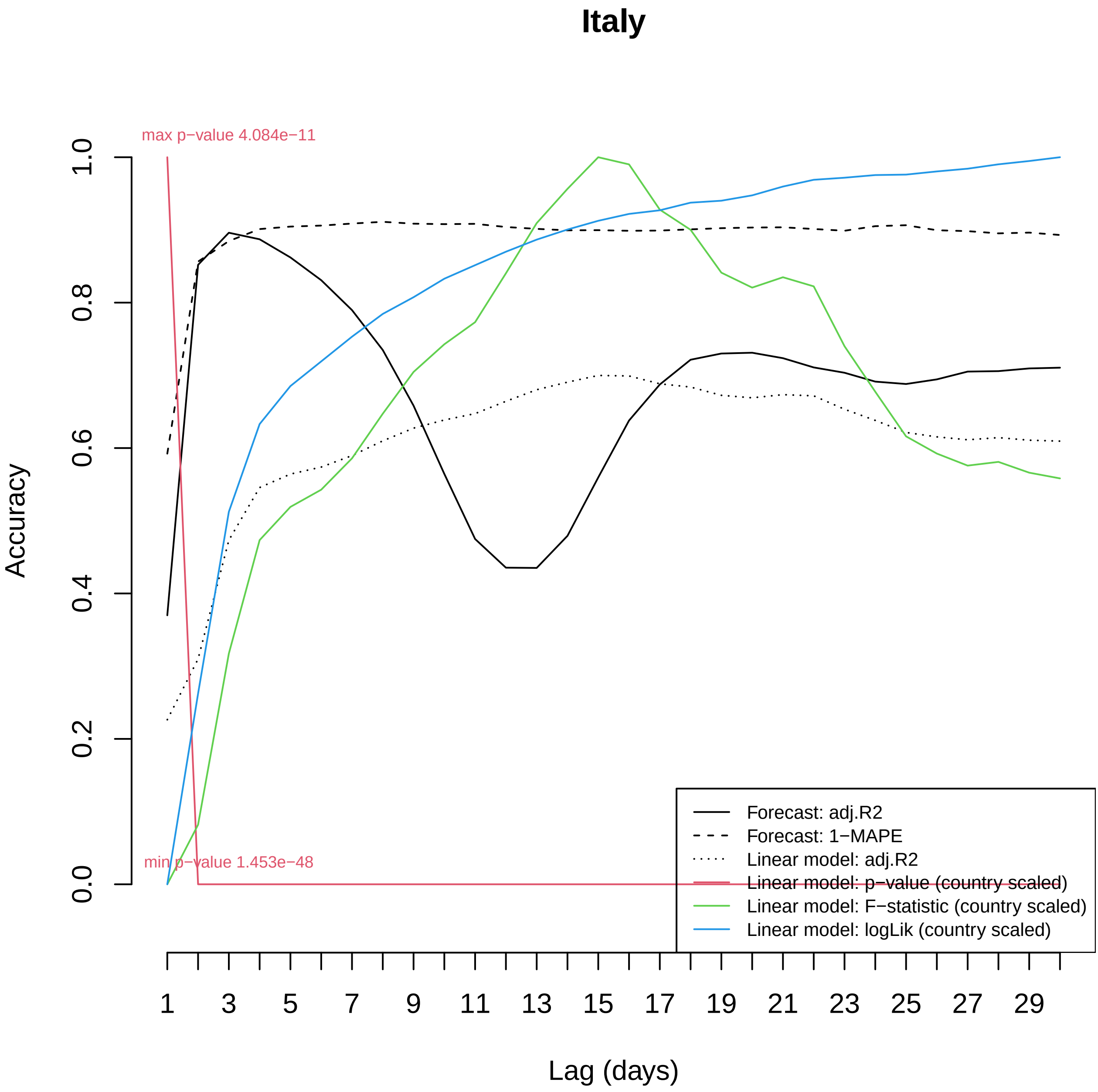

Italy
Accuracy
Lag (days)
max p−value 4.084e−11
min p−value 1.453e−48
1.0
0.8
0.6
0.4
0.2
0.0
1
3
5
7
9
11
13
15
17
19
21
23
25
27
29
Forecast: adj.R2
Forecast: 1−MAPE
Linear model: adj.R2
Linear model: p−value (country scaled)
Linear model: F−statistic (country scaled)
Linear model: logLik (country scaled)

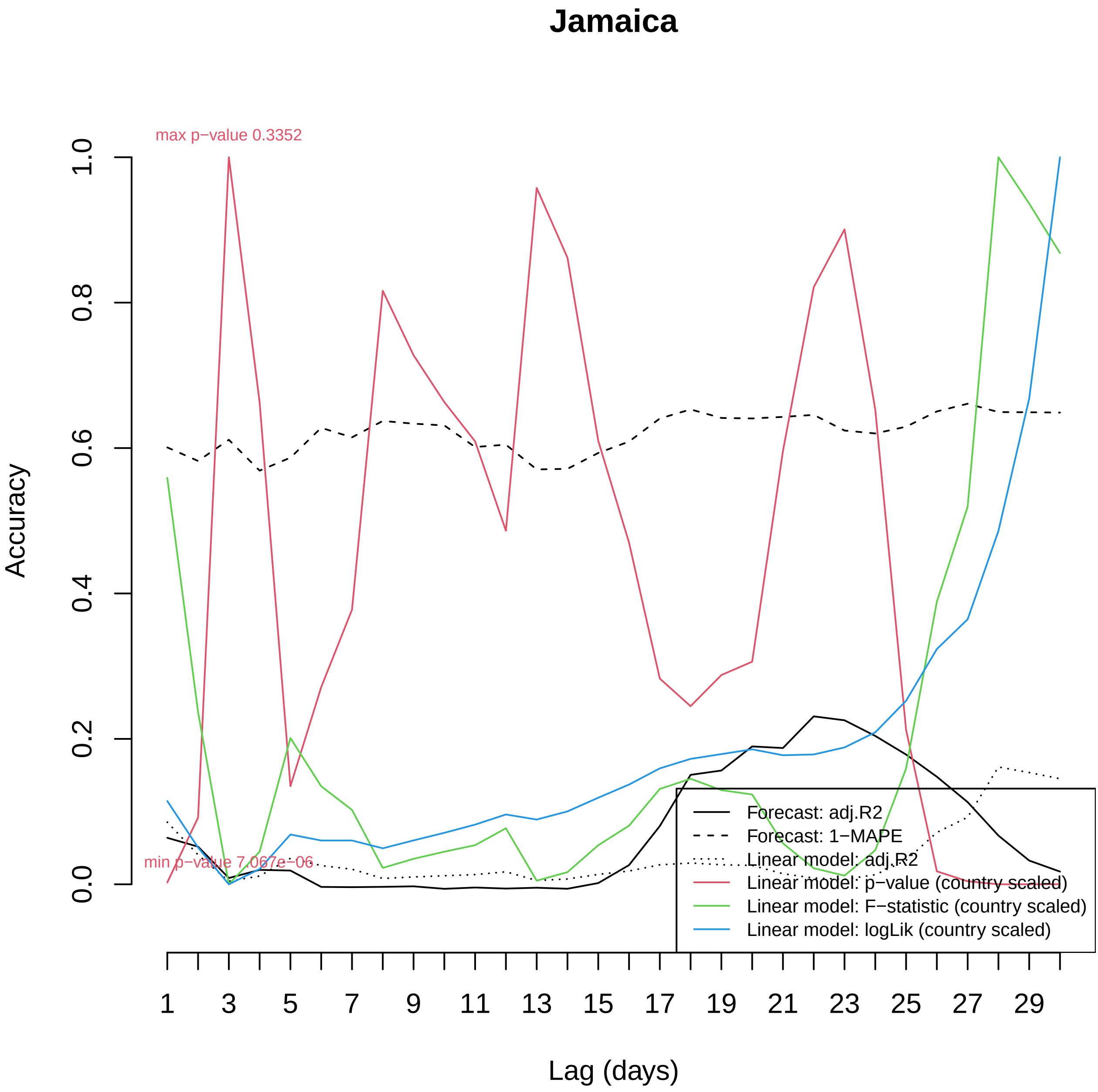

Jamaica
max p−value 0.3352
min p−value 7.067e−06
Accuracy
1.0
0.8
0.6
0.4
0.2
0.0
Lag (days)
1
3
5
7
9
11
13
15
17
19
21
23
25
27
29
Forecast: adj.R2
Forecast: 1−MAPE
Linear model: adj.R2
Linear model: p−value (country scaled)
Linear model: F−statistic (country scaled)
Linear model: logLik (country scaled)

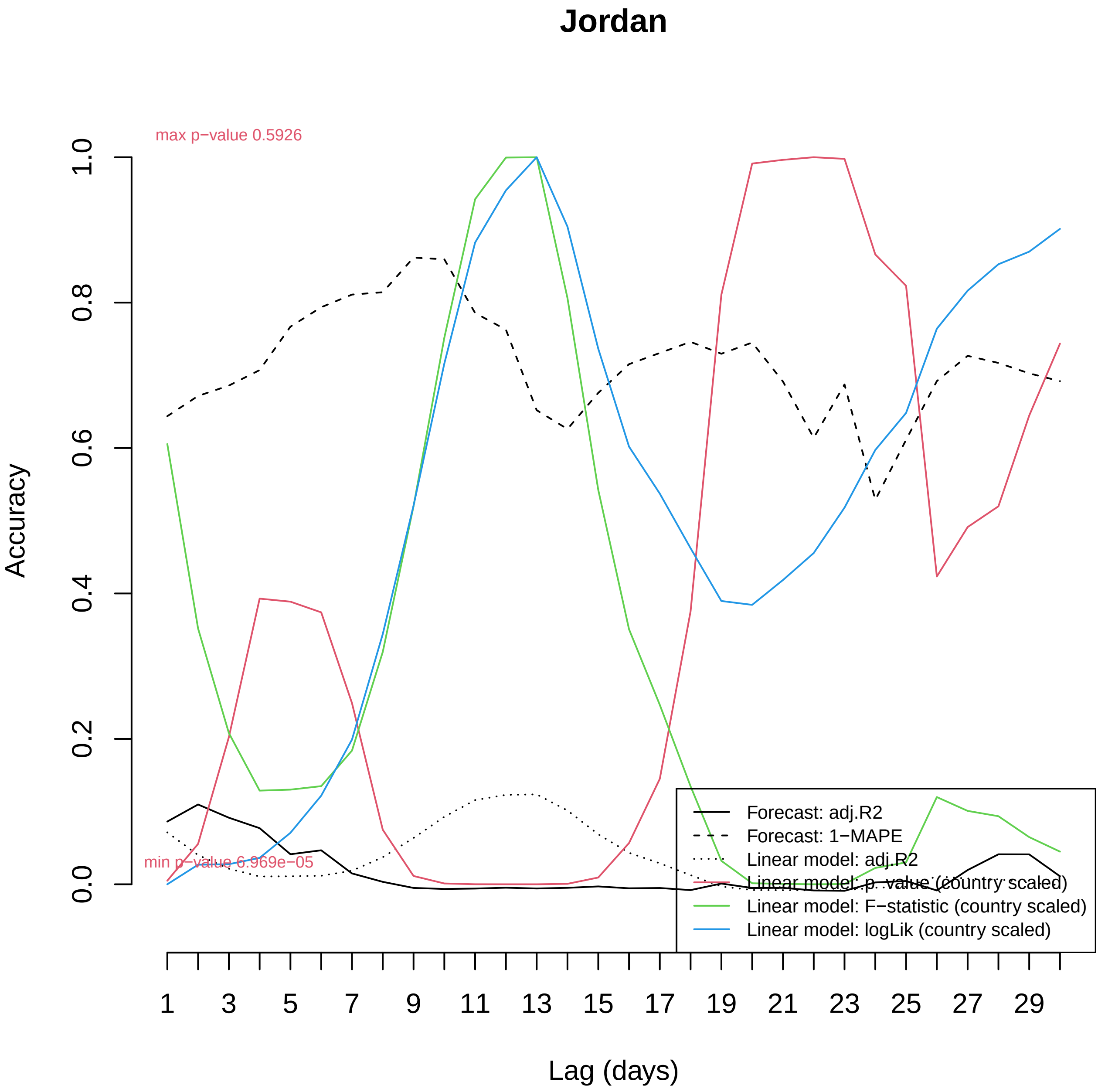

Jordan
max p−value 0.5926
min p−value 6.969e−05
Accuracy
0.0
0.2
0.4
0.6
0.8
1.0
Lag (days)
1
3
5
7
9
11
13
15
17
19
21
23
25
27
29
Forecast: adj.R2
Forecast: 1−MAPE
Linear model: adj.R2
Linear model: p−value (country scaled)
Linear model: F−statistic (country scaled)
Linear model: logLik (country scaled)

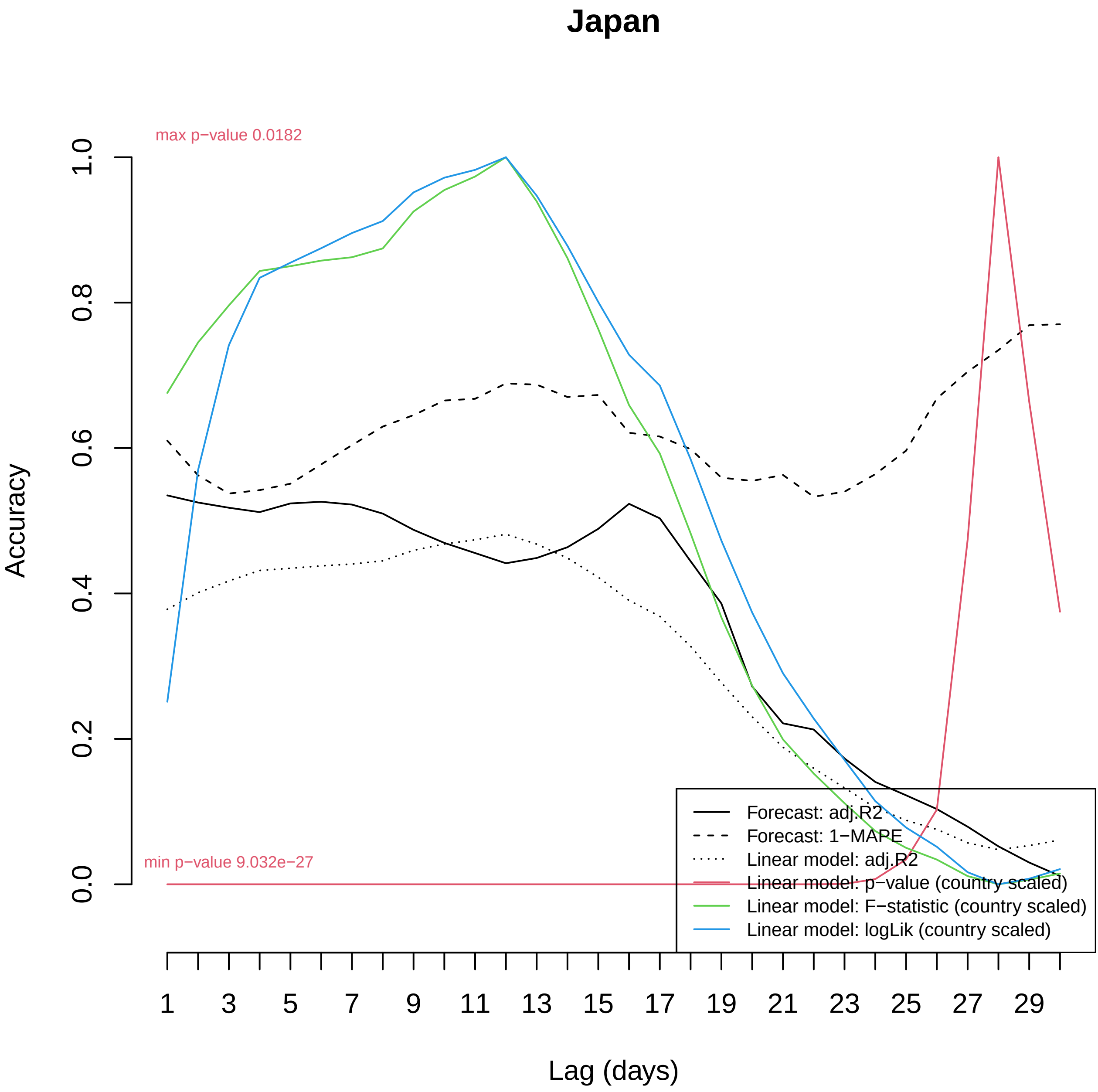

Japan
max p−value 0.0182
min p−value 9.032e−27
Accuracy
0.0
0.2
0.4
0.6
0.8
1.0
Lag (days)
1
3
5
7
9
11
13
15
17
19
21
23
25
27
29
Forecast: adj.R2
Forecast: 1−MAPE
Linear model: adj.R2
Linear model: p−value (country scaled)
Linear model: F−statistic (country scaled)
Linear model: logLik (country scaled)

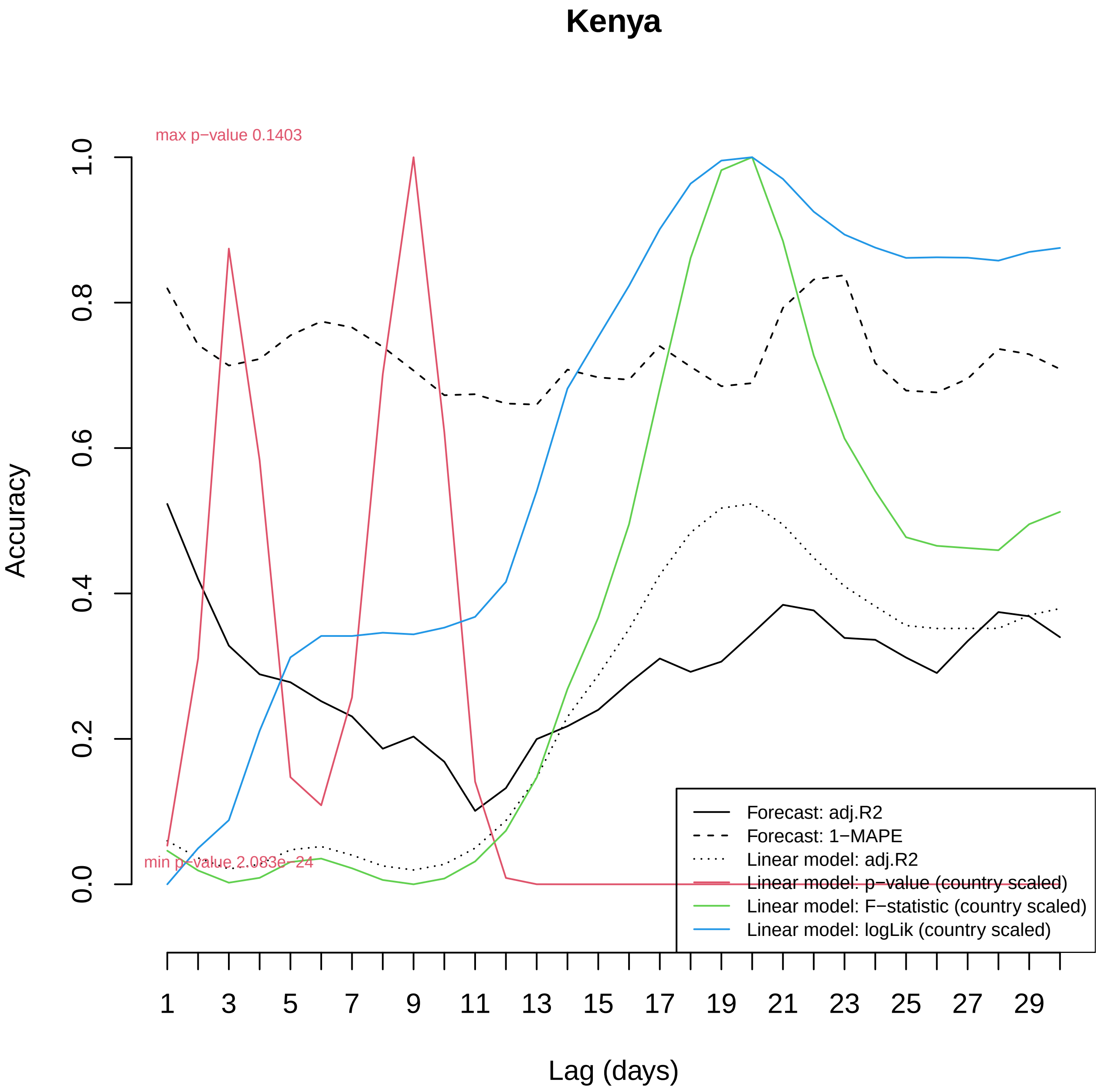

Kenya
max p−value 0.1403
min p−value 2.083e−24
Accuracy
0.0
0.2
0.4
0.6
0.8
1.0
Lag (days)
1
3
5
7
9
11
13
15
17
19
21
23
25
27
29
Forecast: adj.R2
Forecast: 1−MAPE
Linear model: adj.R2
Linear model: p−value (country scaled)
Linear model: F−statistic (country scaled)
Linear model: logLik (country scaled)

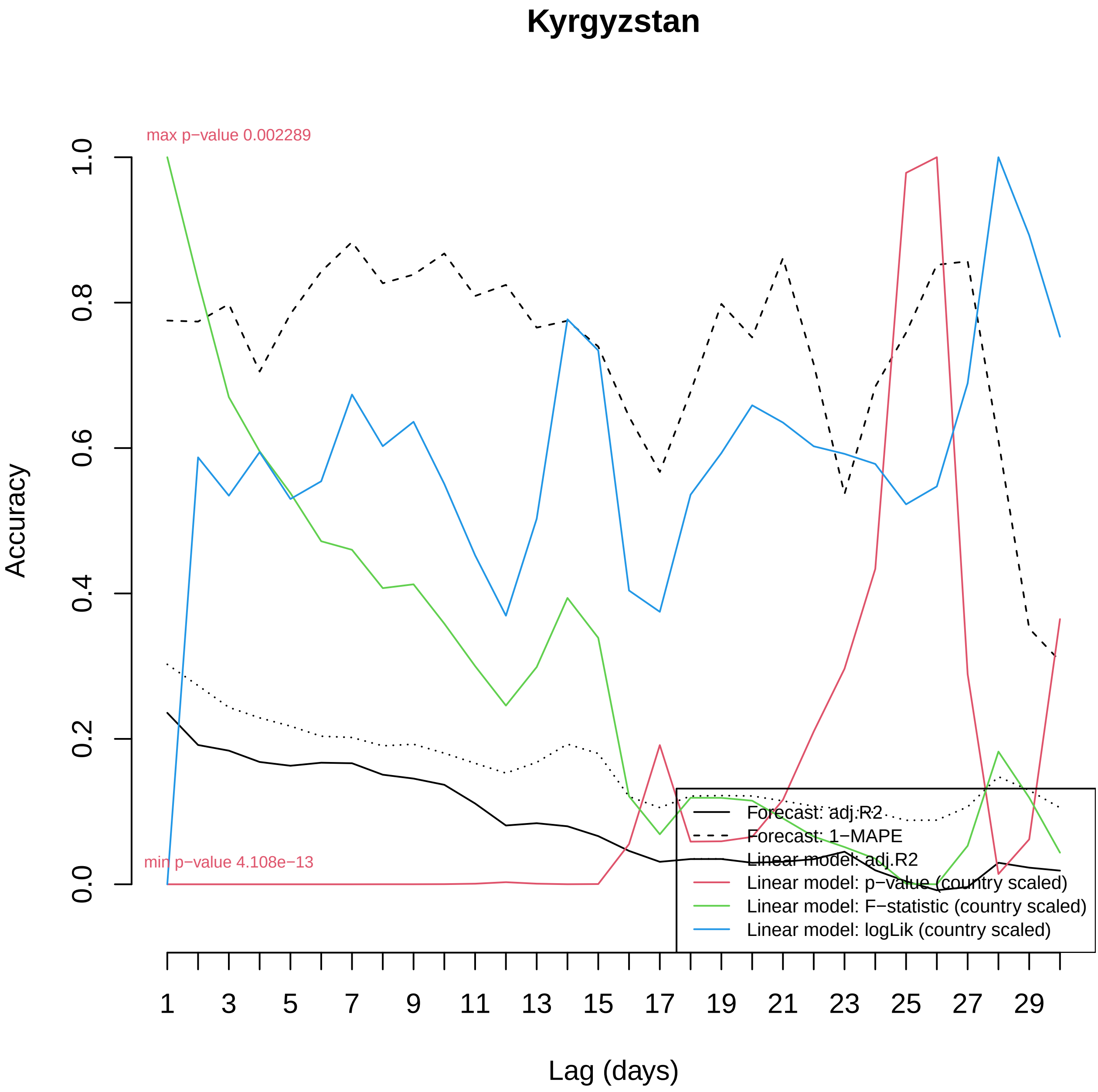

Kyrgyzstan
max p−value 0.002289
min p−value 4.108e−13
Accuracy
0.0
0.2
0.4
0.6
0.8
1.0
Lag (days)
1
3
5
7
9
11
13
15
17
19
21
23
25
27
29
Forecast: adj.R2
Forecast: 1−MAPE
Linear model: adj.R2
Linear model: p−value (country scaled)
Linear model: F−statistic (country scaled)
Linear model: logLik (country scaled)

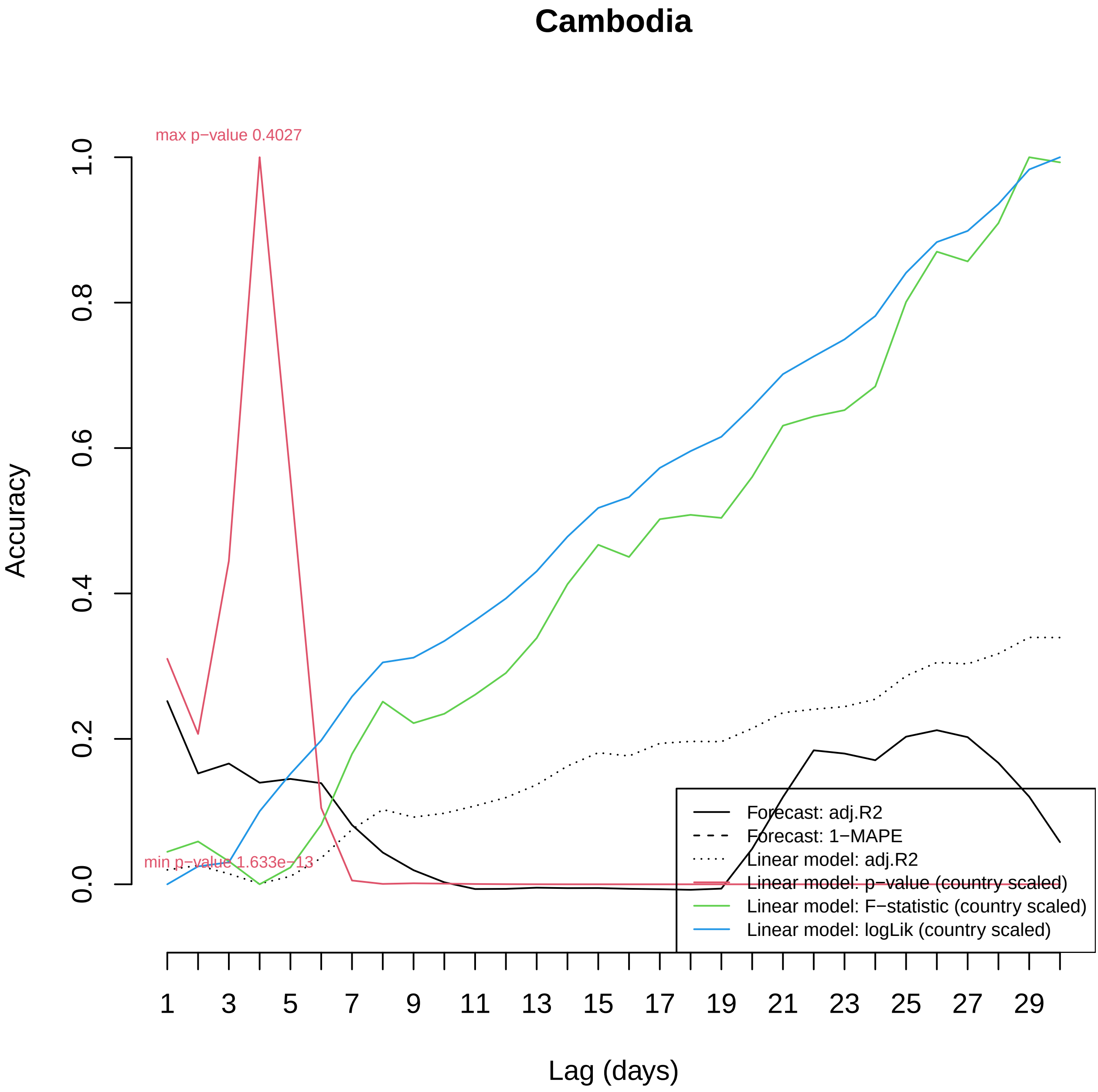
Cambodia
max p−value 0.4027
min p−value 1.633e−13
Accuracy
0.0
0.2
0.4
0.6
0.8
1.0
Lag (days)
1
3
5
7
9
11
13
15
17
19
21
23
25
27
29
Forecast: adj.R2
Forecast: 1−MAPE
Linear model: adj.R2
Linear model: p−value (country scaled)
Linear model: F−statistic (country scaled)
Linear model: logLik (country scaled)

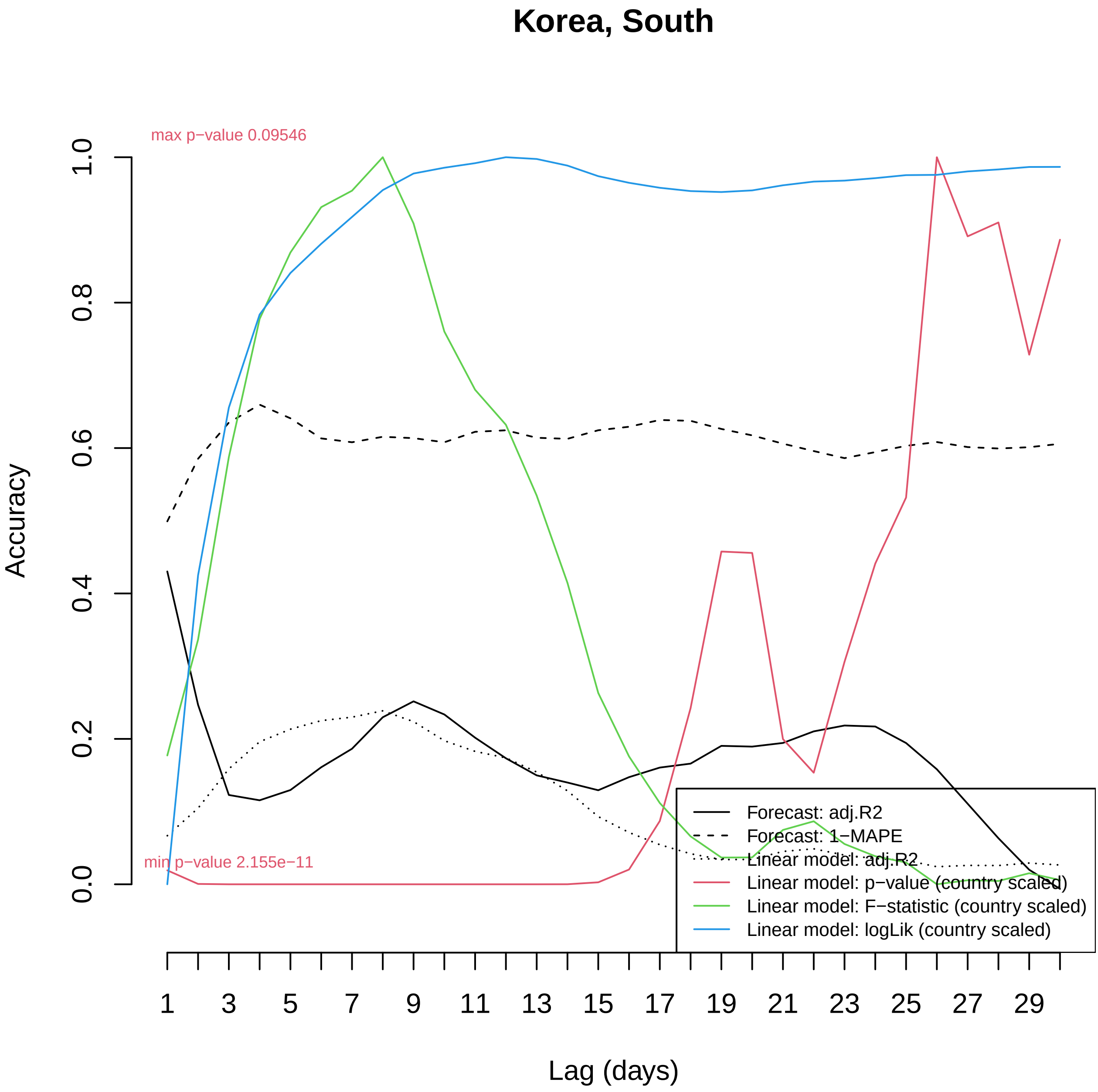

Korea, South
max p−value 0.09546
min p−value 2.155e−11
Accuracy
0.0
0.2
0.4
0.6
0.8
1.0
Lag (days)
1
3
5
7
9
11
13
15
17
19
21
23
25
27
29
Forecast: adj.R2
Forecast: 1−MAPE
Linear model: adj.R2
Linear model: p−value (country scaled)
Linear model: F−statistic (country scaled)
Linear model: logLik (country scaled)

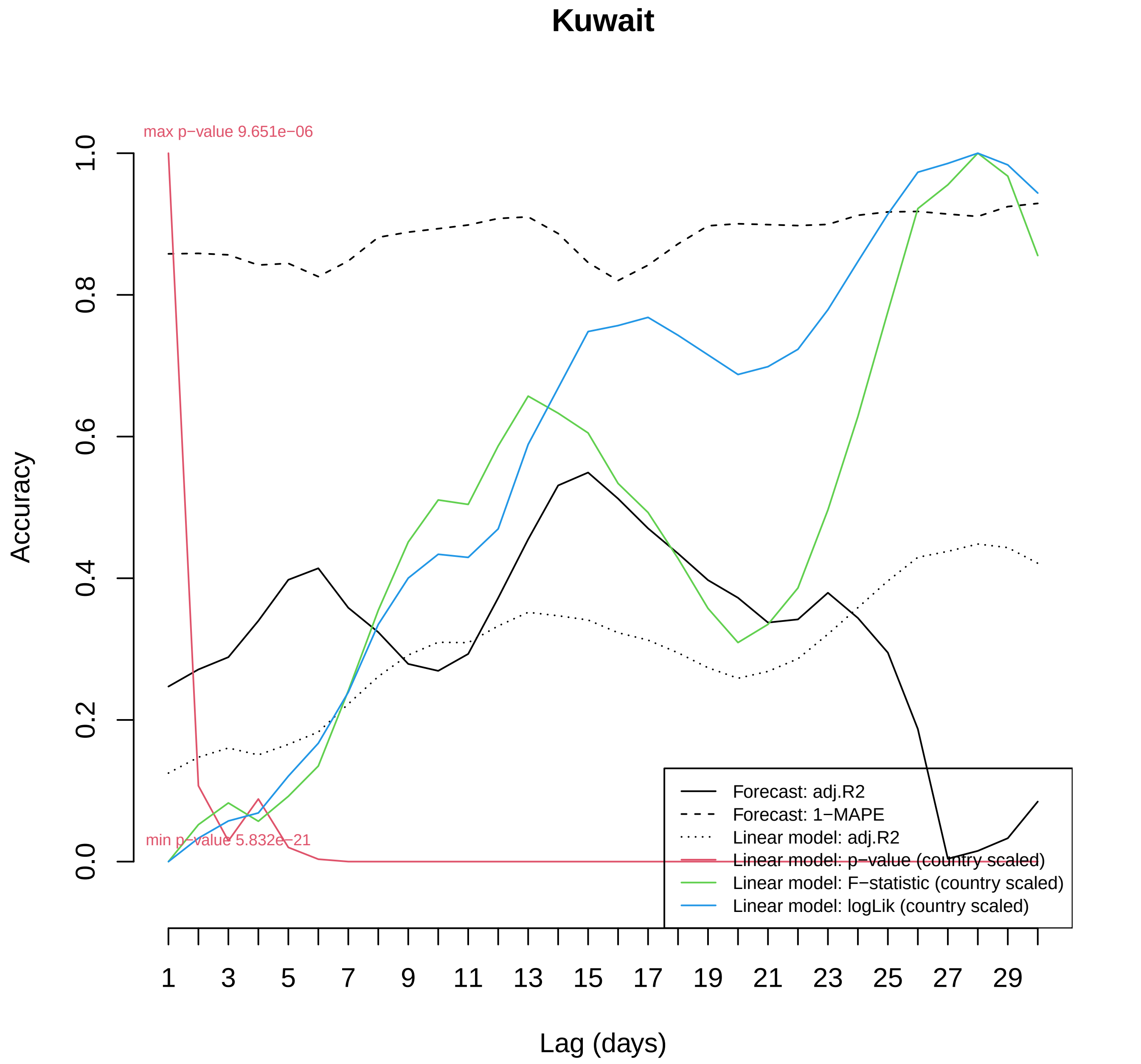

Kuwait
max p−value 9.651e−06
min p−value 5.832e−21
Accuracy
0.0
0.2
0.4
0.6
0.8
1.0
Lag (days)
1
3
5
7
9
11
13
15
17
19
21
23
25
27
29
Forecast: adj.R2
Forecast: 1−MAPE
Linear model: adj.R2
Linear model: p−value (country scaled)
Linear model: F−statistic (country scaled)
Linear model: logLik (country scaled)

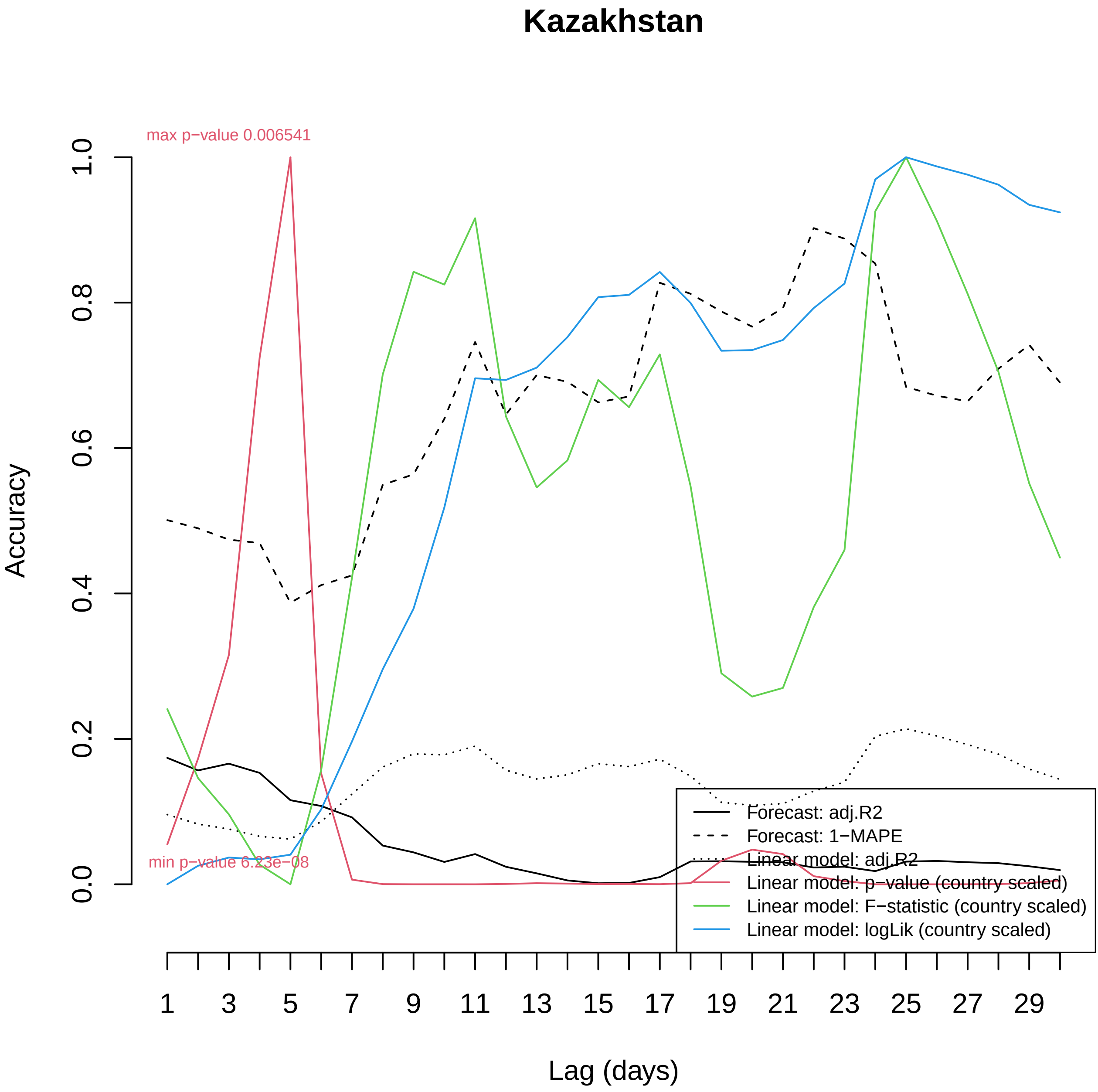

Kazakhstan
max p−value 0.006541
min p−value 6.23e−08
Accuracy
0.0
0.2
0.4
0.6
0.8
1.0
Lag (days)
1
3
5
7
9
11
13
15
17
19
21
23
25
27
29
Forecast: adj.R2
Forecast: 1−MAPE
Linear model: adj.R2
Linear model: p−value (country scaled)
Linear model: F−statistic (country scaled)
Linear model: logLik (country scaled)

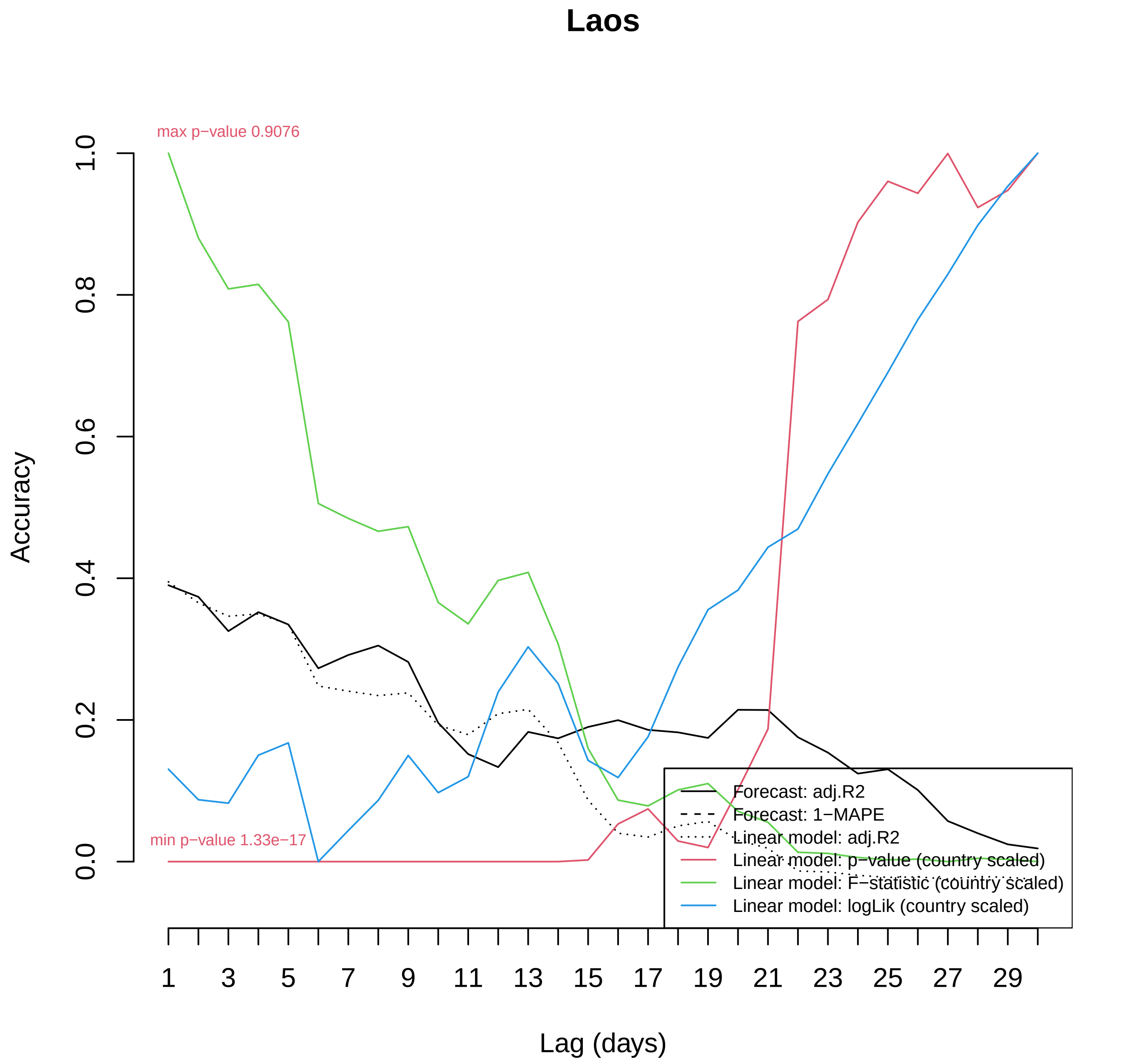

Laos
max p−value 0.9076
min p−value 1.33e−17
Accuracy
0.0
0.2
0.4
0.6
0.8
1.0
Lag (days)
1
3
5
7
9
11
13
15
17
19
21
23
25
27
29
Forecast: adj.R2
Forecast: 1−MAPE
Linear model: adj.R2
Linear model: p−value (country scaled)
Linear model: F−statistic (country scaled)
Linear model: logLik (country scaled)

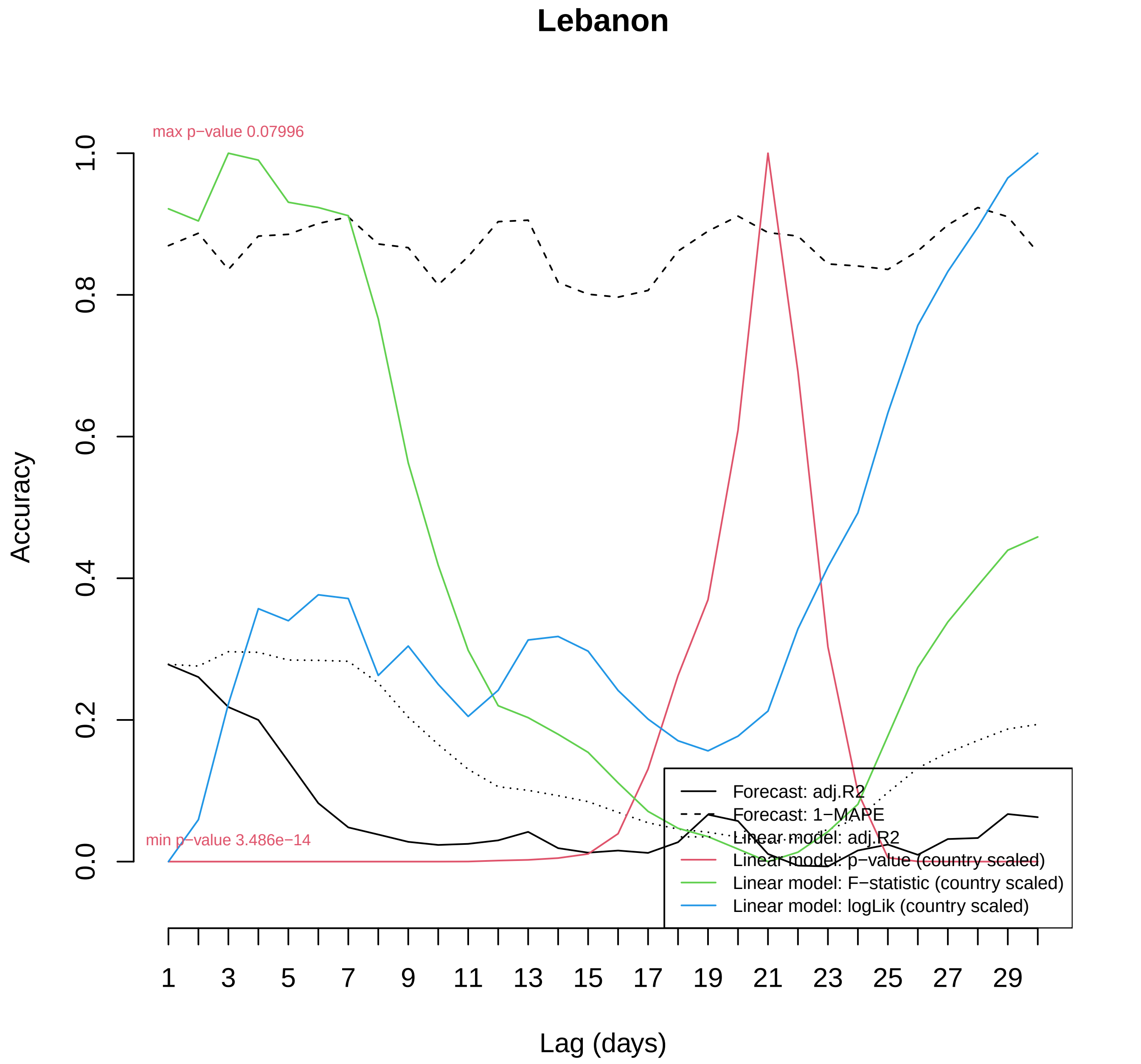
Lebanon
max p−value 0.07996
min p−value 3.486e−14
Accuracy
0.0
0.2
0.4
0.6
0.8
1.0
1
3
5
7
9
11
13
15
17
19
21
23
25
27
29
Lag (days)
Forecast: adj.R2
Forecast: 1−MAPE
Linear model: adj.R2
Linear model: p−value (country scaled)
Linear model: F−statistic (country scaled)
Linear model: logLik (country scaled)

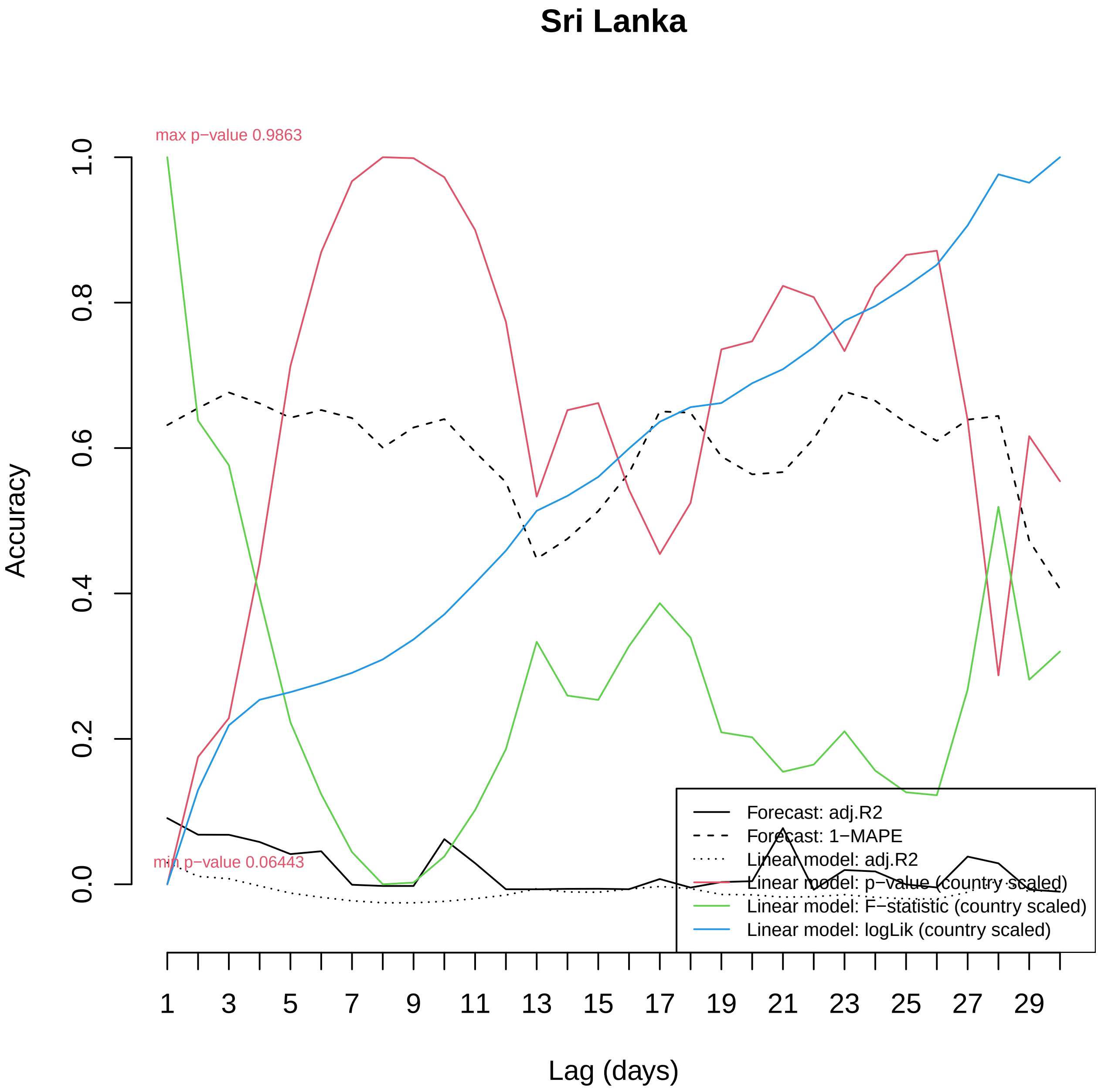

Sri Lanka
max p−value 0.9863
min p−value 0.06443
Accuracy
0.0
0.2
0.4
0.6
0.8
1.0
Lag (days)
1
3
5
7
9
11
13
15
17
19
21
23
25
27
29
Forecast: adj.R2
Forecast: 1−MAPE
Linear model: adj.R2
Linear model: p−value (country scaled)
Linear model: F−statistic (country scaled)
Linear model: logLik (country scaled)

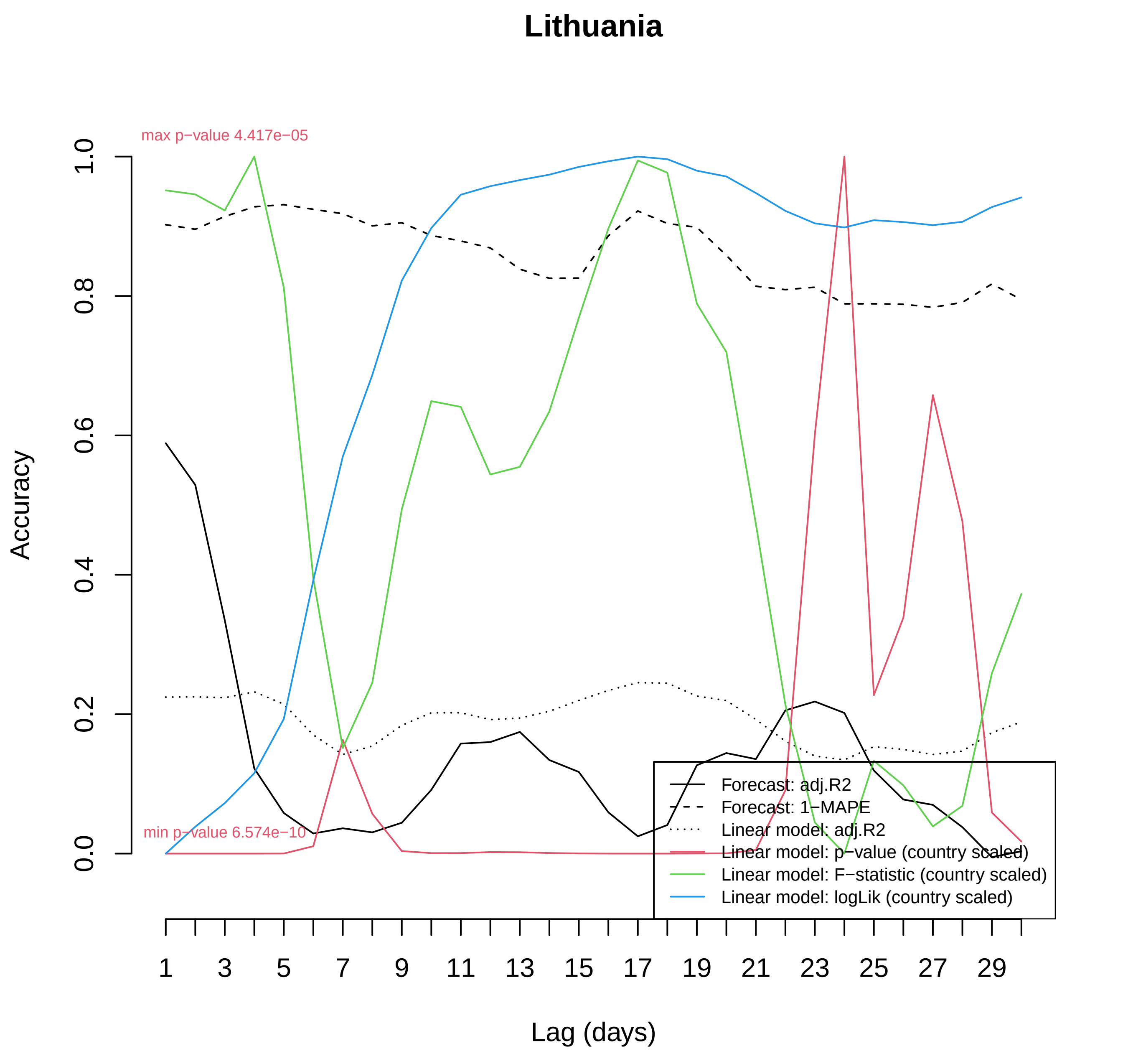
Lithuania
max p−value 4.417e−05
min p−value 6.574e−10
Accuracy
0.0
0.2
0.4
0.6
0.8
1.0
Lag (days)
1
3
5
7
9
11
13
15
17
19
21
23
25
27
29
Forecast: adj.R2
Forecast: 1−MAPE
Linear model: adj.R2
Linear model: p−value (country scaled)
Linear model: F−statistic (country scaled)
Linear model: logLik (country scaled)

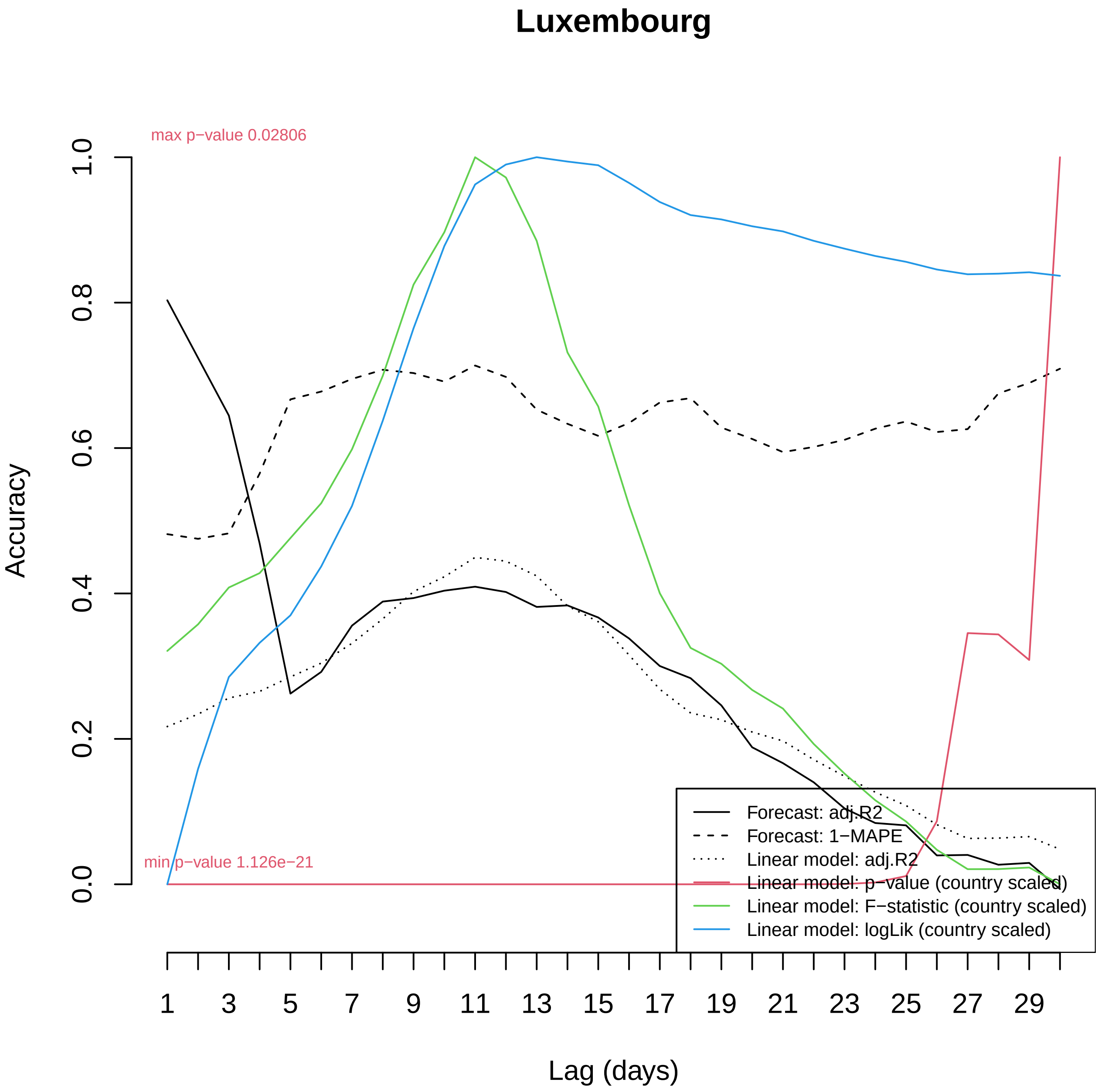

Luxembourg
max p−value 0.02806
min p−value 1.126e−21
Accuracy
0.0
0.2
0.4
0.6
0.8
1.0
Lag (days)
1
3
5
7
9
11
13
15
17
19
21
23
25
27
29
Forecast: adj.R2
Forecast: 1−MAPE
Linear model: adj.R2
Linear model: p−value (country scaled)
Linear model: F−statistic (country scaled)
Linear model: logLik (country scaled)

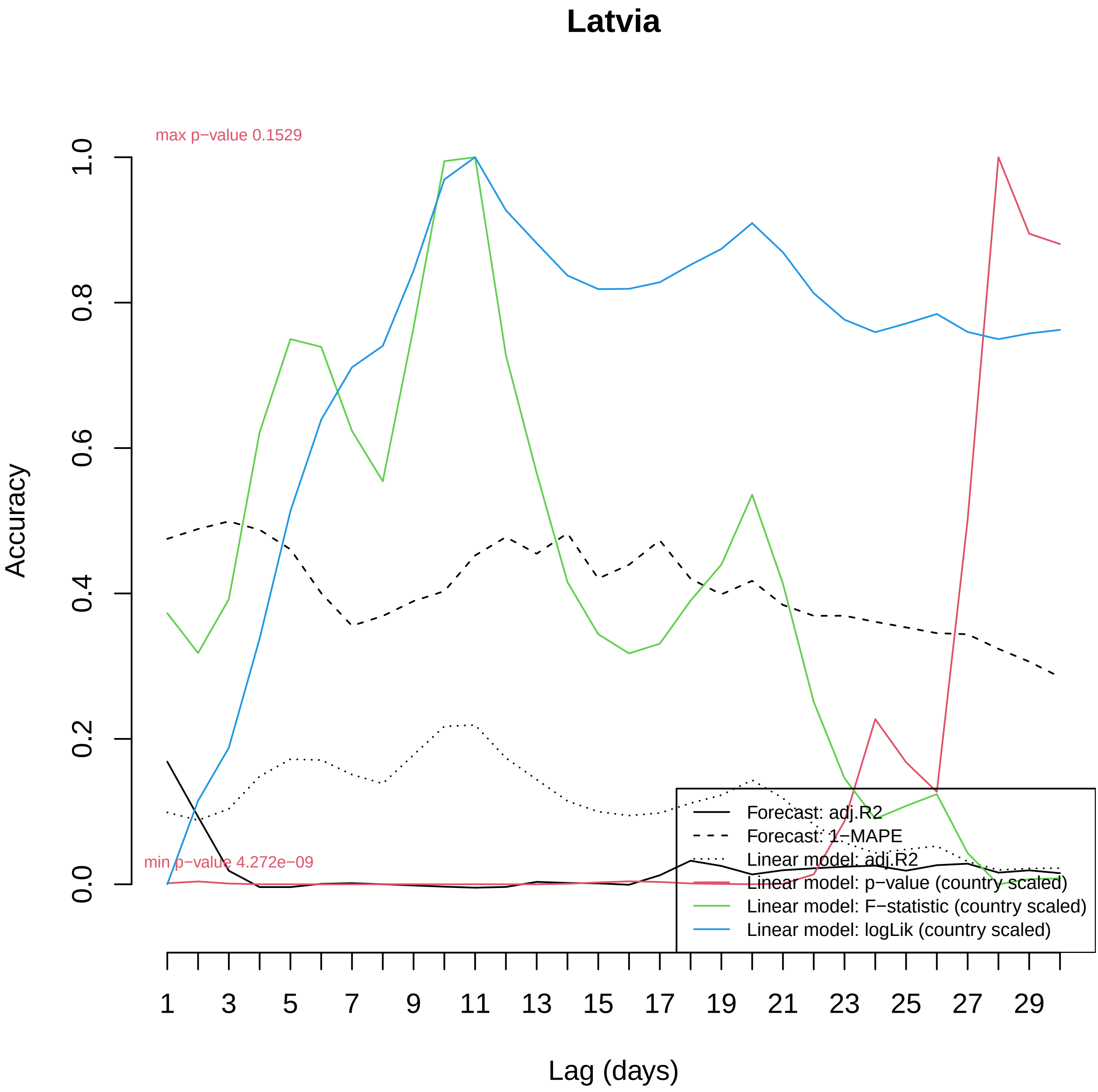

Latvia
max p−value 0.1529
min p−value 4.272e−09
Accuracy
0.0
0.2
0.4
0.6
0.8
1.0
Lag (days)
1
3
5
7
9
11
13
15
17
19
21
23
25
27
29
Forecast: adj.R2
Forecast: 1−MAPE
Linear model: adj.R2
Linear model: p−value (country scaled)
Linear model: F−statistic (country scaled)
Linear model: logLik (country scaled)

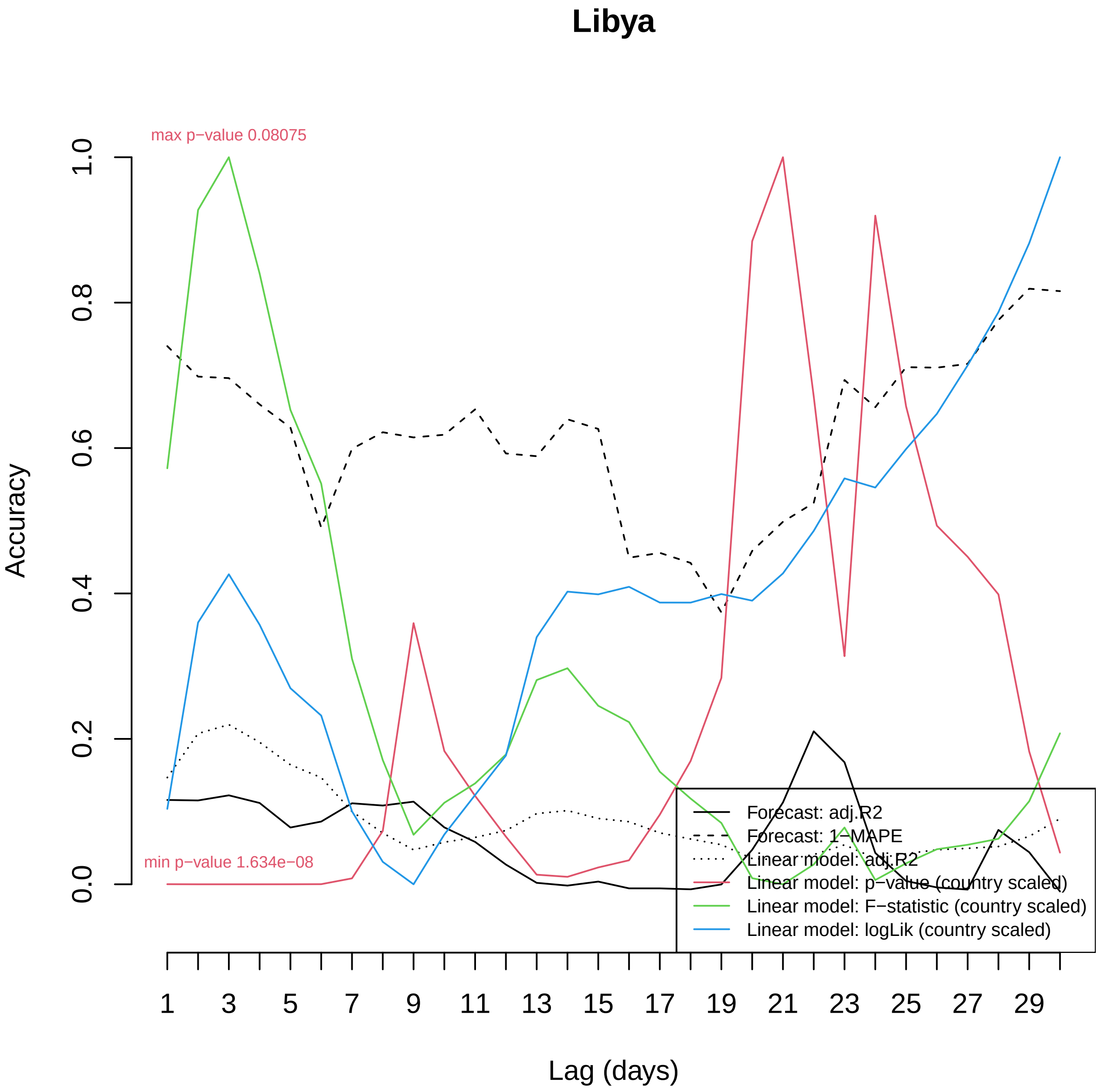

Libya
max p−value 0.08075
min p−value 1.634e−08
Accuracy
0.0
0.2
0.4
0.6
0.8
1.0
1
3
5
7
9
11
13
15
17
19
21
23
25
27
29
Lag (days)
Forecast: adj.R2
Forecast: 1−MAPE
Linear model: adj.R2
Linear model: p−value (country scaled)
Linear model: F−statistic (country scaled)
Linear model: logLik (country scaled)

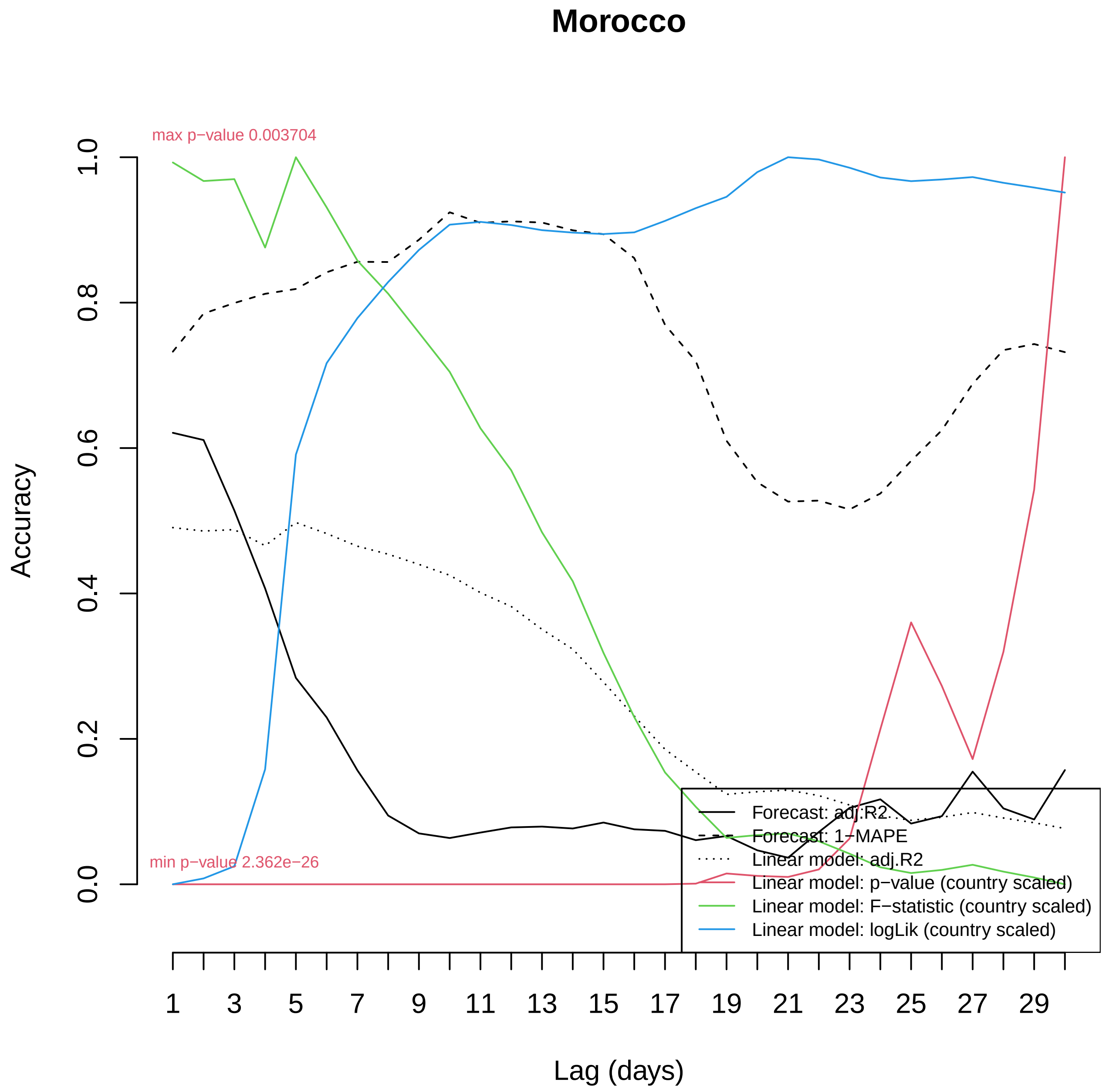

Morocco
max p−value 0.003704
min p−value 2.362e−26
Accuracy
0.0
0.2
0.4
0.6
0.8
1.0
Lag (days)
1
3
5
7
9
11
13
15
17
19
21
23
25
27
29
Forecast: adj.R2
Forecast: 1−MAPE
Linear model: adj.R2
Linear model: p−value (country scaled)
Linear model: F−statistic (country scaled)
Linear model: logLik (country scaled)

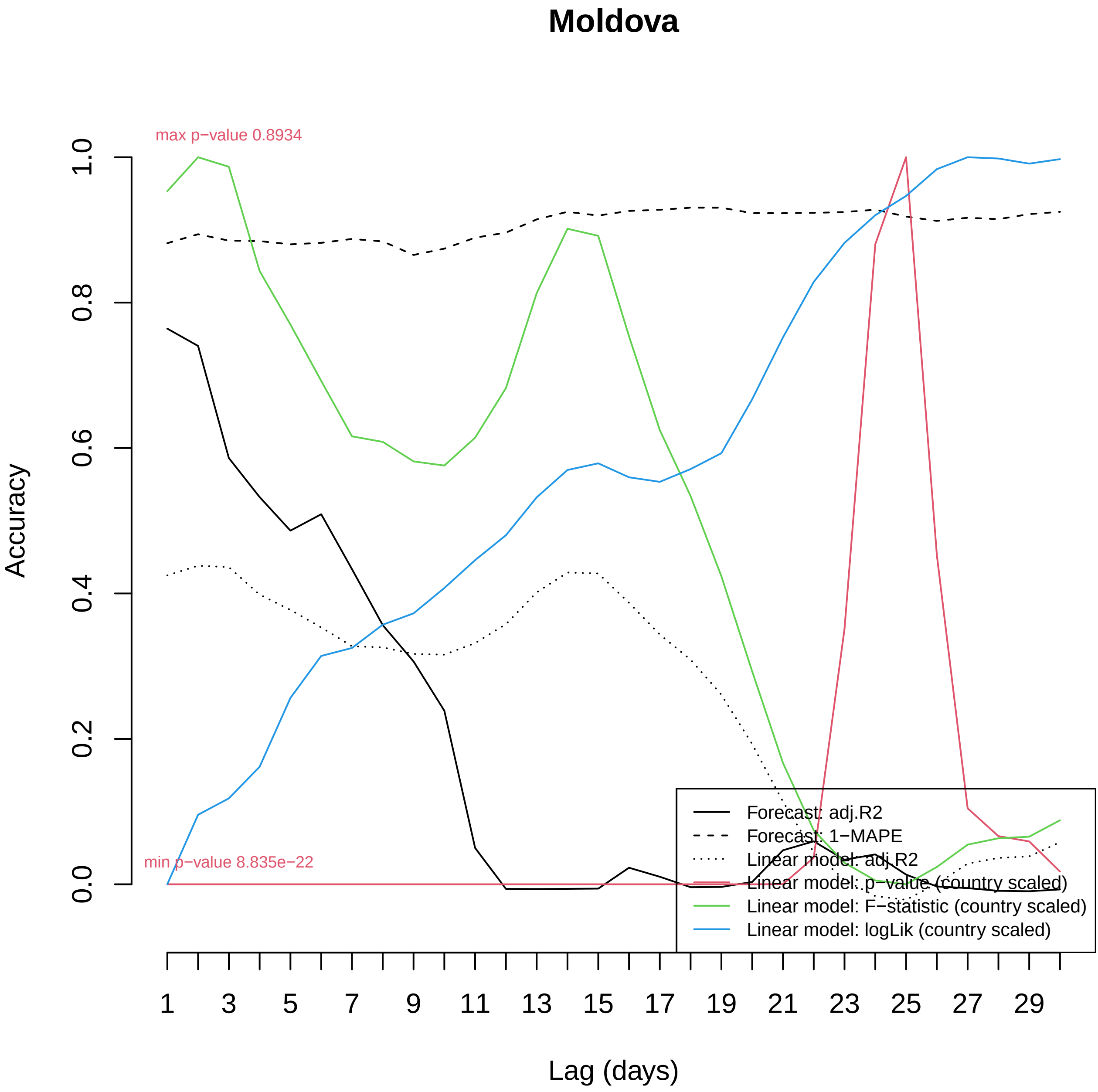

Moldova
max p−value 0.8934
min p−value 8.835e−22
Accuracy
0.0
0.2
0.4
0.6
0.8
1.0
1
3
5
7
9
11
13
15
17
19
21
23
25
27
29
Lag (days)
Forecast: adj.R2
Forecast: 1−MAPE
Linear model: adj.R2
Linear model: p−value (country scaled)
Linear model: F−statistic (country scaled)
Linear model: logLik (country scaled)

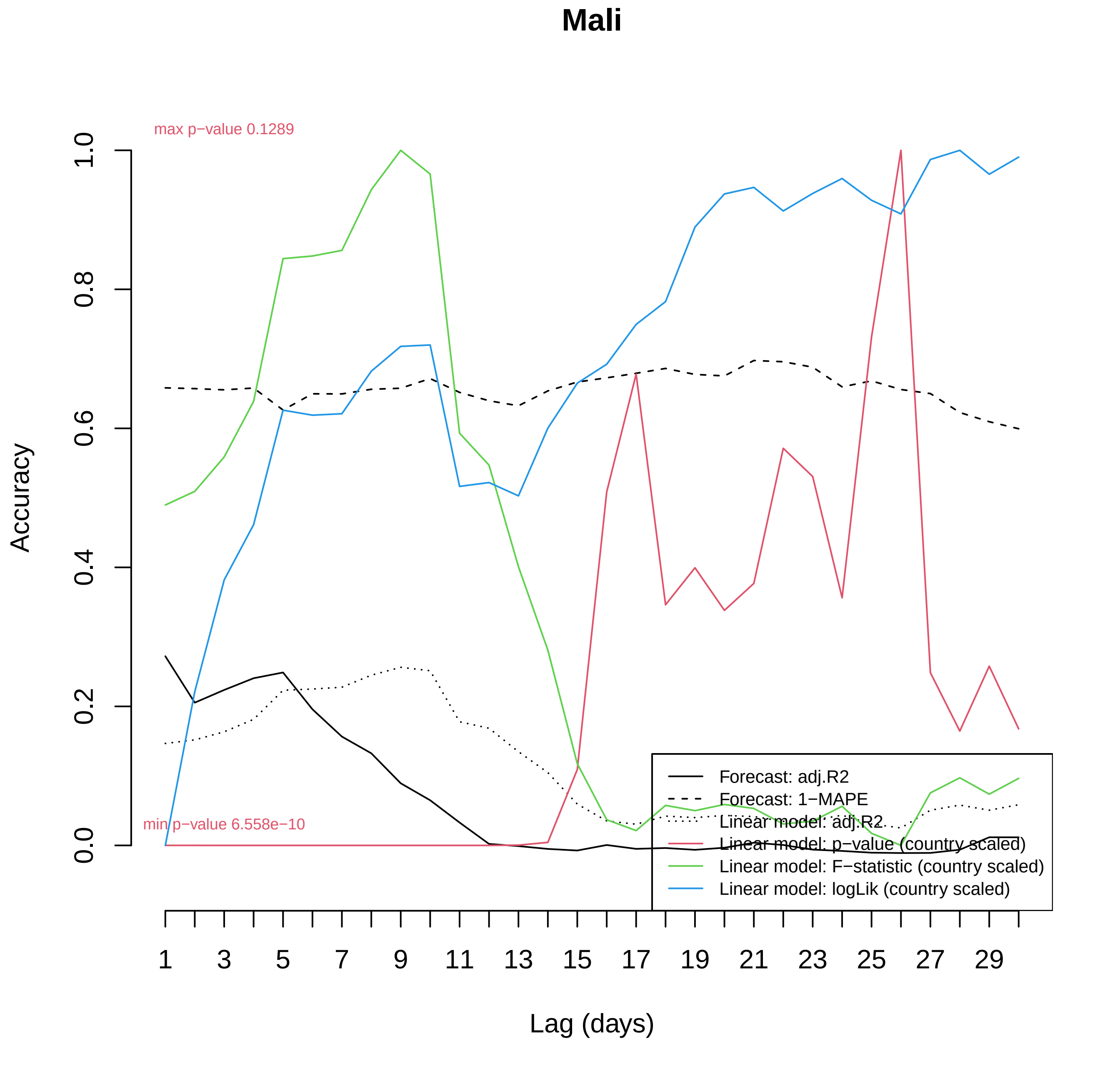

Mali
max p−value 0.1289
min p−value 6.558e−10
Accuracy
0.0
0.2
0.4
0.6
0.8
1.0
Lag (days)
1
3
5
7
9
11
13
15
17
19
21
23
25
27
29
Forecast: adj.R2
Forecast: 1−MAPE
Linear model: adj.R2
Linear model: p−value (country scaled)
Linear model: F−statistic (country scaled)
Linear model: logLik (country scaled)

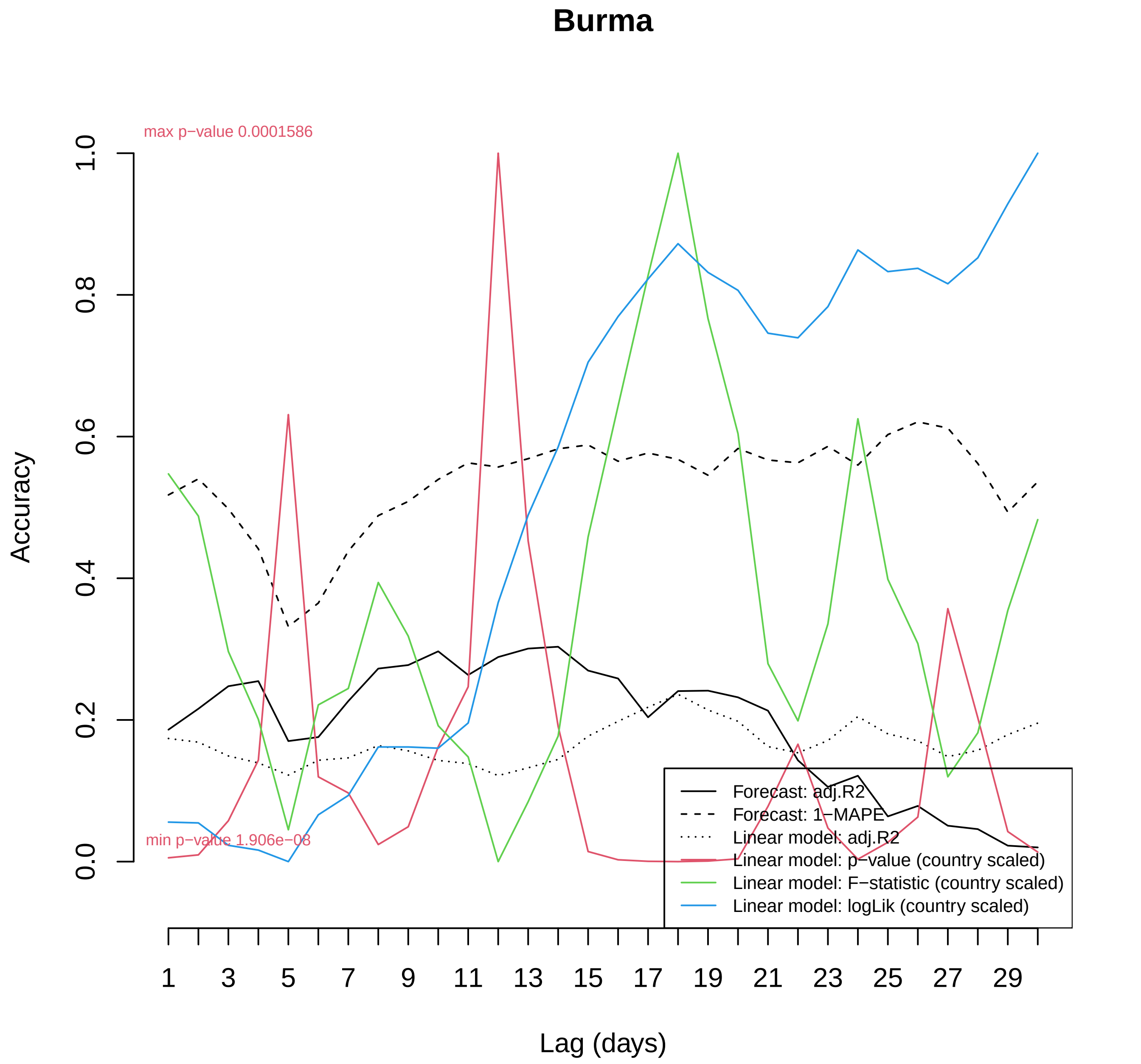

Burma
max p−value 0.0001586
min p−value 1.906e−08
Accuracy
0.0
0.2
0.4
0.6
0.8
1.0
Lag (days)
1
3
5
7
9
11
13
15
17
19
21
23
25
27
29
Forecast: adj.R2
Forecast: 1−MAPE
Linear model: adj.R2
Linear model: p−value (country scaled)
Linear model: F−statistic (country scaled)
Linear model: logLik (country scaled)

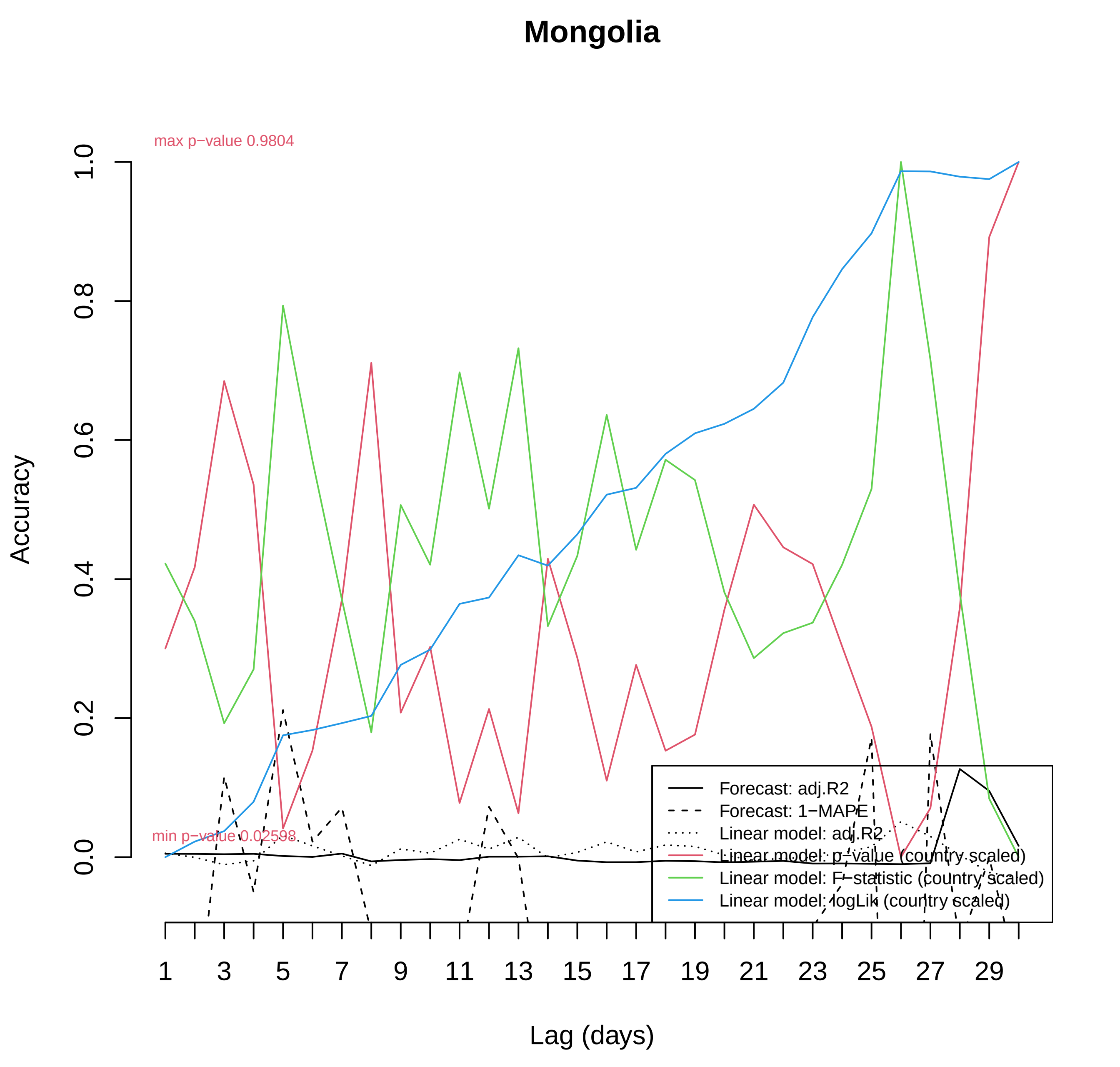

Mongolia
max p−value 0.9804
min p−value 0.02598
Accuracy
0.0
0.2
0.4
0.6
0.8
1.0
Lag (days)
1
3
5
7
9
11
13
15
17
19
21
23
25
27
29
Forecast: adj.R2
Forecast: 1−MAPE
Linear model: adj.R2
Linear model: p−value (country scaled)
Linear model: F−statistic (country scaled)
Linear model: logLik (country scaled)

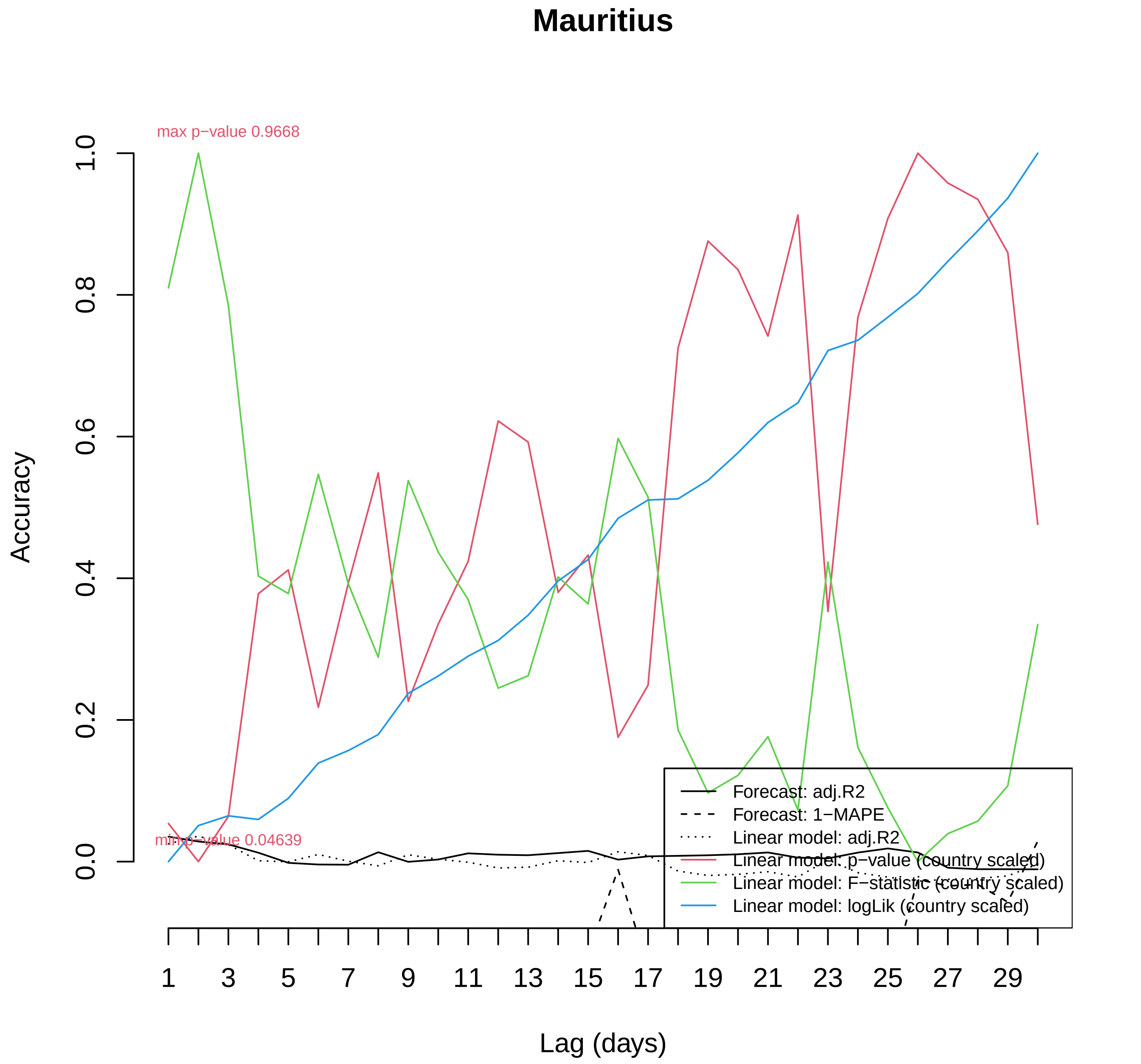
Mauritius
max p−value 0.9668
min p−value 0.04639
Accuracy
0.0
0.2
0.4
0.6
0.8
1.0
Lag (days)
1
3
5
7
9
11
13
15
17
19
21
23
25
27
29
Forecast: adj.R2
Forecast: 1−MAPE
Linear model: adj.R2
Linear model: p−value (country scaled)
Linear model: F−statistic (country scaled)
Linear model: logLik (country scaled)

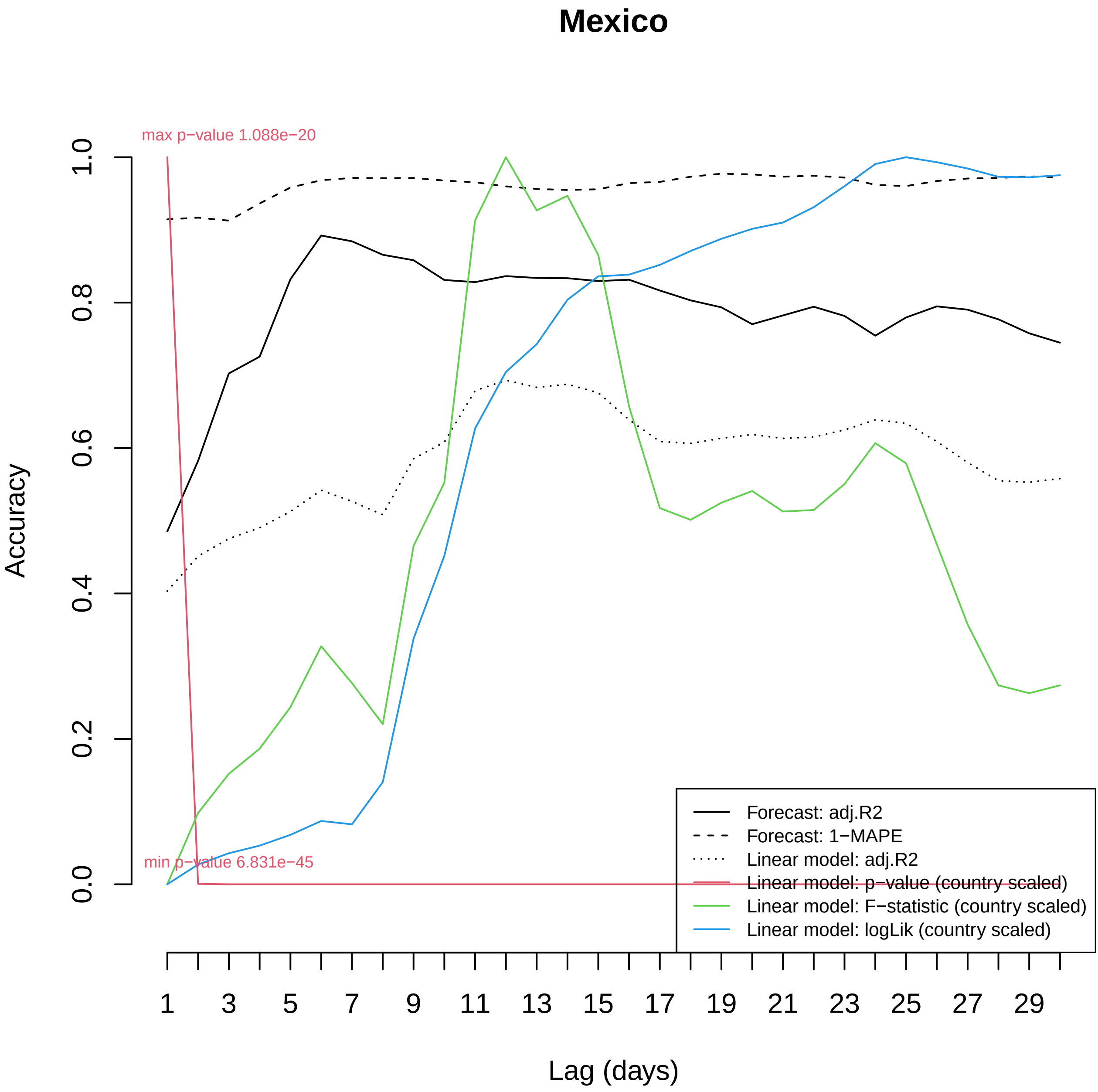

Mexico
max p−value 1.088e−20
min p−value 6.831e−45
Accuracy
0.0
0.2
0.4
0.6
0.8
1.0
1
3
5
7
9
11
13
15
17
19
21
23
25
27
29
Lag (days)
Forecast: adj.R2
Forecast: 1−MAPE
Linear model: adj.R2
Linear model: p−value (country scaled)
Linear model: F−statistic (country scaled)
Linear model: logLik (country scaled)

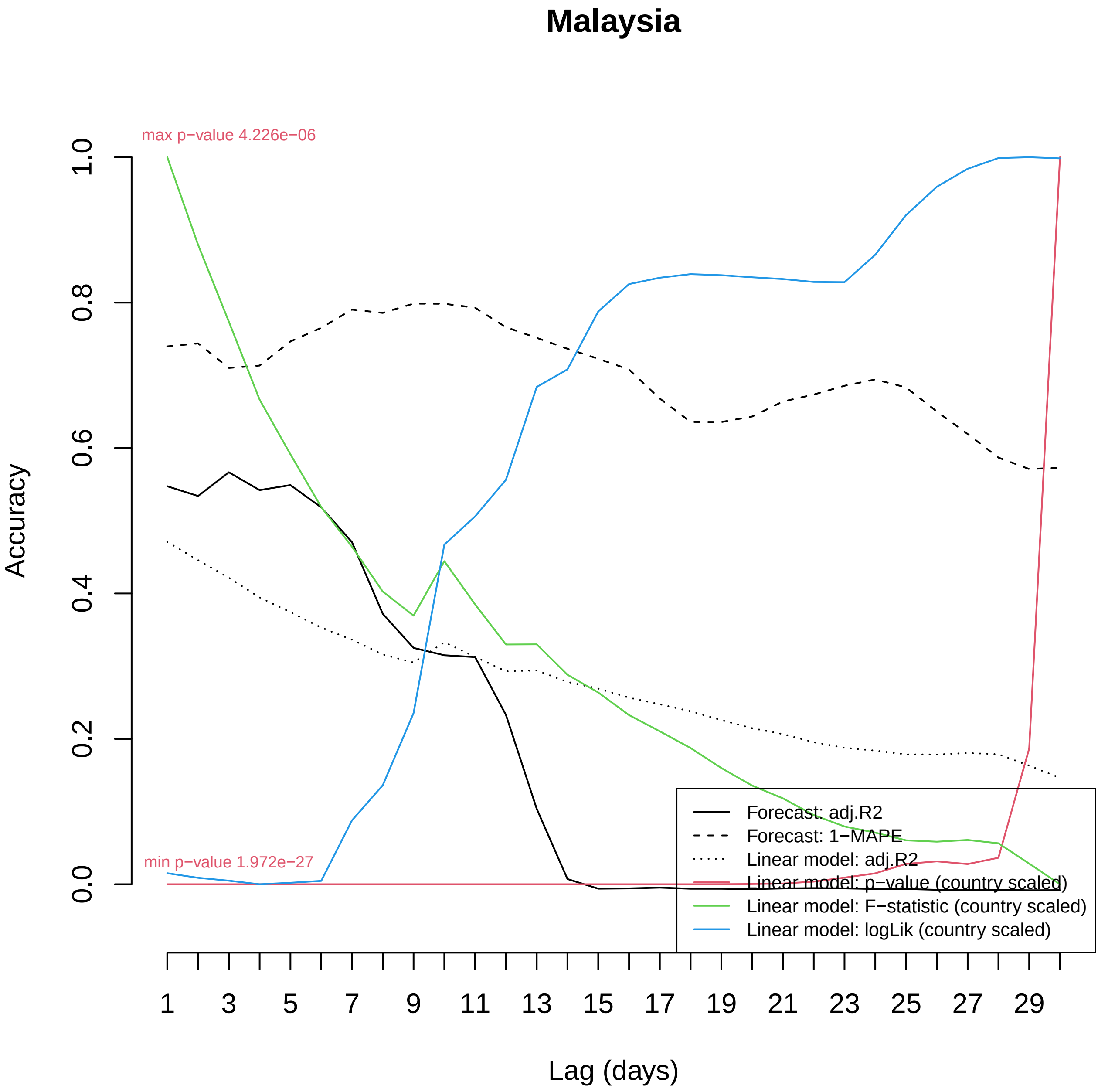

Malaysia
max p−value 4.226e−06
min p−value 1.972e−27
Accuracy
0.0
0.2
0.4
0.6
0.8
1.0
1
3
5
7
9
11
13
15
17
19
21
23
25
27
29
Lag (days)
Forecast: adj.R2
Forecast: 1−MAPE
Linear model: adj.R2
Linear model: p−value (country scaled)
Linear model: F−statistic (country scaled)
Linear model: logLik (country scaled)

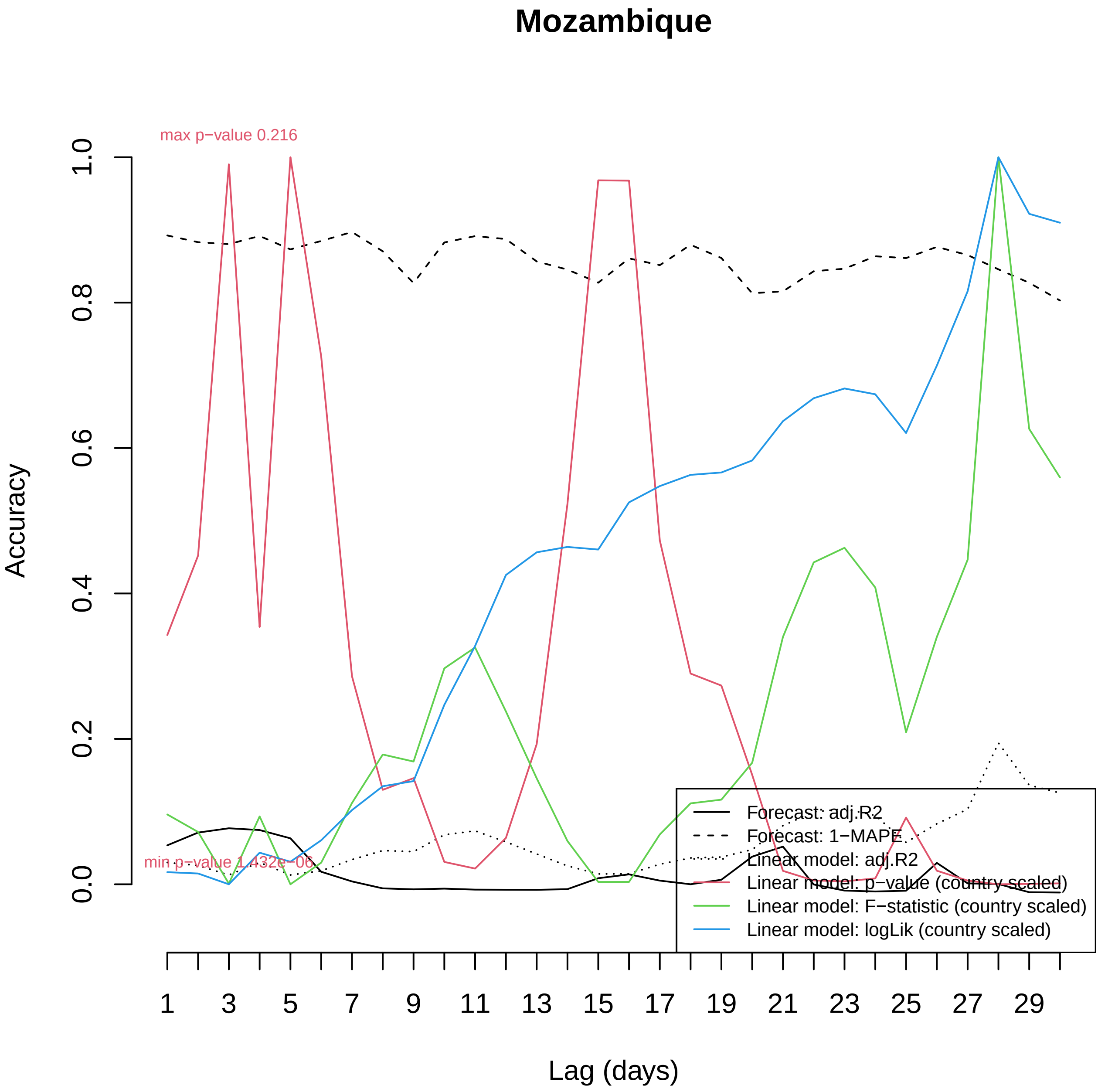
Mozambique
max p−value 0.216
min p−value 1.432e−06
Accuracy
0.0
0.2
0.4
0.6
0.8
1.0
Lag (days)
1
3
5
7
9
11
13
15
17
19
21
23
25
27
29
Forecast: adj.R2
Forecast: 1−MAPE
Linear model: adj.R2
Linear model: p−value (country scaled)
Linear model: F−statistic (country scaled)
Linear model: logLik (country scaled)

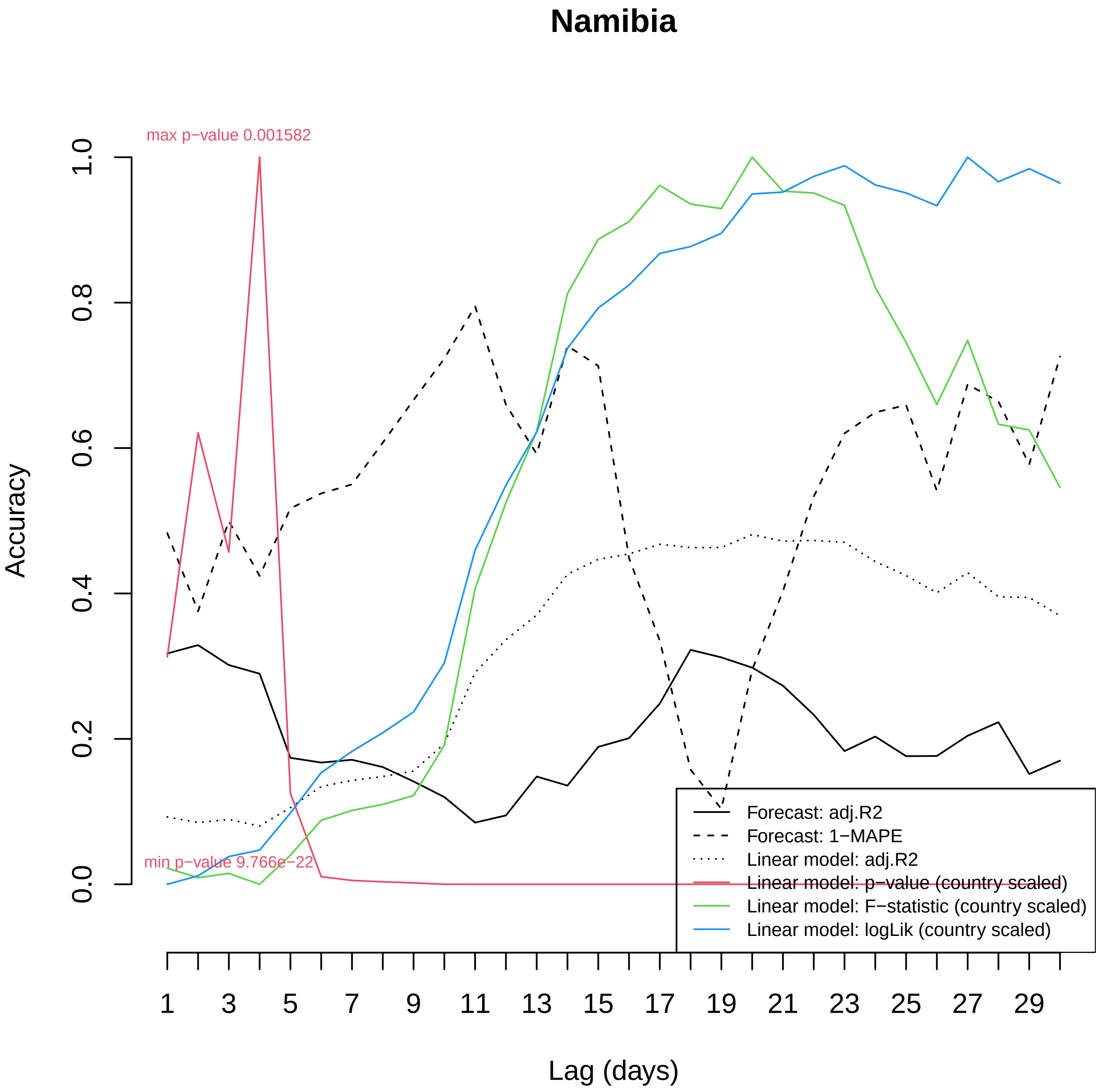

Namibia
max p−value 0.001582
min p−value 9.766e−22
Accuracy
1.0
0.8
0.6
0.4
0.2
0.0
1
3
5
7
9
11
13
15
17
19
21
23
25
27
29
Lag (days)
Forecast: adj.R2
Forecast: 1−MAPE
Linear model: adj.R2
Linear model: p−value (country scaled)
Linear model: F−statistic (country scaled)
Linear model: logLik (country scaled)

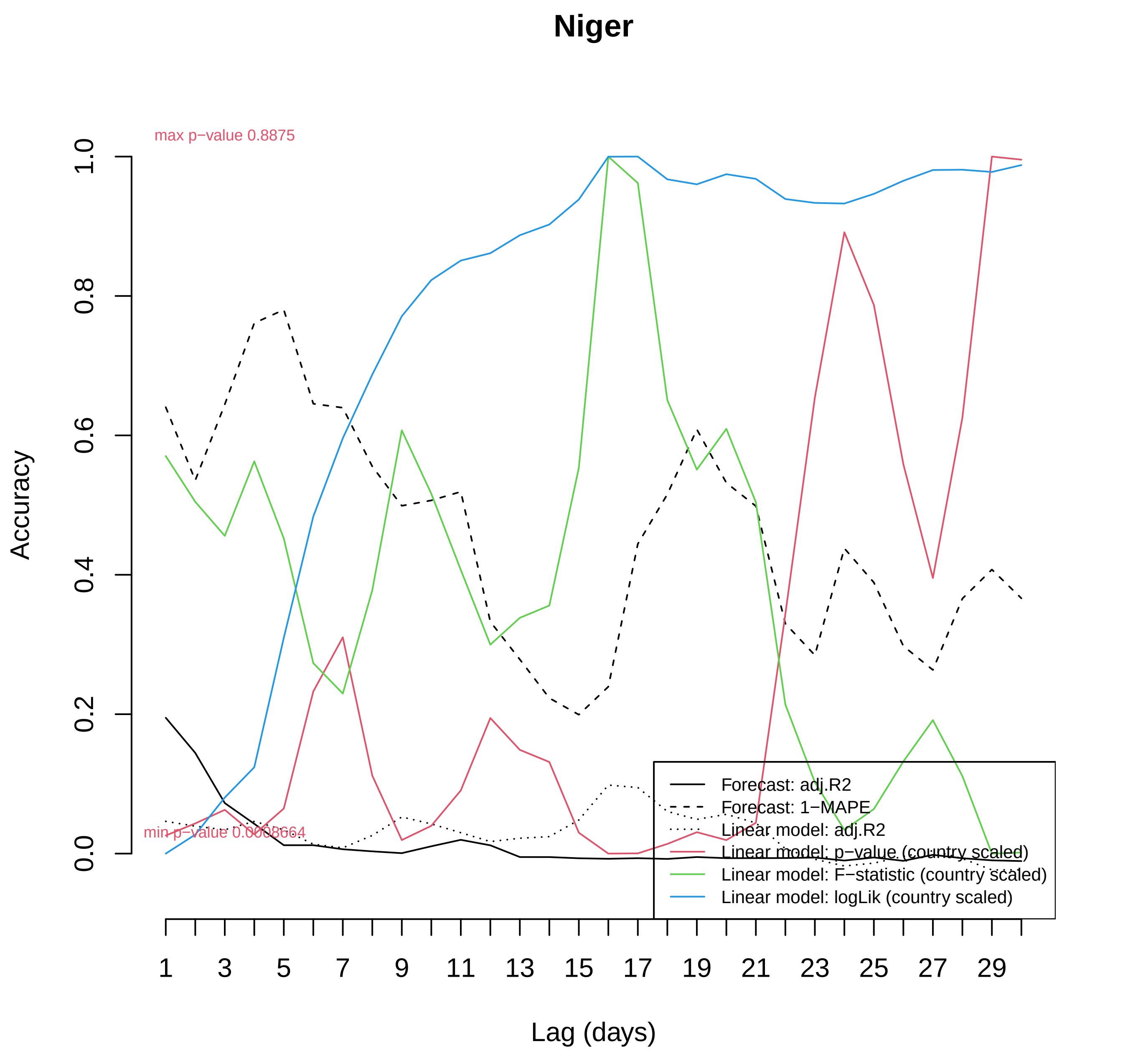

Niger
max p−value 0.8875
min p−value 0.0008664
Accuracy
0.0
0.2
0.4
0.6
0.8
1.0
Lag (days)
1
3
5
7
9
11
13
15
17
19
21
23
25
27
29
Forecast: adj.R2
Forecast: 1−MAPE
Linear model: adj.R2
Linear model: p−value (country scaled)
Linear model: F−statistic (country scaled)
Linear model: logLik (country scaled)

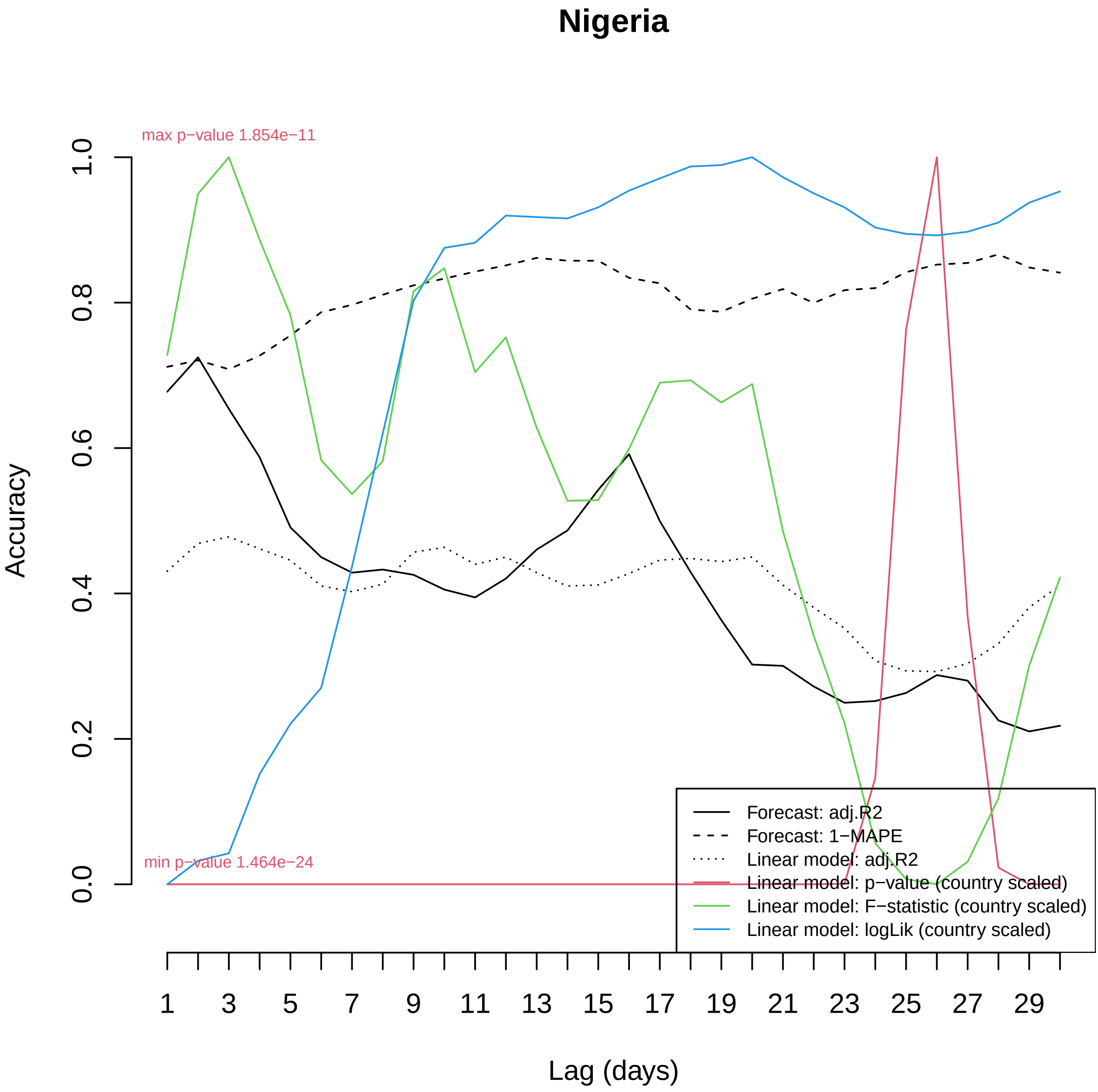
Nigeria
max p−value 1.854e−11
min p−value 1.464e−24
Accuracy
0.0
0.2
0.4
0.6
0.8
1.0
Lag (days)
1
3
5
7
9
11
13
15
17
19
21
23
25
27
29
Forecast: adj.R2
Forecast: 1−MAPE
Linear model: adj.R2
Linear model: p−value (country scaled)
Linear model: F−statistic (country scaled)
Linear model: logLik (country scaled)

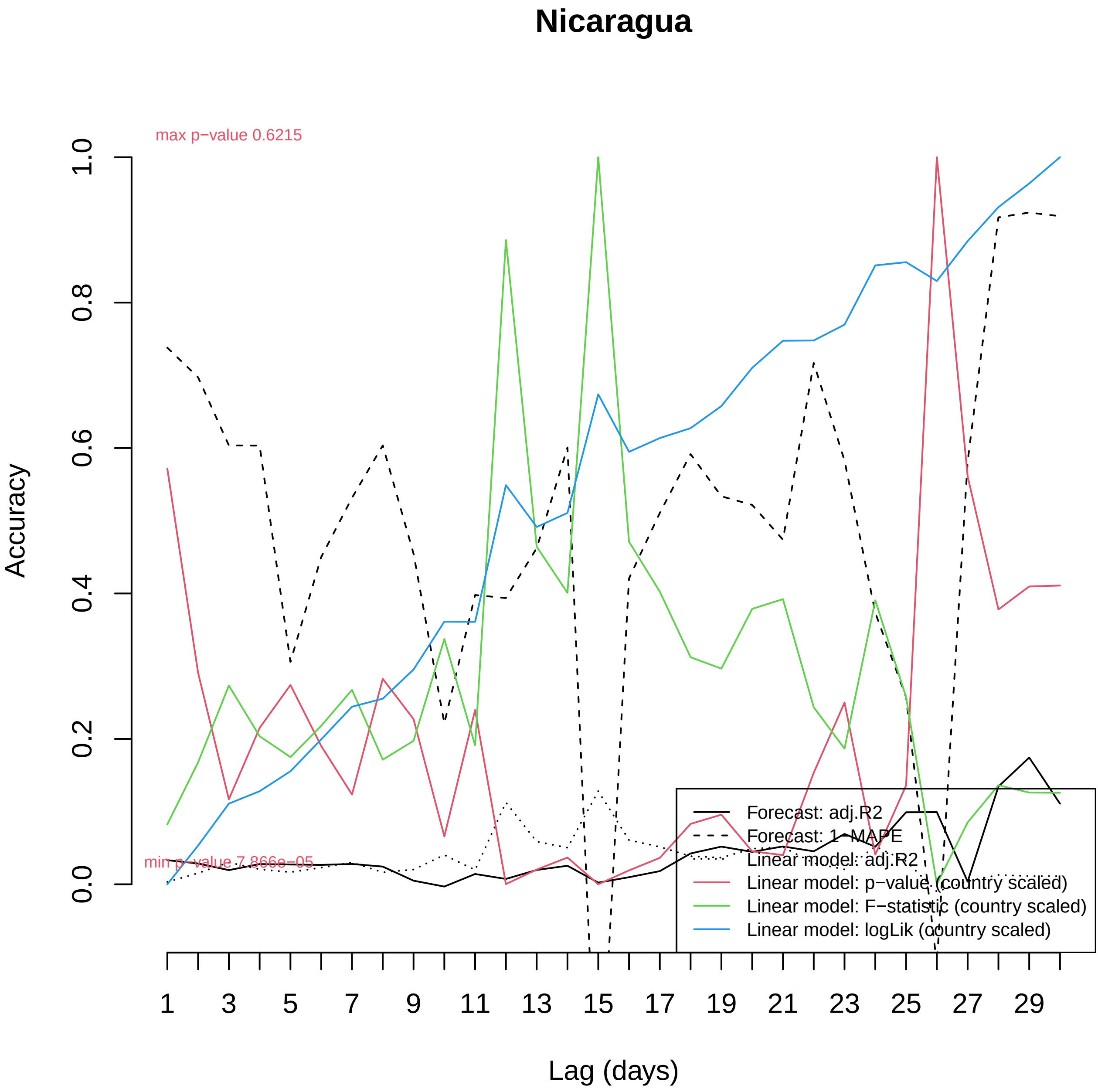

Nicaragua
max p−value 0.6215
min p−value 7.866e−05
Accuracy
0.0
0.2
0.4
0.6
0.8
1.0
1
3
5
7
9
11
13
15
17
19
21
23
25
27
29
Lag (days)
Forecast: adj.R2
Forecast: 1−MAPE
Linear model: adj.R2
Linear model: p−value (country scaled)
Linear model: F−statistic (country scaled)
Linear model: logLik (country scaled)

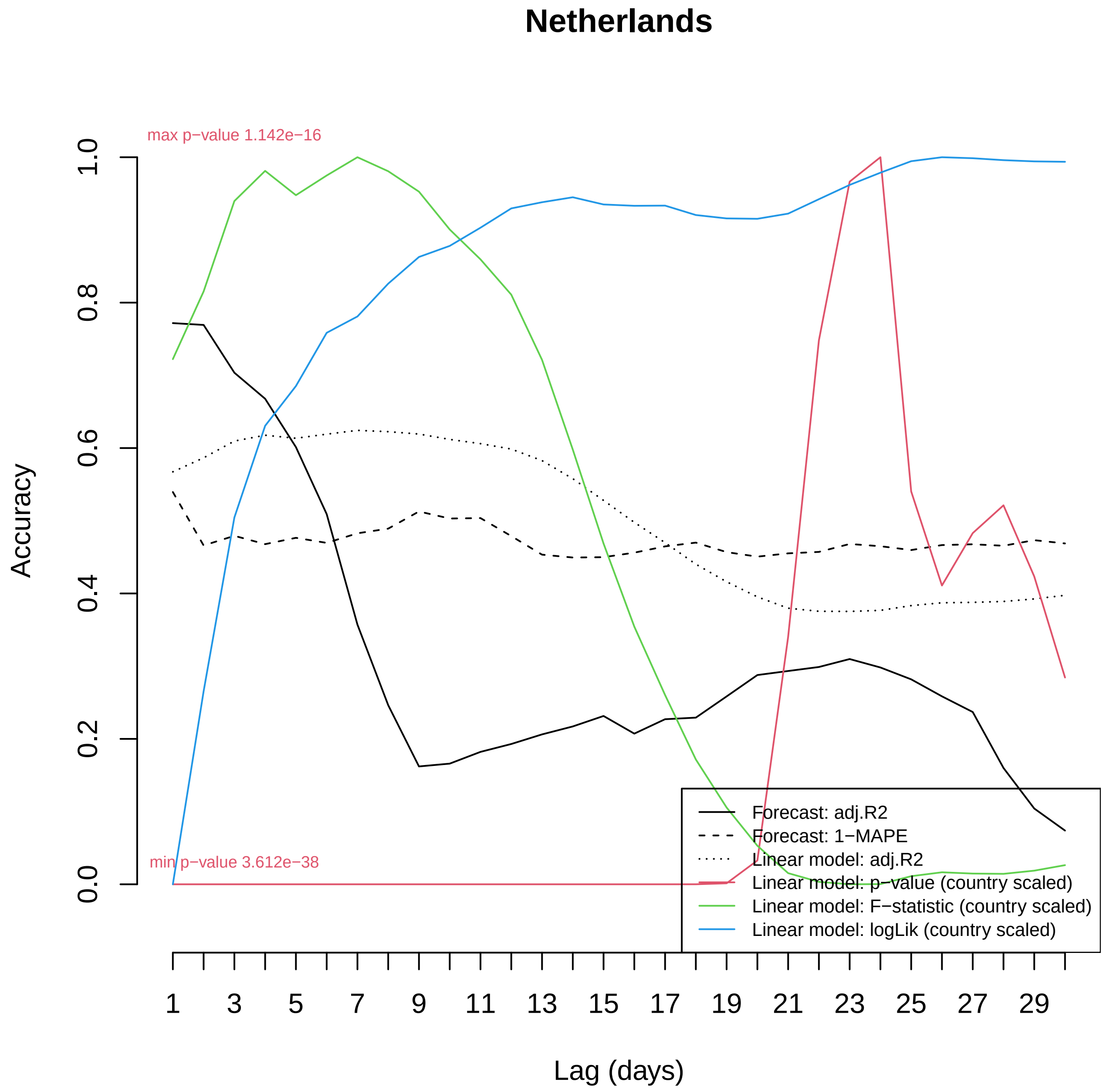

Netherlands
max p−value 1.142e−16
min p−value 3.612e−38
Accuracy
0.0
0.2
0.4
0.6
0.8
1.0
1
3
5
7
9
11
13
15
17
19
21
23
25
27
29
Lag (days)
Forecast: adj.R2
Forecast: 1−MAPE
Linear model: adj.R2
Linear model: p−value (country scaled)
Linear model: F−statistic (country scaled)
Linear model: logLik (country scaled)

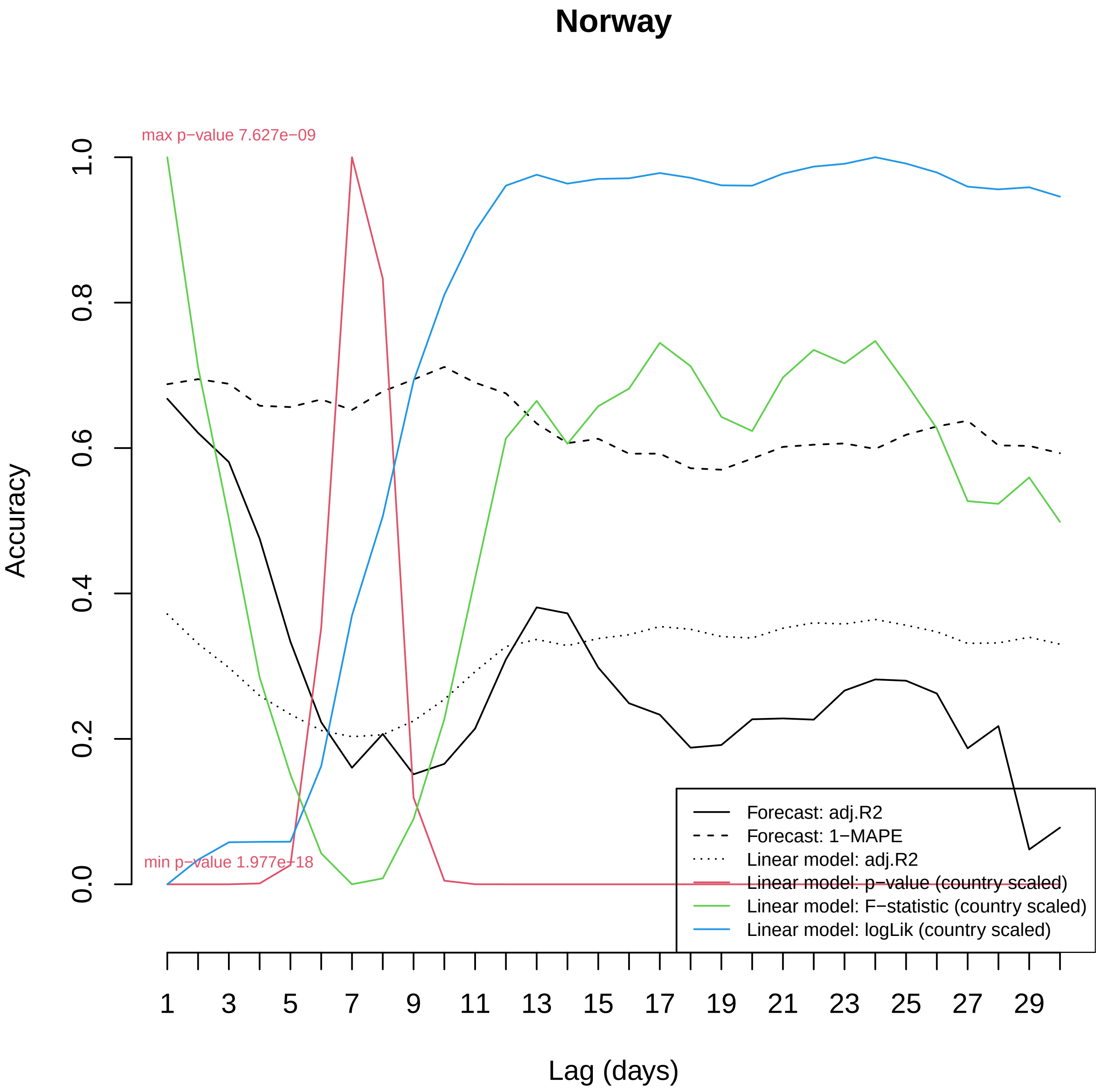
Norway
max p−value 7.627e−09
min p−value 1.977e−18
Accuracy
0.0
0.2
0.4
0.6
0.8
1.0
1
3
5
7
9
11
13
15
17
19
21
23
25
27
29
Lag (days)
Forecast: adj.R2
Forecast: 1−MAPE
Linear model: adj.R2
Linear model: p−value (country scaled)
Linear model: F−statistic (country scaled)
Linear model: logLik (country scaled)

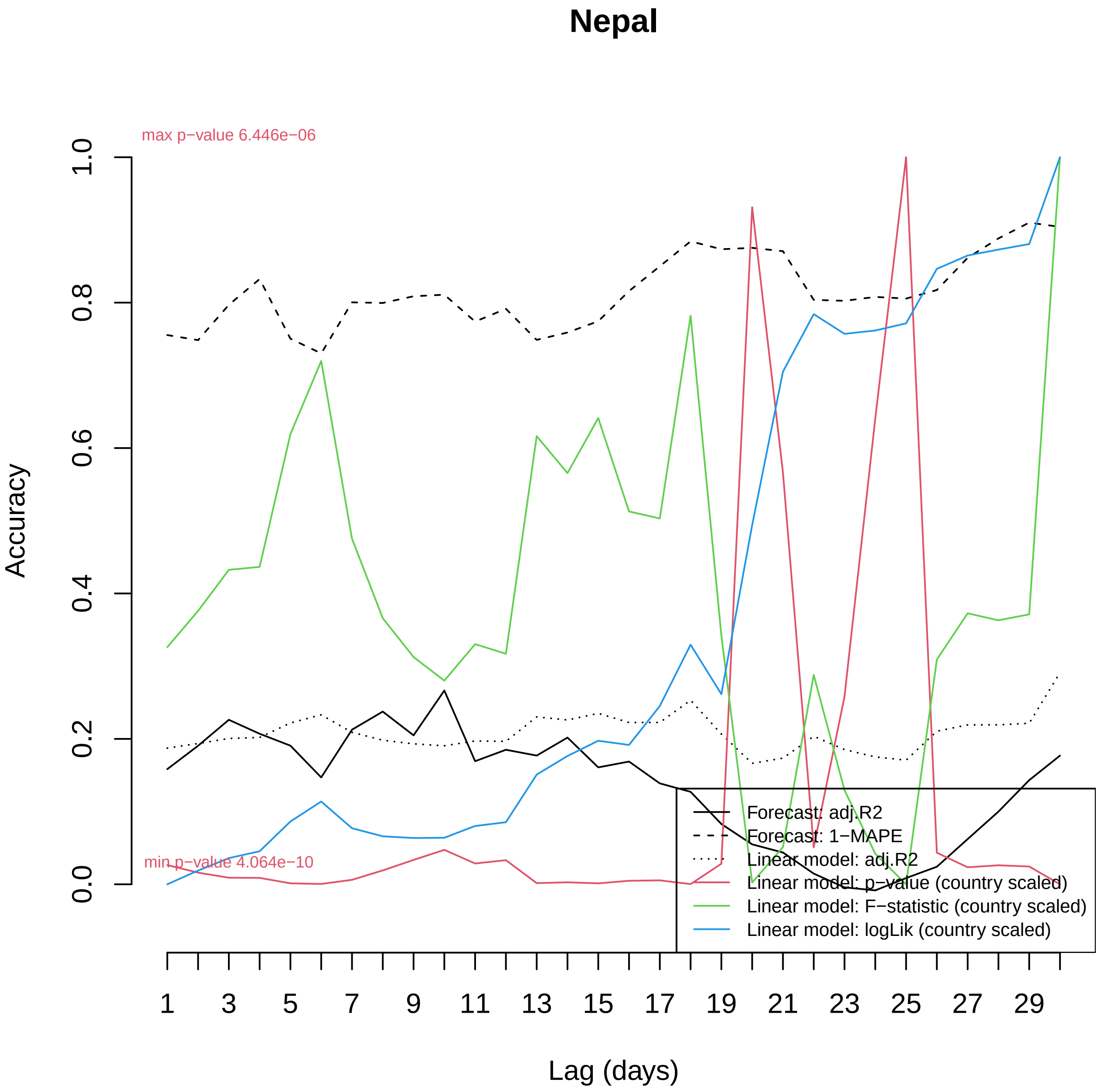

Nepal
max p−value 6.446e−06
min p−value 4.064e−10
Accuracy
0.0
0.2
0.4
0.6
0.8
1.0
Lag (days)
1
3
5
7
9
11
13
15
17
19
21
23
25
27
29
Forecast: adj.R2
Forecast: 1−MAPE
Linear model: adj.R2
Linear model: p−value (country scaled)
Linear model: F−statistic (country scaled)
Linear model: logLik (country scaled)

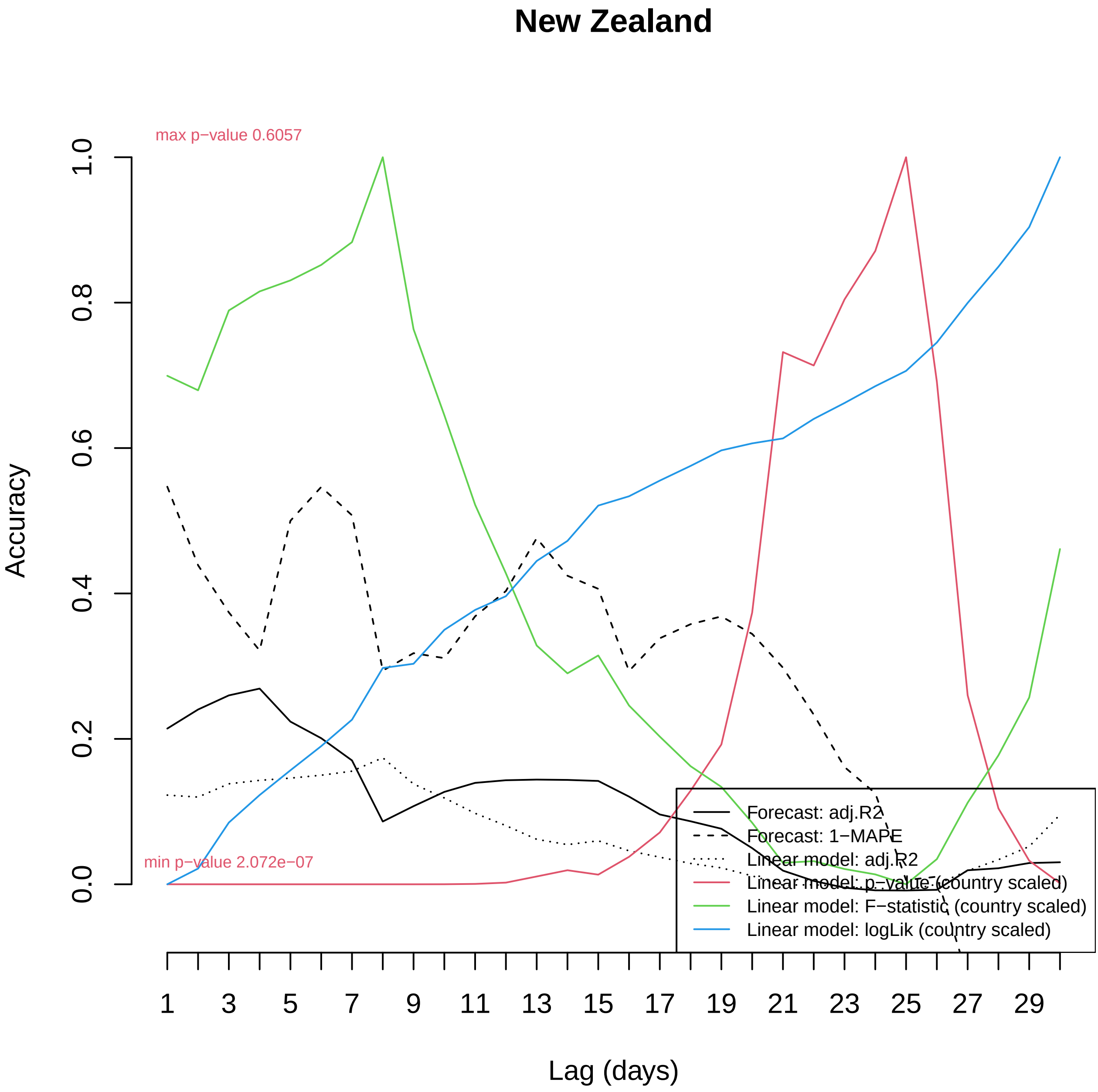

New Zealand
max p−value 0.6057
min p−value 2.072e−07
Accuracy
0.0
0.2
0.4
0.6
0.8
1.0
1
3
5
7
9
11
13
15
17
19
21
23
25
27
29
Lag (days)
Forecast: adj.R2
Forecast: 1−MAPE
Linear model: adj.R2
Linear model: p−value (country scaled)
Linear model: F−statistic (country scaled)
Linear model: logLik (country scaled)

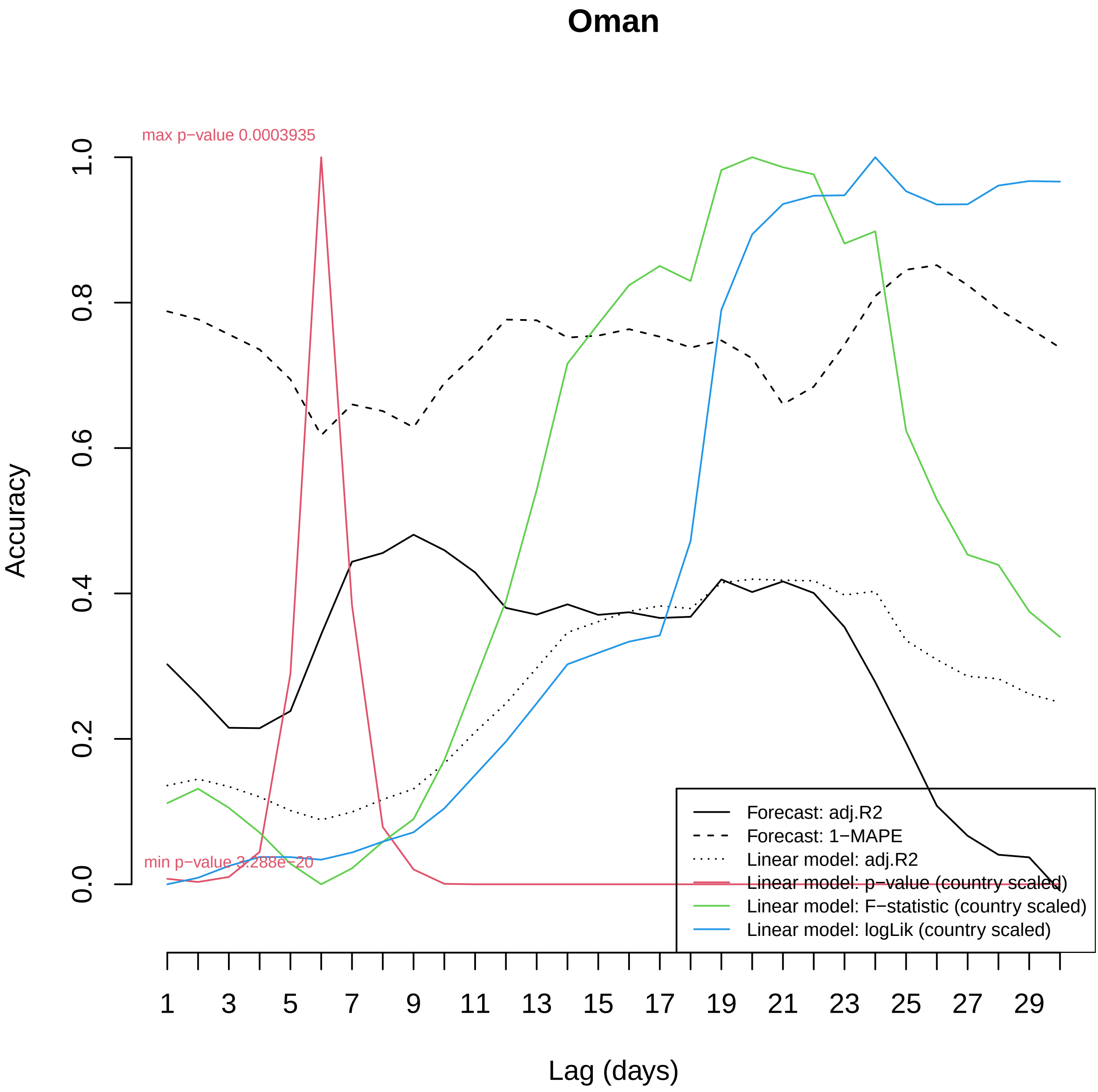

Oman
max p−value 0.0003935
min p−value 3.288e−20
Accuracy
1.0
0.8
0.6
0.4
0.2
0.0
1
3
5
7
9
11
13
15
17
19
21
23
25
27
29
Lag (days)
Forecast: adj.R2
Forecast: 1−MAPE
Linear model: adj.R2
Linear model: p−value (country scaled)
Linear model: F−statistic (country scaled)
Linear model: logLik (country scaled)

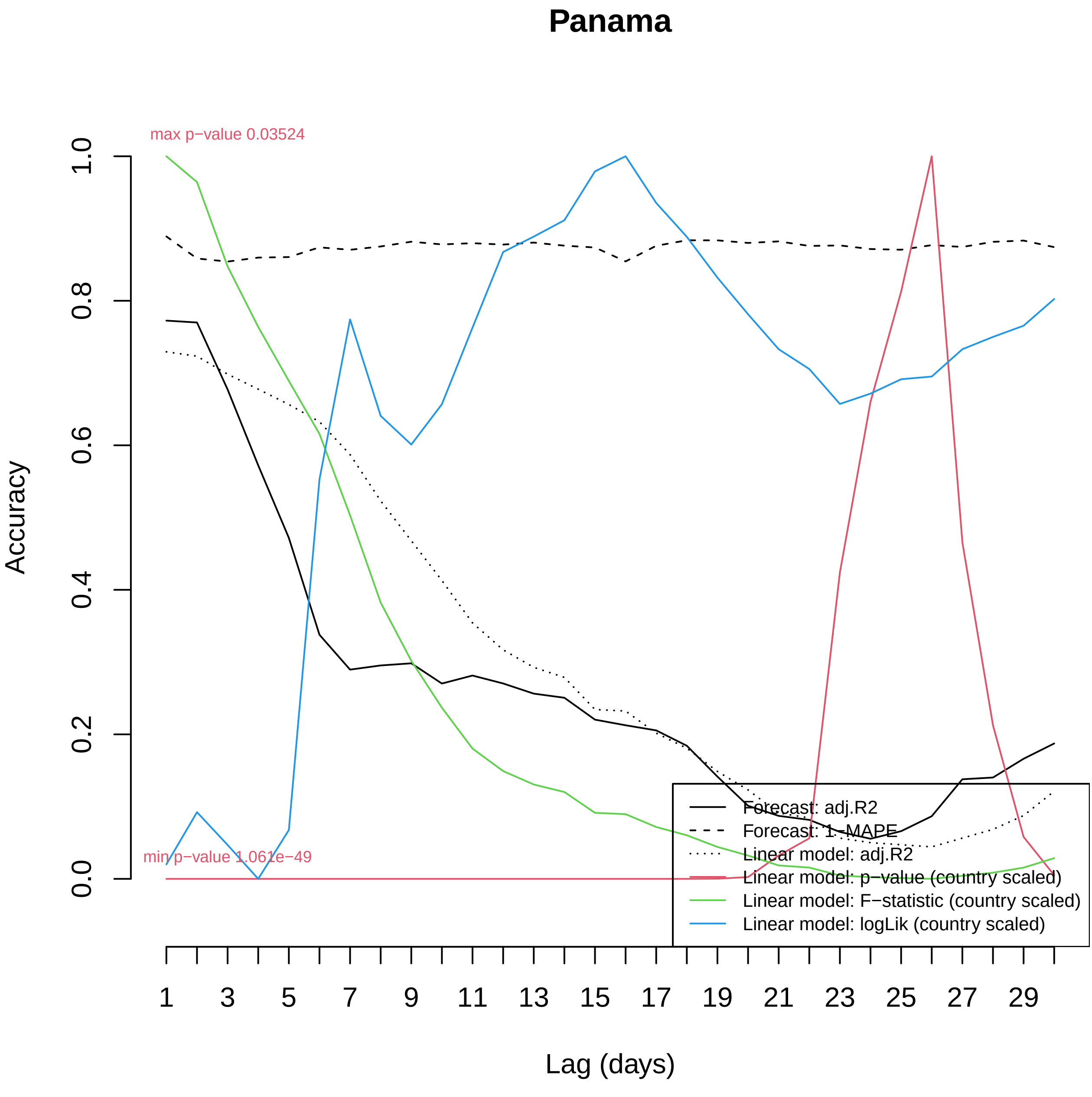

Panama
max p−value 0.03524
min p−value 1.061e−49
Accuracy
0.0
0.2
0.4
0.6
0.8
1.0
Lag (days)
1
3
5
7
9
11
13
15
17
19
21
23
25
27
29
Forecast: adj.R2
Forecast: 1−MAPE
Linear model: adj.R2
Linear model: p−value (country scaled)
Linear model: F−statistic (country scaled)
Linear model: logLik (country scaled)

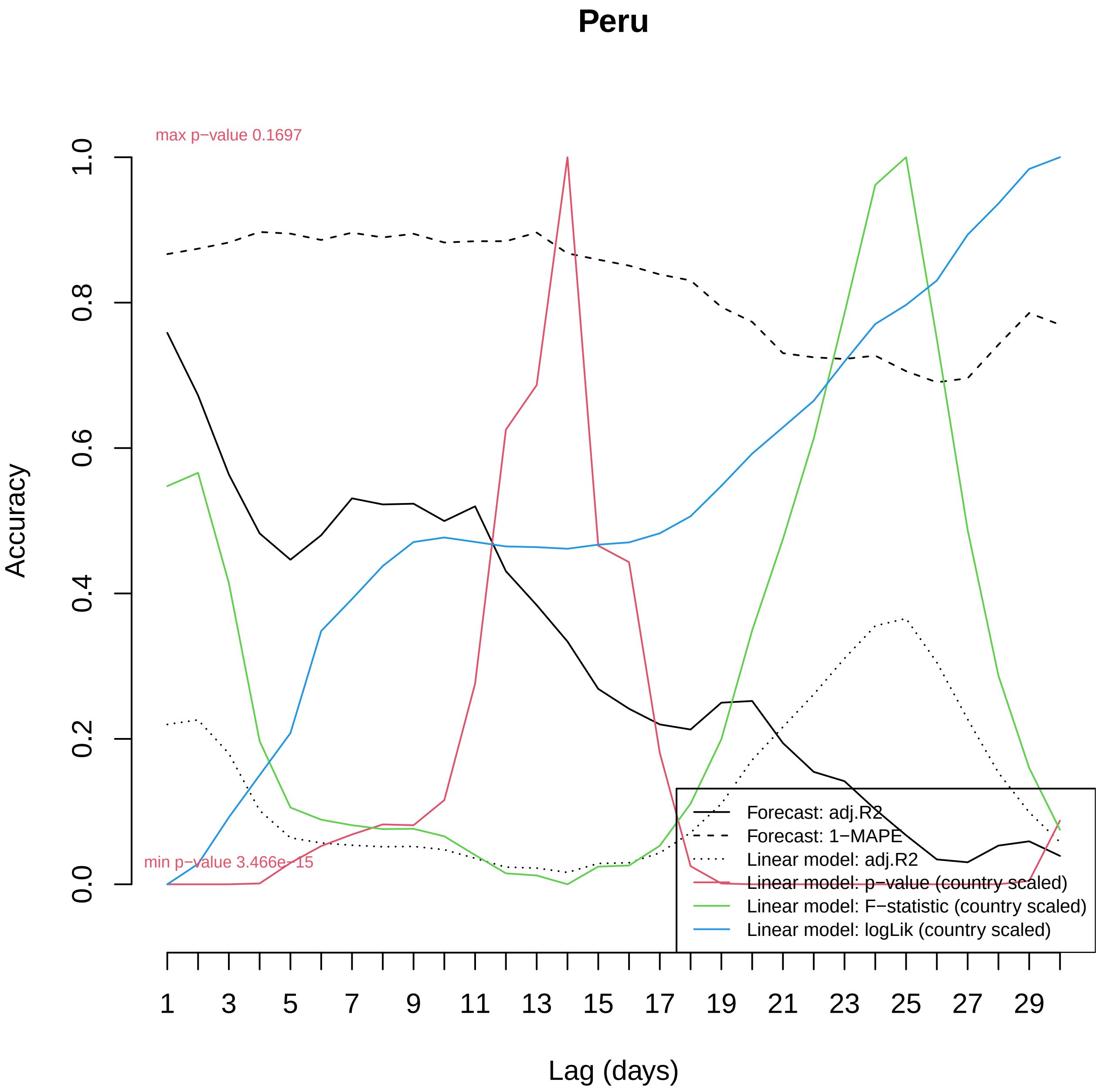

Peru
max p−value 0.1697
min p−value 3.466e−15
Accuracy
0.0
0.2
0.4
0.6
0.8
1.0
1
3
5
7
9
11
13
15
17
19
21
23
25
27
29
Lag (days)
Forecast: adj.R2
Forecast: 1−MAPE
Linear model: adj.R2
Linear model: p−value (country scaled)
Linear model: F−statistic (country scaled)
Linear model: logLik (country scaled)

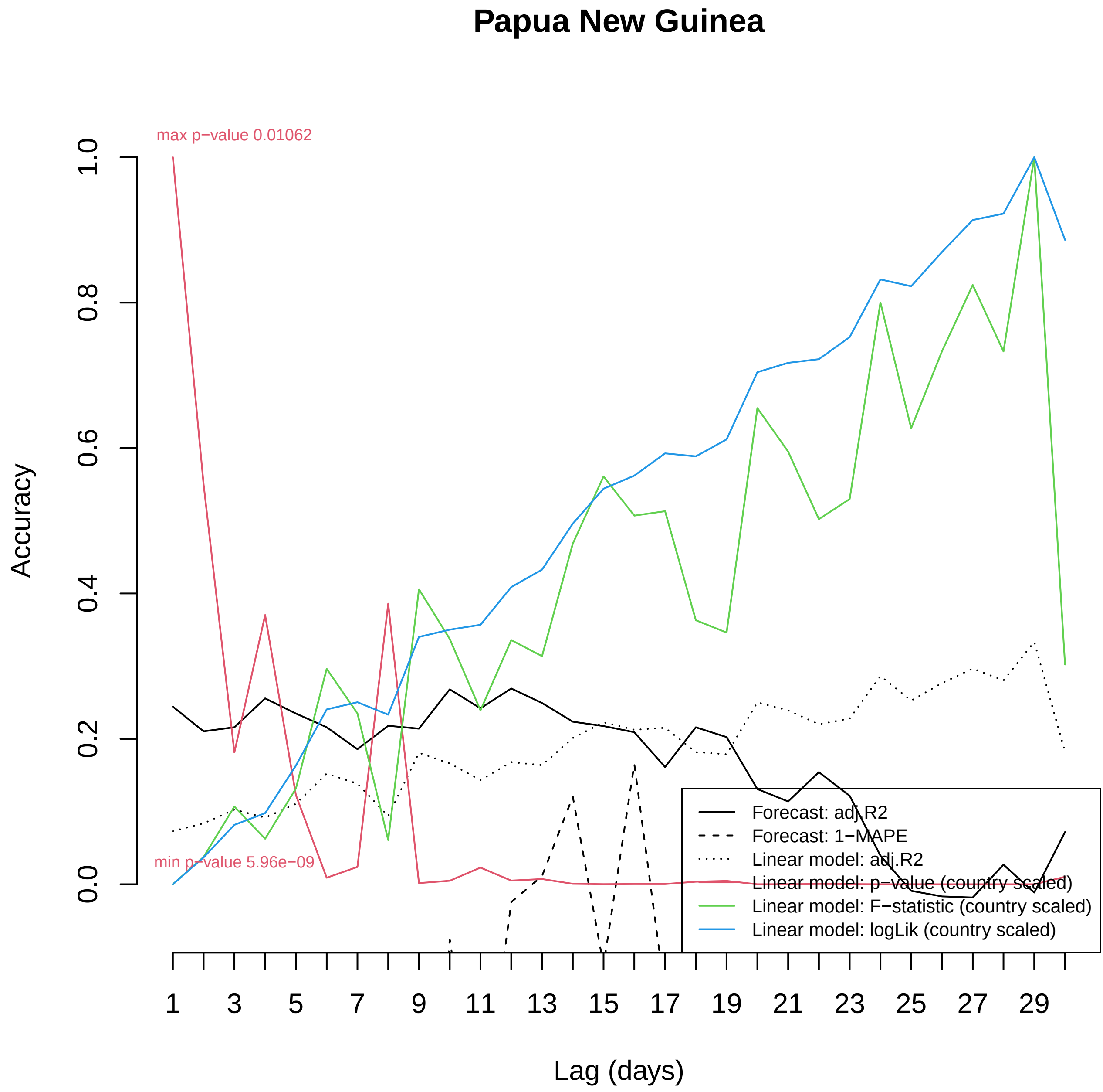

Papua New Guinea
max p−value 0.01062
min p−value 5.96e−09
Accuracy
0.0
0.2
0.4
0.6
0.8
1.0
1
3
5
7
9
11
13
15
17
19
21
23
25
27
29
Lag (days)
Forecast: adj.R2
Forecast: 1−MAPE
Linear model: adj.R2
Linear model: p−value (country scaled)
Linear model: F−statistic (country scaled)
Linear model: logLik (country scaled)

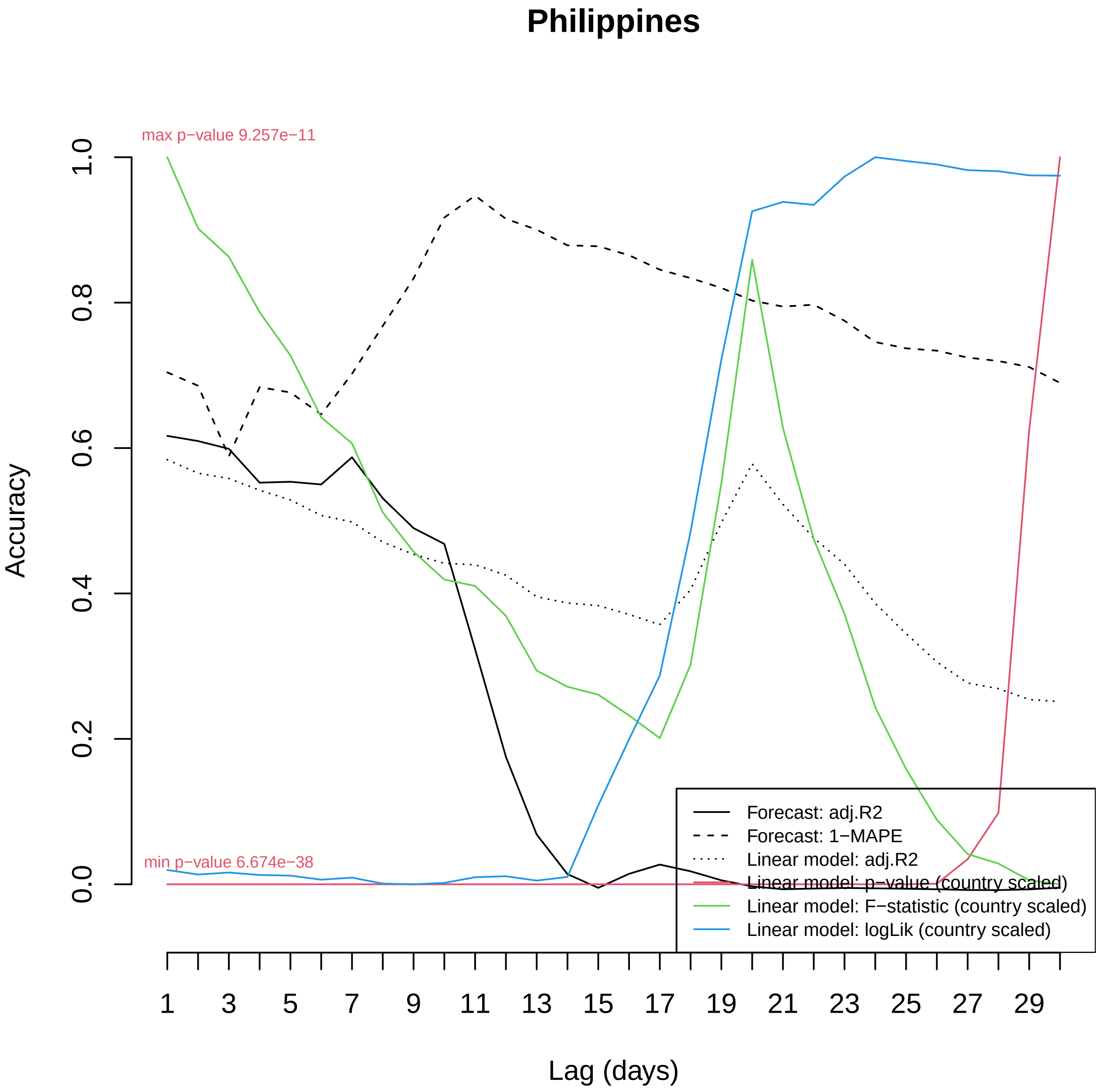

Philippines
max p−value 9.257e−11
min p−value 6.674e−38
Accuracy
0.0
0.2
0.4
0.6
0.8
1.0
Lag (days)
1
3
5
7
9
11
13
15
17
19
21
23
25
27
29
Forecast: adj.R2
Forecast: 1−MAPE
Linear model: adj.R2
Linear model: p−value (country scaled)
Linear model: F−statistic (country scaled)
Linear model: logLik (country scaled)

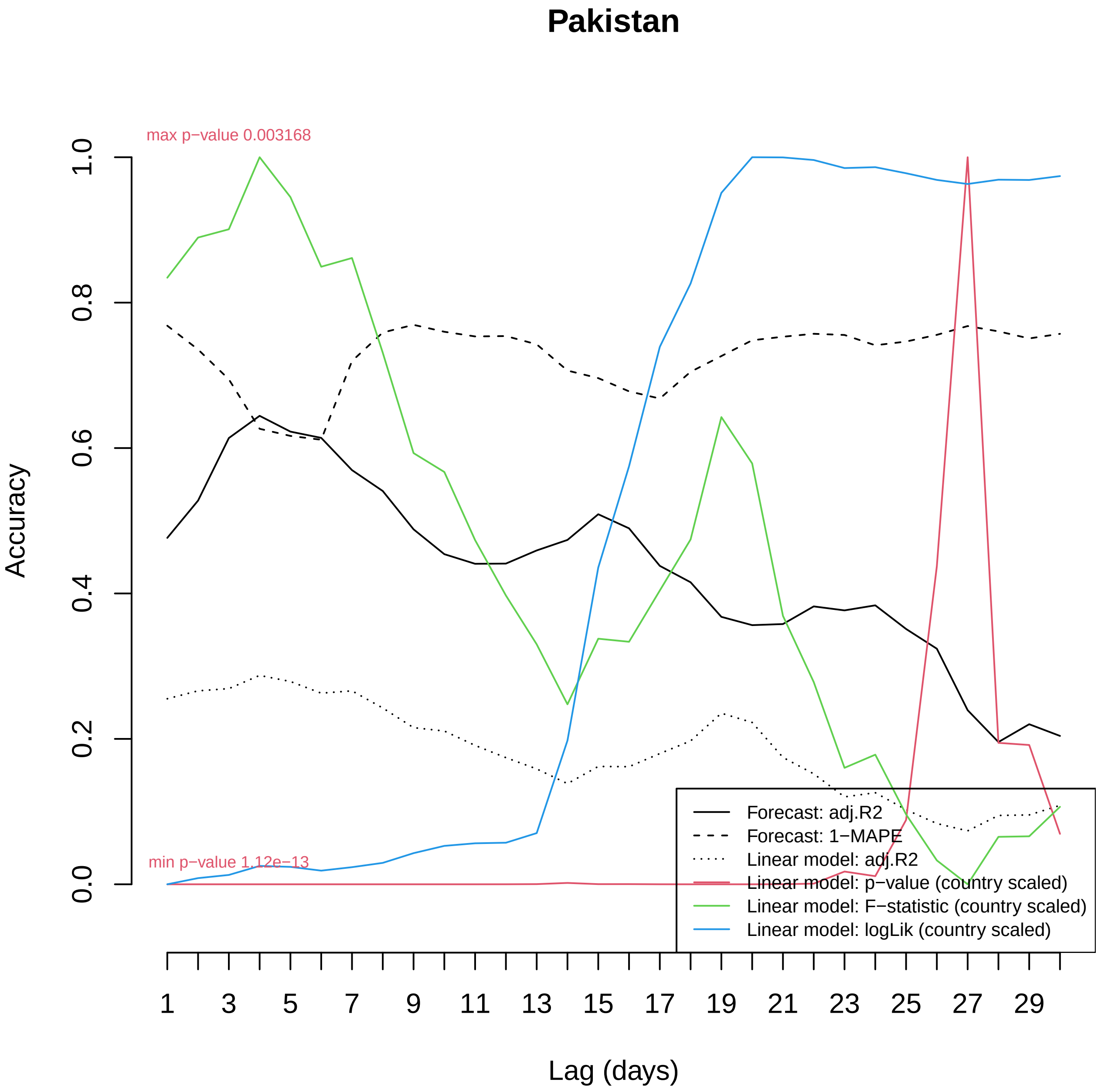

Pakistan
max p−value 0.003168
min p−value 1.12e−13
Accuracy
0.0
0.2
0.4
0.6
0.8
1.0
Lag (days)
1
3
5
7
9
11
13
15
17
19
21
23
25
27
29
Forecast: adj.R2
Forecast: 1−MAPE
Linear model: adj.R2
Linear model: p−value (country scaled)
Linear model: F−statistic (country scaled)
Linear model: logLik (country scaled)

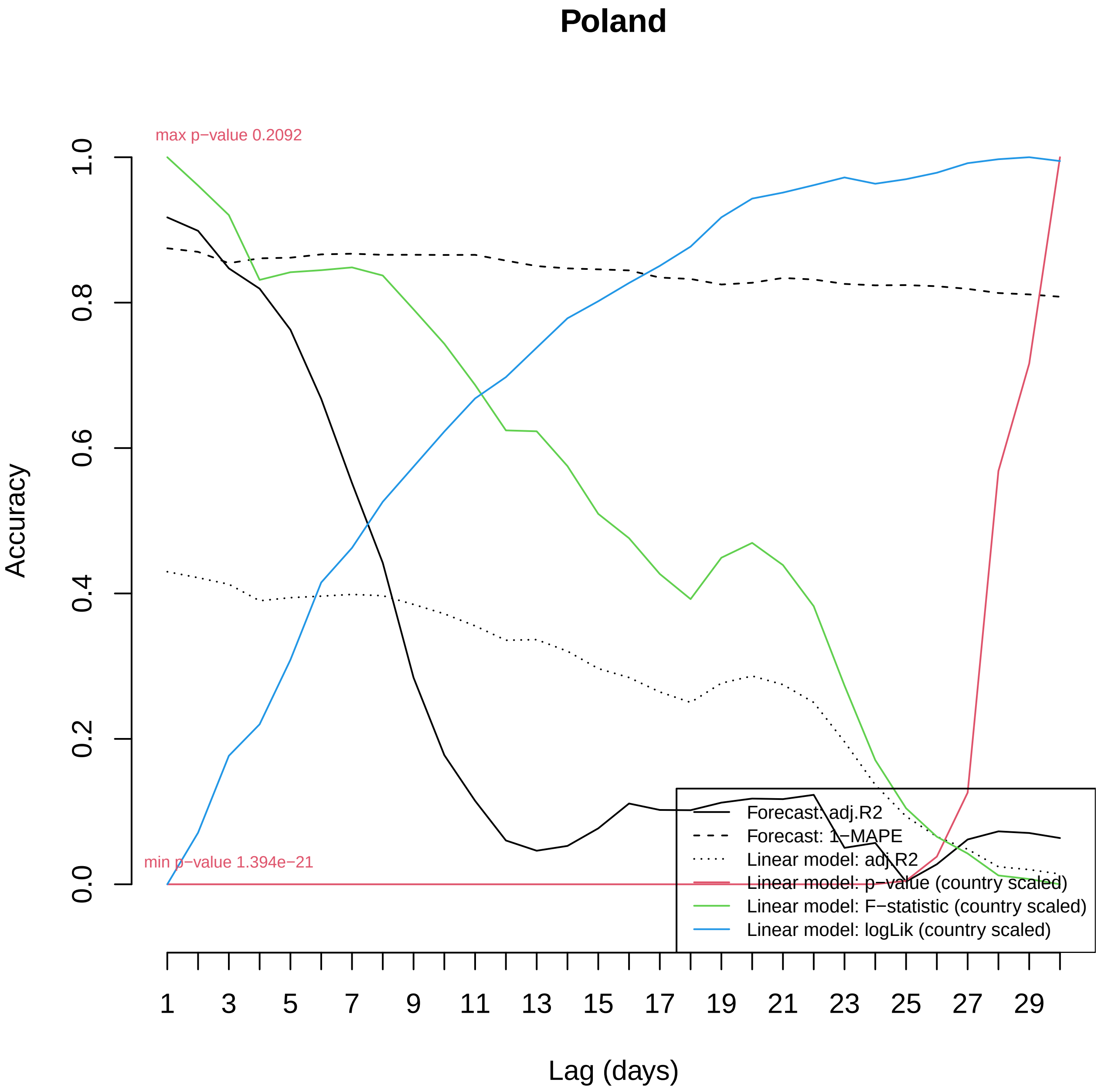

Poland
max p−value 0.2092
min p−value 1.394e−21
Accuracy
1.0
0.8
0.6
0.4
0.2
0.0
1
3
5
7
9
11
13
15
17
19
21
23
25
27
29
Lag (days)
Forecast: adj.R2
Forecast: 1−MAPE
Linear model: adj.R2
Linear model: p−value (country scaled)
Linear model: F−statistic (country scaled)
Linear model: logLik (country scaled)

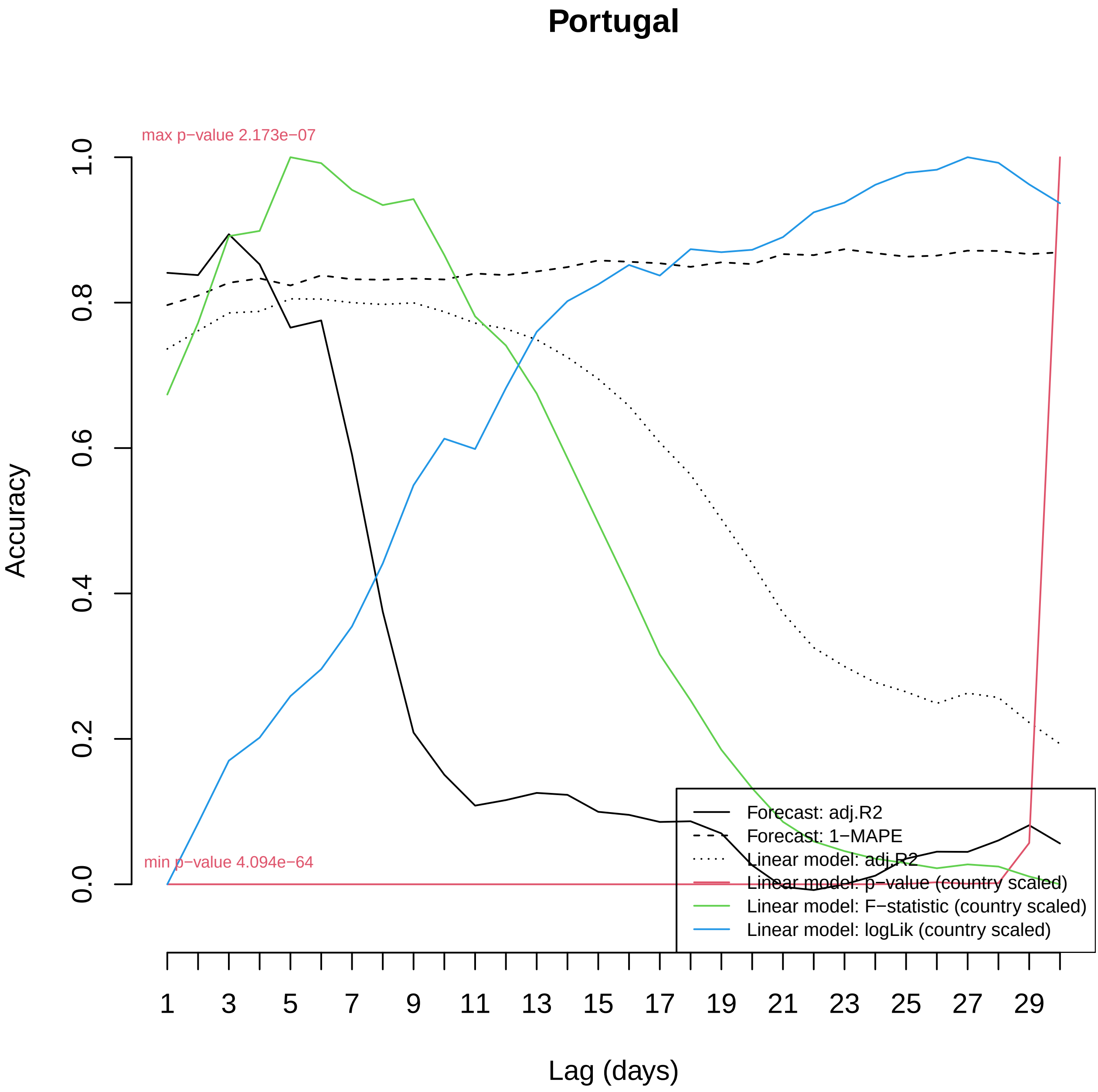
Portugal
max p−value 2.173e−07
min p−value 4.094e−64
Accuracy
0.0
0.2
0.4
0.6
0.8
1.0
Lag (days)
1
3
5
7
9
11
13
15
17
19
21
23
25
27
29
Forecast: adj.R2
Forecast: 1−MAPE
Linear model: adj.R2
Linear model: p−value (country scaled)
Linear model: F−statistic (country scaled)
Linear model: logLik (country scaled)

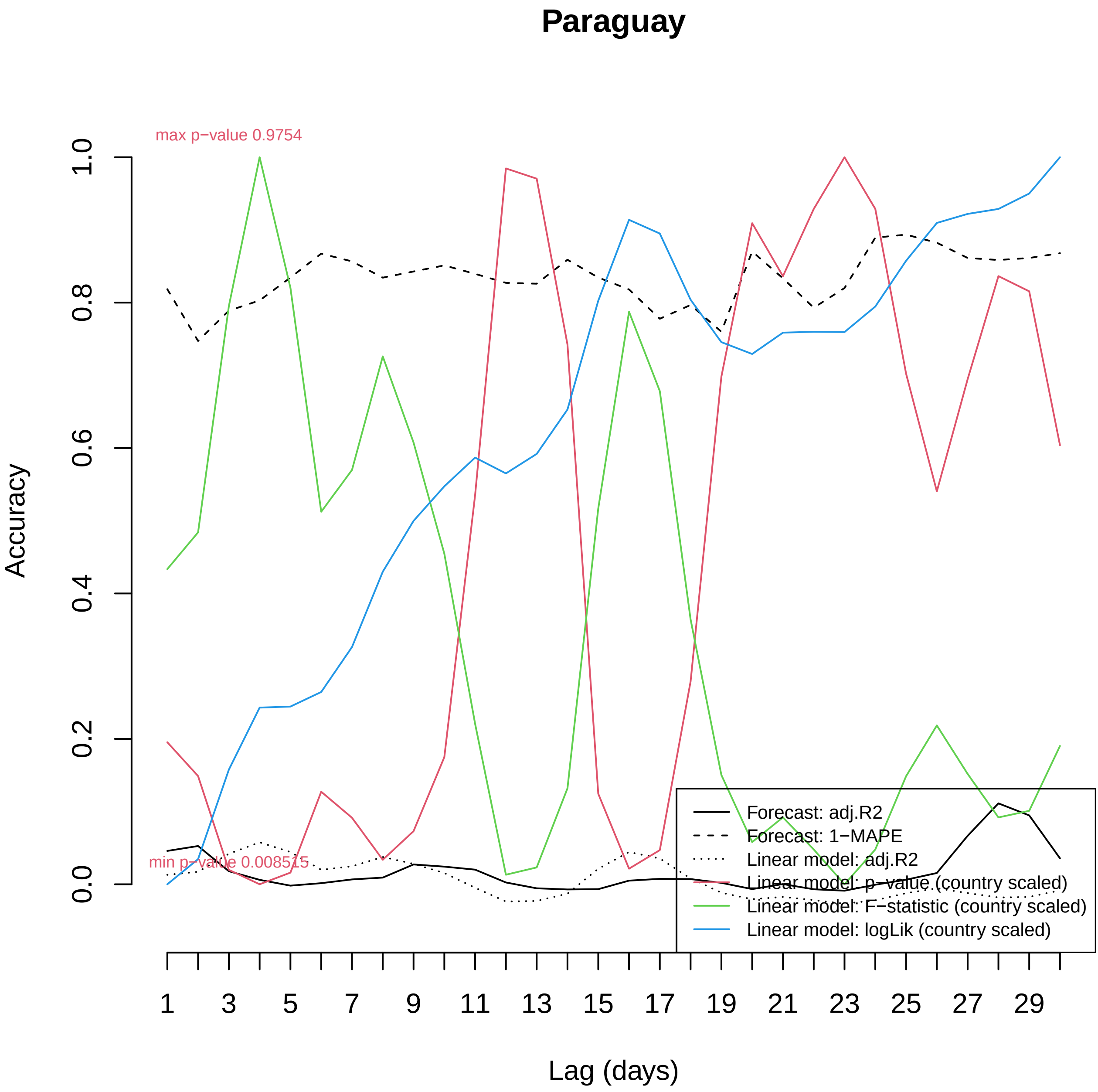

Paraguay
max p−value 0.9754
min p−value 0.008515
Accuracy
0.0
0.2
0.4
0.6
0.8
1.0
Lag (days)
1
3
5
7
9
11
13
15
17
19
21
23
25
27
29
Forecast: adj.R2
Forecast: 1−MAPE
Linear model: adj.R2
Linear model: p−value (country scaled)
Linear model: F−statistic (country scaled)
Linear model: logLik (country scaled)

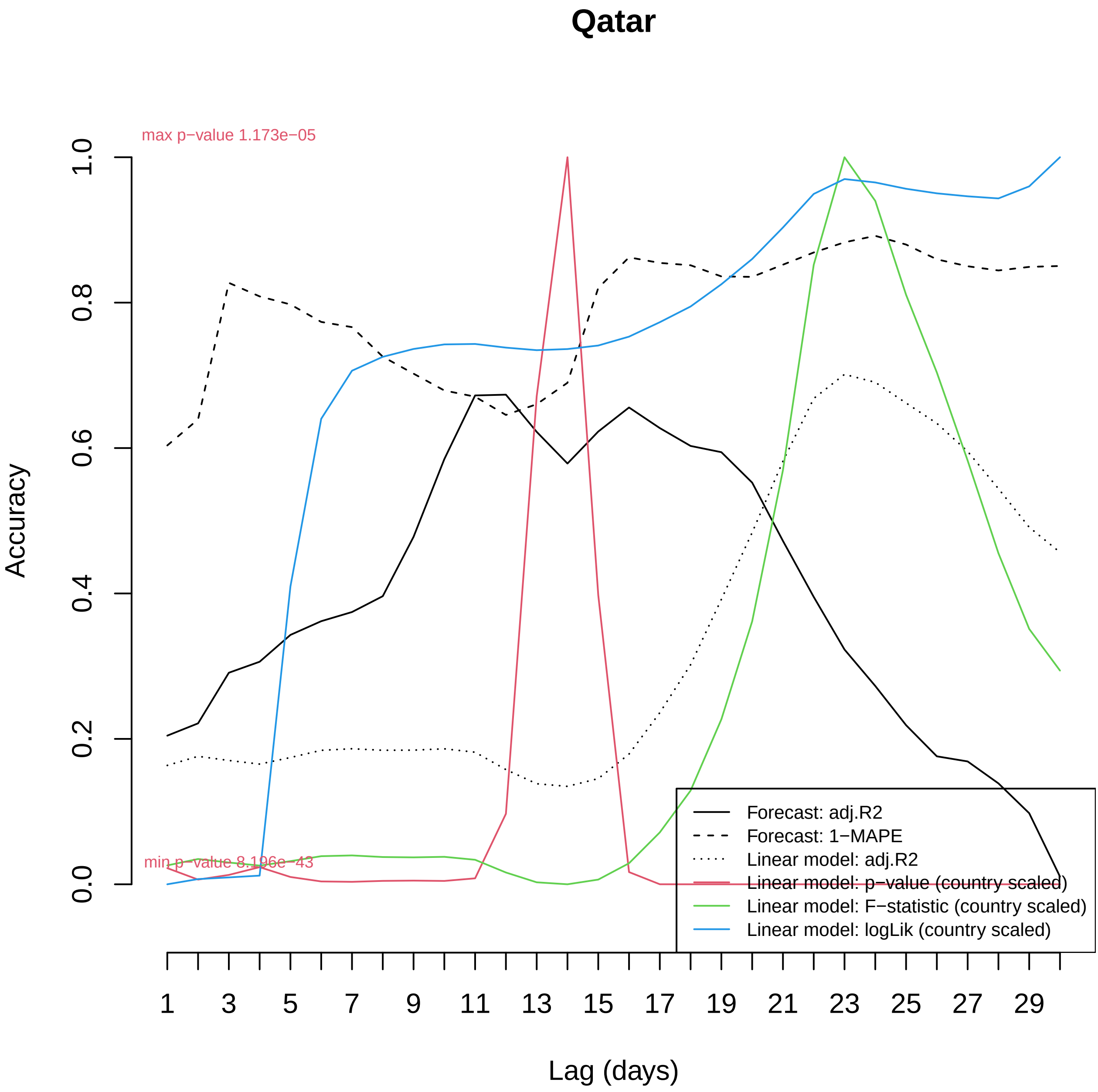

Qatar
max p−value 1.173e−05
min p−value 8.196e−43
Accuracy
0.0
0.2
0.4
0.6
0.8
1.0
Lag (days)
1
3
5
7
9
11
13
15
17
19
21
23
25
27
29
Forecast: adj.R2
Forecast: 1−MAPE
Linear model: adj.R2
Linear model: p−value (country scaled)
Linear model: F−statistic (country scaled)
Linear model: logLik (country scaled)

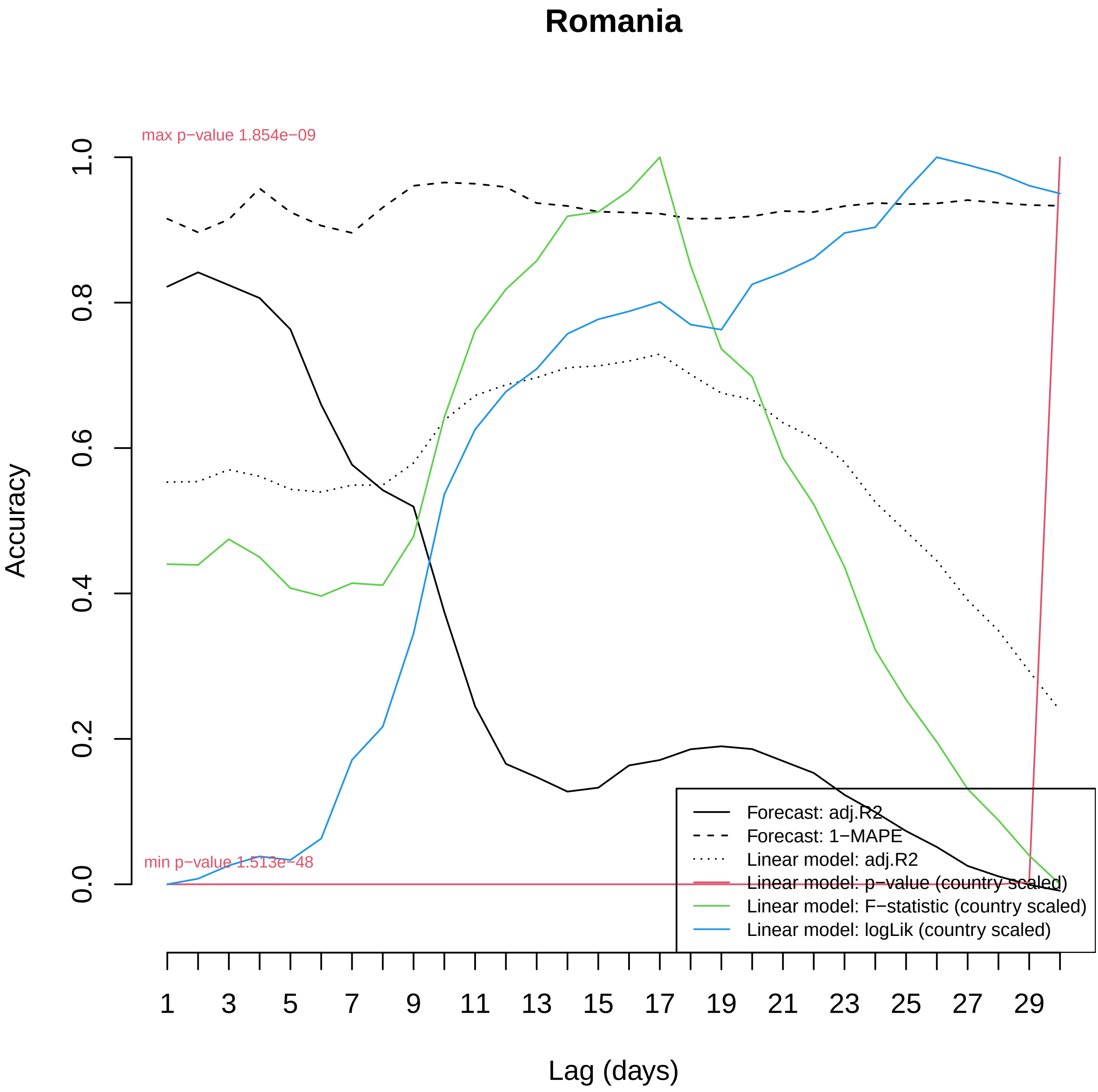

Romania
max p−value 1.854e−09
min p−value 1.513e−48
Accuracy
0.0
0.2
0.4
0.6
0.8
1.0
1
3
5
7
9
11
13
15
17
19
21
23
25
27
29
Lag (days)
Forecast: adj.R2
Forecast: 1−MAPE
Linear model: adj.R2
Linear model: p−value (country scaled)
Linear model: F−statistic (country scaled)
Linear model: logLik (country scaled)

### Serbia

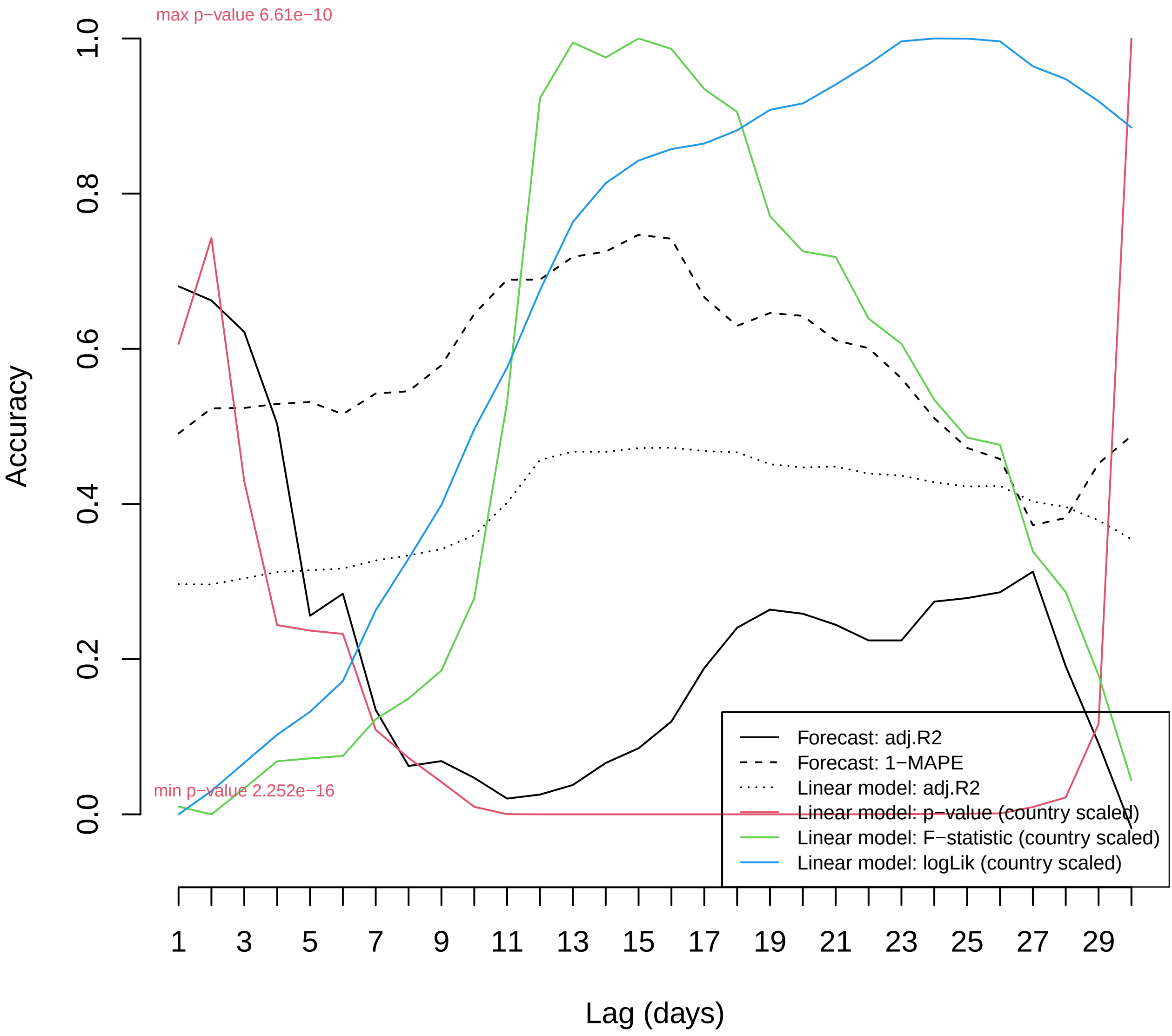

max p−value 6.61e−10
min p−value 2.252e−16
Accuracy
0.0
0.2
0.4
0.6
0.8
1.0
Lag (days)
1
3
5
7
9
11
13
15
17
19
21
23
25
27
29
Forecast: adj.R2
Forecast: 1−MAPE
Linear model: adj.R2
Linear model: p−value (country scaled)
Linear model: F−statistic (country scaled)
Linear model: logLik (country scaled)

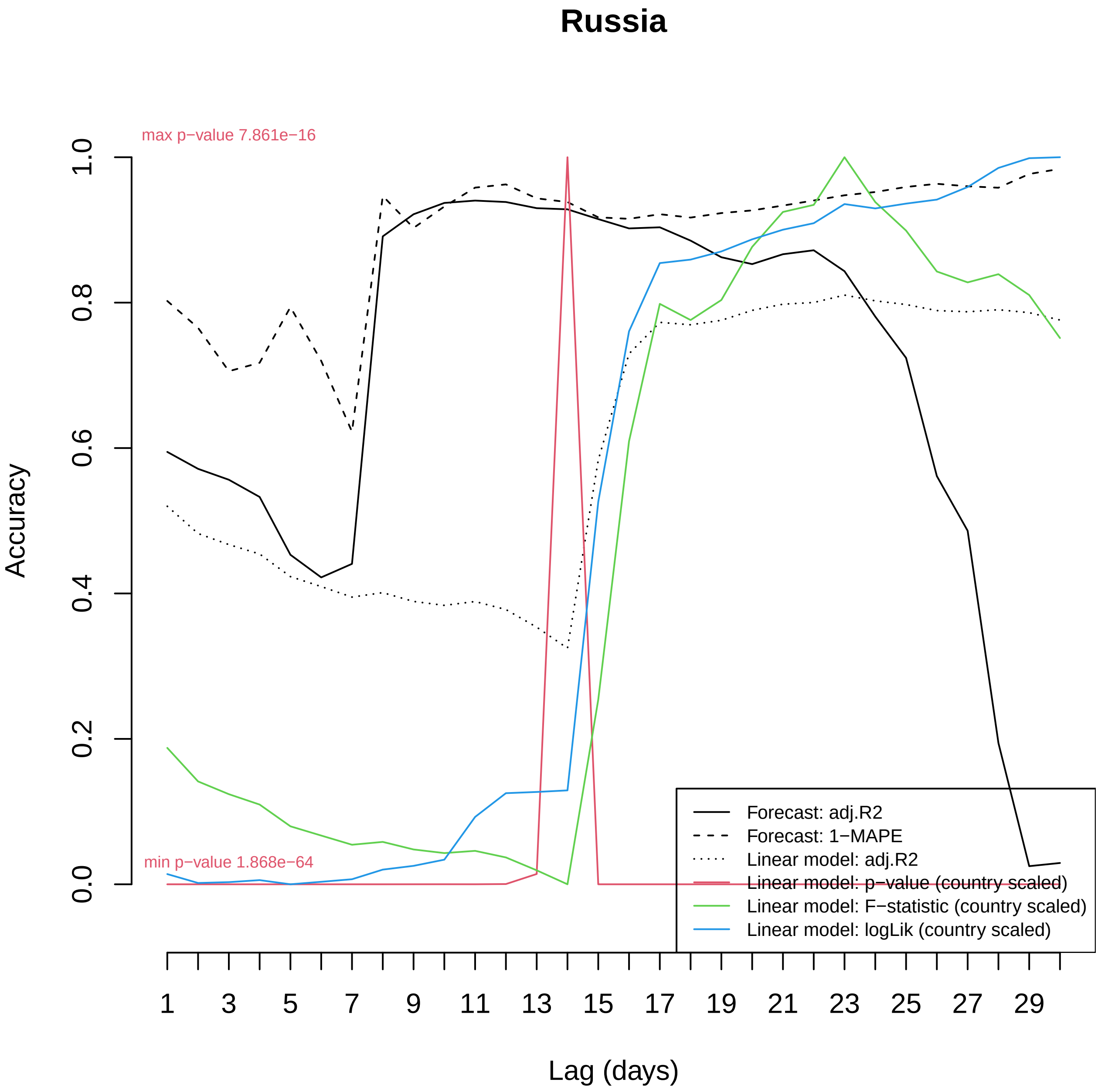

Russia
max p−value 7.861e−16
min p−value 1.868e−64
Accuracy
0.0
0.2
0.4
0.6
0.8
1.0
1
3
5
7
9
11
13
15
17
19
21
23
25
27
29
Lag (days)
Forecast: adj.R2
Forecast: 1−MAPE
Linear model: adj.R2
Linear model: p−value (country scaled)
Linear model: F−statistic (country scaled)
Linear model: logLik (country scaled)

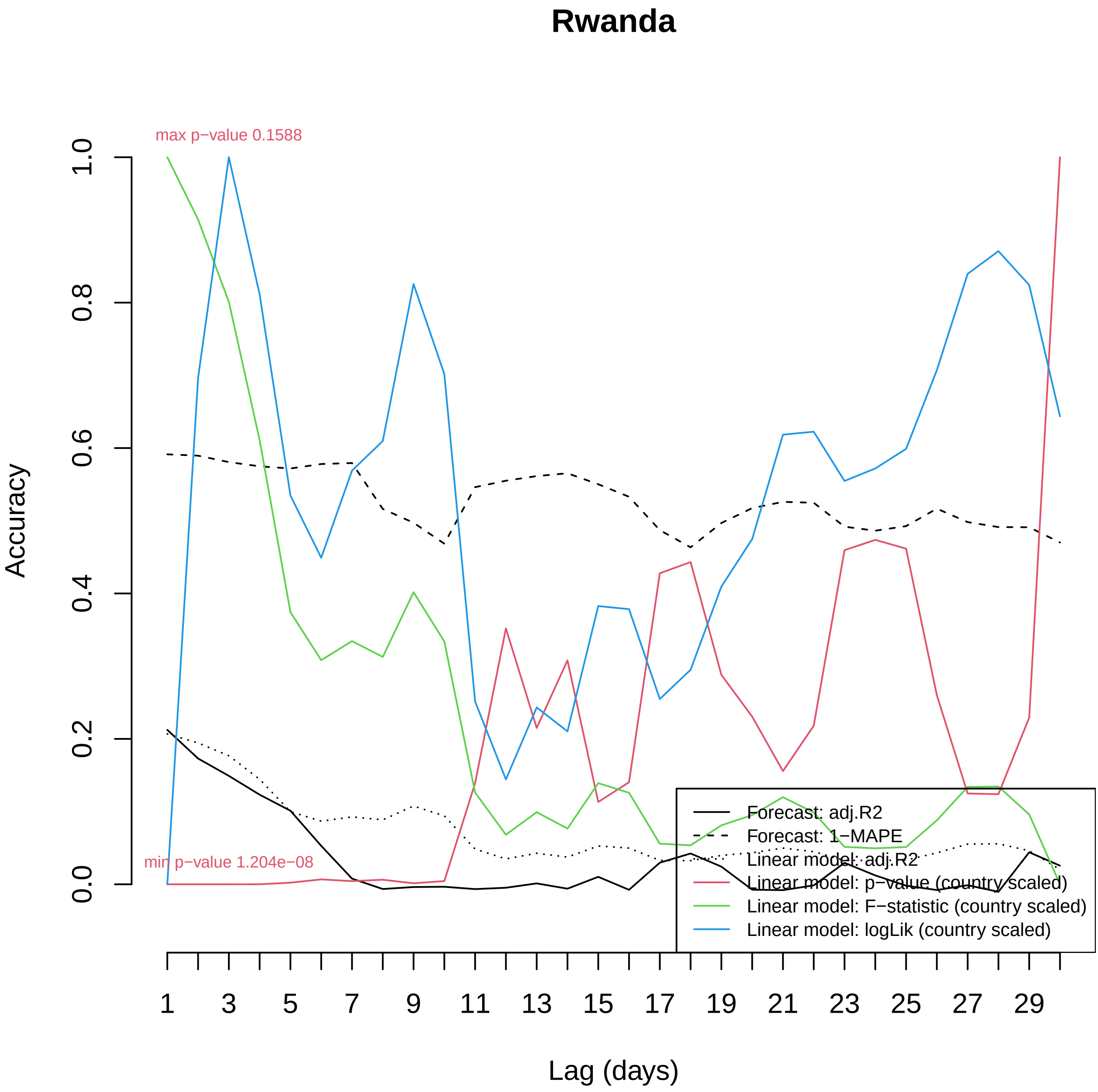

Rwanda
max p−value 0.1588
min p−value 1.204e−08
Accuracy
0.0
0.2
0.4
0.6
0.8
1.0
Lag (days)
1
3
5
7
9
11
13
15
17
19
21
23
25
27
29
Forecast: adj.R2
Forecast: 1−MAPE
Linear model: adj.R2
Linear model: p−value (country scaled)
Linear model: F−statistic (country scaled)
Linear model: logLik (country scaled)

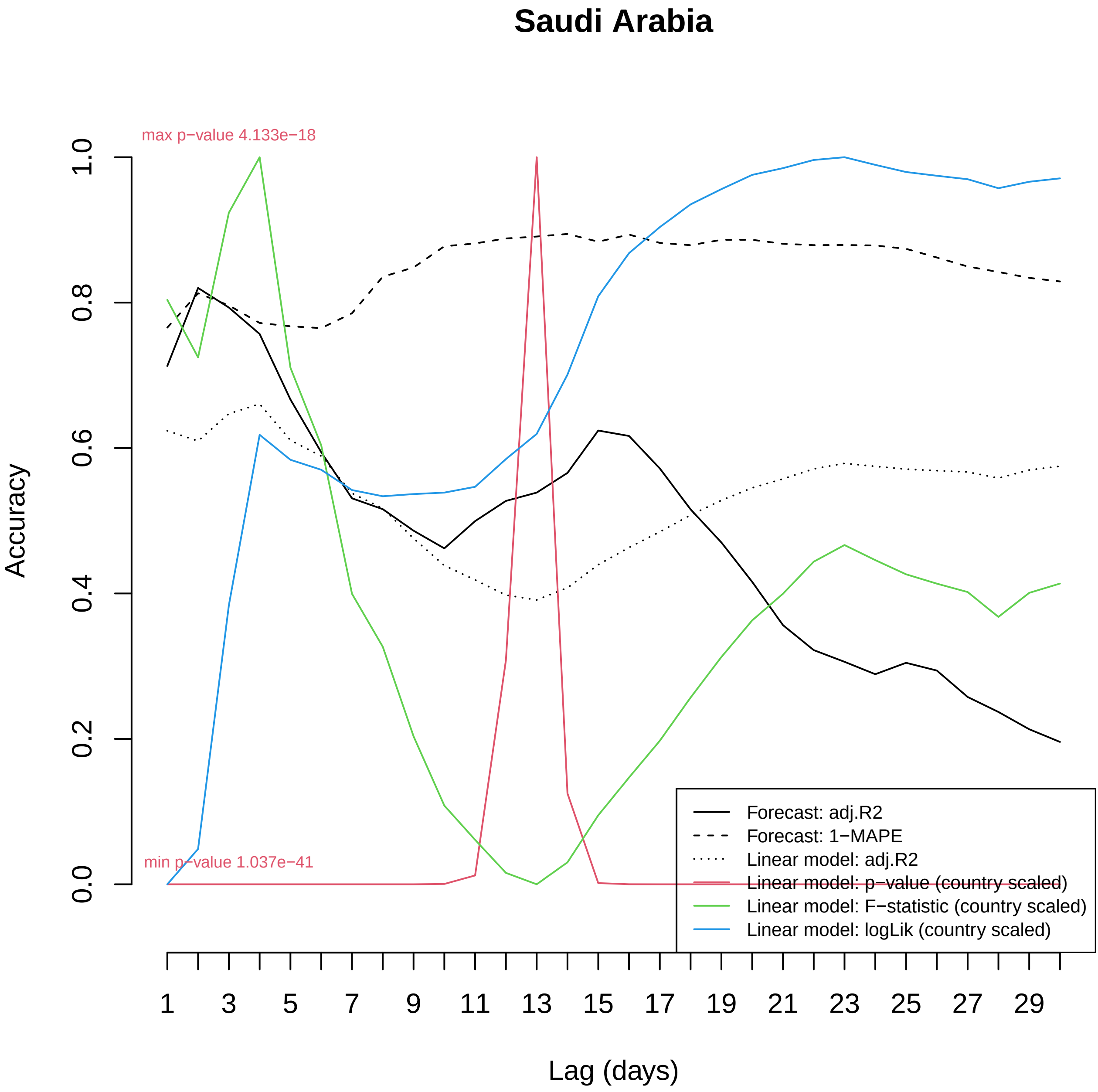
Saudi Arabia
max p−value 4.133e−18
min p−value 1.037e−41
Accuracy
0.0
0.2
0.4
0.6
0.8
1.0
Lag (days)
1
3
5
7
9
11
13
15
17
19
21
23
25
27
29
Forecast: adj.R2
Forecast: 1−MAPE
Linear model: adj.R2
Linear model: p−value (country scaled)
Linear model: F−statistic (country scaled)
Linear model: logLik (country scaled)

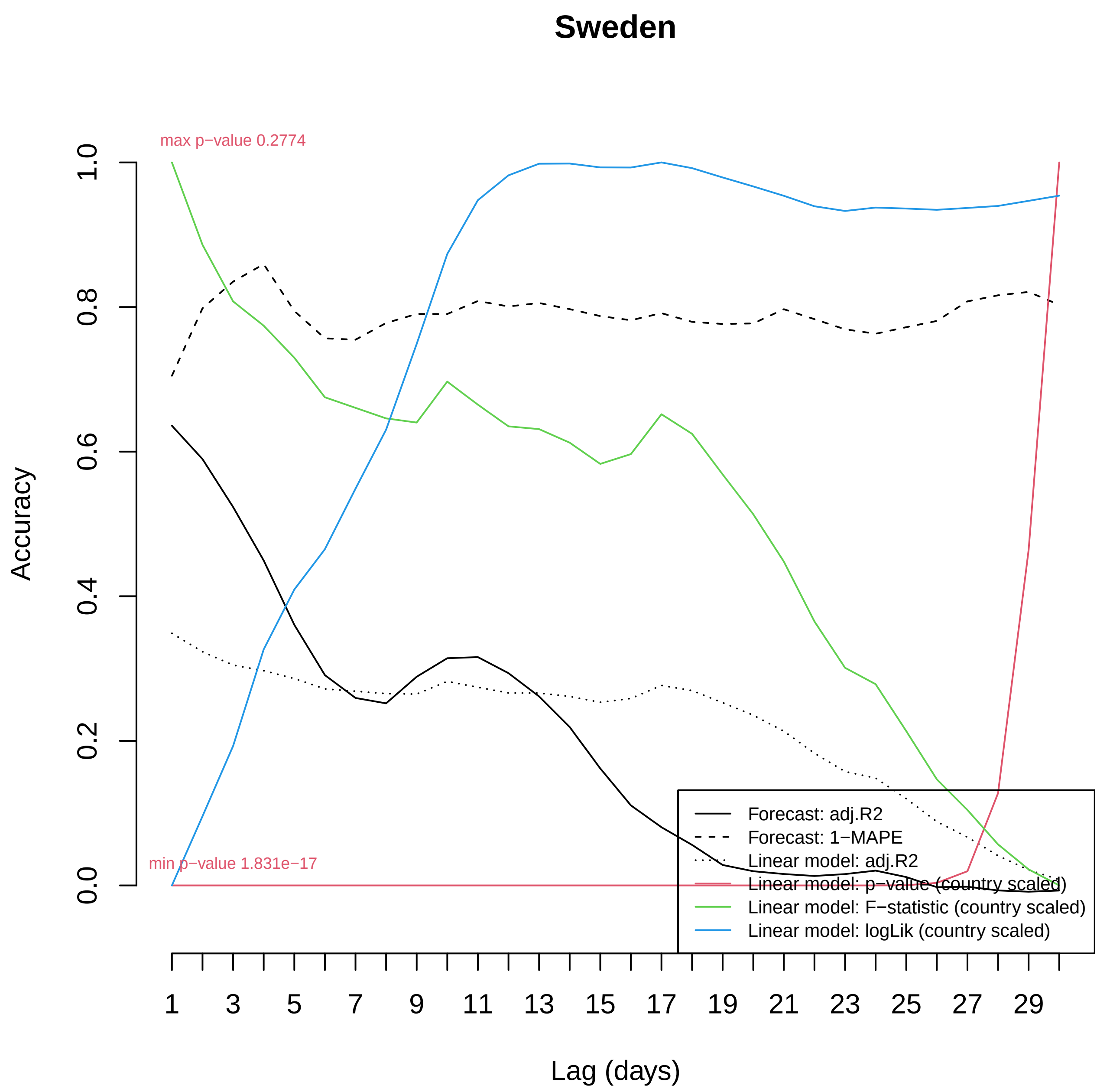
Sweden
max p−value 0.2774
min p−value 1.831e−17
Accuracy
0.0
0.2
0.4
0.6
0.8
1.0
Lag (days)
1
3
5
7
9
11
13
15
17
19
21
23
25
27
29
Forecast: adj.R2
Forecast: 1−MAPE
Linear model: adj.R2
Linear model: p−value (country scaled)
Linear model: F−statistic (country scaled)
Linear model: logLik (country scaled)

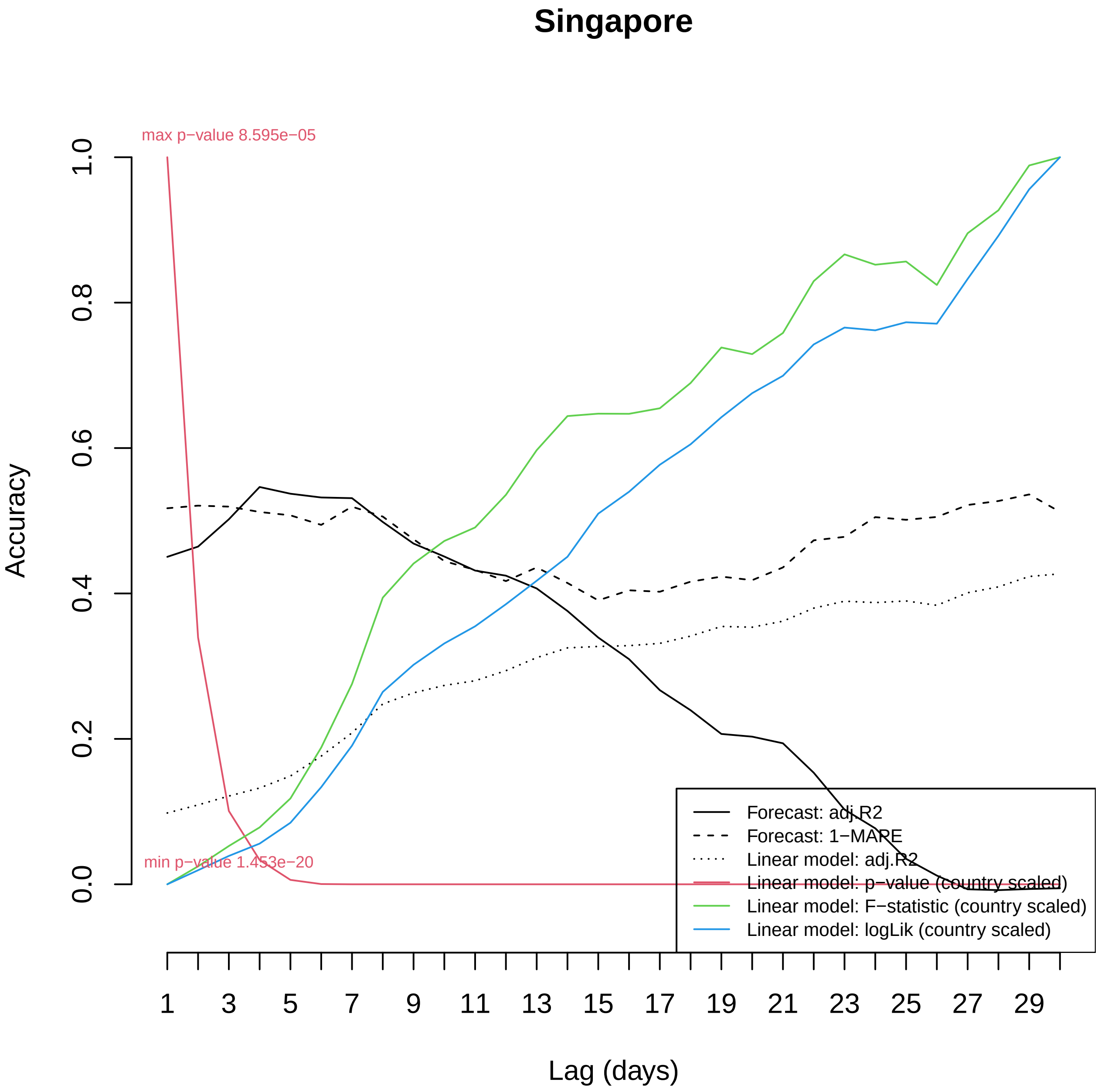

Singapore
max p−value 8.595e−05
min p−value 1.453e−20
Accuracy
0.0
0.2
0.4
0.6
0.8
1.0
Lag (days)
1
3
5
7
9
11
13
15
17
19
21
23
25
27
29
Forecast: adj.R2
Forecast: 1−MAPE
Linear model: adj.R2
Linear model: p−value (country scaled)
Linear model: F−statistic (country scaled)
Linear model: logLik (country scaled)

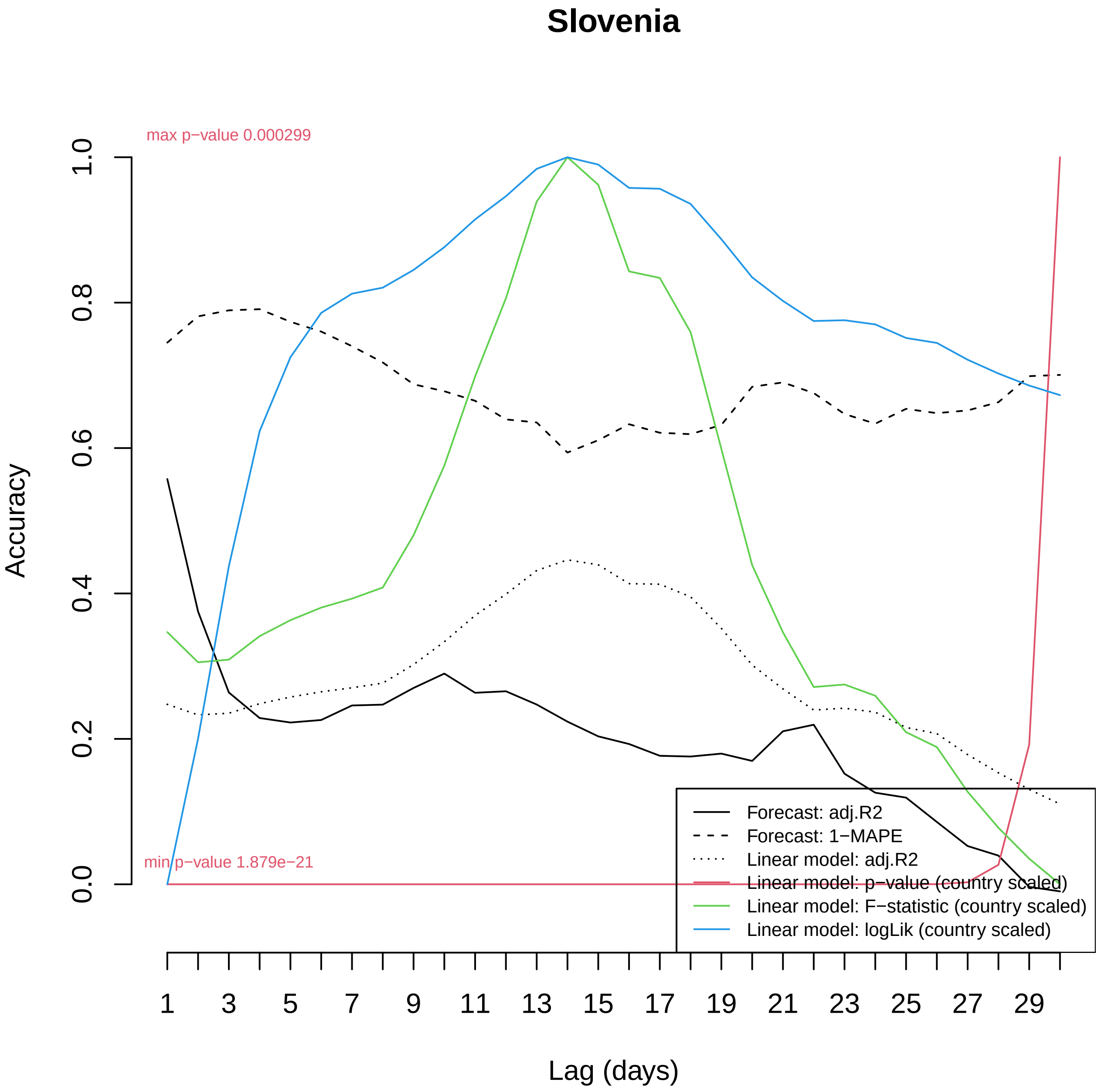

Slovenia
max p−value 0.000299
min p−value 1.879e−21
Accuracy
0.0
0.2
0.4
0.6
0.8
1.0
Lag (days)
1
3
5
7
9
11
13
15
17
19
21
23
25
27
29
Forecast: adj.R2
Forecast: 1−MAPE
Linear model: adj.R2
Linear model: p−value (country scaled)
Linear model: F−statistic (country scaled)
Linear model: logLik (country scaled)

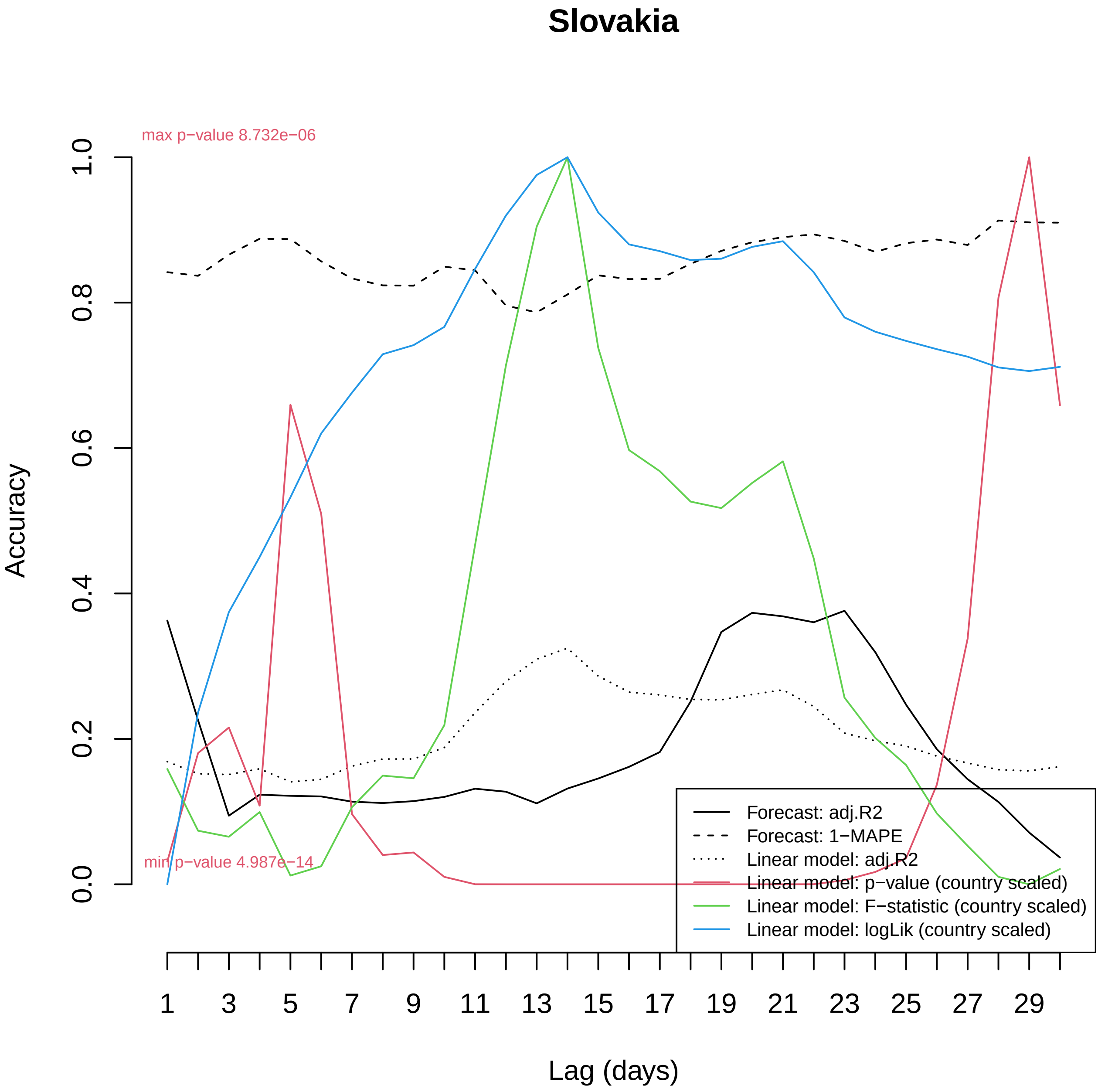
Slovakia
max p−value 8.732e−06
min p−value 4.987e−14
Accuracy
0.0
0.2
0.4
0.6
0.8
1.0
Lag (days)
1
3
5
7
9
11
13
15
17
19
21
23
25
27
29
Forecast: adj.R2
Forecast: 1−MAPE
Linear model: adj.R2
Linear model: p−value (country scaled)
Linear model: F−statistic (country scaled)
Linear model: logLik (country scaled)

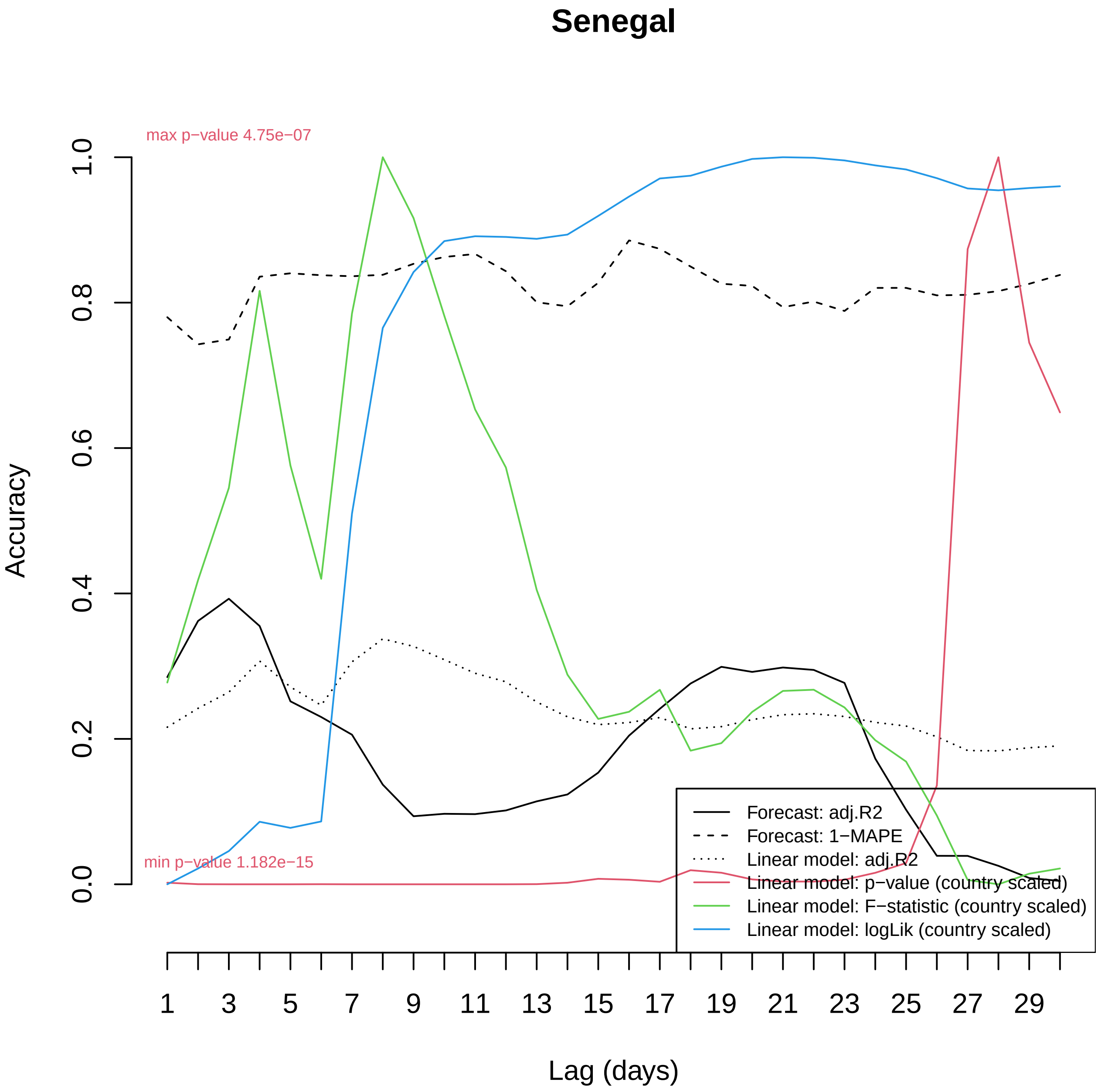

Senegal
max p−value 4.75e−07
min p−value 1.182e−15
Accuracy
0.0
0.2
0.4
0.6
0.8
1.0
Lag (days)
1
3
5
7
9
11
13
15
17
19
21
23
25
27
29
Forecast: adj.R2
Forecast: 1−MAPE
Linear model: adj.R2
Linear model: p−value (country scaled)
Linear model: F−statistic (country scaled)
Linear model: logLik (country scaled)

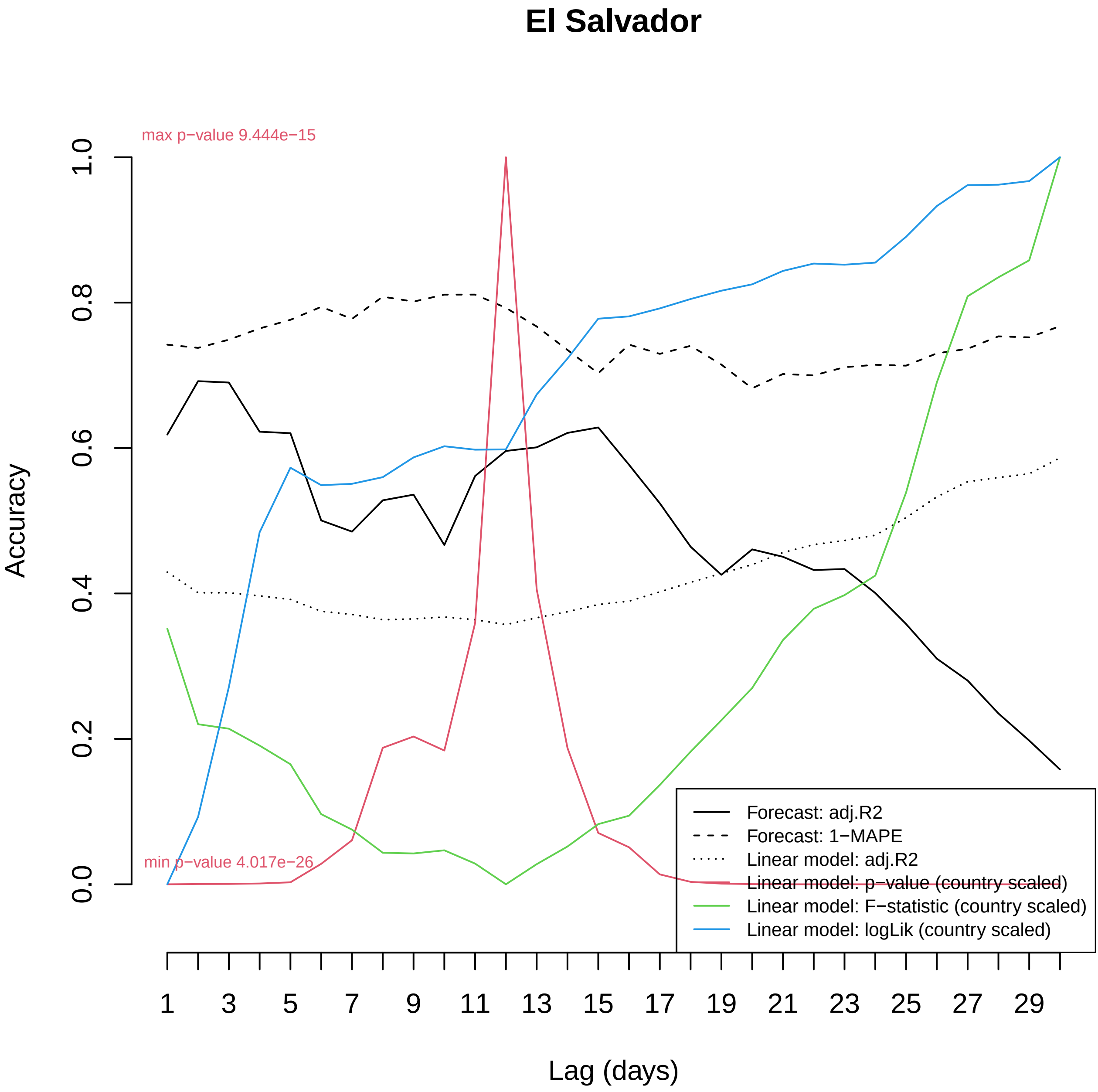

El Salvador
max p−value 9.444e−15
min p−value 4.017e−26
Accuracy
0.0
0.2
0.4
0.6
0.8
1.0
Lag (days)
1
3
5
7
9
11
13
15
17
19
21
23
25
27
29
Forecast: adj.R2
Forecast: 1−MAPE
Linear model: adj.R2
Linear model: p−value (country scaled)
Linear model: F−statistic (country scaled)
Linear model: logLik (country scaled)

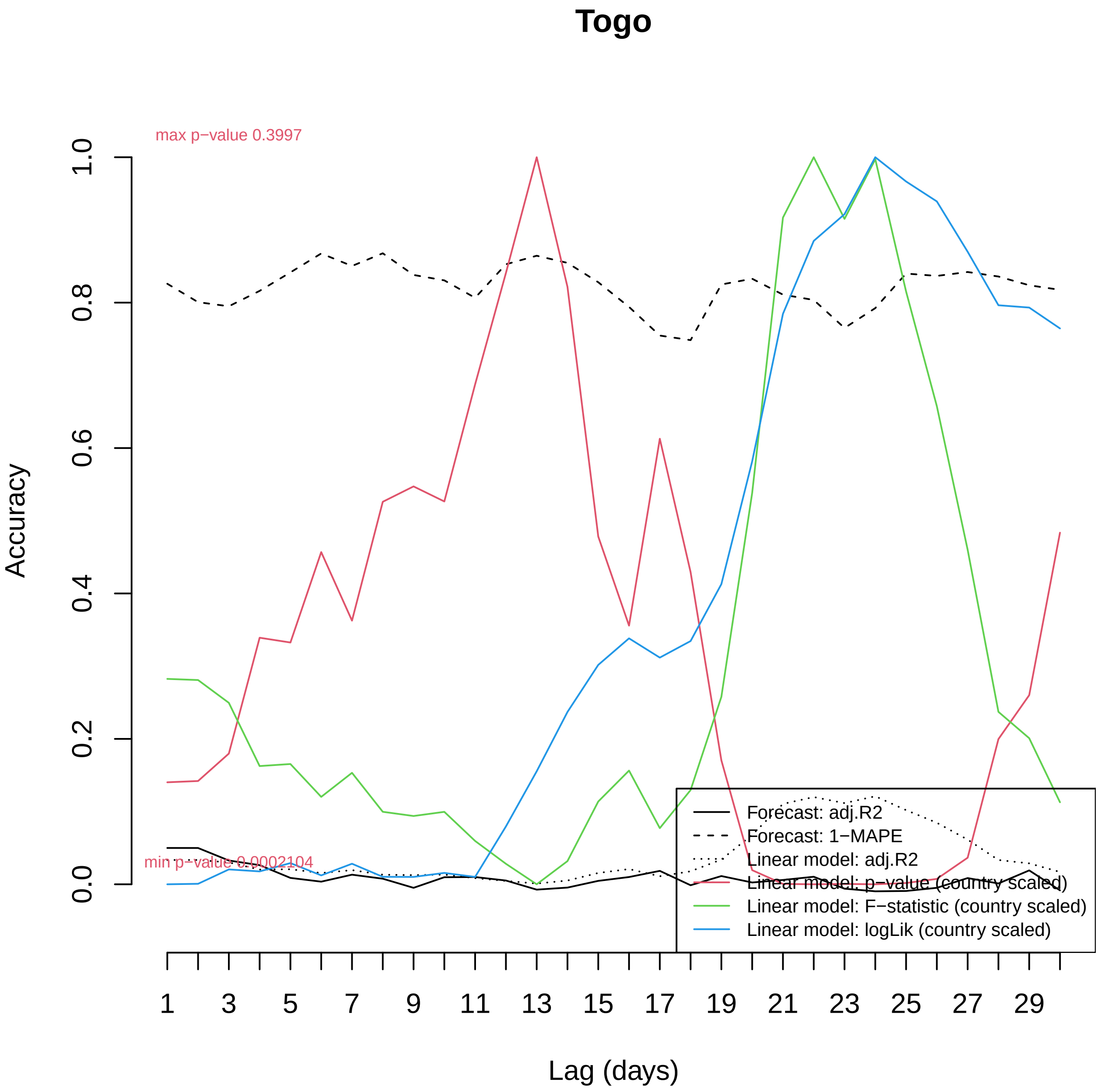

Togo
max p−value 0.3997
min p−value 0.0002104
Accuracy
0.0
0.2
0.4
0.6
0.8
1.0
Lag (days)
1
3
5
7
9
11
13
15
17
19
21
23
25
27
29
Forecast: adj.R2
Forecast: 1−MAPE
Linear model: adj.R2
Linear model: p−value (country scaled)
Linear model: F−statistic (country scaled)
Linear model: logLik (country scaled)

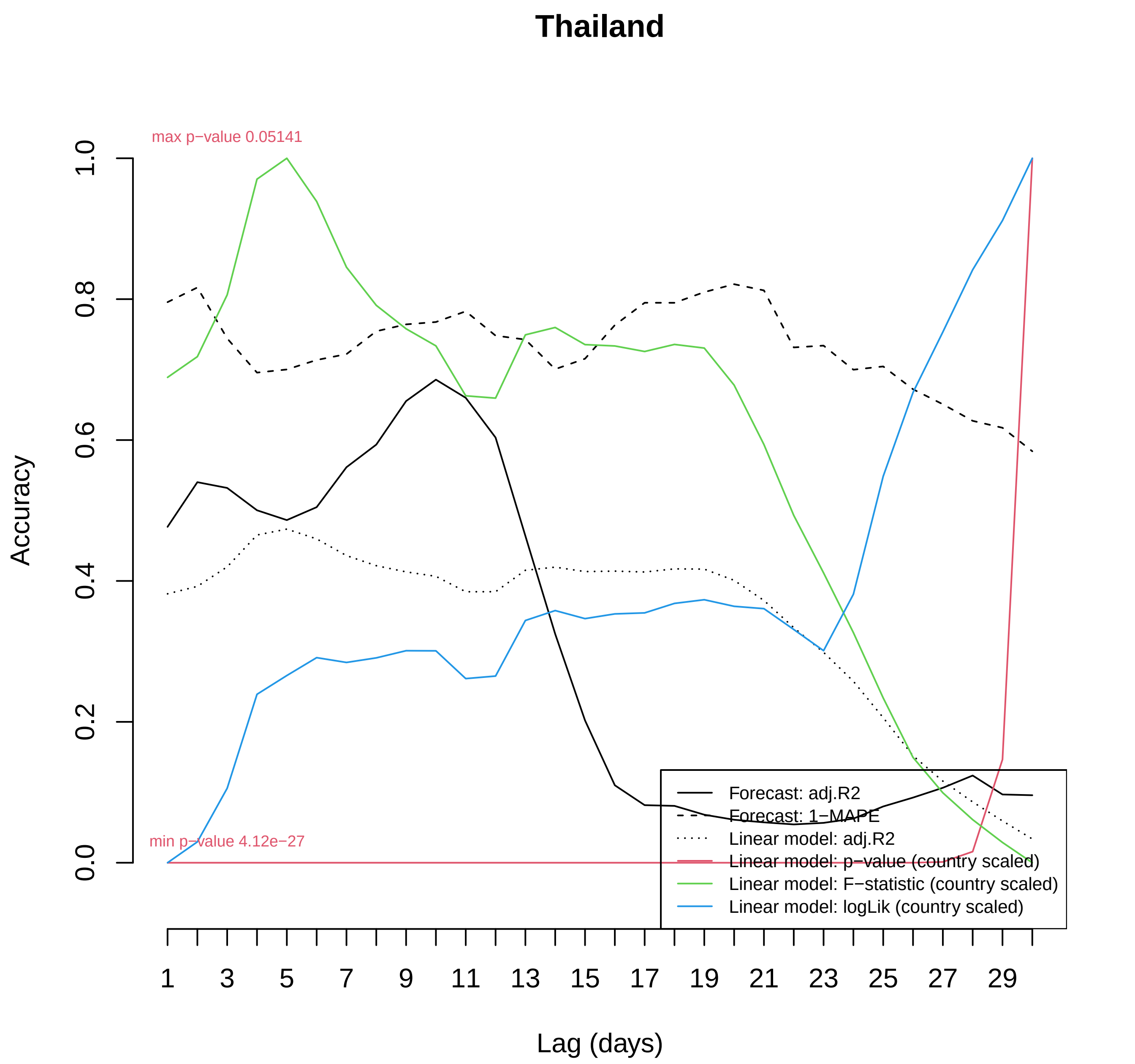

Thailand
max p−value 0.05141
min p−value 4.12e−27
Accuracy
0.0
0.2
0.4
0.6
0.8
1.0
1
3
5
7
9
11
13
15
17
19
21
23
25
27
29
Lag (days)
Forecast: adj.R2
Forecast: 1−MAPE
Linear model: adj.R2
Linear model: p−value (country scaled)
Linear model: F−statistic (country scaled)
Linear model: logLik (country scaled)

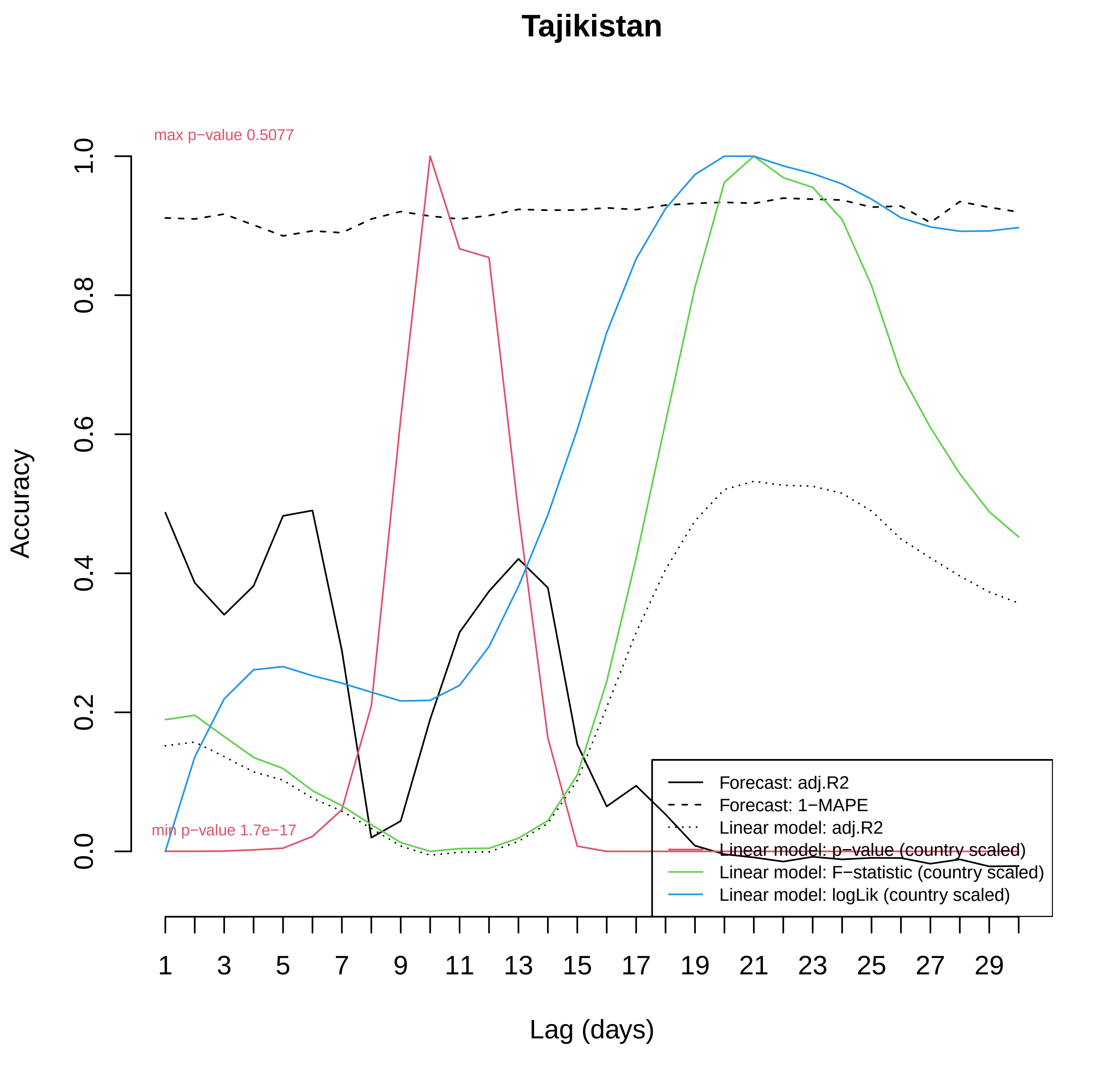
Tajikistan
max p−value 0.5077
min p−value 1.7e−17
Accuracy
0.0
0.2
0.4
0.6
0.8
1.0
1
3
5
7
9
11
13
15
17
19
21
23
25
27
29
Lag (days)
Forecast: adj.R2
Forecast: 1−MAPE
Linear model: adj.R2
Linear model: p−value (country scaled)
Linear model: F−statistic (country scaled)
Linear model: logLik (country scaled)

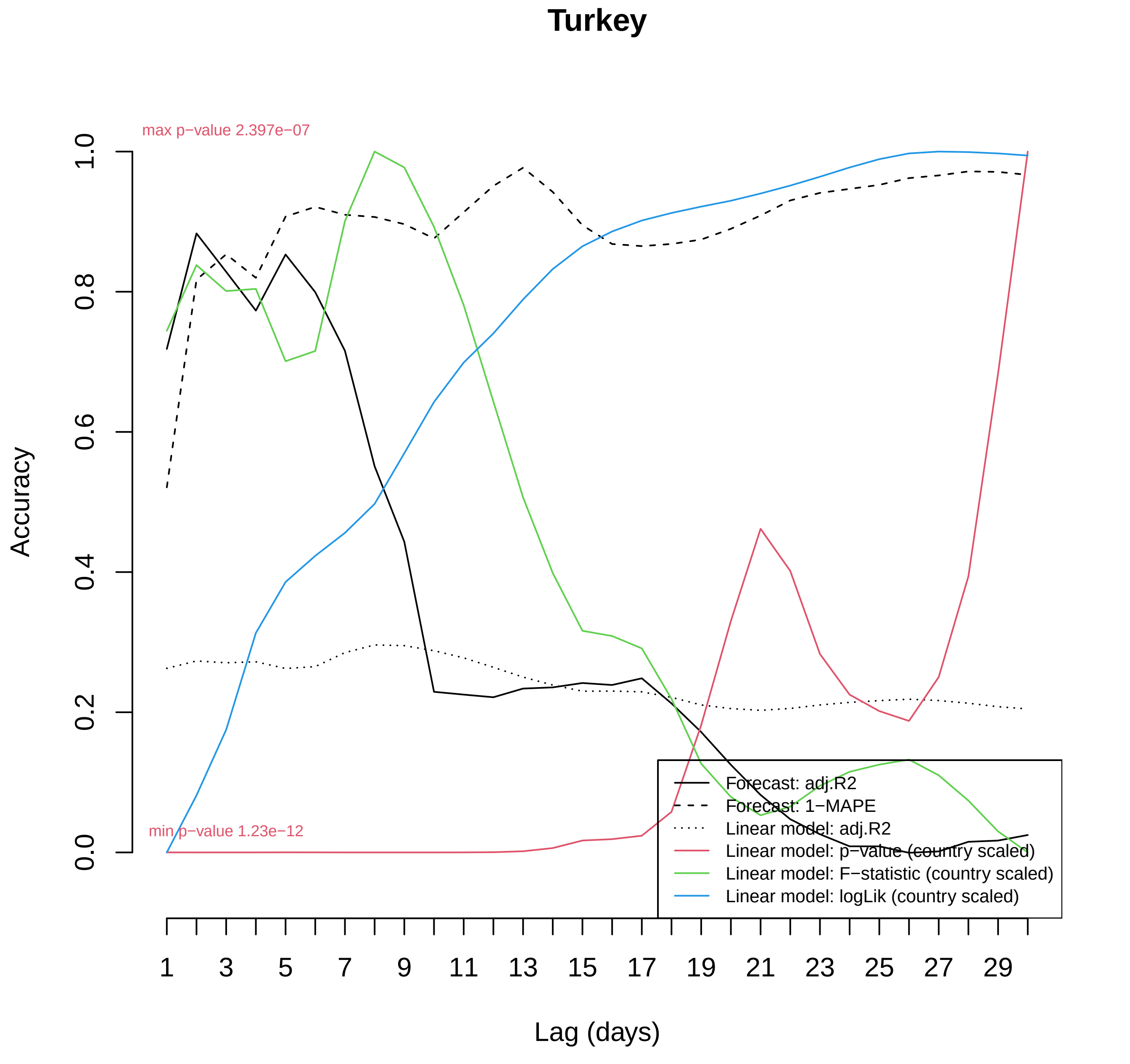
Turkey
max p−value 2.397e−07
min p−value 1.23e−12
Accuracy
0.0
0.2
0.4
0.6
0.8
1.0
1
3
5
7
9
11
13
15
17
19
21
23
25
27
29
Lag (days)
Forecast: adj.R2
Forecast: 1−MAPE
Linear model: adj.R2
Linear model: p−value (country scaled)
Linear model: F−statistic (country scaled)
Linear model: logLik (country scaled)

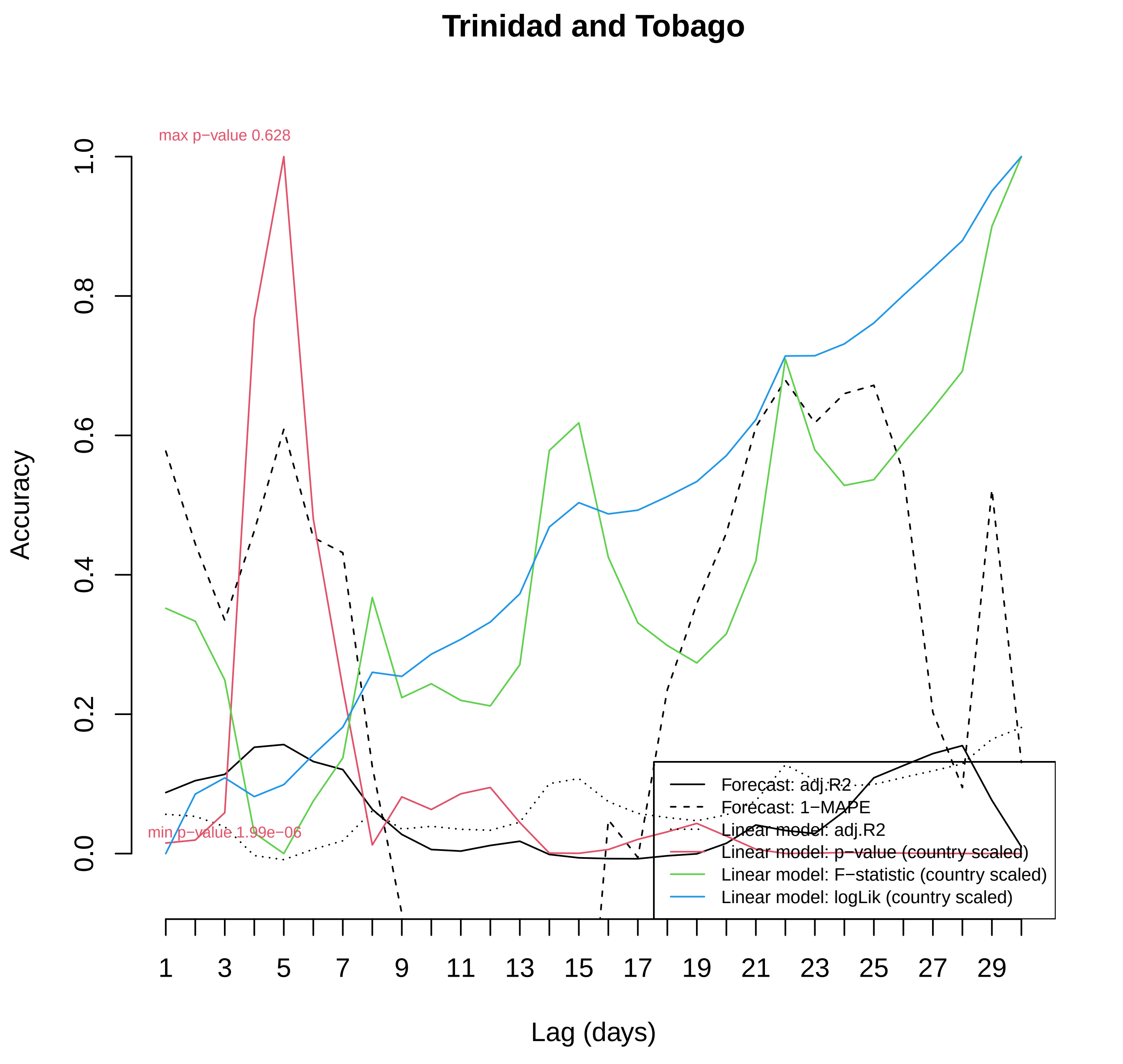

Trinidad and Tobago
max p−value 0.628
min p−value 1.99e−06
Accuracy
0.0
0.2
0.4
0.6
0.8
1.0
Lag (days)
1
3
5
7
9
11
13
15
17
19
21
23
25
27
29
Forecast: adj.R2
Forecast: 1−MAPE
Linear model: adj.R2
Linear model: p−value (country scaled)
Linear model: F−statistic (country scaled)
Linear model: logLik (country scaled)

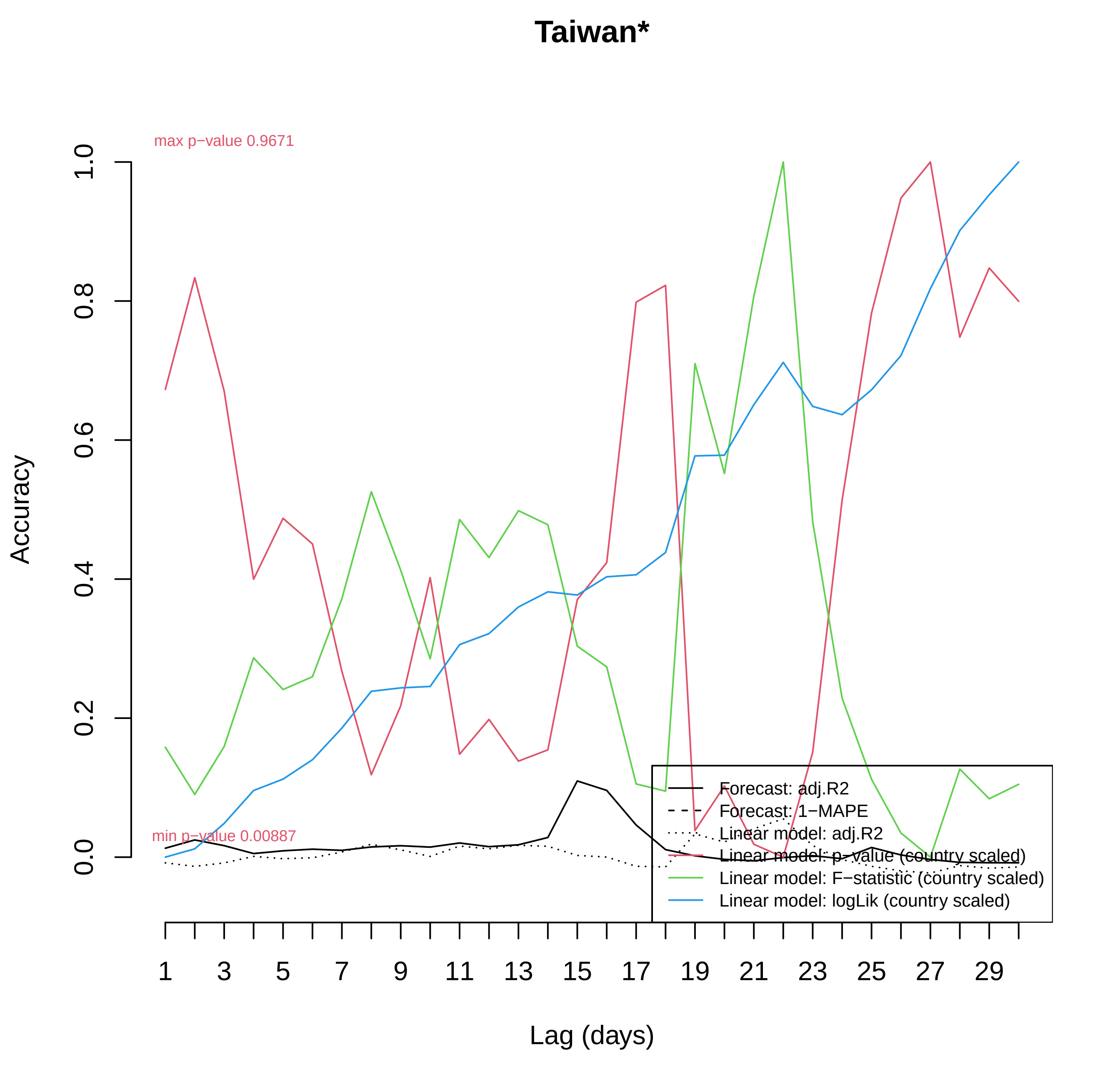

Taiwan*
max p−value 0.9671
min p−value 0.00887
Accuracy
0.0
0.2
0.4
0.6
0.8
1.0
Lag (days)
1
3
5
7
9
11
13
15
17
19
21
23
25
27
29
Forecast: adj.R2
Forecast: 1−MAPE
Linear model: adj.R2
Linear model: p−value (country scaled)
Linear model: F−statistic (country scaled)
Linear model: logLik (country scaled)

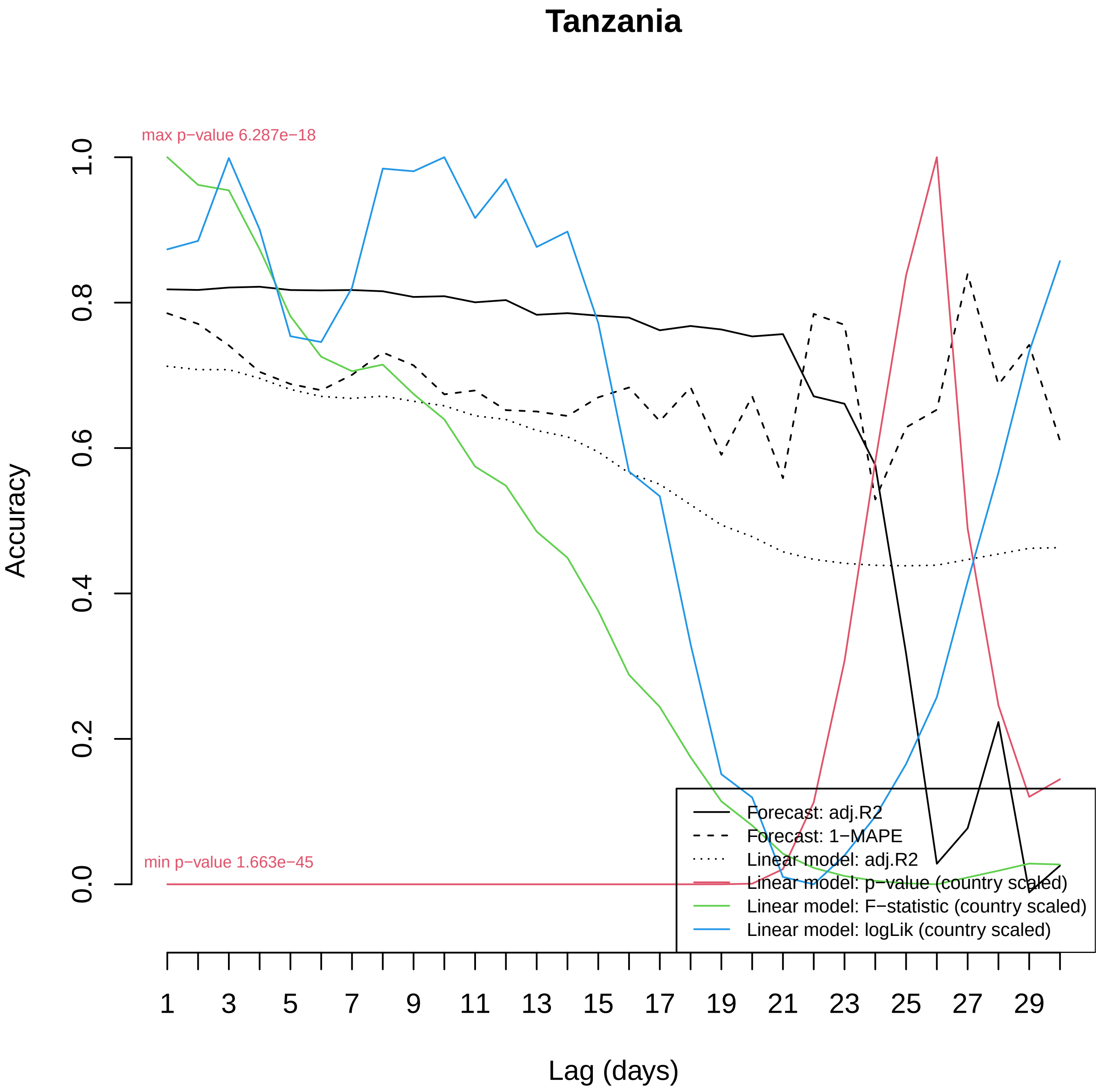

Tanzania
max p−value 6.287e−18
min p−value 1.663e−45
Accuracy
0.0
0.2
0.4
0.6
0.8
1.0
1
3
5
7
9
11
13
15
17
19
21
23
25
27
29
Lag (days)
Forecast: adj.R2
Forecast: 1−MAPE
Linear model: adj.R2
Linear model: p−value (country scaled)
Linear model: F−statistic (country scaled)
Linear model: logLik (country scaled)

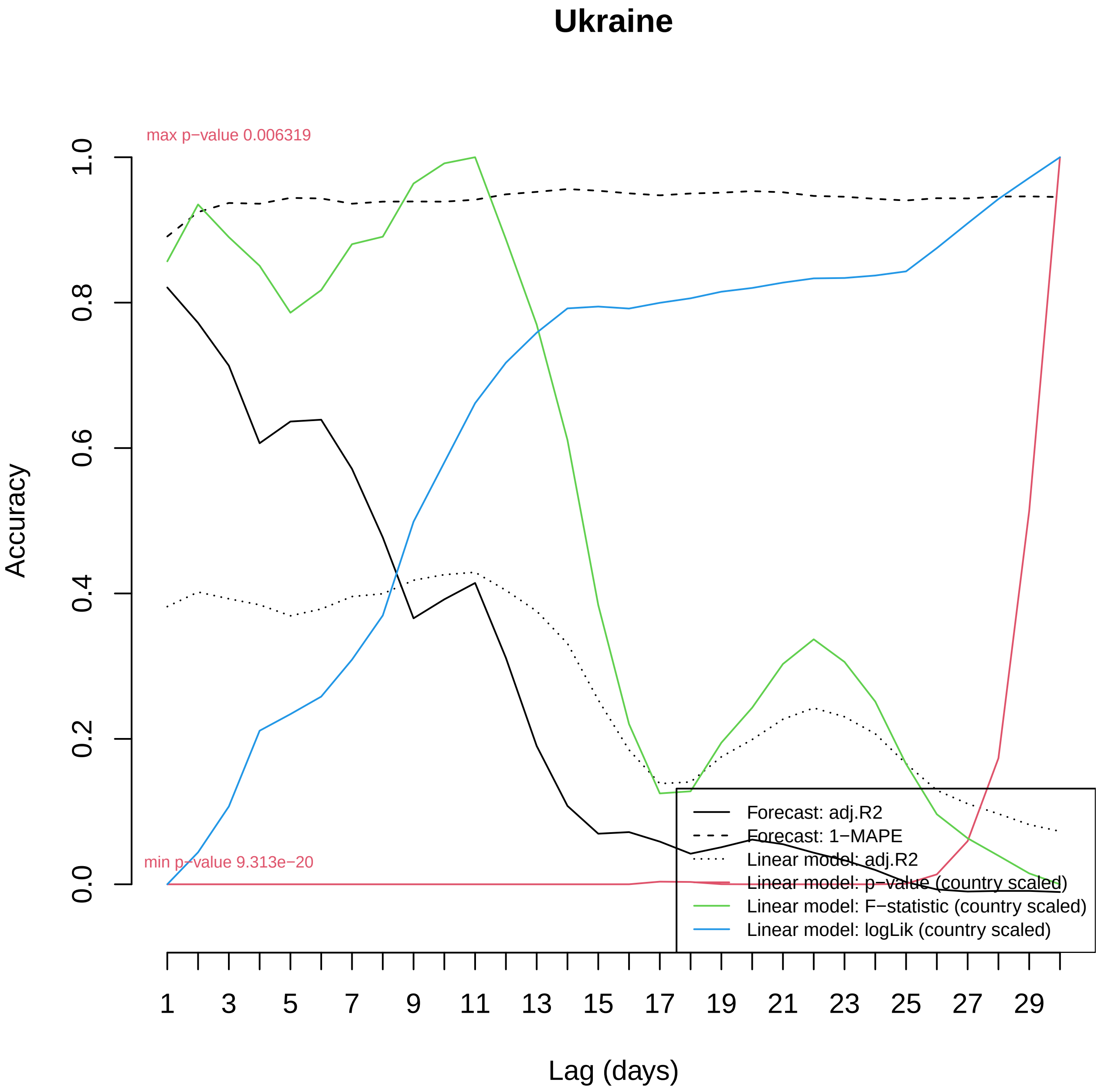

Ukraine
max p−value 0.006319
min p−value 9.313e−20
Accuracy
1.0
0.8
0.6
0.4
0.2
0.0
1
3
5
7
9
11
13
15
17
19
21
23
25
27
29
Lag (days)
Forecast: adj.R2
Forecast: 1−MAPE
Linear model: adj.R2
Linear model: p−value (country scaled)
Linear model: F−statistic (country scaled)
Linear model: logLik (country scaled)

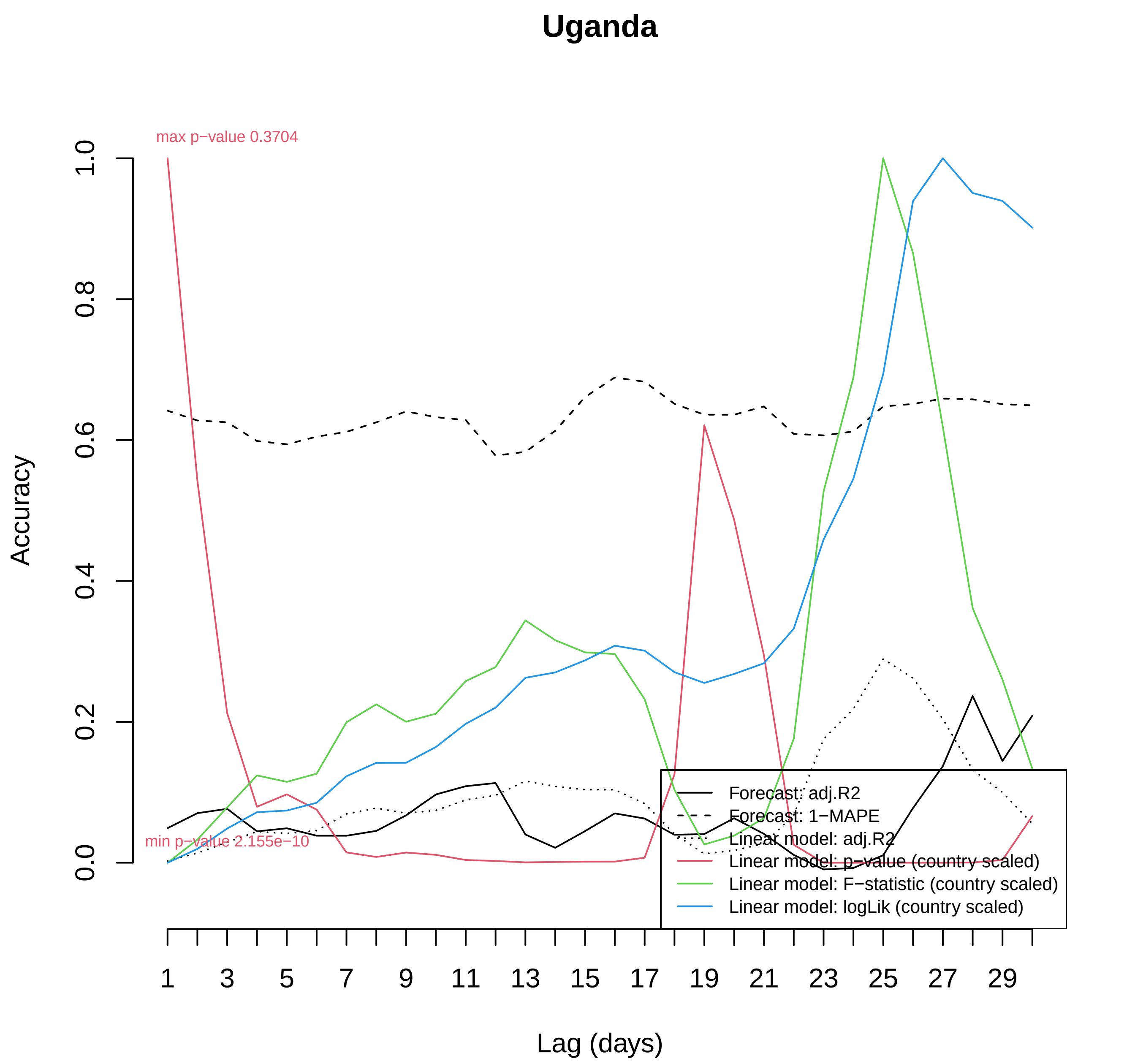

Uganda
max p−value 0.3704
min p−value 2.155e−10
Accuracy
0.0
0.2
0.4
0.6
0.8
1.0
Lag (days)
1
3
5
7
9
11
13
15
17
19
21
23
25
27
29
Forecast: adj.R2
Forecast: 1−MAPE
Linear model: adj.R2
Linear model: p−value (country scaled)
Linear model: F−statistic (country scaled)
Linear model: logLik (country scaled)

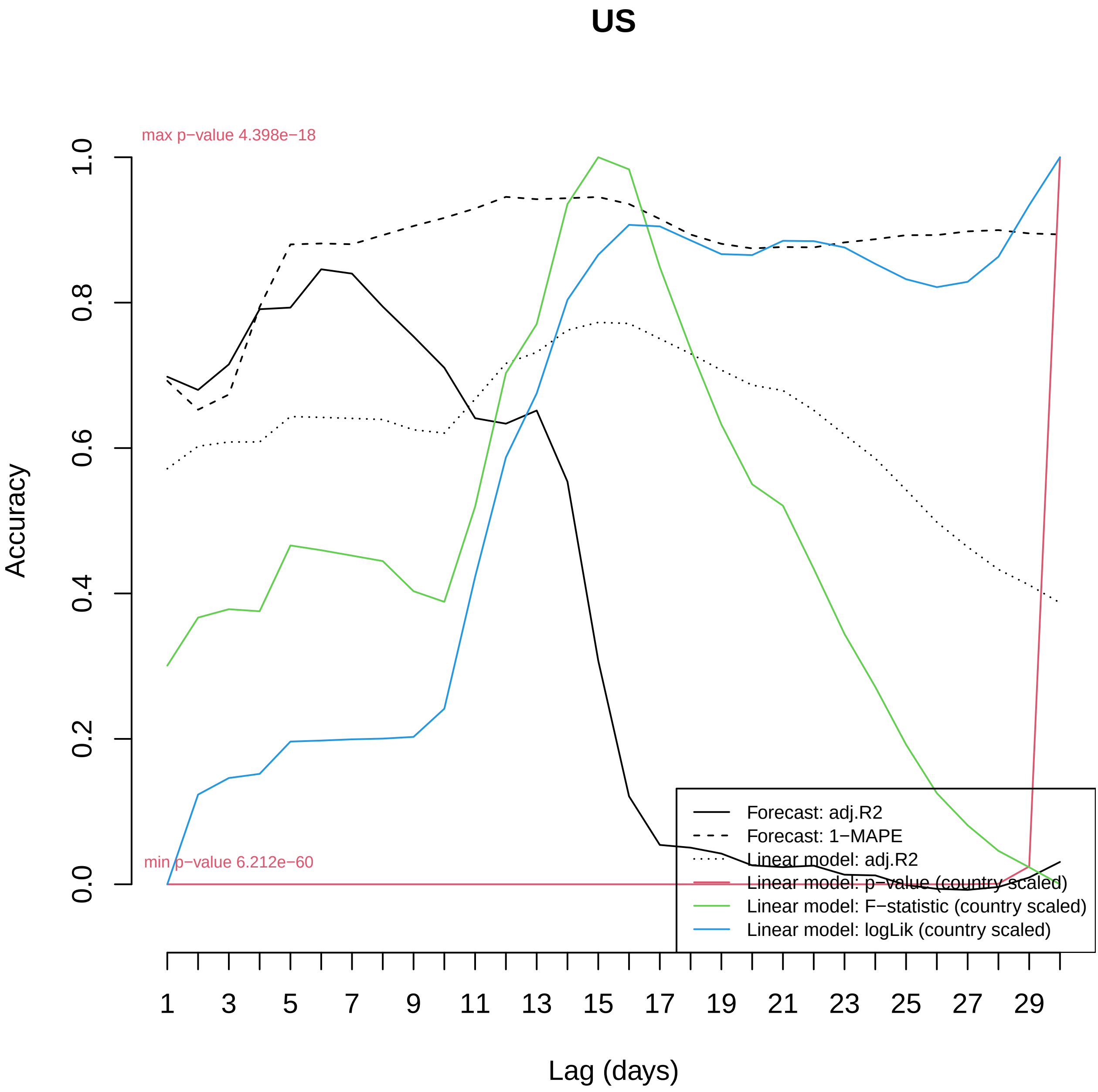

US
max p−value 4.398e−18
min p−value 6.212e−60
Accuracy
0.0
0.2
0.4
0.6
0.8
1.0
1
3
5
7
9
11
13
15
17
19
21
23
25
27
29
Lag (days)
Forecast: adj.R2
Forecast: 1−MAPE
Linear model: adj.R2
Linear model: p−value (country scaled)
Linear model: F−statistic (country scaled)
Linear model: logLik (country scaled)

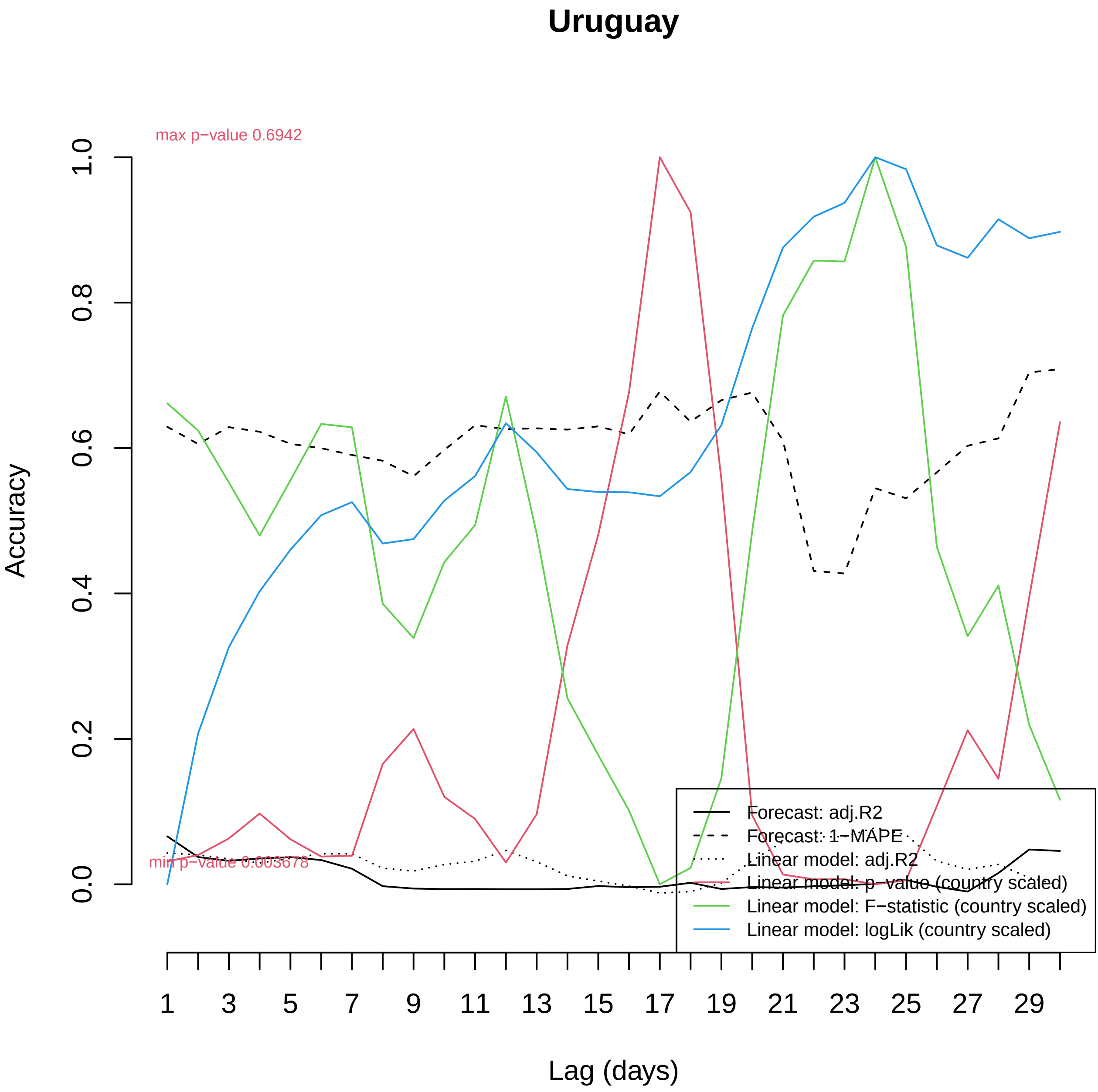
Uruguay
max p−value 0.6942
min p−value 0.003678
Accuracy
0.0
0.2
0.4
0.6
0.8
1.0
Lag (days)
1
3
5
7
9
11
13
15
17
19
21
23
25
27
29
Forecast: adj.R2
Forecast: 1−MAPE
Linear model: adj.R2
Linear model: p−value (country scaled)
Linear model: F−statistic (country scaled)
Linear model: logLik (country scaled)

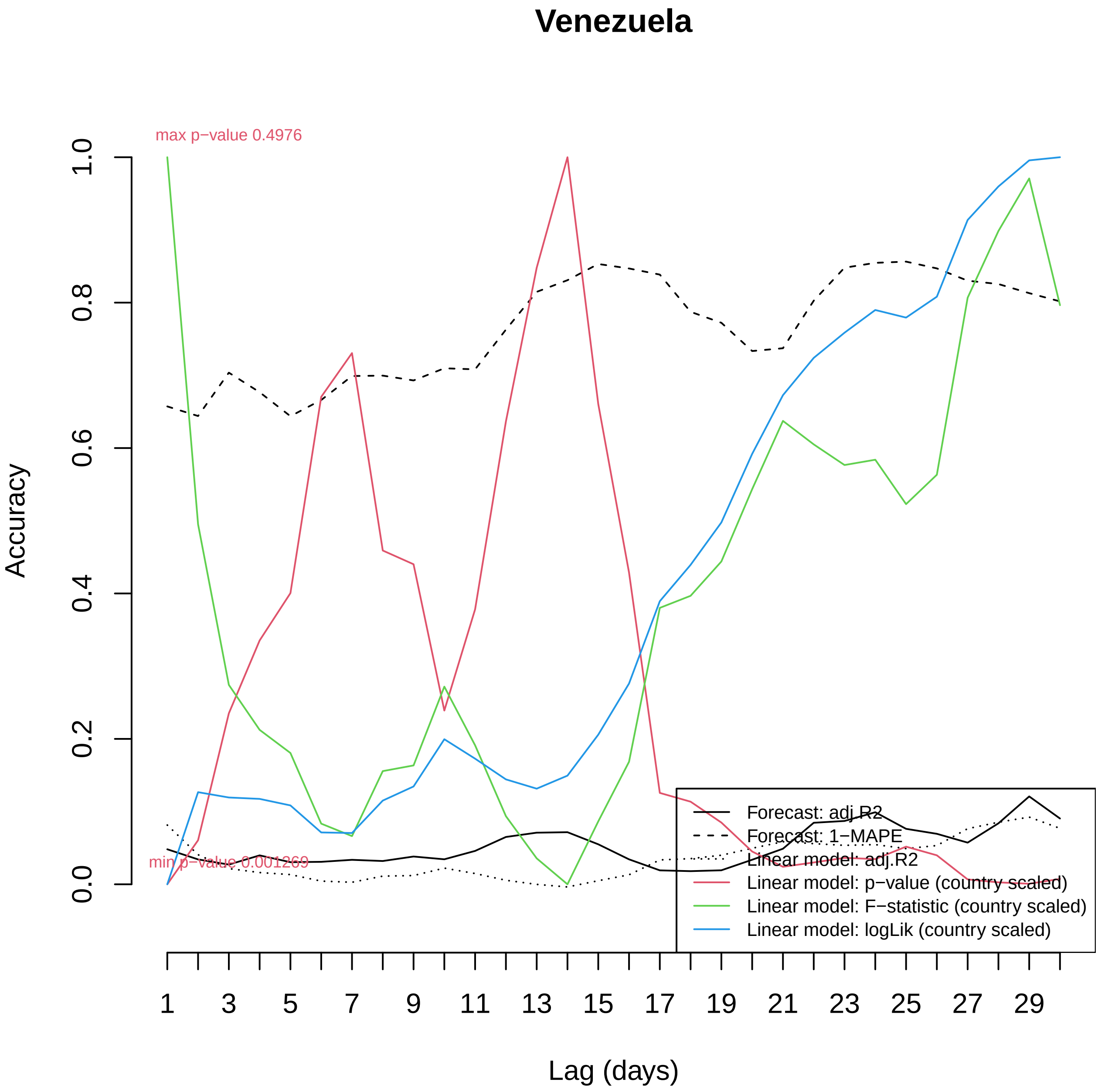

Venezuela
max p−value 0.4976
min p−value 0.001269
Accuracy
0.0
0.2
0.4
0.6
0.8
1.0
Lag (days)
1
3
5
7
9
11
13
15
17
19
21
23
25
27
29
Forecast: adj.R2
Forecast: 1−MAPE
Linear model: adj.R2
Linear model: p−value (country scaled)
Linear model: F−statistic (country scaled)
Linear model: logLik (country scaled)

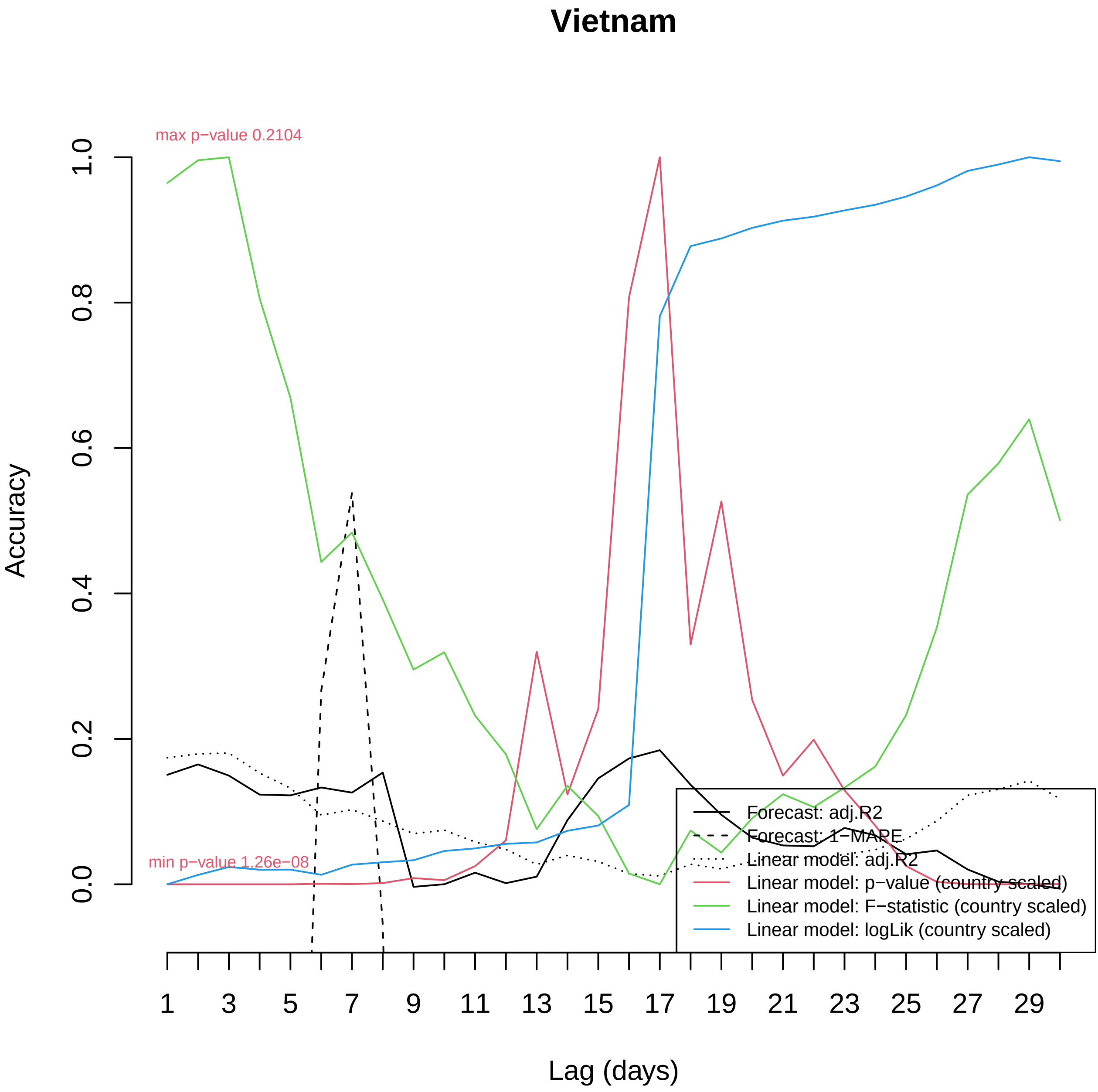

Vietnam
max p−value 0.2104
min p−value 1.26e−08
Accuracy
0.0
0.2
0.4
0.6
0.8
1.0
1
3
5
7
9
11
13
15
17
19
21
23
25
27
29
Lag (days)
Forecast: adj.R2
Forecast: 1−MAPE
Linear model: adj.R2
Linear model: p−value (country scaled)
Linear model: F−statistic (country scaled)
Linear model: logLik (country scaled)

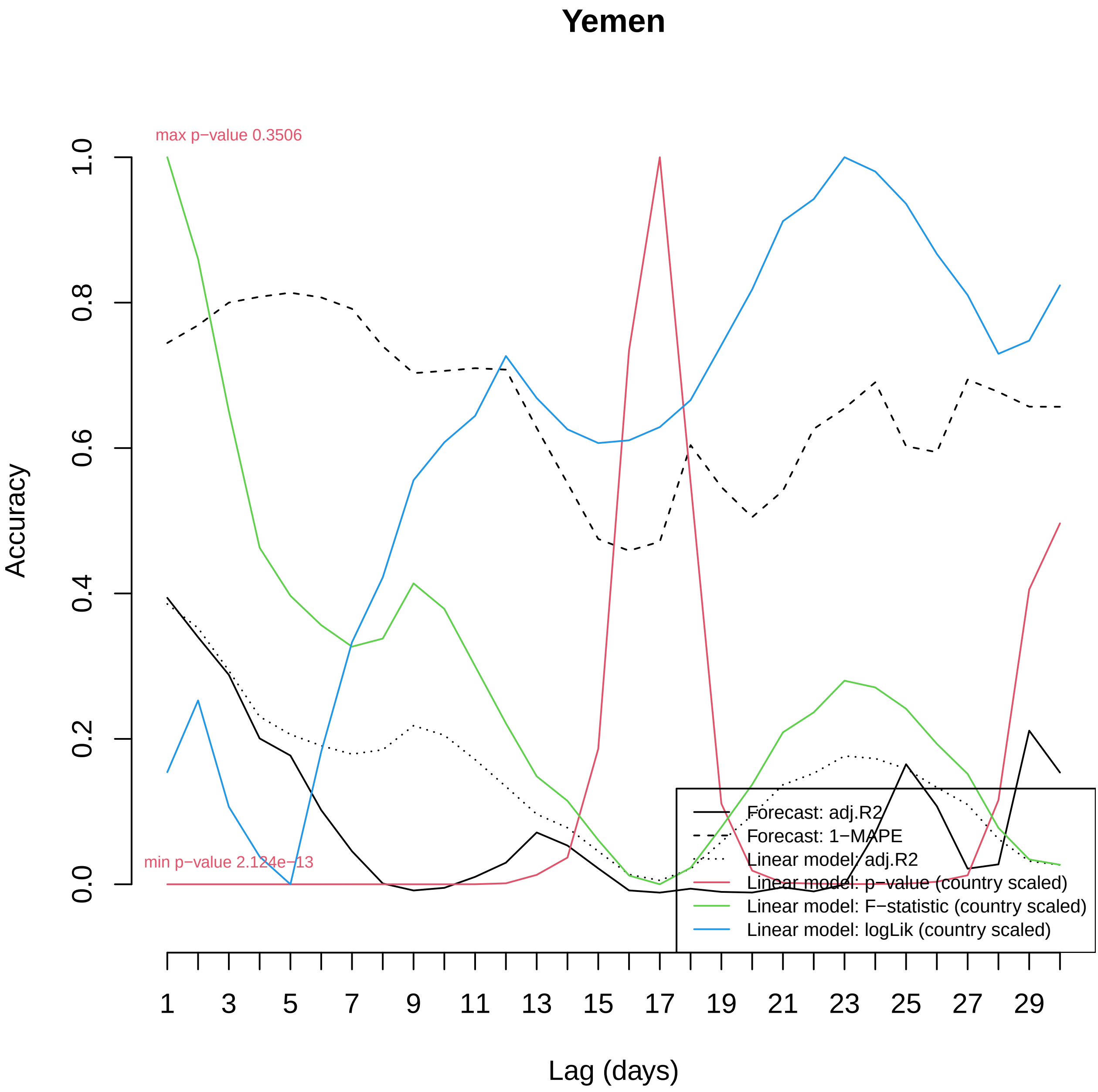
Yemen
max p−value 0.3506
min p−value 2.124e−13
Accuracy
0.0
0.2
0.4
0.6
0.8
1.0
Lag (days)
1
3
5
7
9
11
13
15
17
19
21
23
25
27
29
Forecast: adj.R2
Forecast: 1−MAPE
Linear model: adj.R2
Linear model: p−value (country scaled)
Linear model: F−statistic (country scaled)
Linear model: logLik (country scaled)

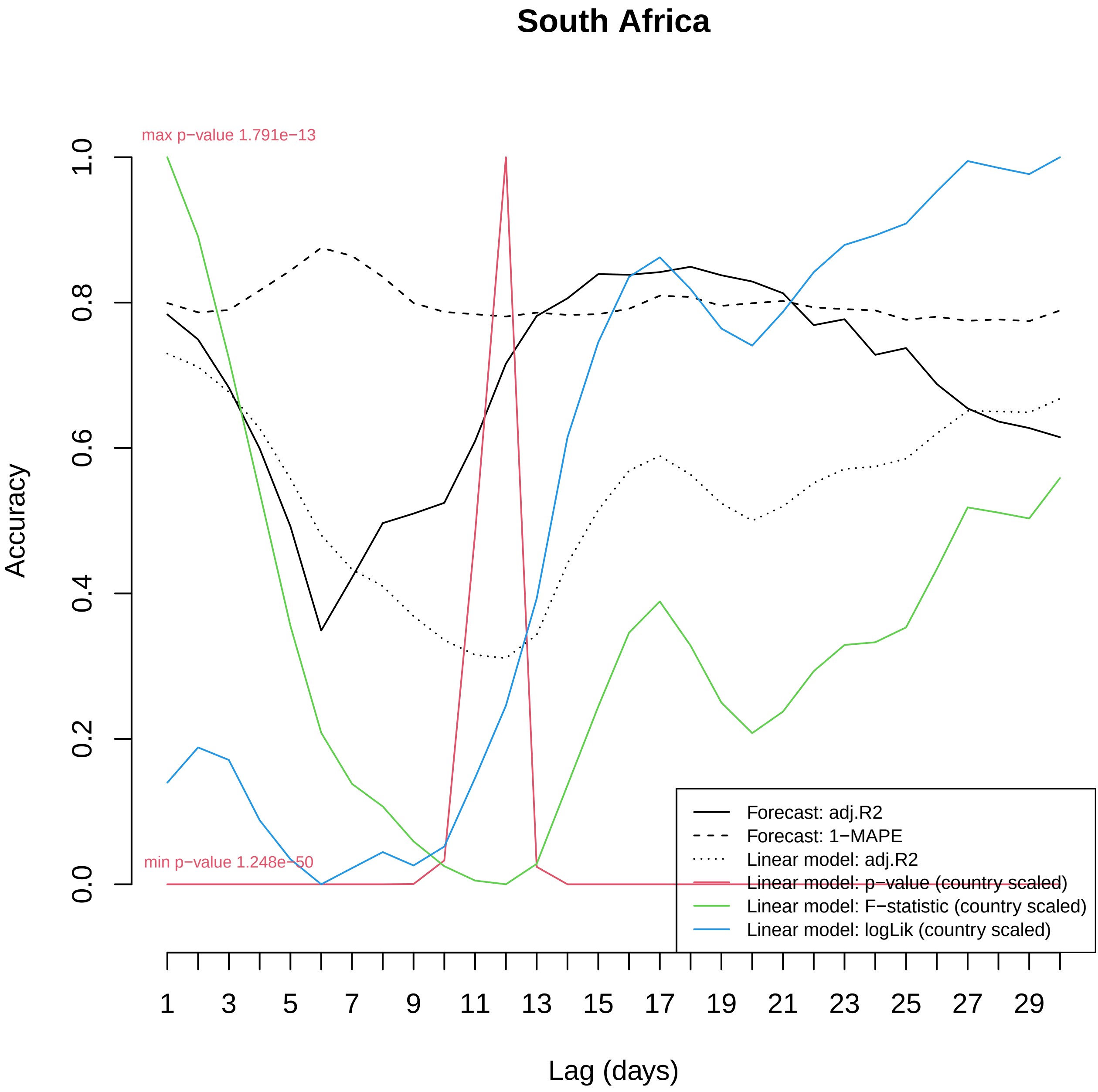

South Africa
max p−value 1.791e−13
min p−value 1.248e−50
Accuracy
0.0
0.2
0.4
0.6
0.8
1.0
Lag (days)
1
3
5
7
9
11
13
15
17
19
21
23
25
27
29
Forecast: adj.R2
Forecast: 1−MAPE
Linear model: adj.R2
Linear model: p−value (country scaled)
Linear model: F−statistic (country scaled)
Linear model: logLik (country scaled)

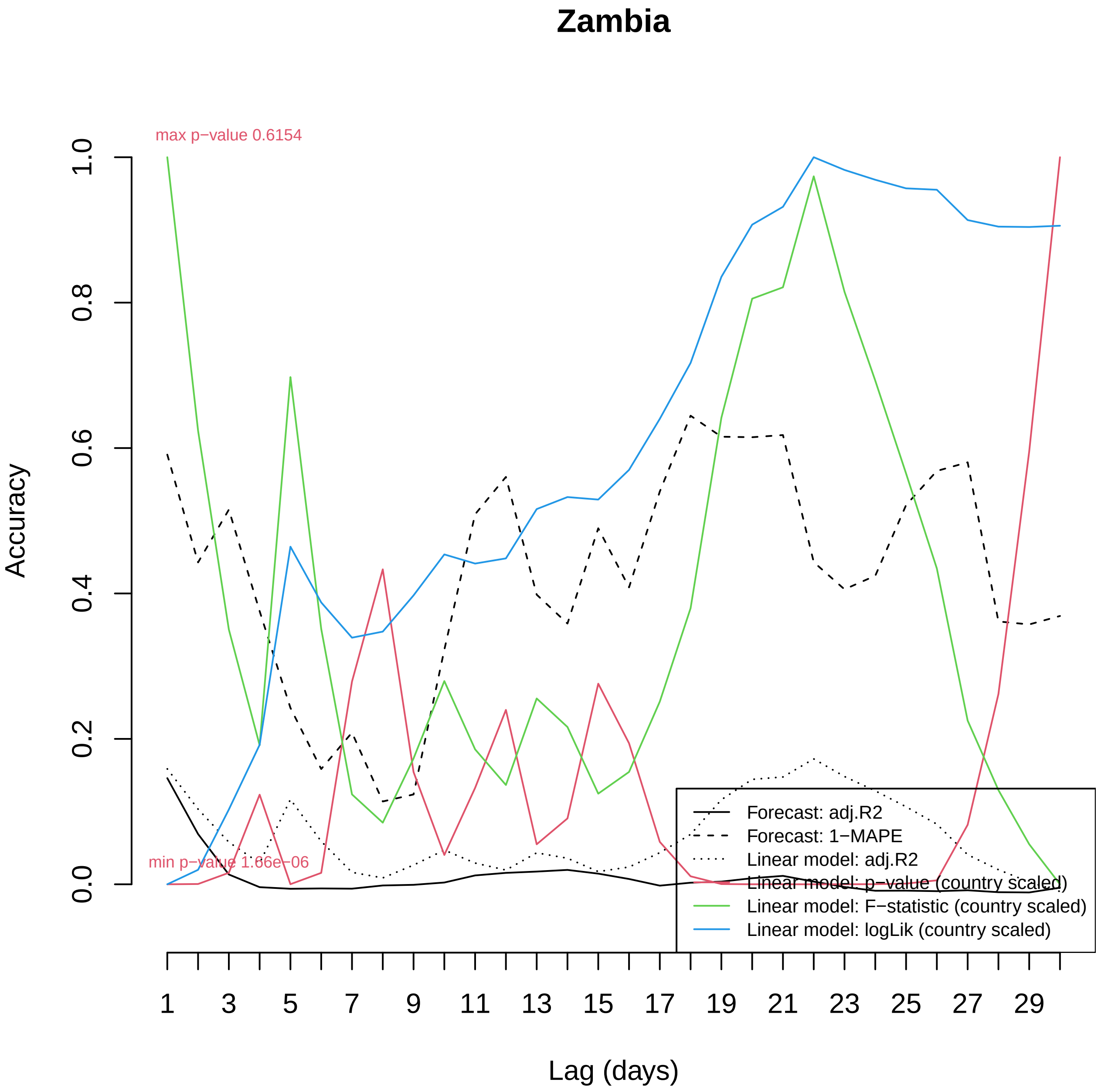

Zambia
max p−value 0.6154
min p−value 1.66e−06
Accuracy
0.0
0.2
0.4
0.6
0.8
1.0
Lag (days)
1
3
5
7
9
11
13
15
17
19
21
23
25
27
29
Forecast: adj.R2
Forecast: 1−MAPE
Linear model: adj.R2
Linear model: p−value (country scaled)
Linear model: F−statistic (country scaled)
Linear model: logLik (country scaled)

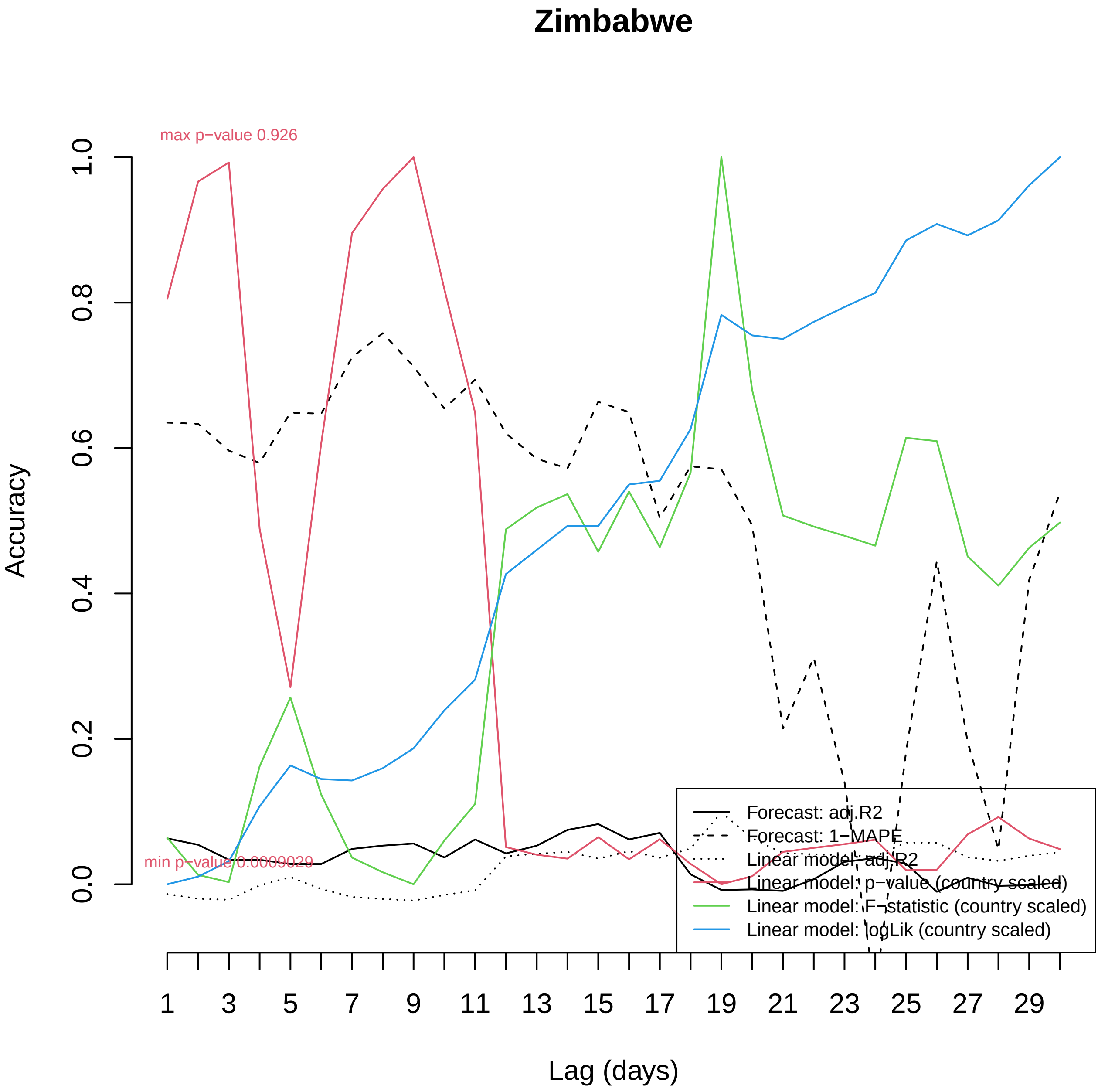
Zimbabwe
max p−value 0.926
min p−value 0.0009029
Accuracy
0.0
0.2
0.4
0.6
0.8
1.0
Lag (days)
1
3
5
7
9
11
13
15
17
19
21
23
25
27
29
Forecast: adj.R2
Forecast: 1−MAPE
Linear model: adj.R2
Linear model: p−value (country scaled)
Linear model: F−statistic (country scaled)
Linear model: logLik (country scaled)
